# Text dialogue analysis Based ChatGPT for Primary Screening of Mild Cognitive Impairment

**DOI:** 10.1101/2023.06.27.23291884

**Authors:** Changyu Wang, Siru Liu, Aiqing Li, Jialin Liu

**Affiliations:** Department of Medical Informatics, West China Medical School, Sichuan University, Chengdu, China; West China College of Stomatology, Sichuan University, Chengdu, China; Department of Biomedical Informatics, Vanderbilt University Medical Center, Nashville, TN, USA; Department of Neurology, West China Hospital, Sichuan University; Information Center, West China Hospital, Sichuan University, Chengdu, China; Department of Otolaryngology-Head and Neck Surgery, West China Hospital, Sichuan University, Chengdu, China

## Abstract

**Background:** AI models tailored to diagnose cognitive impairment have shown excellent results. However, it is unclear whether large linguistic models can rival specialized models by text alone.

**Objectives:** We would explore the effectiveness of ChatGPT for primary screening of mild cognitive impairment (MCI) and standardize the design steps and components of the prompt.

**Methods:** We obtained 174 participants from the DementiaBank screening and classified 70% of them into the training set and 30% into the test set. Three dimensions of variables were incorporated, including: vocabulary, syntax and grammar, and semantics. These variables were generated from published studies and statistical analyses. We used R 4.3.0. for the analysis of variables and diagnostic indicators.

**Results:** The final variables included by published studies included: word frequency and word ratio, phrase frequency and phrase ratio, lexical complexity, syntactic complexity, grammatical components, semantic density, and semantic coherence; variables included in the analysis included: tip-of-the-tongue phenomenon (P < 0.001), difficulty with complex ideas (P < 0.001), and memory issues (P < 0.001). The final GPT4 model achieved the sensitivity (SEN) of 0.8636, specificity (SPE) of 0.9487 and area under the curve (AUC) of 0.9062 on the training set; on the test set, the SEN, SPE and AUC reached 0.7727, 0.8333 and 0.8030, respectively. The prompt consisted of five main parts, including character setting, scoring system setting, indicator setting, output setting, and explanatory information setting.

**Conclusion:** ChatGPT was effective in primary screening of participants with possible MCI. Improved standardization of prompts by professional clinicians would also improve the performance of the model. It is important to note that ChatGPT is not a substitute for a clinician making a diagnosis.

## Introduction

Alzheimer’s disease (AD) is the most common form of dementia, causeing abnormal mental decline, including thinking, memory, and language, and severely affecting the patient’s quality of life.^1^ AD has always been a major health hazard to society and a huge burden to the healthcare system, and this burden will even increase in the future.^2, 3^ There is an urgent need for human society to carry out prevention and intervention for AD. Although AD is progressive and incurable, early detection, diagnosis, and treatment can effectively delay its progression and thus significantly improve the quality of life of patient.^4^ Mild cognitive impairment (MCI) has been recognized as a strong predictor of AD and an early stage of AD, and early diagnosis and intervention of MCI will be of great help to human society in the fight against AD.^4–6^ However, neuropsychological testing for MCI is time-consuming and rigorous, and to accommodate the potentially large MCI population in society, a variety of validated brief cognitive tests, including language tests, memory tests, the Mini-Mental State Exam (MMSE) and the Montreal Cognitive Assessment (MoCA), are widely used for early screening of the disease.^7–9^ Developments in artificial intelligence (AI) are likely to make this process more accessible and allow a wider group of people to benefit.^10^

AI has emerged as a promising tool in healthcare, particularly in the area of cognitive impairment diagnosis.^11, 12^ AI can provide a more accurate and standardized process for diagnosing and predicting disease.^13^ Thabtah et al. conducted a comprehensive analysis of mobile-based AD and MCI screening apps, highlighting the potential of AI applications for early detection and diagnosis of dementia and exploring AI to improve access to healthcare services.^14^ Kalafatis et al. demonstrated the convergent validity of the Integrated Cognitive Assessment (ICA) using cognitive tests such as the MoCA. the AI model of the ICA was able to detect cognitive impairment with an AUC of 0.81 for MCI patients and 0.88 for mild AD patients.^15^ In contrast to custom-built AI models, we would like to see if large language models (LLMs) can be applied to this domain with good outcomes.^16, 17^

ChatGPT is a high-level LLM developed by OpenAI that is designed to generate human-like text based on the input it receives.^18^ GPT-4, the latest version available as of the September 2021 knowledge deadline, has a larger model size (in terms of parameters) and improved text generation capabilities than its predecessors.^19, 20^ The Generative Pretrained Transformer (GPT) model is a neural network architecture for understanding data sequences such as text. ChatGPT is “pre-trained” on a large corpus of text data, enabling it to understand and generate text that is not only grammatically correct, but also contextually relevant and coherent over a long period of time.^21^ With its powerful text analysis capabilities, we were ready to explore whether ChatGPT could be used for primary screening of MCI based on text conversation analysis under the supervision of professional neurologists.^22^ In the meantime, we would discuss the process and components of a standardized prompt design.^23^

It is essential to emphasize that ChatGPT is used only as a primary screening tool and has no diagnostic role.^24^ The ultimate goal of the study is to encourage more people with potential MCI to actively seek medical attention for early detection, early diagnosis and early treatment.

## Methods

### Inclusion and exclusion of participants

We included a total of 174 participants from the DementiaBank English Protocol Delaware Corpus^25^ and the DementiaBank English Pitt Corpus^6, 26^ (Figure 1).

**Figure 1.**
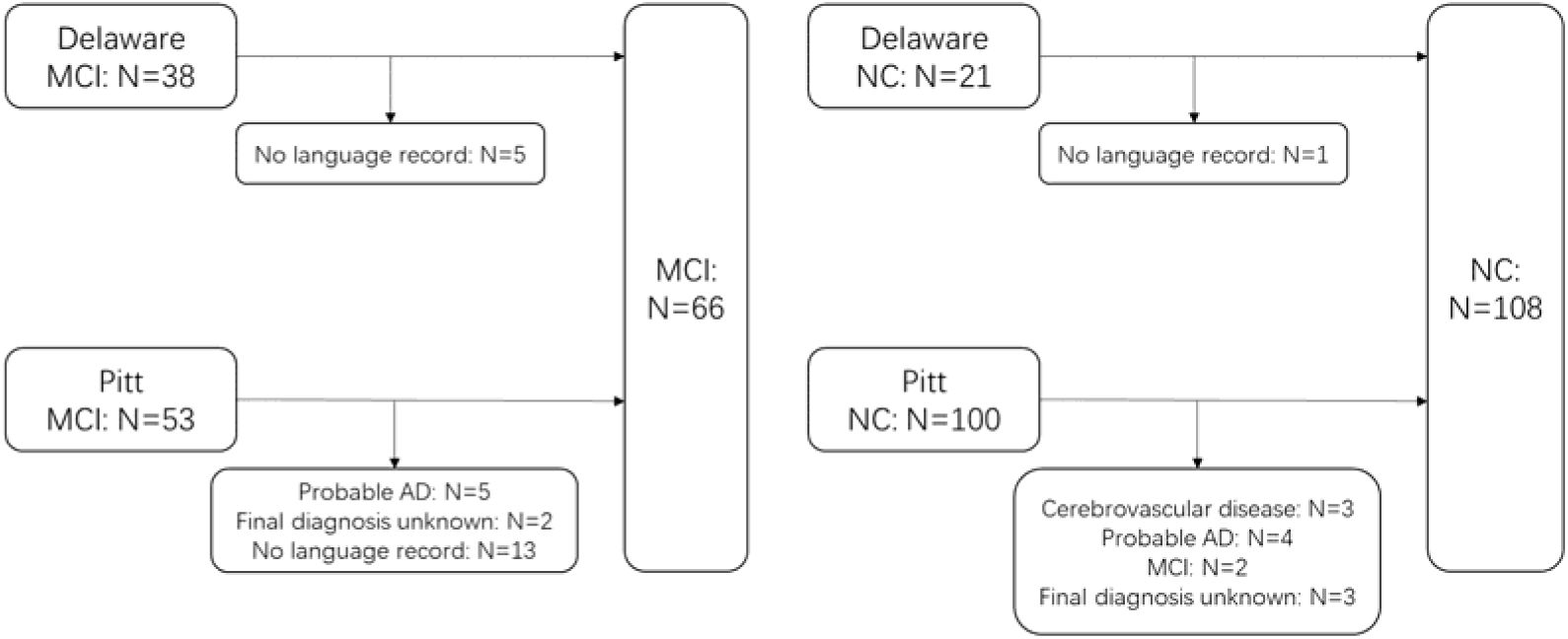
Participant inclusion and exclusion

A total of 66 MCI participants were included: in the Delaware database, we excluded 5 patients with no language record data and finally included 33 participants. In the Pitt database, we finally included 33 participants. Thirty of them were from the dementia group and three from the control group. The specific screening method was as follows: we screened 36 participants from the dementia group by initial diagnosis of MCI, then we excluded 5 participants with probable AD, 2 participants with unknown final diagnosis, 13 participants with no language record data of the patients. We also included 3 participants with a final diagnosis of MCI from the control group.

A total of 108 cognitively normal participants were included: in the Delaware database, we excluded 1 patient with no language record data and finally included 20 participants with 20 language records obtained. In the Pitt database, there were a total of 100 participants in the control group with language record data, and we finally included 88 participants. The specific screening method was as follows: we excluded controls with cerebrovascular disease because participants were finally diagnosed with Probable AD or MCI. Controls with a final diagnosis of unknown, controls with a final diagnosis of Probable AD, controls with a final diagnosis of MCI, and controls with a final diagnosis of cerebrovascular disease were also excluded.

In the end, 66 MCI participants and 108 normal participants were included. MCI mild cognitive impairment, NC normal cognition.

### Data cleaning and distribution

First, only the text dialogues “PARTICIPANT” and “INVESTIGATOR” were kept. Then each sentence was cleaned using the macro code (see Supplementary Information). Finally, a manual check was performed by Wang.

Random numbers were generated using the RAND and RANK functions to renumber all participants. The 70% of participants were set as the training set, with 44 MCI participants and 78 normal participants. The final 30% of participants were set as the test set, with 22 MCI participants and 30 normal participants. (See “Participant Information.xlsx” participant and random grouping sheet).

### Model Prompt Design

Model 1 prompt: We identified the basic indicators for analyzing language features based on published studies, including vocabulary features, syntax and grammar features, and semantics features in three main areas, with a total of seven indicators (Table 1).^27–30^

**Table 1.**
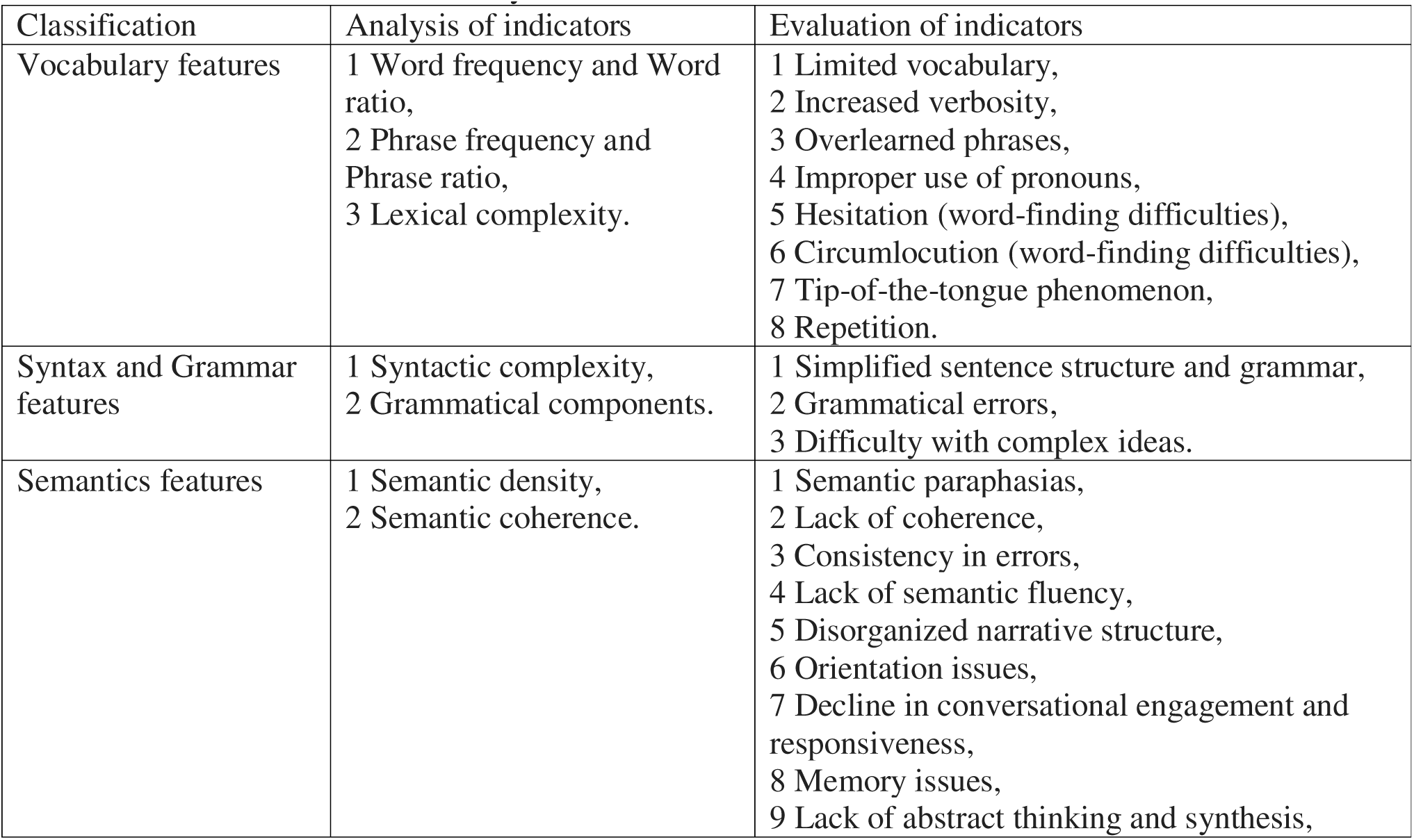
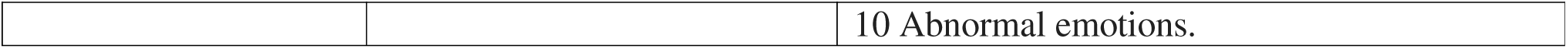
Analysis of indicators and evaluation of indicators

| Classification | Analysis of indicators | Evaluation of indicators |
| --- | --- | --- |
| Vocabulary features | 1 Word frequency and Word ratio,<br>2 Phrase frequency and Phrase ratio,<br>3 Lexical complexity. | 1 Limited vocabulary,<br>2 Increased verbosity,<br>3 Overlearned phrases,<br>4 Improper use of pronouns,<br>5 Hesitation (word-finding difficulties),<br>6 Circumlocution (word-finding difficulties),<br>7 Tip-of-the-tongue phenomenon,<br>8 Repetition. |
| Syntax and Grammar features | 1 Syntactic complexity,<br>2 Grammatical components. | 1 Simplified sentence structure and grammar,<br>2 Grammatical errors,<br>3 Difficulty with complex ideas. |
| Semantics features | 1 Semantic density,<br>2 Semantic coherence. | 1 Semantic paraphasias,<br>2 Lack of coherence,<br>3 Consistency in errors,<br>4 Lack of semantic fluency,<br>5 Disorganized narrative structure,<br>6 Orientation issues,<br>7 Decline in conversational engagement and responsiveness,<br>8 Memory issues,<br>9 Lack of abstract thinking and synthesis, |

Model 2 prompt: At the end of the Model 1 training, participants receive Feedback1, which included more detailed MCI features. This information was used to design the new prompt.

Model 3 prompt: We set a new prompt based on the Feedback 2 generated by model 2. Differently, we extracted 21 severity assessment indicators related to MCI and designed a scoring system (Table 1).

Model 4 prompt: We allowed GPT-4 to design 3 prompts for itself based on the Model 3 prompt and integrated them. This helped to compare the differences between the manual and robotic designs and standardize the format of the subsequent prompts.

Model 5 prompt: The 21 indicators obtained from GPT4-Model3 to assess the severity of MCI were analyzed using statistical methods to select more statistically significant indicators and to reduce bias. And the prompt format of Model 4 was then followed for the design.

All model prompts and feedbacks and detailed explanations of each indicator, are available in the Supplementary Information.

### Statistics Analysis

Descriptive statistics, including the calculation of mean, standard deviation, median, interquartile range, minimum and maximum values, were performed for each variable in each group. We then analyzed the normal distribution and the homogeneity of variance test of the data for the all variables. On the one hand, if the variables are normally distributed and have a uniform variance, a one-way ANOVA is used for the four groups, i.e. true positive (TP), false positive (FP), false negative (FN), and true negative (TN), and an independent samples t-test is used for the two groups, i.e. positive and negative. And then if there were differences in the results of the one-way ANOVA, a two-way comparison was performed using Tukey’s Honestly Significant Difference test as a post hoc test. On the other hand, if the variables are not normally distributed, the Kruskal-Wallis H-test is used for the four groups, and the Mann-Whitney U test was used for the two groups. And then if there were differences in the results of the Kruskal-Wallis H-test, Dunn’s test was applied as a post hoc test. (See “Participant Information.xlsx” Variable_selection_data, and Variable_selection_data (2) Sheet for all variables). Finally, the visualization of violin plots was used to compare the degree of concentration and dispersion of each variable.

Sensitivity (SEN), specificity (SPE), positive predictive value (PPV), negative predictive value (NPV), positive likelihood ratios (PLR), negative likelihood ratios (NLR), accuracy (ACC), and receiver operating characteristics (ROC) curve and area under the curve (AUC) of the different models were compared (see “Participant Information.xlsx” Training_GPT4, Training_GPT3.5, and Testing Sheet for all variables). Analysis was performed using R 4.3.0. All R codes can be found in the Supplementary Information.

## Results

### Inclusion of variables

In addition to the 8 analytic indicators that had been explicitly included in the model 5 prompts, we included 3 evaluative indicators in the final prompt. These 3 indicators were generated from 21 indicators (Table 1) that assess the severity of MCI, and they were effective in discriminating between MCI and cognitively normal participants, namely tip-of-the-tongue phenomenon (P < 0.001), difficulty with complex ideas (P < 0.001), and memory problems (P < 0.001). (Figure 2). Although most of the indicators were effective in distinguishing FP-FN, TP-FN, TN-FP, and TP-TN, only the tip-of-the-tongue phenomenon (p-value without adjustment for multiple comparisons = 0.032, and p-value with adjustment for multiple comparisons = 0.035), difficulty with complex ideas (p-value without adjustment for multiple comparisons = 0.037, and p-value with adjustment for multiple comparisons = 0.012), and memory problem (p-value without adjustment for multiple comparisons = 0.066, and p-value with adjustment for multiple comparisons = 0.034) could effectively discriminate TP-FP (Figure 2). The statistical analysis of all indicators can be found in the Supplementary Information.

**Figure 2.**
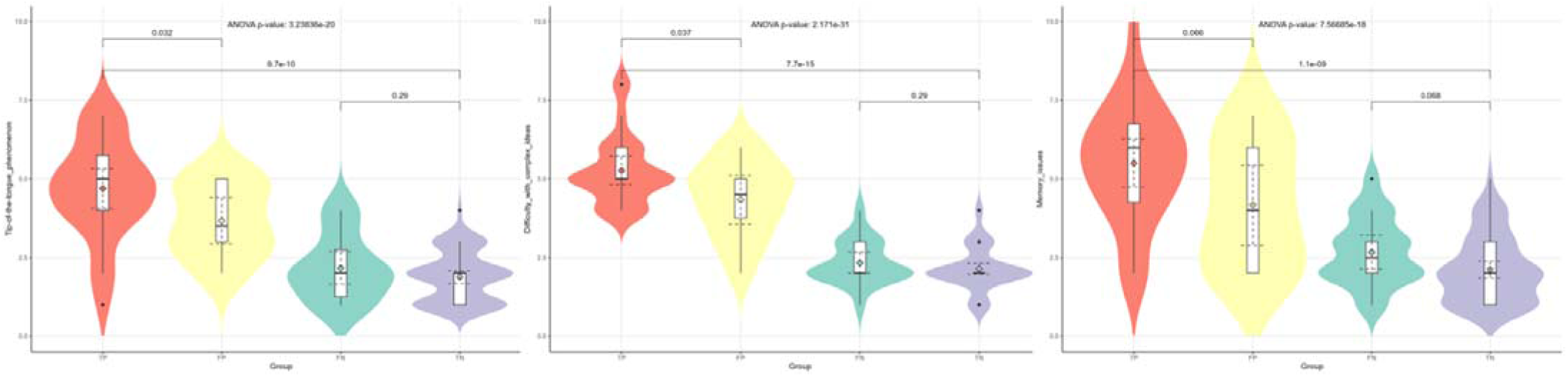
The violin plots of the selected evaluation metrics

Tip-of-the-tongue phenomenon, difficulty with complex ideas, and memory problems, three metrics for evaluating participants’ cognitive functioning, were included in the prompt of model 5. The violin plots include P-values from one-way ANOVA, and P-values from Tukey’s honestly significant difference test without adjustment for multiple comparisons.

### Model Comparison

We found that ChatGPT had acceptable performance in screening for MCI by text analysis alone (Table 2 and Figure 3). On the training set, the SEN, SPE, and AUC of GPT4_Model5 reached 0.8636, 0.9487, and 0.9062, respectively; on the test set, the SEN, SPE, and AUC reached 0.7727, 0.8333, and 0.8030, respectively. The prompt of model5 consisted of five main parts, including character setting, scoring system setting, indicator setting, output setting, and explanatory information setting.

**Figure 3.**
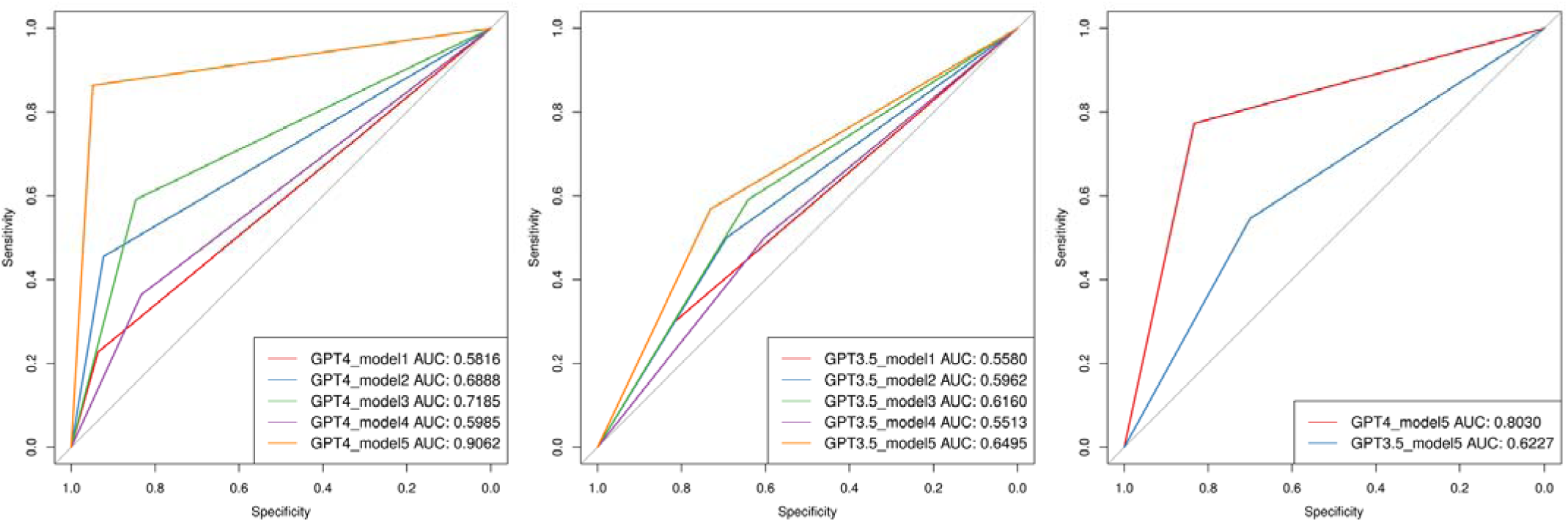
ROC curve

**Table 2.** Diagnostic evaluation indicators

| Dataset | model | TP | FN | FP | TN | SEN | SPE | PPV | NPV | PLR | NLR | Accuracy |
| --- | --- | --- | --- | --- | --- | --- | --- | --- | --- | --- | --- | --- |
| Training set | GPT4_model1 | 10 | 34 | 5 | 73 | 0.2273 | 0.9359 | 0.6667 | 0.6822 | 3.5455 | 0.8257 | 0.6803 |
| Training set | GPT4_model2 | 20 | 24 | 6 | 72 | 0.4545 | 0.9231 | 0.7692 | 0.7500 | 5.9091 | 0.5909 | 0.7541 |
| Training set | GPT4_model3 | 26 | 18 | 12 | 66 | 0.5909 | 0.8462 | 0.6842 | 0.7857 | 3.8409 | 0.4835 | 0.7541 |
| Training set | GPT4_model4 | 16 | 28 | 13 | 65 | 0.3636 | 0.8333 | 0.5517 | 0.6989 | 2.1818 | 0.7636 | 0.6639 |
| Training set | GPT4_model5 | 38 | 6 | 4 | 74 | 0.8636 | 0.9487 | 0.9048 | 0.9250 | 16.8409 | 0.1437 | 0.9180 |
| Training set | GPT3.5_model1 | 13 | 31 | 14 | 64 | 0.2955 | 0.8205 | 0.4815 | 0.6737 | 1.6461 | 0.8587 | 0.6311 |
| Training set | GPT3.5_model2 | 22 | 22 | 24 | 54 | 0.5000 | 0.6923 | 0.4783 | 0.7105 | 1.6250 | 0.7222 | 0.6230 |
| Training set | GPT3.5_model3 | 26 | 18 | 28 | 50 | 0.5909 | 0.6410 | 0.4815 | 0.7353 | 1.6461 | 0.6382 | 0.6230 |
| Training set | GPT3.5_model4 | 22 | 22 | 31 | 47 | 0.5000 | 0.6026 | 0.4151 | 0.6812 | 1.2581 | 0.8298 | 0.5656 |
| Training set | GPT3.5_model5 | 25 | 19 | 21 | 57 | 0.5682 | 0.7308 | 0.5435 | 0.7500 | 2.1104 | 0.5909 | 0.6721 |
| Test set | GPT4_model5 | 17 | 5 | 5 | 25 | 0.7727 | 0.8333 | 0.7727 | 0.8333 | 4.6364 | 0.2727 | 0.8077 |
| Test set | GPT3.5_model5 | 12 | 10 | 9 | 21 | 0.5455 | 0.7000 | 0.5714 | 0.6774 | 1.8182 | 0.6494 | 0.6346 |

By comparing model 1, model 2 and model 3 (GPT4-AUC: 0.5816, 0.6888, and 0.7185, respectively; GPT3.5-AUC: 0.5580, 0.5962, and 0.6160, respectively), we found that providing ChatGPT with more detailed prompts would effectively improve its ability to screen for MCI.

And comparing model 3 and model 4 (GPT4-AUC: 0.7185, and 0.5985, respectively; GPT3.5-AUC: 0.6160, and 0.5513, respectively) showed that the physician-designed prompt was superior to the ChatGPT-designed prompt.

The analysis and screening of the included indicators would effectively improve the performance of the model, as shown by the comparison of model 3 and model 5 (GPT4-AUC: 0.7185, and 0.9062, respectively; GPT3.5-AUC: 0.6160, and 0.6495 respectively).

In addition, we verified that GPT4 outperformed GPT3.5 in terms of logical ability, which was reflected in the following two points: firstly, the AUC of GPT4 was consistently higher than that of GPT3.5 under the same model in the same data set; secondly, the improvement in prompt was more significant for the performance improvement of GPT4 compared to GPT3.5.

In the training set, the final model 5 achieves an AUC of 0.9062 in GPT-4 and 0.6459 in GPT-3.5. Model 5 outperforms models 1 through 4 in both GPT-4 and GPT3.5. In the test set, model 5 achieves an AUC of 0.8030 in GPT-4 and 0.6227.

## Discussion

### ChatGPT based on language analysis

It is critical to note that the diagnosis of MCI is often based on a variety of factors, including clinical observations, cognitive testing, and reports from individuals and their families about their daily functioning. Language analysis can provide useful information, but is only one piece of the puzzle.^31^ In general, people with MCI may show subtle differences in language use compared to people with normal cognitive status. some possible indications of MCI in language use may include 8 lexical features: limited vocabulary, increased verbosity, overlearned phrases, incorrect use of pronouns, hesitation (word-finding difficulties), circumlocution (word-finding difficulties), tip-of-the-tongue phenomenon, and repetition; 3 syntax and grammar features: simplified sentence structure and grammar, grammatical errors, and difficulty with complex ideas; and 10 semantic features: semantic paraphasias, lack of coherence, consistency in errors, lack of semantic fluency, disorganized narrative structure, orientation issues, decline in conversational engagement and responsiveness, memory issues, lack of abstract thinking and synthesis, and abnormal emotions. Among them, the tip-of-the-tongue phenomenon, difficulty with complex ideas, and memory issues deserve particular attention.

It is important to emphasize that ChatGPT based on linguistic analysis cannot be used to diagnose MCI, which must be done by a specialist. ChatGPT is only used as a primary screening of subjects in the hope of detecting more potential MCI language features. This will help to identify the patient at an early stage and assist the physician in the assessment.

### Prompt Design

At this stage, the prompts designed by professional physicians are still better than those designed by ChatGPT itself, comparing model 4 with model 3 and model 5. This is probably because ChatGPT cannot effectively decide which indicators are more important for decision making.

The prompt can be designed in four steps. In the first step, high quality literature, preferably high quality systematic reviews, was found and proven valid indicators were extracted and added to the prompts with interpretation of these indicators. In the second step, ChatGPT was required to output the results by analyzing these indicators, while interpreting the association between each patient’s symptoms and the indicators. The new explanatory information was collected, organized, reviewed and finally added to the prompt. In the third step, the indicators were scored and these scores were statistically analyzed to select those that better discriminated between the experimental and control groups. Finally, by standardizing the format of the prompt, the indicators selected in step three were incorporated and the information collected in step two was used to interpret them.

The prompt consists of five main parts (Figure 4). The first part is character setting, i.e., designing a role for the ChatGPT. In this research, ChatGPT was set up as a physician’s assistant who, by analyzing indicators, would first classify the participants into the probable MCI group and the probable normal cognitive function group, and provide detailed reasons for doing so, in order to facilitate the diagnosis by a professional physician. The second part is the establishment scoring system. Its purpose is to quantify the narrative indicators to facilitate subsequent statistical analysis. The third part is the definition of indicators. Indicators are divided into those included based on the literature and those included based on the analysis. There is a need to ensure the quality rather than the quantity of the included indicators. The fourth part is output setting. This part requires matching with the first part. The fifth part is the explanatory information setting. Providing valid explanatory information derived from the literature or analysis for each indicator could improve the performance of the models.

**Figure 4.**
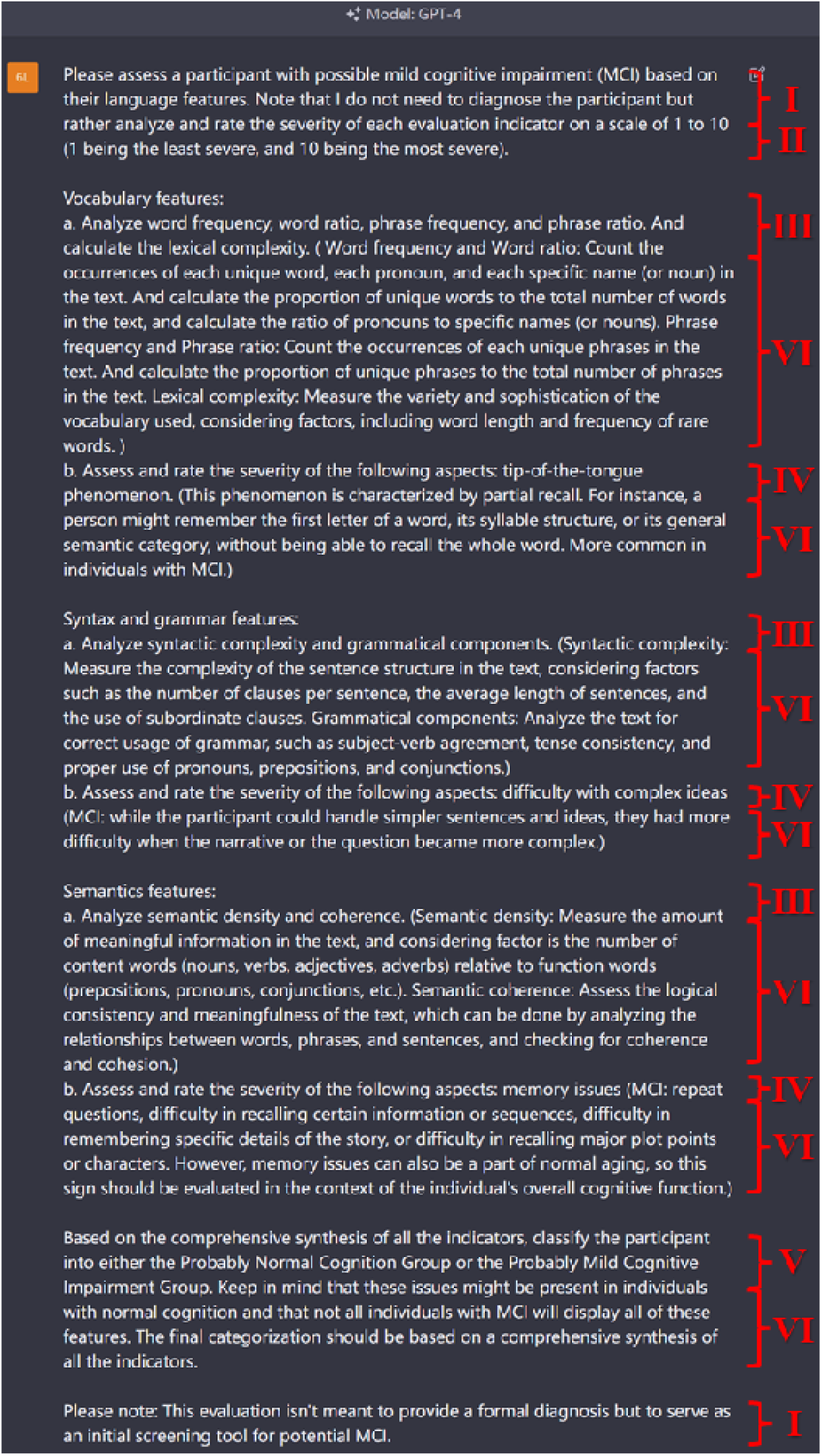
Prompt Design

The prompt consists of 5 main parts. The character setting is shown as LJ. The scoring system setting is shown as LJ. The indicator settings are shown as LJ and LJ. Indicators include those included based on literature (i.e. shown as LJ) and those included based on analysis (i.e. shown as LJ). The output setting is shown as LJ. The explanatory information settings are shown as LJ.

## Limitation

First, we found no valid indicators to distinguish TN-FN, which increased the likelihood that ChatGPT would make an error in assigning participants to the potentially cognitively normal group. Secondly, we only compared GPT4 with GPT3.5. Although the prompts were reviewed by expert clinicians and the model performed well statistically, there was still no guarantee that ChatGPT could be used as a tool for primary screening for MCI. In a follow-up study, we intend to compare GPT4 with clinicians at different levels to determine its clinical role in primary screening. Thirdly, our test dataset used data that had never been trained, all of which had to be requested and some of which were only published in 2022,^25^ but there was still no guarantee that ChatGPT had not learned them, which is a problem to be faced in the era of large models. For this reason, we intend to include information on patients from our institution to ensure that the data are completely unknown to ChatGPT.

## Conclusion

In both the training and test sets, ChatGPT could effectively discriminate participants with possible MCI. Meanwhile, the standardised improvement of prompts by professional physicians would improve the performance of the model. It should be noted, however, that the use of ChatGPT must follow medical ethics and cannot replace doctors in diagnosis. Through the study, we hope to screen a large number of people who may have MCI and help them to go to hospital for diagnosis, so that ‘early detection, early diagnosis and early treatment’ can be achieved, delaying or even preventing the progression of MCI to AD.

## Statement

The article was written entirely by human. All authors are responsible and accountable for the originality, accuracy, and integrity of the work. Information that could lead to the identification of patients has been removed from the manuscript and supplementary information. Readers can contact the corresponding author to request access to this data.

## Supporting information

Data(GPT4_vs_GPT3.5_model1-5)

Participant Information

## Data Availability

All data produced in the present study are available upon reasonable request to the authors

