## Supplementary Information for "Text dialogue analysis Based ChatGPT for Primary Screening of Mild Cognitive Impairment"

Catalog

**Macro code**	**2**

**Model prompts**	**11**

**Model1 prompt**	11

**Model2 prompt**	13

**Model3 prompt**	15

**Model4 prompt**	18

**Model5 prompt**	23

**Feedback**	**25**

**Feedback1**	25

**Feedback2**	26

**R codes**	**36**

**R code 1**	36

**R code 2**	40

**R code 3**	42

**Statistical analysis**	**45**

**Statistical analysis 1: mean, standard deviation, median, interquartile spacing, minimum, maximum values**	45

**Statistical analysis 2: normal distribution, Homogeneity of variance test, Independent samples t-test for the two groups, Mann-Whitney U test for the two groups**	48

**Statistical analysis 3 and Violin plots: one-way ANOVA for the four groups (Tukey's Honestly Significant Difference test as a post hoc test), and Kruskal-Wallis H-test for the four groups (Dunn's test was applied as a post hoc test)**	49

**Macro code**: Use the macro code to clean up each dialogue of "PARTICIPANT" and "INVESTIGATOR".

Sub macro 1()

'

' macro 1 macro

'

'

Selection.Find.ClearFormatting

Selection.Find.Replacement.ClearFormatting

With Selection.Find

.Text = "^#"

.Replacement.Text = ""

.Forward = True

.Wrap = wdFindContinue

.Format = False

.MatchCase = False

.MatchWholeWord = False

.MatchByte = True

.MatchWildcards = False

.MatchSoundsLike = False

.MatchAllWordForms = False

End With

Selection.Find.Execute Replace:=wdReplaceAll

With Selection.Find

.Text = Chr(21) & "_" & Chr(21)

.Replacement.Text = ""

.Forward = True

.Wrap = wdFindContinue

.Format = False

.MatchCase = False

.MatchWholeWord = False

.MatchByte = True

.MatchWildcards = False

.MatchSoundsLike = False

.MatchAllWordForms = False

End With

Selection.Find.Execute Replace:=wdReplaceAll

With Selection.Find

.Text = " _ "

.Replacement.Text = ""

.Forward = True

.Wrap = wdFindContinue

.Format = False

.MatchCase = False

.MatchWholeWord = False

.MatchByte = True

.MatchWildcards = False

.MatchSoundsLike = False

.MatchAllWordForms = False

End With

Selection.Find.Execute Replace:=wdReplaceAll

With Selection.Find

.Text = ChrW(8225)

.Replacement.Text = ""

.Forward = True

.Wrap = wdFindContinue

.Format = False

.MatchCase = False

.MatchWholeWord = False

.MatchByte = True

.MatchWildcards = False

.MatchSoundsLike = False

.MatchAllWordForms = False

End With

Selection.Find.Execute Replace:=wdReplaceAll

With Selection.Find

.Text = "+//"

.Replacement.Text = ""

.Forward = True

.Wrap = wdFindContinue

.Format = False

.MatchCase = False

.MatchWholeWord = False

.MatchByte = True

.MatchWildcards = False

.MatchSoundsLike = False

.MatchAllWordForms = False

End With

Selection.Find.Execute Replace:=wdReplaceAll

With Selection.Find

.Text = "+/"

.Replacement.Text = ""

.Forward = True

.Wrap = wdFindContinue

.Format = False

.MatchCase = False

.MatchWholeWord = False

.MatchByte = True

.MatchWildcards = False

.MatchSoundsLike = False

.MatchAllWordForms = False

End With

Selection.Find.Execute Replace:=wdReplaceAll

With Selection.Find

.Text = "+<"

.Replacement.Text = ""

.Forward = True

.Wrap = wdFindContinue

.Format = False

.MatchCase = False

.MatchWholeWord = False

.MatchByte = True

.MatchWildcards = False

.MatchSoundsLike = False

.MatchAllWordForms = False

End With

Selection.Find.Execute Replace:=wdReplaceAll

With Selection.Find

.Text = "+""/"

.Replacement.Text = ""

.Forward = True

.Wrap = wdFindContinue

.Format = False

.MatchCase = False

.MatchWholeWord = False

.MatchByte = True

.MatchWildcards = False

.MatchSoundsLike = False

.MatchAllWordForms = False

End With

Selection.Find.Execute Replace:=wdReplaceAll

With Selection.Find

.Text = "&=sighs"

.Replacement.Text = ""

.Forward = True

.Wrap = wdFindContinue

.Format = False

.MatchCase = False

.MatchWholeWord = False

.MatchByte = True

.MatchWildcards = False

.MatchSoundsLike = False

.MatchAllWordForms = False

End With

Selection.Find.Execute Replace:=wdReplaceAll

With Selection.Find

.Text = "&=coughs"

.Replacement.Text = ""

.Forward = True

.Wrap = wdFindContinue

.Format = False

.MatchCase = False

.MatchWholeWord = False

.MatchByte = True

.MatchWildcards = False

.MatchSoundsLike = False

.MatchAllWordForms = False

End With

Selection.Find.Execute Replace:=wdReplaceAll

With Selection.Find

.Text = "&=laughs"

.Replacement.Text = ""

.Forward = True

.Wrap = wdFindContinue

.Format = False

.MatchCase = False

.MatchWholeWord = False

.MatchByte = True

.MatchWildcards = False

.MatchSoundsLike = False

.MatchAllWordForms = False

End With

Selection.Find.Execute Replace:=wdReplaceAll

With Selection.Find

.Text = "[+ exc]"

.Replacement.Text = ""

.Forward = True

.Wrap = wdFindContinue

.Format = False

.MatchCase = False

.MatchWholeWord = False

.MatchByte = True

.MatchWildcards = False

.MatchSoundsLike = False

.MatchAllWordForms = False

End With

Selection.Find.Execute Replace:=wdReplaceAll

With Selection.Find

.Text = "[+ gram]"

.Replacement.Text = ""

.Forward = True

.Wrap = wdFindContinue

.Format = False

.MatchCase = False

.MatchWholeWord = False

.MatchByte = True

.MatchWildcards = False

.MatchSoundsLike = False

.MatchAllWordForms = False

End With

Selection.Find.Execute Replace:=wdReplaceAll

With Selection.Find

.Text = "*INV"

.Replacement.Text = "INVESTIGATOR"

.Forward = True

.Wrap = wdFindContinue

.Format = False

.MatchCase = False

.MatchWholeWord = False

.MatchByte = True

.MatchWildcards = False

.MatchSoundsLike = False

.MatchAllWordForms = False

End With

Selection.Find.Execute Replace:=wdReplaceAll

With Selection.Find

.Text = "*PAR"

.Replacement.Text = "PARTICIPANT"

.Forward = True

.Wrap = wdFindContinue

.Format = False

.MatchCase = False

.MatchWholeWord = False

.MatchByte = True

.MatchWildcards = False

.MatchSoundsLike = False

.MatchAllWordForms = False

End With

Selection.Find.Execute Replace:=wdReplaceAll

With Selection.Find

.Text = "[//]"

.Replacement.Text = "…"

.Forward = True

.Wrap = wdFindContinue

.Format = False

.MatchCase = False

.MatchWholeWord = False

.MatchByte = True

.MatchWildcards = False

.MatchSoundsLike = False

.MatchAllWordForms = False

End With

Selection.Find.Execute Replace:=wdReplaceAll

With Selection.Find

.Text = "[/]"

.Replacement.Text = "…"

.Forward = True

.Wrap = wdFindContinue

.Format = False

.MatchCase = False

.MatchWholeWord = False

.MatchByte = True

.MatchWildcards = False

.MatchSoundsLike = False

.MatchAllWordForms = False

End With

Selection.Find.Execute Replace:=wdReplaceAll

With Selection.Find

.Text = "&+"

.Replacement.Text = "…"

.Forward = True

.Wrap = wdFindContinue

.Format = False

.MatchCase = False

.MatchWholeWord = False

.MatchByte = True

.MatchWildcards = False

.MatchSoundsLike = False

.MatchAllWordForms = False

End With

Selection.Find.Execute Replace:=wdReplaceAll

With Selection.Find

.Text = "&-"

.Replacement.Text = "…"

.Forward = True

.Wrap = wdFindContinue

.Format = False

.MatchCase = False

.MatchWholeWord = False

.MatchByte = True

.MatchWildcards = False

.MatchSoundsLike = False

.MatchAllWordForms = False

End With

Selection.Find.Execute Replace:=wdReplaceAll

With Selection.Find

.Text = "&~"

.Replacement.Text = "…"

.Forward = True

.Wrap = wdFindContinue

.Format = False

.MatchCase = False

.MatchWholeWord = False

.MatchByte = True

.MatchWildcards = False

.MatchSoundsLike = False

.MatchAllWordForms = False

End With

Selection.Find.Execute Replace:=wdReplaceAll

With Selection.Find

.Text = "+"

.Replacement.Text = ""

.Forward = True

.Wrap = wdFindContinue

.Format = False

.MatchCase = False

.MatchWholeWord = False

.MatchByte = True

.MatchWildcards = False

.MatchSoundsLike = False

.MatchAllWordForms = False

End With

Selection.Find.Execute Replace:=wdReplaceAll

With Selection.Find

.Text = "(.)"

.Replacement.Text = ""

.Forward = True

.Wrap = wdFindContinue

.Format = False

.MatchCase = False

.MatchWholeWord = False

.MatchByte = True

.MatchWildcards = False

.MatchSoundsLike = False

.MatchAllWordForms = False

End With

Selection.Find.Execute Replace:=wdReplaceAll

With Selection.Find

.Text = "(..)"

.Replacement.Text = ""

.Forward = True

.Wrap = wdFindContinue

.Format = False

.MatchCase = False

.MatchWholeWord = False

.MatchByte = True

.MatchWildcards = False

.MatchSoundsLike = False

.MatchAllWordForms = False

End With

Selection.Find.Execute Replace:=wdReplaceAll

With Selection.Find

.Text = "(...)"

.Replacement.Text = "…"

.Forward = True

.Wrap = wdFindContinue

.Format = False

.MatchCase = False

.MatchWholeWord = False

.MatchByte = True

.MatchWildcards = False

.MatchSoundsLike = False

.MatchAllWordForms = False

End With

Selection.Find.Execute Replace:=wdReplaceAll

With Selection.Find

.Text = "&=clears:throat"

.Replacement.Text = ""

.Forward = True

.Wrap = wdFindContinue

.Format = False

.MatchCase = False

.MatchWholeWord = False

.MatchByte = True

.MatchWildcards = False

.MatchSoundsLike = False

.MatchAllWordForms = False

End With

Selection.Find.Execute Replace:=wdReplaceAll

End Sub

**Model1 prompt:** Design based on published studies

It is important to stress that you do not need a diagnosis, only an initial screening. That is, I would like you to be assessed through the conversation between PARTICIPANT and INVESTIGATOR below.

Assessment requirements:

Firstly, please assess the Vocabulary features of PARTICIPANT at the Word frequency, Word ratio, Phrase ratio, Difficulty finding the right words, Lexical complexity, Limited vocabulary.

Word frequency: Count the occurrences of each unique word in the text. Unique words are distinct words that appear in the text, not taking into account their repetitions. Each word is counted only once, regardless of how many times it occurs.

Word ratio: Calculate the proportion of unique words to the total number of words in the text.

Phrase frequency: Count the occurrences of each unique phrases in the text. Unique phrases are distinct combinations of words that appear in the text, not taking into account their repetitions. Each phrase is counted only once, regardless of how many times it occurs.

Phrase ratio: Calculate the proportion of unique phrases to the total number of phrases in the text.

Difficulty finding the right words: Frequent pauses or use of filler words, like "uh" or "um," as the participant searches for the desired word.

Lexical complexity: Measure the variety and sophistication of the vocabulary used, considering factors such as word length and frequency of rare words.

Limited vocabulary: Repeated use of the same words or simple words, with a lack of variety or sophistication.

Secondly, please assess the Semantics features of PARTICIPANT, from Semantic density, Inability to convey meaning, Semantic coherence, Lack of coherence.

Semantic density: Measure the amount of meaningful information in the text, considering factors such as the number of content words (nouns, verbs, adjectives, adverbs) relative to function words (prepositions, pronouns, conjunctions, etc.).

Inability to convey meaning: Difficulty in expressing ideas clearly, leading to confusion or misunderstanding.

Semantic coherence: Assess the logical consistency and meaningfulness of the text. This can be done by analyzing the relationships between words, phrases, and sentences, and checking for coherence and cohesion.

Lack of coherence: Difficulty in maintaining a logical flow of ideas, with thoughts appearing disjointed or unrelated.

Thirdly, please assess the Syntax and Grammar features of PARTICIPANT, from Syntactic complexity, Simplified sentence structure, Grammatical components, Grammatical errors.

Syntactic complexity: Measure the complexity of the sentence structure in the text, considering factors such as the number of clauses per sentence, the average length of sentences, and the use of subordinate clauses.

Simplified sentence structure: Predominantly short, simple sentences or a lack of complex sentence structures.

Grammatical components: Analyze the text for correct usage of grammar, such as subject-verb agreement, tense consistency, and proper use of pronouns, prepositions, and conjunctions.

Grammatical errors: Frequent mistakes in verb tense, subject-verb agreement, or other aspects of grammar.

Finally, please assign PARTICIPANT to the Probably Normal group or the Probably Mild Cognitive Impairment group, only.

Mild cognitive impairment is an early stage of memory loss or other cognitive ability loss (such as language or visual/spatial perception) in individuals who maintain the ability to independently perform most activities of daily living. I know that it is very difficult to distinguish mild cognitive impairment from normal status by language alone. I hope you will be able to group boldly by subtle language gaps.

**Model2 prompt:** Re-design based on Feedback1

Attention! You do not need to diagnose, just follow the steps below to initially screen for PARTICIPANT with possibly mild cognitive impairment (MCI). Some differences between MCI and normal cognition are also given, please analyse them in detail, step by step.

Firstly, please assess the Vocabulary features of PARTICIPANT at the Word frequency and Word ratio, Phrase frequency and Phrase ratio, Word-finding difficulties, Repetition, Lexical complexity, Limited vocabulary.

Word frequency: Count the occurrences of each unique word in the text. Unique words are distinct words that appear in the text, not taking into account their repetitions. Each word is counted only once, regardless of how many times it occurs.

Word ratio: Calculate the proportion of unique words to the total number of words in the text.

Phrase frequency: Count the occurrences of each unique phrases in the text. Unique phrases are distinct combinations of words that appear in the text, not taking into account their repetitions.

Phrase ratio: Calculate the proportion of unique phrases to the total number of phrases in the text.

Word-finding difficulties: Frequent pauses or use of filler words, like "uh" or "um,". MCI: more frequent instances, longer time to find the right word, or using more general terms. Normal: fewer instances, and better lexical retrieval.

Repetition: The participant repeats certain phrases or ideas a few times throughout the conversation. MCI: more frequent repetition. Normal: less frequent repetition (some repetition of words but not excessive) and better memory of what has been mentioned.

Lexical complexity: Measure the variety and sophistication of the vocabulary used, considering factors, including word length and frequency of rare words.

Limited vocabulary: Repeated use of the same words or simple words. MCI: slightly reduced vocabulary, and more frequent use of simpler words or common terms; Normal: richer vocabulary (low limitation).

Secondly, please assess the Semantics features of PARTICIPANT, from Semantic density and Inability to convey meaning, Semantic coherence and Lack of coherence.

Semantic density: Measure the amount of meaningful information in the text, and considering factor is the number of content words (nouns, verbs, adjectives, adverbs) relative to function words (prepositions, pronouns, conjunctions, etc.).

Inability to convey meaning: Difficulty in expressing ideas clearly, leading to confusion or misunderstanding. Normal: the participant was able to describe the scene clearly.

Semantic coherence: Assess the logical consistency and meaningfulness of the text, which can be done by analyzing the relationships between words, phrases, and sentences, and checking for coherence and cohesion.

Lack of coherence: Difficulty in maintaining a logical flow of ideas, with thoughts appearing disjointed or unrelated. MCI: struggle with organizing thoughts coherently or maintaining a clear focus in conversation; Normal: greater coherence and organization in their storytelling and conversations.

Thirdly, please assess the Syntax and Grammar features of PARTICIPANT, from Syntactic complexity and Simplified sentence structure, Grammatical components and Grammatical errors.

Syntactic complexity: Measure the complexity of the sentence structure in the text, considering factors such as the number of clauses per sentence, the average length of sentences, and the use of subordinate clauses.

Simplified sentence structure: A lack of complex sentence structures. MCI: simpler sentence structures; Normal: in longer dialogues there is a greater variety of sentence structures (moderately complex), but in short dialogues the sentences may turn out to be simple.

Grammatical components: Analyze the text for correct usage of grammar, such as subject-verb agreement, tense consistency, and proper use of pronouns, prepositions, and conjunctions. MCI: avoidance of complex grammatical constructions; Normal: better use of complex grammatical constructions.

Grammatical errors: Frequent mistakes in verb tense, subject-verb agreement, or other aspects of grammar. MCI: more frequent grammatical errors, such as mistakes in verb tense and subject-verb agreement; Normal: fewer grammatical errors (a few errors, but not significant).

Finally, Please dichotomise PARTICIPANT into the Probably Normal group or the Probably Mild Cognitive Impairment group. PARTICIPANT with MCI might exhibit more difficulty with vocabulary, sentence structure, repetition, word-finding, coherence, and grammar. However, it is not definitive and requires a comprehensive judgement.

**Model3 prompt**: Re-design based on Feedback2

You do not need to diagnose, just the analysis indicators and evaluation indicators below to initially screen for PARTICIPANT with possibly mild cognitive impairment (MCI). Also, please rate the severity of each evaluation indicator on a scale from 1 to 10 for the PARTICIPANT. (1 as the least severe and 10 as the most severe).

Firstly, please analyse the Vocabulary features of PARTICIPANT, including Word frequency and Word ratio, Phrase frequency and Phrase ratio, and Lexical complexity. Then assess the following aspects based on Vocabulary features: 1 Limited vocabulary, 2 Increased verbosity, 3 Overlearned phrases, 4 Improper use of pronouns, 5 Hesitation (Word-finding difficulties), 6 Circumlocution (Word-finding difficulties), 7 Tip-of-the-tongue phenomenon, and 8 Repetition.

Word frequency and Word ratio: Count the occurrences of each unique word, each pronoun, and each specific name (or noun) in the text. And calculate the proportion of unique words to the total number of words in the text, and calculate the ratio of pronouns to specific names (or nouns).

Phrase frequency and Phrase ratio: Count the occurrences of each unique phrases in the text. And calculate the proportion of unique phrases to the total number of phrases in the text.

Lexical complexity: Measure the variety and sophistication of the vocabulary used, considering factors, including word length and frequency of rare words.

Limited vocabulary: MCI: slightly reduced vocabulary, and more frequent use of the same words, simple words, or common terms; Normal: richer vocabulary.

Increased verbosity: MCI: more verbose and using more words than necessary.

Overlearned phrases: MCI: overuse of cliches or stock phrases. Normal: usually demonstrate more variety in their language use.

Improper use of pronouns: MCI: a tendency to use more pronouns than specific names (or nouns), or difficulty with pronouns (mixing up "he" and "she" or "I" and "you").

Hesitation (Word-finding difficulties): Frequent pauses or use of filler words, like "uh", "um" or "…". MCI: more disfluencies, longer time to find the right word, or using more filler words. Normal: fluent speech with fewer filled and unfilled pauses.

Circumlocution (Word-finding difficulties): MCI: talking in a roundabout way due to difficulty finding the specific word; using many words where fewer would do, especially in a deliberate attempt to be vague or evasive; using a word close to, but not quite the intended word; or substitute more general words for more specific ones.

Tip-of-the-tongue phenomenon: this phenomenon is characterized by partial recall. For instance, a person might remember the first letter of a word, its syllable structure, or its general semantic category, without being able to recall the whole word. More common in individuals with MCI.

Repetition: MCI: repeat the same stories, questions, or statements within a short span of time. Normal: some repetition of words but not excessive.

Secondly, please analyse the Syntax and Grammar features, including Syntactic complexity, and Grammatical components. Then assess the following aspects based on Syntax and Grammar features: 1 Simplified sentence structure and grammar, 2 Grammatical errors, and 3 Difficulty with complex ideas.

Syntactic complexity: Measure the complexity of the sentence structure in the text, considering factors such as the number of clauses per sentence, the average length of sentences, and the use of subordinate clauses.

Grammatical components: Analyze the text for correct usage of grammar, such as subject-verb agreement, tense consistency, and proper use of pronouns, prepositions, and conjunctions.

Simplified sentence structure and grammar: MCI: preference for simpler structures in both long and short sentences, and avoidance of complex grammatical constructions; Normal: in long dialogues there is a greater variety of sentence structures (have a balanced usage of simple and complex sentences) and better use of complex grammatical constructions; but in short dialogues the sentences and grammar may turn out to be simple.

Grammatical errors: MCI: more frequent grammatical errors, such as mistakes in verb tense and subject-verb agreement; Normal: fewer grammatical errors (a few errors, but not significant).

Difficulty with complex ideas: MCI: while the participant could handle simpler sentences and ideas, they had more difficulty when the narrative or the question became more complex.

Thirdly, please analyse the Semantics features of PARTICIPANT, including Semantic density, and Semantic coherence. Then assess the following aspects according to the Semantics features: 1 Semantic paraphasias, 2 Lack of coherence, 3 Consistency in errors, 4 Lack of semantic fluency, 5 Disorganized narrative structure, 6 Orientation issues, 7 Decline in conversational engagement and responsiveness, 8 Memory issues, 9 Lack of abstract thinking and synthesis, and 10 Abnormal emotions.

Semantic density: Measure the amount of meaningful information in the text, and considering factor is the number of content words (nouns, verbs, adjectives, adverbs) relative to function words (prepositions, pronouns, conjunctions, etc.).

Semantic coherence: Assess the logical consistency and meaningfulness of the text, which can be done by analyzing the relationships between words, phrases, and sentences, and checking for coherence and cohesion.

Semantic paraphasias: Difficulty in expressing ideas clearly, and errors in word choice that often involve selecting a word that is related in meaning to the intended word or odd combinations of words. Normal: the participant was able to describe the scene clearly.

Lack of coherence: MCI: struggle with organizing thoughts coherently, or maintaining a clear focus in conversation; may change the topic frequently and abruptly; more frequent use of indirect speech or going off on tangents. Normal: greater coherence and organization in their storytelling and conversations.

Consistency in errors: MCI: may make consistent language errors that are unusual for their educational and cultural background.

Lack of semantic fluency: MCI: may have trouble naming items in a category or describing a process or sequence of events.

Disorganized narrative structure: MCI: struggling to maintain a coherent line of thought, and a narrative that jumps around chronologically or lacks key elements.

Orientation Issues: MCI: confusion about date, time, place, or sequence of events

Decline in conversational engagement and responsiveness: MCI: decreased initiation of conversation and less spontaneity; difficulties in following a conversation, responding to questions, or initiating topics; and tendency to steer conversations back to familiar topics or to ignore or inappropriately respond to comments or questions from their conversation partner. Normal: active engagement and spontaneous responses in conversation.

Memory issues: MCI: repeat questions, difficulty in recalling certain information or sequences, difficulty in remembering specific details of the story, or difficulty in recalling major plot points or characters. However, memory issues can also be a part of normal aging, so this sign should be evaluated in the context of the individual's overall cognitive function.

Lack of abstract thinking and synthesis: MCI: despite detailed description, inability to synthesize the overall situation, or trouble understanding or expressing abstract concepts.

Abnormal emotions: MCI: changes in tone or style of speech, lack of Initiative, or increased irritability, depression or apathy.

Finally, please dichotomise PARTICIPANT into either the Probably Normal Cognition Group or the Probably Mild Cognitive Impairment Group (without other outcomes). It is important to note that some of these issues may be present in normal cognitive participants and not all of these assessment indicators may be present. You will need to make a comprehensive synthesis of all the indicators before making a conclusion.

**Model4 prompt:** Designed by GPT-4 with Model3 prompt as the basis.

Please assess a participant with possible mild cognitive impairment (MCI) based on their language features. Note that I do not need to diagnose the participant but rather analyze and rate the severity of each evaluation indicator on a scale of 1 to 10 (1 being the least severe, and 10 being the most severe).

Vocabulary features:

a. Analyze word frequency, word ratio, phrase frequency, and phrase ratio.

b. Calculate the lexical complexity.

c. Assess and rate the severity of the following aspects: limited vocabulary, increased verbosity, overlearned phrases, improper use of pronouns, hesitation, circumlocution, tip-of-the-tongue phenomenon, and repetition.

Syntax and grammar features:

a. Analyze syntactic complexity and grammatical components.

b. Assess and rate the severity of the following aspects: simplified sentence structure and grammar, grammatical errors, and difficulty with complex ideas.

Semantics features:

a. Analyze semantic density and coherence.

b. Assess and rate the severity of the following aspects: semantic paraphasias, lack of coherence, consistency in errors, lack of semantic fluency, disorganized narrative structure, orientation issues, decline in conversational engagement and responsiveness, memory issues, lack of abstract thinking and synthesis, and abnormal emotions.

Based on the comprehensive synthesis of all the indicators, classify the participant into either the Probably Normal Cognition Group or the Probably Mild Cognitive Impairment Group. Keep in mind that these issues might be present in individuals with normal cognition and that not all individuals with MCI will display all of these features. The final categorization should be based on a comprehensive synthesis of all the indicators.

Please note: This evaluation isn't meant to provide a formal diagnosis but to serve as an initial screening tool for potential MCI.

**Designed by GPT-4:**

Please help me in refining the following question request to make it more conducive for you (GPT4) to answer: You do not need to diagnose, just the analysis indicators and evaluation indicators below to initially screen for PARTICIPANT with possibly mild cognitive impairment (MCI). Also, please rate the severity of each evaluation indicator on a scale from 1 to 10 for the PARTICIPANT. (1 as the least severe and 10 as the most severe). Firstly, please analyse the Vocabulary features of PARTICIPANT, including Word frequency and Word ratio, Phrase frequency and Phrase ratio, and Lexical complexity. Then assess the following aspects based on Vocabulary features: 1 Limited vocabulary, 2 Increased verbosity, 3 Overlearned phrases, 4 Improper use of pronouns, 5 Hesitation (Word-finding difficulties), 6 Circumlocution (Word-finding difficulties), 7 Tip-of-the-tongue phenomenon, and 8 Repetition. Word frequency and Word ratio: Count the occurrences of each unique word, each pronoun, and each specific name (or noun) in the text. And calculate the proportion of unique words to the total number of words in the text, and calculate the ratio of pronouns to specific names (or nouns). Phrase frequency and Phrase ratio: Count the occurrences of each unique phrases in the text. And calculate the proportion of unique phrases to the total number of phrases in the text. Lexical complexity: Measure the variety and sophistication of the vocabulary used, considering factors, including word length and frequency of rare words. Limited vocabulary: MCI: slightly reduced vocabulary, and more frequent use of the same words, simple words, or common terms; Normal: richer vocabulary. Increased verbosity: MCI: more verbose and using more words than necessary. Overlearned phrases: MCI: overuse of cliches or stock phrases. Normal: usually demonstrate more variety in their language use. Improper use of pronouns: MCI: a tendency to use more pronouns than specific names (or nouns), or difficulty with pronouns (mixing up "he" and "she" or "I" and "you"). Hesitation (Word-finding difficulties): Frequent pauses or use of filler words, like "uh", "um" or "…". MCI: more disfluencies, longer time to find the right word, or using more filler words. Normal: fluent speech with fewer filled and unfilled pauses. Circumlocution (Word-finding difficulties): MCI: talking in a roundabout way due to difficulty finding the specific word; using many words where fewer would do, especially in a deliberate attempt to be vague or evasive; using a word close to, but not quite the intended word; or substitute more general words for more specific ones. Tip-of-the-tongue phenomenon: this phenomenon is characterized by partial recall. For instance, a person might remember the first letter of a word, its syllable structure, or its general semantic category, without being able to recall the whole word. More common in individuals with MCI. Repetition: MCI: repeat the same stories, questions, or statements within a short span of time. Normal: some repetition of words but not excessive. Secondly, please analyse the Syntax and Grammar features, including Syntactic complexity, and Grammatical components. Then assess the following aspects based on Syntax and Grammar features: 1 Simplified sentence structure and grammar, 2 Grammatical errors, and 3 Difficulty with complex ideas. Syntactic complexity: Measure the complexity of the sentence structure in the text, considering factors such as the number of clauses per sentence, the average length of sentences, and the use of subordinate clauses. Grammatical components: Analyze the text for correct usage of grammar, such as subject-verb agreement, tense consistency, and proper use of pronouns, prepositions, and conjunctions. Simplified sentence structure and grammar: MCI: preference for simpler structures in both long and short sentences, and avoidance of complex grammatical constructions; Normal: in long dialogues there is a greater variety of sentence structures (have a balanced usage of simple and complex sentences) and better use of complex grammatical constructions; but in short dialogues the sentences and grammar may turn out to be simple. Grammatical errors: MCI: more frequent grammatical errors, such as mistakes in verb tense and subject-verb agreement; Normal: fewer grammatical errors (a few errors, but not significant). Difficulty with complex ideas: MCI: while the participant could handle simpler sentences and ideas, they had more difficulty when the narrative or the question became more complex. Thirdly, please analyse the Semantics features of PARTICIPANT, including Semantic density, and Semantic coherence. Then assess the following aspects according to the Semantics features: 1 Semantic paraphasias, 2 Lack of coherence, 3 Consistency in errors, 4 Lack of semantic fluency, 5 Disorganized narrative structure, 6 Orientation Issues, 7 Decline in conversational engagement and responsiveness, 8 Memory issues, 9 Lack of abstract thinking and synthesis, and 10 Abnormal emotions. Semantic density: Measure the amount of meaningful information in the text, and considering factor is the number of content words (nouns, verbs, adjectives, adverbs) relative to function words (prepositions, pronouns, conjunctions, etc.). Semantic coherence: Assess the logical consistency and meaningfulness of the text, which can be done by analyzing the relationships between words, phrases, and sentences, and checking for coherence and cohesion. Semantic paraphasias: Difficulty in expressing ideas clearly, and errors in word choice that often involve selecting a word that is related in meaning to the intended word or odd combinations of words. Normal: the participant was able to describe the scene clearly. Lack of coherence: MCI: struggle with organizing thoughts coherently, or maintaining a clear focus in conversation; may change the topic frequently and abruptly; more frequent use of indirect speech or going off on tangents. Normal: greater coherence and organization in their storytelling and conversations. Consistency in errors: MCI: may make consistent language errors that are unusual for their educational and cultural background. Lack of semantic fluency: MCI: may have trouble naming items in a category or describing a process or sequence of events. Disorganized narrative structure: MCI: struggling to maintain a coherent line of thought, and a narrative that jumps around chronologically or lacks key elements. Orientation Issues: MCI: confusion about date, time, place, or sequence of events Decline in conversational engagement and responsiveness: MCI: decreased initiation of conversation and less spontaneity; difficulties in following a conversation, responding to questions, or initiating topics; and tendency to steer conversations back to familiar topics or to ignore or inappropriately respond to comments or questions from their conversation partner. Normal: active engagement and spontaneous responses in conversation. Memory issues: MCI: repeat questions, difficulty in recalling certain information or sequences, difficulty in remembering specific details of the story, or difficulty in recalling major plot points or characters. However, memory issues can also be a part of normal aging, so this sign should be evaluated in the context of the individual's overall cognitive function. Lack of abstract thinking and synthesis: MCI: despite detailed description, inability to synthesize the overall situation, or trouble understanding or expressing abstract concepts. Abnormal emotions: MCI: changes in tone or style of speech, lack of Initiative, or increased irritability, depression or apathy. Finally, please dichotomise PARTICIPANT into either the Probably Normal Cognition Group or the Probably Mild Cognitive Impairment Group (without other outcomes). It is important to note that some of these issues may be present in normal cognitive participants and not all of these assessment indicators may be present. You will need to make a comprehensive synthesis of all the indicators before making a conclusion.

ChatGPT

1 / 3

Certainly, your request is very detailed, but as an AI, I can't perform the real-time analysis required to evaluate an individual for potential mild cognitive impairment. However, I can help you refine your request to make it more feasible for a human analyst or a specialized AI tool. Here's a suggested revision:

Please provide an initial assessment of the PARTICIPANT's potential for Mild Cognitive Impairment (MCI), based on the analysis of the following indicators:

1. Vocabulary features: Analyze the PARTICIPANT's word and phrase frequency and ratio, as well as lexical complexity. The evaluation should include:
   - Limited vocabulary
   - Increased verbosity
   - Overuse of overlearned phrases
   - Misuse of pronouns
   - Hesitations and circumlocutions (indicative of word-finding difficulties)
   - Tip-of-the-tongue phenomena
   - Repetition of words or phrases
2. Syntax and Grammar features: Evaluate the syntactic complexity and grammatical accuracy of the PARTICIPANT's speech. Look for:
   - Simplified sentence structure and grammar
   - Frequent grammatical errors
   - Difficulty with expressing complex ideas
3. Semantic features: Assess the semantic density and coherence of the PARTICIPANT's speech. Keep an eye out for:
   - Semantic paraphasias
   - Lack of coherence in speech
   - Consistent errors in language use
   - Lack of semantic fluency
   - Disorganized narrative structure
   - Orientation issues
   - Decline in conversational engagement and responsiveness
   - Memory issues
   - Difficulty with abstract thinking and synthesis
   - Abnormal emotional responses

After evaluating these features, please categorize the PARTICIPANT as either probably having normal cognition or probably having MCI. Keep in mind that these issues might be present in individuals with normal cognition and that not all individuals with MCI will display all of these features. The final categorization should be based on a comprehensive synthesis of all the indicators.

Please note: This evaluation isn't meant to provide a formal diagnosis but to serve as an initial screening tool for potential MCI.

ChatGPT

2 / 3

Please help me refine the following request for assessing a participant with possible mild cognitive impairment (MCI) based on their language features. Note that I do not need to diagnose the participant but rather analyze and rate the severity of each evaluation indicator on a scale of 1 to 10 (1 being the least severe, and 10 being the most severe).

1. Vocabulary features: a. Analyze word frequency, word ratio, phrase frequency, and phrase ratio. b. Calculate the lexical complexity. c. Assess and rate the severity of the following aspects: limited vocabulary, increased verbosity, overlearned phrases, improper use of pronouns, hesitation, circumlocution, tip-of-the-tongue phenomenon, and repetition.
2. Syntax and grammar features: a. Analyze syntactic complexity and grammatical components. b. Assess and rate the severity of the following aspects: simplified sentence structure and grammar, grammatical errors, and difficulty with complex ideas.
3. Semantics features: a. Analyze semantic density and coherence. b. Assess and rate the severity of the following aspects: semantic paraphasias, lack of coherence, consistency in errors, lack of semantic fluency, disorganized narrative structure, orientation issues, decline in conversational engagement and responsiveness, memory issues, lack of abstract thinking and synthesis, and abnormal emotions.
4. Based on the comprehensive synthesis of all the indicators, classify the participant into either the Probably Normal Cognition Group or the Probably Mild Cognitive Impairment Group.

ChatGPT

3 / 3

Sure, I'll help to rephrase your question request to make it more straightforward and suitable for me to respond. Here's a simplified version:

I'm trying to identify potential signs of mild cognitive impairment (MCI) in a participant. I don't need a diagnosis, but an analysis of the following indicators in their speech:

1. Vocabulary features: Analyze word and phrase frequency, word ratio, phrase ratio, and lexical complexity. Rate the severity of the following conditions in the participant on a scale of 1 (least severe) to 10 (most severe): limited vocabulary, increased verbosity, overuse of overlearned phrases, improper use of pronouns, hesitations or circumlocutions indicating word-finding difficulties, the tip-of-the-tongue phenomenon, and repetition.
2. Syntax and Grammar features: Analyze syntactic complexity and grammatical components. Rate the severity of the following conditions: simplified sentence structure and grammar, grammatical errors, and difficulty with complex ideas.
3. Semantics features: Analyze semantic density and coherence. Rate the severity of the following conditions: semantic paraphasias, lack of coherence, consistency in errors, lack of semantic fluency, disorganized narrative structure, orientation issues, decline in conversational engagement and responsiveness, memory issues, lack of abstract thinking and synthesis, and abnormal emotions.

After this, please classify the participant into one of two groups based on your evaluation of the indicators: Probably Normal Cognition or Probably Mild Cognitive Impairment. Remember, not all indicators may be present and some may be present in those with normal cognition. The conclusion should be based on a comprehensive synthesis of all indicators.

**Model5 prompt**: Design by analyzing the severity of each indicator obtained from GPT4-Model3 and following the Model4 prompt format.

Please assess a participant with possible mild cognitive impairment (MCI) based on their language features. Note that I do not need to diagnose the participant but rather analyze and rate the severity of each evaluation indicator on a scale of 1 to 10 (1 being the least severe, and 10 being the most severe).

Vocabulary features:

a. Analyze word frequency, word ratio, phrase frequency, and phrase ratio. And calculate the lexical complexity. ( Word frequency and Word ratio: Count the occurrences of each unique word, each pronoun, and each specific name (or noun) in the text. And calculate the proportion of unique words to the total number of words in the text, and calculate the ratio of pronouns to specific names (or nouns). Phrase frequency and Phrase ratio: Count the occurrences of each unique phrases in the text. And calculate the proportion of unique phrases to the total number of phrases in the text. Lexical complexity: Measure the variety and sophistication of the vocabulary used, considering factors, including word length and frequency of rare words. )

b. Assess and rate the severity of the following aspects: tip-of-the-tongue phenomenon. (This phenomenon is characterized by partial recall. For instance, a person might remember the first letter of a word, its syllable structure, or its general semantic category, without being able to recall the whole word. More common in individuals with MCI.)

Syntax and grammar features:

a. Analyze syntactic complexity and grammatical components. (Syntactic complexity: Measure the complexity of the sentence structure in the text, considering factors such as the number of clauses per sentence, the average length of sentences, and the use of subordinate clauses. Grammatical components: Analyze the text for correct usage of grammar, such as subject-verb agreement, tense consistency, and proper use of pronouns, prepositions, and conjunctions.)

b. Assess and rate the severity of the following aspects: difficulty with complex ideas (MCI: while the participant could handle simpler sentences and ideas, they had more difficulty when the narrative or the question became more complex.)

Semantics features:

a. Analyze semantic density and coherence. (Semantic density: Measure the amount of meaningful information in the text, and considering factor is the number of content words (nouns, verbs, adjectives, adverbs) relative to function words (prepositions, pronouns, conjunctions, etc.). Semantic coherence: Assess the logical consistency and meaningfulness of the text, which can be done by analyzing the relationships between words, phrases, and sentences, and checking for coherence and cohesion.)

b. Assess and rate the severity of the following aspects: memory issues (MCI: repeat questions, difficulty in recalling certain information or sequences, difficulty in remembering specific details of the story, or difficulty in recalling major plot points or characters. However, memory issues can also be a part of normal aging, so this sign should be evaluated in the context of the individual's overall cognitive function.)

Based on the comprehensive synthesis of all the indicators, classify the participant into either the Probably Normal Cognition Group or the Probably Mild Cognitive Impairment Group. Keep in mind that these issues might be present in individuals with normal cognition and that not all individuals with MCI will display all of these features. The final categorization should be based on a comprehensive synthesis of all the indicators.

Please note: This evaluation isn't meant to provide a formal diagnosis but to serve as an initial screening tool for potential MCI.

**Feedback1**:

1. Vocabulary:
   - MCI: Slightly reduced vocabulary, more frequent use of simpler words or common terms, and more instances of word repetition.
   - Normal: Richer vocabulary, less frequent repetition, and better lexical retrieval.
2. Sentence structure:
   - MCI: Simpler sentence structures, avoidance of complex grammatical constructions.
   - Normal: More varied sentence structures and better use of complex grammatical constructions.
3. Repetition:
   - MCI: More frequent repetition of words, phrases, or ideas, difficulty recalling whether they've mentioned something before.
   - Normal: Less frequent repetition and better memory of what has been mentioned.
4. Word-finding difficulties:
   - MCI: More frequent instances of word-finding difficulties, longer time to find the right word, or using more general terms.
   - Normal: Fewer instances of word-finding difficulties and better word retrieval.
5. Coherence and organization:
   - MCI: Struggle with organizing thoughts coherently or maintaining a clear focus in conversation.
   - Normal: Greater coherence and organization in their storytelling and conversations.
6. Grammatical errors:
   - MCI: More frequent grammatical errors, such as mistakes in verb tense and subject-verb agreement.
   - Normal: Fewer grammatical errors and better language production

Normal:

Vocabulary:

1. Limited vocabulary: Low limitation; some repetition of words but not excessive.

Semantics:

1. Inability to convey meaning: Low; the participant was able to describe the scene clearly.

Syntax and Grammar:

1. Syntactic complexity: Moderate complexity; not overly simplified.
2. Grammatical errors: A few errors, but not significant.

**Feedback2**:

Here's a summary of the points that distinguish mild cognitive impairment (MCI) from normal cognition:

1. Lack of Synthesis: Despite detailed description, inability to synthesize the overall situation may hint at problems with abstract thinking.
2. Sentence Complexity: A shift towards simpler sentence structures could indicate difficulty with complex language tasks.

Additional linguistic indicators that could be considered:

4. Conversational Engagement: Decreased initiation of conversation and less spontaneity might suggest MCI.
5. Story Recall: Difficulty in recalling major plot points or characters could be a sign of MCI.
6. Temporal Orientation: Difficulty with understanding and awareness of time, date, and sequence of events.
7. Abstract Thinking: Difficulty in understanding and explaining abstract concepts.
8. Attention to Detail: Missing subtle details while describing pictures.
9. Response Latency: Increased response time could indicate MCI.
10. Speech Fluency: More disfluencies, like rate of speech, filled and unfilled pauses, and the flow of speech.
11. Use of Overlearned Phrases: Overuse of cliches or stock phrases.

Points related to the participant's conversation:

12. Word-finding Difficulties: Increased difficulty finding the right words.
13. Repetition: Tendency to repeat certain phrases.
14. Simplified Language: Shift towards simpler words and sentences.
15. Difficulty Following Complex Conversations: Trouble understanding or responding to complex prompts.
16. Slower Speech or Pauses: Frequent pauses mid-sentence or slower speech.

Other potential linguistic indicators:

17. Tip-of-the-tongue Phenomenon: More common in individuals with MCI.
18. Frequent Pauses or Hesitations: Excessive pauses may indicate struggle to find the right words.
19. Indirect Speech: More frequent use of indirect speech or going off on tangents.
20. Difficulty with Pronouns: Mixing up "he" and "she" or "I" and "you".
21. Word Substitution: Using a word close to but not quite the intended word.
22. Reduced Speech Fluency: More "ums" and "uhs" and other filler words.
23. Difficulty Following Conversations: Harder time following complex conversations or instructions.
24. Difficulty with Abstract Concepts: Harder time understanding or explaining abstract concepts.
25. Changes in Tone or Style of Speech: Speaking in a different tone or style.
26. Changes in Written Language: More spelling or grammar errors in their written language.

Please note, all these are potential signs and do not definitively diagnose MCI. A comprehensive evaluation by a professional is necessary for an accurate diagnosis.

Sure, here are the key points differentiating mild cognitive impairment (MCI) from normal cognition as outlined in your text:

1. **Hesitation**: In conversations, hesitation can indicate a need for more processing time, which might be a sign of MCI. In normal cognition, responses usually flow more fluently with less noticeable hesitation.
2. **Detailed description but lack of synthesis**: While detailed descriptions are a normal part of conversation, the inability to synthesize or summarize these details into an overarching theme can suggest MCI. A healthy cognitive state typically allows individuals to both describe details and synthesize them into a coherent whole.
3. **Sentence complexity**: Preference for simpler sentence structures could indicate difficulty with complex language tasks, possibly suggesting MCI. In contrast, individuals with normal cognition usually have a balanced usage of simple and complex sentences.
4. **Conversational engagement**: Decreased initiation of conversation and less spontaneity in responses can suggest MCI. On the other hand, normal cognition is typically marked by active engagement and spontaneous responses in conversation.
5. **Story recall**: Difficulty recalling major plot points or characters can indicate MCI. Normally, individuals can recall and retell stories with relative ease.
6. **Temporal orientation**: Problems with understanding and awareness of time, date, and sequence of events can be indicative of MCI. Individuals with normal cognition generally maintain a clear understanding of these elements.
7. **Abstract thinking**: Difficulty in understanding and explaining abstract concepts or proverbs could suggest MCI, whereas normal cognition allows individuals to handle abstract concepts effectively.
8. **Attention to detail**: While describing pictures, individuals with MCI might miss subtle details. People with normal cognition are generally more attentive to such details.
9. **Response latency**: Increased response time can indicate MCI. In normal cognition, response times to questions or prompts are generally quicker.
10. **Speech fluency**: Disfluencies might be more common in individuals with MCI. Normal cognition is generally characterized by fluent speech with fewer filled and unfilled pauses.
11. **Use of overlearned phrases**: Overuse of cliches or stock phrases might be more common in MCI. Individuals with normal cognition usually demonstrate more variety in their language use.
12. **Word-finding difficulties and repetition**: People with MCI might exhibit increased difficulty finding the right words during conversations and might repeat phrases. These issues are less common in normal cognition.
13. **Difficulty following complex conversations or instructions**: If an individual has trouble understanding or responding to complex prompts, it could suggest cognitive impairment. In normal cognition, individuals can generally follow complex instructions and conversations more easily.
14. **Simplified language and slower speech**: Some people with MCI might start to use simpler words and sentences and speak more slowly. In contrast, normal cognition usually involves a balance of complex and simple language structures and a steady speech rate.

These linguistic and cognitive indicators can provide clues about an individual's cognitive health. However, these signs are often subtle and may not be definitively indicative of MCI. A comprehensive evaluation by a professional is necessary for an accurate diagnosis.

Below are the methods mentioned for distinguishing mild cognitive impairment (MCI) from normal cognition:

1. **Pauses and Speech Fluency**: One sign of MCI might be longer pauses, slow speech, or difficulty finding the right words during an oral conversation. This is because MCI can cause difficulties with language production, which might manifest as these speech fluency issues.
2. **Memory Issues**: MCI often affects memory, so if an individual struggles to recall certain information or sequences, this might suggest MCI. However, memory issues can also be a part of normal aging, so this sign should be evaluated in the context of the individual's overall cognitive function.
3. **Repetitive Speech**: Repetition could be a sign of MCI, as it might indicate issues with working memory. However, it can also occur in normal speech, so this sign should be evaluated alongside other potential indicators of MCI.
4. **Attention and Concentration**: The ability to maintain attention or concentrate during a conversation can be affected in MCI. This can be assessed through various cognitive tests that measure attention and concentration.
5. **Orientation and Time Awareness**: Confusion regarding time, place, or personal details could indicate MCI. This is usually assessed through questioning during a clinical examination.
6. **Executive Functioning**: Tasks involving planning, organizing, or abstract thinking can be challenging for someone with MCI. These aspects are often evaluated through specific cognitive tests that measure executive function, such as the Trail Making Test or the Wisconsin Card Sorting Test.
7. **Emotional Changes**: Changes in mood or personality, such as increased anxiety or depression, can sometimes accompany cognitive changes in MCI. These changes are typically assessed through clinical interviews or questionnaires.
8. **Repetition and Word-Finding Difficulties**: Frequent use of filler words and a noticeable amount of repetition in dialogue could indicate challenges with working memory and language production, which are common signs of MCI.
9. **Simplified Sentence Structure and Limited Vocabulary**: A lack of complexity in sentence structure and a reliance on common words and phrases might suggest difficulties with executive functioning and cognitive flexibility, which are often affected in MCI.
10. **Lack of Coherence**: If an individual's thoughts appear disjointed or unrelated, this might indicate struggles with organizing thoughts coherently, which can be a sign of MCI.
11. **Inability to Convey Meaning**: Struggles to express ideas clearly or a lack of detail in storytelling might suggest MCI.
12. **Circumlocution**: Using many words where fewer would do, especially in a deliberate attempt to be vague or evasive, might be a compensatory strategy when the individual can't remember a specific word.
13. **Language Production Rate**: Speaking more slowly or taking longer to express thoughts might suggest MCI.
14. **Content**: Going off-topic or losing the thread of their conversation could suggest MCI.
15. **Narrative Structure**: Difficulties in maintaining a clear and consistent narrative structure could be indicative of MCI.
16. **Inference Making**: Struggles with understanding things that are not explicitly stated might suggest MCI.
17. **Abstract Thinking**: Reduced ability to understand and use metaphors or other forms of abstract language might suggest MCI.
18. **Pragmatic Skills**: Difficulty understanding sarcasm, humor, or other subtle uses of language might suggest MCI.
19. **Increased Verbosity**: Using more words than necessary to express a simple idea could suggest MCI.
20. **Disorganized Narrative**: Having difficulty maintaining a coherent narrative could suggest MCI.
21. **Grammatical Errors**: Making frequent grammatical errors could suggest MCI.
22. **Difficulty Understanding Complex Sentences or Instructions**

The methods to distinguish Mild Cognitive Impairment (MCI) from normal cognition as mentioned in your provided texts are:

1. **Observing pauses and speech fluency**: Longer pauses, slow speech, or difficulty finding the right words could indicate MCI.
2. **Checking for memory issues**: Struggles with certain aspects of memory not covered in general assessments could indicate MCI.
3. **Looking for repetitive speech**: Repetition of certain details and phrases could be a sign of MCI.
4. **Assessing attention and concentration**: Difficulty maintaining attention or concentrating can indicate MCI.
5. **Testing orientation and time awareness**: Signs of confusion regarding time, place, or personal details could suggest MCI.
6. **Evaluating executive functioning**: Difficulties with tasks involving planning, organizing, or abstract thinking can indicate MCI.
7. **Noting emotional changes**: Changes in mood or personality, such as increased anxiety or depression, can sometimes accompany cognitive changes indicating MCI.
8. **Looking for word-finding difficulties**: Frequent use of filler words and repetition in dialogue could suggest difficulties with working memory, a common sign of MCI.
9. **Examining sentence structure and vocabulary**: Simplified sentences and a notable reliance on common words and phrases could suggest difficulties with executive functioning and cognitive flexibility.
10. **Checking for coherence**: Disjointed or unrelated thoughts could indicate struggles with organizing thoughts coherently.
11. **Checking ability to convey meaning**: Struggles to express ideas clearly, leading to confusion or lack of details could suggest MCI.
12. **Observing inconsistencies in storytelling**: Changes in story details midway could suggest MCI.
13. **Noting overuse of filler words**: Frequent use of "uh" and "um" could indicate a struggle to find the right words, suggesting MCI.
14. **Looking for repetition of ideas**: Repeating the same information with slight variations could suggest MCI.
15. **Checking for ambiguity**: Unclear descriptions of things or events could suggest MCI.

Remember, these are potential indicators and not definitive signs of MCI. A comprehensive assessment by healthcare professionals is essential for an accurate diagnosis

Sure, here are the methods mentioned for distinguishing mild cognitive impairment (MCI) from normal cognition:

1. **Observe Memory Issues**: Look for patterns where individuals may forget conversations or appointments more frequently than before. They might repeat questions, not remembering they've asked them already.
2. **Notice Attention Difficulties**: People with MCI may have a harder time focusing on tasks or may be easily distracted. This could manifest as difficulty following along with more complex conversation topics or multitasking.
3. **Check for Executive Functioning Problems**: Look for problems in planning, organization, problem-solving, and executing tasks. If a person has trouble making plans, following through with them, or making decisions, this could indicate MCI.
4. **Look for Orientation Issues**: People with MCI might show signs of confusion about the date, time, or place.
5. **Observe Changes in Mood or Behavior**: Changes in personality, increased irritability, depression, and apathy can sometimes be associated with MCI.
6. **Check for Difficulty with Visual Perception**: Trouble judging distance, understanding visual images, or differentiating colors could also indicate MCI.
7. **Look for Apathy or Lack of Initiative**: A person with MCI might lose interest in activities they previously enjoyed, or might not feel motivated to initiate activities or tasks.
8. **Look for Increased Verbosity**: People with MCI may be more verbose and provide more information than is necessary, which can be a compensatory strategy for cognitive difficulties.
9. **Observe Frequent Topic Shifts**: Frequent and abrupt changes in topic could indicate difficulties in maintaining a coherent line of thought.
10. **Check for Semantic Paraphasias**: These are errors in word choice that often involve selecting a word that is related in meaning to the intended word.
11. **Look for Circumlocution**: This involves talking in a roundabout way to get to a point when a person cannot think of the specific word they want to use.
12. **Look for Difficulty with Pronouns**: People with MCI might have trouble keeping track of who 'he', 'she', or 'they' are when pronouns are used in a conversation.
13. **Check for Disorganized Narrative Structure**: People with MCI may struggle with organizing their thoughts coherently, which might result in a narrative that jumps around chronologically or lacks key elements.
14. **Observe Decline in Conversational Engagement and Responsiveness**: This may manifest as a tendency to ignore or not respond appropriately to the conversational partner's comments or questions.
15. **Look for Increased Pauses and Filler Words**: Individuals with MCI may show increased use of pauses and filler words such as "uh", "um", or "you know", indicating difficulty in finding the right words or maintaining the flow of conversation.
16. **Check for Subtle Changes in Story Recall**: Individuals with MCI might struggle to remember specific details of the story, even if they can remember the overall theme.
17. **Look for Difficulty with Complex Ideas**: While the participant could handle simpler sentences and ideas, they had more difficulty when the narrative or the question became more complex.
18. **Check for Repetition of Content Words and Phrases**: Individuals with MCI may repeat certain words or phrases, even in the same context. This could be a subtle sign of memory difficulty.
19. **Look for Length and Complexity of Utterances**: In some cases, individuals with MCI may produce shorter utterances and may show a tendency towards simplified syntactic structures.
20. **Look for Consistency in Errors**: People with MCI may make consistent language errors that are unusual for their educational and cultural background.
21. **Check for Difficulty with Abstract Concepts**

Here's a summarised list of the methods mentioned in the text for distinguishing mild cognitive impairment (MCI) from normal cognition:

**General Cognitive and Behavioral Indicators:**

1. Frequent memory issues, such as forgetting conversations or appointments.
2. Difficulties with attention, such as being easily distracted or having trouble focusing on complex tasks.
3. Problems with executive functioning, such as planning, organization, and problem-solving.
4. Confusion about date, time, or place.
5. Changes in mood or behavior, including personality changes, irritability, depression, and apathy.
6. Trouble with visual perception, such as judging distance or differentiating colors.
7. Apathy or lack of initiative, such as losing interest in activities they previously enjoyed.

**Linguistic Indicators:**

1. Increased verbosity or providing more information than necessary.
2. Frequent and abrupt changes in conversation topic.
3. Semantic paraphasias or errors in word choice.
4. Circumlocution or talking in a roundabout way due to difficulty finding the specific word.
5. Difficulty with pronouns, such as losing track of who 'he', 'she', or 'they' are in a conversation.
6. Disorganized narrative structure and struggling to maintain a coherent line of thought.
7. Decline in conversational engagement and responsiveness.
8. Increased use of pauses and filler words.
9. Subtle changes in story recall.
10. Struggles with maintaining conversational coherence.
11. Difficulty handling complex ideas or sentences.
12. Repetition of content words and phrases.
13. Tendency towards simplified syntactic structures.
14. Decreased fluency in speech.
15. Simplified grammar or syntax.
16. Impaired comprehension of complex language, indirect requests, or figurative language.
17. Semantic anomalies or odd combinations of words.
18. Difficulty maintaining a conversation topic or frequently shifting topics.
19. Consistent language errors.
20. Trouble understanding or expressing abstract concepts.
21. Difficulty constructing a coherent narrative.
22. Changes in speech rate and fluency.
23. Reduced complexity of expressed ideas.
24. Reduced use of specific terms and increased use of general or vague terms.
25. Difficulty following complex instructions.
26. Changes in pragmatic aspects of communication, such as turn-taking, topic maintenance, and understanding non-literal language.

**Memory-Related and Cognitive Signs:**

1. Increased forgetfulness, including forgetting important dates, events, or conversations.
2. Difficulty with complex tasks, such as taking longer to complete routine tasks.
3. Impaired decision-making.
4. Spatial and temporal disorientation, such as trouble with directions or losing track of time.
5. Decreased attention span and easily getting distracted.

**Behavioral Signs:**

1. Mood and personality changes, including increased anxiety, apathy, depression, or irritability.

These indicators, while helpful, are not definitive proof of MCI and can be associated with normal aging, stress, or other conditions. Therefore, a comprehensive clinical evaluation is necessary for a definitive diagnosis.

The language changes associated with MCI and early Alzheimer's disease can be subtle and may not be immediately apparent in the context of everyday conversation or a single brief interaction. However, over time and with repeated interactions, certain patterns of speech and language use may become more noticeable. These can include:

1. Difficulty finding the right words: Individuals may struggle to find the correct words during conversation, leading to pauses, filler words like "um" or "uh", or circumlocution (talking around the word they can't remember). They might also substitute more general words for more specific ones (e.g., "thing" instead of "car").
2. Repetitive speech: They might repeat the same stories, questions, or statements within a short span of time, often without realizing that they're doing so.
3. Issues with semantic fluency: They may have trouble naming items in a category or describing a process or sequence of events. This can be a sign of difficulty accessing stored knowledge.
4. Decreased complexity in sentence structure: Their speech may become simpler over time, with shorter sentences and a reduction in the use of complex grammatical constructions.
5. Difficulty understanding complex sentences: They might have trouble following complex instructions or understanding sentences that have multiple clauses or indirect references.
6. Problems with topic maintenance: They might have difficulty staying on topic during conversation, frequently shifting to unrelated subjects or going off on tangents.
7. Challenges with abstract language: They might struggle to understand or use figurative language, such as idioms, metaphors, and sarcasm.
8. Altered conversational dynamics: They might interrupt more, have difficulty taking turns in conversation, or not respond appropriately to conversational cues.

It's important to note that these changes can also occur in the context of normal aging, stress, fatigue, or other medical conditions. Thus, it's essential that these observations be considered in the broader context of the individual's overall health, cognitive function, and daily functioning.

The methods for distinguishing mild cognitive impairment (MCI) from normal cognition, as mentioned, can be summarized as follows:

1. **Disfluencies**: Monitor for an increase in interruptions in speech or fillers, which can indicate word-finding difficulties.
2. **Content**: Look for changes in the content of speech, such as vagueness, irrelevance, or a lack of specificity.
3. **Conversational Engagement**: Pay attention to difficulties in following a conversation, responding to questions, or initiating topics, and tendency to steer conversations back to familiar topics.
4. **Story Recall**: Assess the ability to accurately recall and recount a recently heard story.
5. **Narrative Construction**: Evaluate the ability to tell a story or describe a complex process. Less detailed, less structured, and less coherent narratives could suggest MCI.
6. **Abstract Language**: Monitor for difficulties in understanding or using abstract language, metaphors, and idioms.
7. **Pronoun Use**: Note if there's a tendency to use pronouns more often than specific names or nouns, making speech harder to follow.
8. **Temporal Orientation**: Check for confusion about dates, times, and sequencing of events.
9. **Topic Maintenance**: Look for difficulty in maintaining the topic of conversation and frequent digression.
10. **Discourse Management**: Evaluate the ability to construct a coherent narrative or argument.
11. **Understanding Complex Instructions**: Check for difficulty in understanding and following complex instructions or explanations.
12. **Semantic Paraphasia**: Look for instances of using a word related in meaning but not correct in context.
13. **Decreased Fluency**: Note if the individual speaks more slowly, with more pauses, indicating difficulty with word retrieval.
14. **Pragmatic Skills**: Monitor for difficulties in using language in social contexts, such as not taking turns properly in conversation, lack of eye contact, or inappropriate facial expressions or body language.
15. **Metaphorical Language**: Check for difficulties in understanding or using metaphorical or abstract language.
16. **Reading and Writing Skills**: Note a decline in ability to read and write complex sentences or understand complex written information.
17. **Semantic Fluency**: Check for struggle with tasks that require naming as many items as possible from a certain category in a given time.
18. **Narrative Skills**: Monitor for trouble in narrating a complex story or event.
19. **Abstract Thinking**: Check for difficulty in handling abstract concepts and engaging in abstract thinking.
20. **Monitoring and Correcting Errors**: Note if the individual does not notice when they've made a speech or language error, and if they don't correct themselves as often.
21. **Consistency in Complexity**: Look for noticeable variance in complexity, with some sentences being overly simplified and others unnecessarily complex.
22. **Word-finding difficulties**: Observe frequent use of filler words which might indicate difficulty in finding the right words.
23. **Repetition**: Note repetition of certain elements or ideas, which can be a sign of MCI.
24. **Slower speech**: Check for slower speech rate or longer pauses as they try to find the right words or remember details.
25. **Less detailed descriptions**: Look for basic and less detailed descriptions, tending to simpler language.
26. **Simplified sentence structure**: Check for a tendency towards simpler sentences.
27. **Misinterpretation of the scene**: Look for misinterpretations of scenes or scenarios, which can be a sign of MCI.
28. **Discourse Analysis**: Analyze the structure and content of spontaneous speech, such as how ideas are organized, connected, and expressed.
29. **Speech Tempo and Fluency**: Monitor for changes in the speed, rhythm, and fluency of speech.

**R code 1**: for TP, FN, TN, and FP, the four groups

\# Install and load the necessary packages

\# install.packages("readxl")

\# install.packages("dplyr")

\# install.packages("tidyverse")

library(readxl)

library(dplyr)

library(tidyverse)

\# Load the data

data <- read_excel("F:/study2022-2122/chatGPT/AD/Sorted data/Participant Information - 4 VS 3.5.xlsx", sheet = "Variable_selection_data")

\# Calculate descriptive statistics

descriptive_stats <- data %>%

group_by(Group) %>%

summarise(across(everything(), list(mean = mean, sd = sd, median = median, IQR = IQR, min = min, max = max), .names = "{.col}.{.fn}"))

\# Install and load the necessary package

\# install.packages("writexl")

library(writexl)

\# Write the data frame to an Excel file

write_xlsx(descriptive_stats, "F:/study2022-2122/chatGPT/AD/Sorted data/Descriptive_Statistics.xlsx")

\# Install and load the necessary package

\# install.packages("car")

library(car)

\# Initialize a data frame to store the results

normality_and_homogeneity <- data.frame()

\# Loop through each variable

for (variable in names(data)[-1]) {

\# Perform the Shapiro-Wilk test for normality

shapiro_result <- shapiro.test(data[[variable]])$p.value

\# Perform Levene's test for homogeneity of variance

levene_result <- leveneTest(data[[variable]] ~ Group, data = data, center = median)$Pr[1]

\# Combine the results

results <- data.frame(Variable = variable, Shapiro_Wilk_p_value = shapiro_result, Levene_p_value = levene_result)

\# Add the results to the data frame

normality_and_homogeneity <- rbind(normality_and_homogeneity, results)

}

\# Write the data frame to an Excel file

write_xlsx(normality_and_homogeneity, "F:/study2022-2122/chatGPT/AD/Sorted data/Normality_and_Homogeneity.xlsx")

\# Install and load the necessary packages

\# install.packages("FSA")

library(FSA)

\# Initialize a data frame to store the results

kruskal_and_dunn <- data.frame()

\# Loop through each variable

for (variable in names(data)[-1]) {

\# Perform the Kruskal-Wallis H test

kruskal_result <- kruskal.test(data[[variable]] ~ Group, data = data)$p.value

\# Perform Dunn's test if the Kruskal-Wallis H test is significant

if (kruskal_result < 0.05) {

dunn_result <- dunnTest(data[[variable]] ~ Group, data = data, method = "bonferroni")$res

} else {

dunn_result <- NA

}

\# Combine the results

results <- data.frame(Variable = variable, Kruskal_Wallis_p_value = kruskal_result, Dunn_p_value = dunn_result)

\# Add the results to the data frame

kruskal_and_dunn <- rbind(kruskal_and_dunn, results)

}

\# Write the data frame to an Excel file

write_xlsx(kruskal_and_dunn, "F:/study2022-2122/chatGPT/AD/Sorted data/Kruskal_and_Dunn.xlsx")

\# Initialize a data frame to store the results

anova_and_tukey <- data.frame()

\# Loop through each variable

for (variable in names(data)[-1]) {

\# Perform the one-way ANOVA test

aov_result <- summary(aov(data[[variable]] ~ Group, data = data))[[1]]["Group", "Pr(>F)"]

\# Perform Tukey's HSD test if the ANOVA test is significant

if (aov_result < 0.05) {

tukey_result <- TukeyHSD(aov(data[[variable]] ~ Group, data = data))$Group

} else {

tukey_result <- NA

}

\# Combine the results

results <- data.frame(Variable = variable, ANOVA_p_value = aov_result, Tukey_p_value = tukey_result)

\# Add the results to the data frame

anova_and_tukey <- rbind(anova_and_tukey, results)

}

\# Write the data frame to an Excel file

write_xlsx(anova_and_tukey, "F:/study2022-2122/chatGPT/AD/Sorted data/ANOVA_and_Tukey.xlsx")

\# Install and load the necessary packages

\# install.packages(c("ggplot2", "RColorBrewer", "ggpubr"))

library(ggplot2)

library(RColorBrewer)

library(ggpubr)

\# Define the colors

colors <- brewer.pal(4, "Set3")

\# Define the order of the groups

group_order <- c("TP", "FP", "FN", "TN")

\# Define the comparisons for Tukey's HSD test

comparisons <- list(c("TP", "FP"), c("TP", "TN"), c("TN", "FN"))

\# Loop through each variable

for (variable in names(data)[-1]) {

\# Perform the one-way ANOVA test

aov_result <- summary(aov(data[[variable]] ~ Group, data = data))[[1]]["Group", "Pr(>F)"]

\# Create the violin plot

p <- ggplot(data, aes(x = factor(Group, levels = group_order), y = data[[variable]], fill = Group)) +

geom_violin(scale = "width", trim = FALSE, color = NA) +

geom_boxplot(width = 0.1, fill = "white") +

stat_summary(fun = mean, geom = "point", shape = 23, size = 3) +

stat_summary(fun.data = function(x){mean_cl_normal(x, conf.int = 0.95)}, geom = "errorbar", width = 0.2, linetype = "dashed", color = "grey20") +

scale_fill_manual(values = colors) +

scale_y_continuous(limits = c(0, 10)) +

labs(x = "Group", y = variable) +

theme_bw() +

theme(panel.border = element_blank(), panel.grid.major = element_line(colour = "grey90"), panel.grid.minor = element_blank(), axis.line = element_line(colour = "black"), legend.position = "none") +

annotate("text", x = 2.5, y = 10, label = paste("ANOVA p-value:", format(aov_result, digits = 6, scientific = TRUE)), hjust = 0.5, vjust = 1)

\# Add Tukey's HSD test annotations

if (aov_result < 0.05) {

tukey_result <- TukeyHSD(aov(data[[variable]] ~ Group, data = data))$Group

for (i in 1:length(comparisons)) {

comparison <- comparisons[[i]]

p_value <- tukey_result[paste(comparison, collapse = "-"), "p adj"]

p <- p + stat_compare_means(comparisons = list(comparison), label.x.npc = "left", label.y = 10 - i, method = "t.test",

label = paste("p =", format(p_value, digits = 6, scientific = TRUE)))

}

}

\# Save the plot as an image

ggsave(paste0("F:/study2022-2122/chatGPT/AD/Sorted data/", variable, ".png"), plot = p, width = 10, height = 7)

}

**R code 2**: for P, and N, the two groups

\# Install and load the necessary packages

\# install.packages("readxl")

\# install.packages("dplyr")

\# install.packages("tidyverse")

library(readxl)

library(dplyr)

library(tidyverse)

\# Load the data

data <- read_excel("F:/study2022-2122/chatGPT/AD/Sorted data/Participant Information - 4 VS 3.5.xlsx", sheet = "Variable_selection_data (2)")

\# Calculate descriptive statistics

descriptive_stats <- data %>%

group_by(Group) %>%

summarise(across(everything(), list(mean = mean, sd = sd, median = median, IQR = IQR, min = min, max = max), .names = "{.col}.{.fn}"))

\# Install and load the necessary package

\# install.packages("writexl")

library(writexl)

\# Write the data frame to an Excel file

write_xlsx(descriptive_stats, "F:/study2022-2122/chatGPT/AD/Sorted data/Descriptive_Statistics2.xlsx")

\# Install and load the necessary package

\# install.packages("car")

library(car)

\# Initialize a data frame to store the results

normality_and_homogeneity <- data.frame()

\# Loop through each variable

for (variable in names(data)[-1]) {

\# Perform the Shapiro-Wilk test for normality

shapiro_result <- shapiro.test(data[[variable]])$p.value

\# Perform Levene's test for homogeneity of variance

levene_result <- leveneTest(data[[variable]] ~ Group, data = data, center = median)$Pr[1]

\# Combine the results

results <- data.frame(Variable = variable, Shapiro_Wilk_p_value = shapiro_result, Levene_p_value = levene_result)

\# Add the results to the data frame

normality_and_homogeneity <- rbind(normality_and_homogeneity, results)

}

\# Write the data frame to an Excel file

write_xlsx(normality_and_homogeneity, "F:/study2022-2122/chatGPT/AD/Sorted data/Normality_and_Homogeneity2.xlsx")

\# Perform independent t-tests

t_tests <- lapply(names(data)[-1], function(x) {

result <- tryCatch(t.test(as.formula(paste(x, "~ Group")), data = data)$p.value,

error = function(e) NA)

data.frame(Variable = x, t_test_p_value = result)

})

\# Combine the results into one data frame

t_tests_results <- bind_rows(t_tests)

\# Write the results to an Excel file

write_xlsx(t_tests_results, "F:/study2022-2122/chatGPT/AD/Sorted data/t_test_results.xlsx")

\# Perform Mann-Whitney U tests

wilcox_tests <- lapply(names(data)[-1], function(x) {

result <- tryCatch(wilcox.test(as.formula(paste(x, "~ Group")), data = data)$p.value,

error = function(e) NA)

data.frame(Variable = x, wilcox_test_p_value = result)

})

\# Combine the results into one data frame

wilcox_tests_results <- bind_rows(wilcox_tests)

\# Write the results to an Excel file

write_xlsx(wilcox_tests_results, "F:/study2022-2122/chatGPT/AD/Sorted data/wilcox_test_results.xlsx")

**R code 3**: Sensitivity, Specificity, Positive Predictive Value, Negative Predictive Value, Positive Likelihood Ratios, Negative Likelihood Ratios, Accuracy, and AUC-ROC.

\# Install necessary R packages

\# install.packages(c("readxl", "pROC", "MLmetrics", "writexl"))

\# Load necessary R packages

library(readxl)

library(pROC)

library(MLmetrics)

library(writexl)

library(RColorBrewer)

\# Load the data

\# df <- read_excel("F:/study2022-2122/chatGPT/AD/Sorted data/Participant Information - 4 VS 3.5.xlsx", sheet = "Training_GPT4")

\# df <- read_excel("F:/study2022-2122/chatGPT/AD/Sorted data/Participant Information - 4 VS 3.5.xlsx", sheet = "Training_GPT3.5")

\# df <- read_excel("F:/study2022-2122/chatGPT/AD/Sorted data/Participant Information - 4 VS 3.5.xlsx", sheet = "Testing")

\# Define the models and perform the analysis

\# model_names <- c('GPT4_model1', 'GPT4_model2', 'GPT4_model3', 'GPT4_model4', 'GPT4_model5')

\# model_names <- c('GPT3.5_model1', 'GPT3.5_model2', 'GPT3.5_model3', 'GPT3.5_model4', 'GPT3.5_model5')

\# model_names <- c('GPT4_model5', 'GPT3.5_model5')

stats_list <- list()

for (model in model_names) {

prediction <- df[[model]]

actual <- df$Group

TP <- sum((prediction == 1) & (actual == 1))

FN <- sum((prediction == 0) & (actual == 1))

FP <- sum((prediction == 1) & (actual == 0))

TN <- sum((prediction == 0) & (actual == 0))

sensitivity <- TP / (TP + FN)

specificity <- TN / (FP + TN)

PPV <- TP / (TP + FP)

NPV <- TN / (FN + TN)

PLR <- sensitivity / (1 - specificity)

NLR <- (1 - sensitivity) / specificity

accuracy <- (TP + TN) / (TP + FP + FN + TN)

stats_list[[model]] <- c(TP = TP, FN = FN, FP = FP, TN = TN, Sensitivity = sensitivity, Specificity = specificity, PPV = PPV, NPV = NPV, PLR = PLR, NLR = NLR, Accuracy = accuracy)

}

stats_df <- do.call(rbind, stats_list)

\# Write the dataframe to a csv file

\# write.csv(stats_df, "F:/study2022-2122/chatGPT/AD/Sorted data/stats_1.csv", row.names = FALSE)

\# write.csv(stats_df, "F:/study2022-2122/chatGPT/AD/Sorted data/stats_2.csv", row.names = FALSE)

\# write.csv(stats_df, "F:/study2022-2122/chatGPT/AD/Sorted data/stats_3.csv", row.names = FALSE)

\# Define the output file path

output_file <- "F:/study2022-2122/chatGPT/AD/Sorted data/ROC.pdf"

\# Open a new PDF file

pdf(file = output_file, width = 7, height = 7)

\# Define a vector of colors

\# colors <- brewer.pal(5, "Set1")

\# colors <- brewer.pal(2, "Set1")

\# Plot the AUC-ROC

roc_list <- lapply(model_names, function(model) {

roc(df$Group, df[[model]], plot = FALSE)

})

\# Compute AUC for each model

auc_values <- lapply(roc_list, auc)

\# Create model names with AUC values, now formatted with four decimal places

model_names_with_auc <- paste(model_names, "AUC:", formatC(as.numeric(unlist(auc_values)), format = "f", digits = 4))

\# Plot ROC curve for the first model with reversed xlim parameter and asp = 1

plot(roc_list[[1]], col = colors[1], xlim = c(1, 0))

\# Add ROC curves for the rest of the models

for(i in 2:length(roc_list)) {

lines(roc_list[[i]], col = colors[i])

}

\# Add legend

legend("bottomright", legend = model_names_with_auc, col = colors, lty = 1, cex = 1.2) # Reduce legend size

\# Close the PDF device

dev.off()

**Statistical analysis 1: mean, standard deviation, median, interquartile spacing, minimum and maximum values**

| Group | FN | FP | TN | TP | N | P |
| --- | --- | --- | --- | --- | --- | --- |
| Limited_vocabulary.mean | 2.94 | 5.58 | 3.18 | 4.96 | 3.55 | 4.14 |
| Limited_vocabulary.sd | 1.06 | 1.24 | 1.11 | 1.68 | 1.42 | 1.76 |
| Limited_vocabulary.median | 3.00 | 6.00 | 3.00 | 4.50 | 3.00 | 4.00 |
| Limited_vocabulary.IQR | 1.75 | 1.25 | 2.00 | 3.00 | 1.00 | 2.25 |
| Limited_vocabulary.min | 2.00 | 3.00 | 1.00 | 2.00 | 1.00 | 2.00 |
| Limited_vocabulary.max | 5.00 | 7.00 | 6.00 | 7.00 | 7.00 | 7.00 |
| Increased_verbosity.mean | 2.50 | 4.25 | 2.47 | 3.88 | 2.74 | 3.32 |
| Increased_verbosity.sd | 1.04 | 1.36 | 0.92 | 1.68 | 1.18 | 1.60 |
| Increased_verbosity.median | 2.50 | 4.00 | 2.00 | 3.00 | 2.00 | 3.00 |
| Increased_verbosity.IQR | 1.00 | 2.25 | 1.00 | 2.00 | 1.00 | 2.25 |
| Increased_verbosity.min | 1.00 | 2.00 | 1.00 | 2.00 | 1.00 | 1.00 |
| Increased_verbosity.max | 5.00 | 6.00 | 5.00 | 8.00 | 6.00 | 8.00 |
| Overlearned_phrases.mean | 1.61 | 3.25 | 1.64 | 2.92 | 1.88 | 2.39 |
| Overlearned_phrases.sd | 0.70 | 2.18 | 0.67 | 1.57 | 1.18 | 1.43 |
| Overlearned_phrases.median | 1.50 | 2.00 | 2.00 | 2.50 | 2.00 | 2.00 |
| Overlearned_phrases.IQR | 1.00 | 1.75 | 1.00 | 1.00 | 1.00 | 2.00 |
| Overlearned_phrases.min | 1.00 | 1.00 | 1.00 | 1.00 | 1.00 | 1.00 |
| Overlearned_phrases.max | 3.00 | 8.00 | 4.00 | 7.00 | 8.00 | 7.00 |
| Improper_use_of_pronouns.mean | 1.39 | 2.00 | 1.24 | 2.58 | 1.36 | 2.09 |
| Improper_use_of_pronouns.sd | 0.50 | 1.13 | 0.53 | 1.53 | 0.70 | 1.34 |
| Improper_use_of_pronouns.median | 1.00 | 2.00 | 1.00 | 2.00 | 1.00 | 2.00 |
| Improper_use_of_pronouns.IQR | 1.00 | 1.00 | 0.00 | 1.75 | 1.00 | 2.00 |
| Improper_use_of_pronouns.min | 1.00 | 1.00 | 1.00 | 1.00 | 1.00 | 1.00 |
| Improper_use_of_pronouns.max | 2.00 | 5.00 | 4.00 | 6.00 | 5.00 | 6.00 |
| Hesitation.mean | 4.72 | 6.08 | 4.18 | 6.73 | 4.47 | 5.91 |
| Hesitation.sd | 1.23 | 1.00 | 1.47 | 1.34 | 1.56 | 1.63 |
| Hesitation.median | 4.50 | 6.00 | 4.00 | 7.00 | 4.00 | 6.00 |
| Hesitation.IQR | 1.00 | 1.25 | 2.00 | 2.00 | 3.00 | 3.00 |
| Hesitation.min | 2.00 | 4.00 | 1.00 | 4.00 | 1.00 | 2.00 |
| Hesitation.max | 7.00 | 7.00 | 7.00 | 9.00 | 7.00 | 9.00 |
| Circumlocution.mean | 2.61 | 4.75 | 2.32 | 4.92 | 2.69 | 3.98 |
| Circumlocution.sd | 0.92 | 1.60 | 0.99 | 1.32 | 1.41 | 1.64 |
| Circumlocution.median | 2.50 | 4.50 | 2.00 | 5.00 | 2.00 | 4.00 |
| Circumlocution.IQR | 1.00 | 2.00 | 1.00 | 2.00 | 1.00 | 2.00 |
| Circumlocution.min | 1.00 | 2.00 | 1.00 | 2.00 | 1.00 | 1.00 |
| Circumlocution.max | 5.00 | 8.00 | 6.00 | 7.00 | 8.00 | 7.00 |
| Tip-of-the-tongue_phenomenon.mean | 2.17 | 3.67 | 1.88 | 4.69 | 2.15 | 3.66 |
| Tip-of-the-tongue_phenomenon.sd | 1.04 | 1.15 | 0.79 | 1.57 | 1.07 | 1.85 |
| Tip-of-the-tongue_phenomenon.median | 2.00 | 3.50 | 2.00 | 5.00 | 2.00 | 4.00 |
| Tip-of-the-tongue_phenomenon.IQR | 1.50 | 2.00 | 1.00 | 1.75 | 2.00 | 3.00 |
| Tip-of-the-tongue_phenomenon.min | 1.00 | 2.00 | 1.00 | 1.00 | 1.00 | 1.00 |
| Tip-of-the-tongue_phenomenon.max | 4.00 | 5.00 | 4.00 | 7.00 | 5.00 | 7.00 |
| Repetition.mean | 2.83 | 5.42 | 2.94 | 4.88 | 3.32 | 4.05 |
| Repetition.sd | 0.92 | 1.62 | 1.24 | 1.70 | 1.57 | 1.75 |
| Repetition.median | 3.00 | 6.00 | 3.00 | 4.00 | 3.00 | 4.00 |
| Repetition.IQR | 1.00 | 1.50 | 2.00 | 2.75 | 2.00 | 2.00 |
| Repetition.min | 1.00 | 2.00 | 1.00 | 2.00 | 1.00 | 1.00 |
| Repetition.max | 5.00 | 7.00 | 7.00 | 8.00 | 7.00 | 8.00 |
| Simplified_sentence_structure_and_grammar.mean | 2.72 | 5.17 | 2.92 | 5.35 | 3.27 | 4.27 |
| Simplified_sentence_structure_and_grammar.sd | 0.75 | 1.27 | 1.15 | 1.62 | 1.42 | 1.86 |
| Simplified_sentence_structure_and_grammar.median | 3.00 | 5.00 | 3.00 | 5.50 | 3.00 | 4.00 |
| Simplified_sentence_structure_and_grammar.IQR | 1.00 | 2.00 | 1.00 | 3.00 | 2.00 | 3.00 |
| Simplified_sentence_structure_and_grammar.min | 2.00 | 3.00 | 1.00 | 3.00 | 1.00 | 2.00 |
| Simplified_sentence_structure_and_grammar.max | 4.00 | 7.00 | 7.00 | 8.00 | 7.00 | 8.00 |
| Grammatical_errors.mean | 1.56 | 2.92 | 1.55 | 2.77 | 1.76 | 2.27 |
| Grammatical_errors.sd | 0.62 | 1.31 | 0.81 | 1.11 | 1.02 | 1.11 |
| Grammatical_errors.median | 1.50 | 2.50 | 1.00 | 2.50 | 2.00 | 2.00 |
| Grammatical_errors.IQR | 1.00 | 2.00 | 1.00 | 1.00 | 1.00 | 1.00 |
| Grammatical_errors.min | 1.00 | 1.00 | 1.00 | 1.00 | 1.00 | 1.00 |
| Grammatical_errors.max | 3.00 | 5.00 | 6.00 | 6.00 | 6.00 | 6.00 |
| Difficulty_with_complex_ideas.mean | 2.33 | 4.33 | 2.14 | 5.27 | 2.47 | 4.07 |
| Difficulty_with_complex_ideas.sd | 0.69 | 1.23 | 0.70 | 1.12 | 1.12 | 1.74 |
| Difficulty_with_complex_ideas.median | 2.00 | 4.50 | 2.00 | 5.00 | 2.00 | 4.00 |
| Difficulty_with_complex_ideas.IQR | 1.00 | 1.25 | 0.00 | 1.00 | 1.00 | 3.00 |
| Difficulty_with_complex_ideas.min | 1.00 | 2.00 | 1.00 | 4.00 | 1.00 | 1.00 |
| Difficulty_with_complex_ideas.max | 4.00 | 6.00 | 4.00 | 8.00 | 6.00 | 8.00 |
| Semantic_paraphasias.mean | 1.89 | 2.92 | 1.68 | 3.65 | 1.87 | 2.93 |
| Semantic_paraphasias.sd | 0.83 | 1.16 | 0.83 | 1.41 | 0.99 | 1.48 |
| Semantic_paraphasias.median | 2.00 | 3.00 | 2.00 | 3.00 | 2.00 | 3.00 |
| Semantic_paraphasias.IQR | 1.00 | 2.00 | 1.00 | 1.75 | 1.00 | 2.00 |
| Semantic_paraphasias.min | 1.00 | 1.00 | 1.00 | 1.00 | 1.00 | 1.00 |
| Semantic_paraphasias.max | 4.00 | 5.00 | 6.00 | 6.00 | 6.00 | 6.00 |
| Lack_of_coherence.mean | 2.17 | 4.33 | 1.89 | 4.69 | 2.27 | 3.66 |
| Lack_of_coherence.sd | 0.71 | 1.15 | 0.73 | 1.44 | 1.19 | 1.72 |
| Lack_of_coherence.median | 2.00 | 4.50 | 2.00 | 5.00 | 2.00 | 3.00 |
| Lack_of_coherence.IQR | 1.00 | 2.00 | 1.00 | 1.00 | 1.75 | 3.00 |
| Lack_of_coherence.min | 1.00 | 3.00 | 1.00 | 2.00 | 1.00 | 1.00 |
| Lack_of_coherence.max | 3.00 | 6.00 | 4.00 | 7.00 | 6.00 | 7.00 |
| Consistency_in_errors.mean | 1.44 | 3.50 | 1.29 | 3.58 | 1.63 | 2.70 |
| Consistency_in_errors.sd | 0.62 | 1.17 | 0.49 | 1.70 | 1.02 | 1.72 |
| Consistency_in_errors.median | 1.00 | 3.50 | 1.00 | 4.00 | 1.00 | 2.00 |
| Consistency_in_errors.IQR | 1.00 | 1.50 | 1.00 | 2.75 | 1.00 | 3.00 |
| Consistency_in_errors.min | 1.00 | 2.00 | 1.00 | 1.00 | 1.00 | 1.00 |
| Consistency_in_errors.max | 3.00 | 5.00 | 3.00 | 7.00 | 5.00 | 7.00 |
| Lack_of_semantic_fluency.mean | 2.56 | 4.67 | 2.08 | 5.04 | 2.47 | 4.02 |
| Lack_of_semantic_fluency.sd | 1.10 | 1.15 | 0.85 | 1.25 | 1.30 | 1.70 |
| Lack_of_semantic_fluency.median | 2.00 | 5.00 | 2.00 | 5.00 | 2.00 | 4.00 |
| Lack_of_semantic_fluency.IQR | 1.00 | 0.25 | 1.75 | 2.00 | 1.00 | 2.25 |
| Lack_of_semantic_fluency.min | 1.00 | 2.00 | 1.00 | 3.00 | 1.00 | 1.00 |
| Lack_of_semantic_fluency.max | 5.00 | 6.00 | 5.00 | 7.00 | 6.00 | 7.00 |
| Disorganized_narrative_structure.mean | 2.28 | 4.75 | 2.00 | 4.81 | 2.42 | 3.77 |
| Disorganized_narrative_structure.sd | 0.83 | 0.97 | 0.78 | 1.23 | 1.28 | 1.65 |
| Disorganized_narrative_structure.median | 2.00 | 5.00 | 2.00 | 5.00 | 2.00 | 4.00 |
| Disorganized_narrative_structure.IQR | 1.00 | 1.25 | 1.00 | 1.75 | 1.00 | 3.00 |
| Disorganized_narrative_structure.min | 1.00 | 3.00 | 1.00 | 2.00 | 1.00 | 1.00 |
| Disorganized_narrative_structure.max | 4.00 | 6.00 | 4.00 | 7.00 | 6.00 | 7.00 |
| Orientation_issues.mean | 1.22 | 2.25 | 1.11 | 2.92 | 1.28 | 2.23 |
| Orientation_issues.sd | 0.43 | 0.97 | 0.31 | 1.79 | 0.62 | 1.63 |
| Orientation_issues.median | 1.00 | 2.00 | 1.00 | 3.00 | 1.00 | 2.00 |
| Orientation_issues.IQR | 0.00 | 0.25 | 0.00 | 2.50 | 0.00 | 2.00 |
| Orientation_issues.min | 1.00 | 1.00 | 1.00 | 1.00 | 1.00 | 1.00 |
| Orientation_issues.max | 2.00 | 4.00 | 2.00 | 8.00 | 4.00 | 8.00 |
| Decline_in_conversational_engagement_and_responsiveness.mean | 1.72 | 3.50 | 1.76 | 3.77 | 2.03 | 2.93 |
| Decline_in_conversational_engagement_and_responsiveness.sd | 0.57 | 1.17 | 0.72 | 1.39 | 1.02 | 1.52 |
| Decline_in_conversational_engagement_and_responsiveness.median | 2.00 | 3.00 | 2.00 | 3.00 | 2.00 | 3.00 |
| Decline_in_conversational_engagement_and_responsiveness.IQR | 1.00 | 1.00 | 1.00 | 2.00 | 1.00 | 2.00 |
| Decline_in_conversational_engagement_and_responsiveness.min | 1.00 | 2.00 | 1.00 | 1.00 | 1.00 | 1.00 |
| Decline_in_conversational_engagement_and_responsiveness.max | 3.00 | 6.00 | 4.00 | 7.00 | 6.00 | 7.00 |
| Memory_issues.mean | 2.67 | 4.17 | 2.12 | 5.50 | 2.44 | 4.34 |
| Memory_issues.sd | 1.08 | 1.99 | 1.06 | 1.90 | 1.44 | 2.13 |
| Memory_issues.median | 2.50 | 4.00 | 2.00 | 6.00 | 2.00 | 4.00 |
| Memory_issues.IQR | 1.00 | 4.00 | 2.00 | 2.50 | 2.00 | 4.00 |
| Memory_issues.min | 1.00 | 2.00 | 1.00 | 2.00 | 1.00 | 1.00 |
| Memory_issues.max | 5.00 | 7.00 | 5.00 | 10.00 | 7.00 | 10.00 |
| Lack_of_abstract_thinking_and_synthesis.mean | 2.06 | 4.25 | 1.88 | 4.73 | 2.24 | 3.64 |
| Lack_of_abstract_thinking_and_synthesis.sd | 0.73 | 1.22 | 0.71 | 1.54 | 1.18 | 1.83 |
| Lack_of_abstract_thinking_and_synthesis.median | 2.00 | 4.00 | 2.00 | 5.00 | 2.00 | 3.00 |
| Lack_of_abstract_thinking_and_synthesis.IQR | 0.00 | 2.25 | 1.00 | 2.00 | 2.00 | 3.00 |
| Lack_of_abstract_thinking_and_synthesis.min | 1.00 | 3.00 | 1.00 | 1.00 | 1.00 | 1.00 |
| Lack_of_abstract_thinking_and_synthesis.max | 4.00 | 6.00 | 3.00 | 7.00 | 6.00 | 7.00 |
| Abnormal_emotions.mean | 1.06 | 1.92 | 1.14 | 2.00 | 1.26 | 1.61 |
| Abnormal_emotions.sd | 0.24 | 1.00 | 0.35 | 1.10 | 0.57 | 0.97 |
| Abnormal_emotions.median | 1.00 | 2.00 | 1.00 | 2.00 | 1.00 | 1.00 |
| Abnormal_emotions.IQR | 0.00 | 1.25 | 0.00 | 1.00 | 0.00 | 1.00 |
| Abnormal_emotions.min | 1.00 | 1.00 | 1.00 | 1.00 | 1.00 | 1.00 |
| Abnormal_emotions.max | 2.00 | 4.00 | 2.00 | 6.00 | 4.00 | 6.00 |

**Statistical analysis 2: normal distribution and Homogeneity of variance test, Independent samples t-test for the two groups, Mann-Whitney U test for the two groups**

| Variable | Shapiro Wilk p value | Levene p value (4groups) | Levene p value (2groups) | t test p value | Wilcox test p value |
| --- | --- | --- | --- | --- | --- |
| Limited_vocabulary | 2.05E-08 | 0.001709 | 0.072775 | 0.063193 | 0.084555 |
| Increased_verbosity | 2.94E-09 | 0.011126 | 0.250229 | 0.040403 | 0.046519 |
| Overlearned_phrases | 3.28E-14 | 0.0026 | 0.200604 | 0.052202 | 0.021069 |
| Improper_use_of_pronouns | 4.45E-16 | 2.47E-05 | 0.000492 | 0.001387 | 0.000148 |
| Hesitation | 0.000625 | 0.456978 | 0.898278 | 8.07E-06 | 1.97E-05 |
| Circumlocution | 3.31E-08 | 0.042575 | 0.070296 | 3.65E-05 | 1.09E-05 |
| Tip-of-the-tongue_phenomenon | 5.87E-09 | 0.008003 | 1.61E-05 | 6.68E-06 | 9.99E-06 |
| Repetition | 5.56E-07 | 0.044777 | 0.39898 | 0.025511 | 0.021743 |
| Simplified_sentence_structure_and_grammar | 1.07E-08 | 0.001791 | 0.015923 | 0.002728 | 0.002846 |
| Grammatical_errors | 1.63E-12 | 0.061091 | 0.981456 | 0.012865 | 0.001877 |
| Difficulty_with_complex_ideas | 1.29E-09 | 0.016002 | 0.000157 | 8.56E-07 | 3.57E-07 |
| Semantic_paraphasias | 5.56E-11 | 0.017012 | 0.00147 | 7.28E-05 | 2.05E-05 |
| Lack_of_coherence | 2.97E-09 | 0.000503 | 0.00095 | 1.15E-05 | 4.79E-06 |
| Consistency_in_errors | 1.83E-13 | 4.27E-09 | 0.000979 | 0.000347 | 0.000152 |
| Lack_of_semantic_fluency | 6.6E-08 | 0.185818 | 0.013909 | 1.61E-06 | 1.36E-06 |
| Disorganized_narrative_structure | 6.99E-08 | 0.038433 | 0.025199 | 1.33E-05 | 7.45E-06 |
| Orientation_issues | 8.23E-17 | 4.44E-11 | 9.05E-08 | 0.000526 | 7.7E-05 |
| Decline_in_conversational_engagement_and_responsiveness | 2.95E-10 | 0.00597 | 0.002418 | 0.000743 | 0.000419 |
| Memory_issues | 1.16E-08 | 0.001493 | 0.000854 | 1.56E-06 | 5.33E-07 |
| Lack_of_abstract_thinking_and_synthesis | 8.35E-10 | 7.41E-05 | 2.33E-05 | 2.53E-05 | 3.44E-05 |
| Abnormal_emotions | 1.34E-17 | 1.42E-05 | 0.011444 | 0.029043 | 0.007973 |

**Statistical analysis 3 and Violin plots: one-way ANOVA for the four groups (Tukey's Honestly Significant Difference test as a post hoc test), and Kruskal-Wallis H-test for the four groups (Dunn's test was applied as a post hoc test)**

| Variable | Comparison | ANOVA p value | Tukey p value | Kruskal Wallis p value | Dunn p value |
| --- | --- | --- | --- | --- | --- |
| Limited_vocabulary | FP-FN | 5.43E-12 | 7.38E-07 | 9.55E-09 | 1.41E-05 |
| Limited_vocabulary | TN-FN | 5.43E-12 | 0.893038 | 9.55E-09 | 0.462826 |
| Limited_vocabulary | TP-FN | 5.43E-12 | 4.40E-06 | 9.55E-09 | 5.77E-06 |
| Limited_vocabulary | TN-FP | 5.43E-12 | 8.89E-08 | 9.55E-09 | 4.43E-05 |
| Limited_vocabulary | TP-FP | 5.43E-12 | 0.491525 | 9.55E-09 | 0.29404 |
| Limited_vocabulary | TP-TN | 5.43E-12 | 7.83E-08 | 9.55E-09 | 5.01E-06 |
| Increased_verbosity | FP-FN | 2.89E-08 | 0.00067 | 2.4E-06 | 0.000986 |
| Increased_verbosity | TN-FN | 2.89E-08 | 0.999674 | 2.4E-06 | 0.826812 |
| Increased_verbosity | TP-FN | 2.89E-08 | 0.001162 | 2.4E-06 | 4.17E-05 |
| Increased_verbosity | TN-FP | 2.89E-08 | 2.6E-05 | 2.4E-06 | 0.004439 |
| Increased_verbosity | TP-FP | 2.89E-08 | 0.810829 | 2.4E-06 | 0.308544 |
| Increased_verbosity | TP-TN | 2.89E-08 | 5.35E-06 | 2.4E-06 | 5.85E-05 |
| Overlearned_phrases | FP-FN | 1.32E-07 | 0.000983 | 4.39E-06 | 0.005053 |
| Overlearned_phrases | TN-FN | 1.32E-07 | 0.999788 | 4.39E-06 | 0.898141 |
| Overlearned_phrases | TP-FN | 1.32E-07 | 0.001425 | 4.39E-06 | 0.001278 |
| Overlearned_phrases | TN-FP | 1.32E-07 | 8.12E-05 | 4.39E-06 | 0.000562 |
| Overlearned_phrases | TP-FP | 1.32E-07 | 0.841885 | 4.39E-06 | 0.970642 |
| Overlearned_phrases | TP-TN | 1.32E-07 | 1.8E-05 | 4.39E-06 | 9.82E-06 |
| Improper_use_of_pronouns | FP-FN | 2.72E-08 | 0.264526 | 4.23E-07 | 0.098426 |
| Improper_use_of_pronouns | TN-FN | 2.72E-08 | 0.92722 | 4.23E-07 | 0.268252 |
| Improper_use_of_pronouns | TP-FN | 2.72E-08 | 0.000187 | 4.23E-07 | 0.003725 |
| Improper_use_of_pronouns | TN-FP | 2.72E-08 | 0.039846 | 4.23E-07 | 0.001939 |
| Improper_use_of_pronouns | TP-FP | 2.72E-08 | 0.257754 | 4.23E-07 | 0.337871 |
| Improper_use_of_pronouns | TP-TN | 2.72E-08 | 1.68E-08 | 4.23E-07 | 7.63E-08 |
| Hesitation | FP-FN | 2.29E-12 | 0.042874 | 6.62E-10 | 0.022594 |
| Hesitation | TN-FN | 2.29E-12 | 0.450687 | 6.62E-10 | 0.23827 |
| Hesitation | TP-FN | 2.29E-12 | 2.98E-05 | 6.62E-10 | 0.00021 |
| Hesitation | TN-FP | 2.29E-12 | 0.000127 | 6.62E-10 | 0.000214 |
| Hesitation | TP-FP | 2.29E-12 | 0.530553 | 6.62E-10 | 0.413858 |
| Hesitation | TP-TN | 2.29E-12 | 4.79E-12 | 6.62E-10 | 3.95E-10 |
| Circumlocution | FP-FN | 9.71E-19 | 8.51E-06 | 6.1E-13 | 0.001332 |
| Circumlocution | TN-FN | 9.71E-19 | 0.764025 | 6.1E-13 | 0.298389 |
| Circumlocution | TP-FN | 9.71E-19 | 5.15E-09 | 6.1E-13 | 2.7E-06 |
| Circumlocution | TN-FP | 9.71E-19 | 2.06E-09 | 6.1E-13 | 1.67E-05 |
| Circumlocution | TP-FP | 9.71E-19 | 0.971612 | 6.1E-13 | 0.72239 |
| Circumlocution | TP-TN | 9.71E-19 | 8.22E-15 | 6.1E-13 | 5.39E-12 |
| Tip-of-the-tongue_phenomenon | FP-FN | 3.24E-20 | 0.001537 | 4.06E-12 | 0.004859 |
| Tip-of-the-tongue_phenomenon | TN-FN | 3.24E-20 | 0.744236 | 4.06E-12 | 0.440409 |
| Tip-of-the-tongue_phenomenon | TP-FN | 3.24E-20 | 3.07E-11 | 4.06E-12 | 6.39E-05 |
| Tip-of-the-tongue_phenomenon | TN-FP | 3.24E-20 | 3.08E-06 | 4.06E-12 | 5.82E-06 |
| Tip-of-the-tongue_phenomenon | TP-FP | 3.24E-20 | 0.035276 | 4.06E-12 | 0.329547 |
| Tip-of-the-tongue_phenomenon | TP-TN | 3.24E-20 | 0 | 4.06E-12 | 5.64E-12 |
| Repetition | FP-FN | 1.8E-11 | 6.95E-06 | 3.08E-08 | 0.000203 |
| Repetition | TN-FN | 1.8E-11 | 0.991032 | 3.08E-08 | 0.843724 |
| Repetition | TP-FN | 1.8E-11 | 1.49E-05 | 3.08E-08 | 2.19E-05 |
| Repetition | TN-FP | 1.8E-11 | 2.85E-07 | 3.08E-08 | 0.000138 |
| Repetition | TP-FP | 1.8E-11 | 0.673485 | 3.08E-08 | 0.536145 |
| Repetition | TP-TN | 1.8E-11 | 4.85E-08 | 3.08E-08 | 1.43E-06 |
| Simplified_sentence_structure_and_grammar | FP-FN | 1.07E-15 | 3.05E-06 | 1.73E-11 | 4.78E-05 |
| Simplified_sentence_structure_and_grammar | TN-FN | 1.07E-15 | 0.926962 | 1.73E-11 | 0.670178 |
| Simplified_sentence_structure_and_grammar | TP-FN | 1.07E-15 | 1.41E-09 | 1.73E-11 | 7.89E-06 |
| Simplified_sentence_structure_and_grammar | TN-FP | 1.07E-15 | 3.56E-07 | 1.73E-11 | 8.5E-07 |
| Simplified_sentence_structure_and_grammar | TP-FP | 1.07E-15 | 0.97553 | 1.73E-11 | 0.986796 |
| Simplified_sentence_structure_and_grammar | TP-TN | 1.07E-15 | 4.79E-13 | 1.73E-11 | 1.63E-09 |
| Grammatical_errors | FP-FN | 3.37E-09 | 0.000633 | 8.9E-09 | 0.001736 |
| Grammatical_errors | TN-FN | 3.37E-09 | 0.999974 | 8.9E-09 | 0.769719 |
| Grammatical_errors | TP-FN | 3.37E-09 | 0.000179 | 8.9E-09 | 7.26E-05 |
| Grammatical_errors | TN-FP | 3.37E-09 | 2.93E-05 | 8.9E-09 | 0.000111 |
| Grammatical_errors | TP-FP | 3.37E-09 | 0.966984 | 8.9E-09 | 0.958666 |
| Grammatical_errors | TP-TN | 3.37E-09 | 3.55E-07 | 8.9E-09 | 4.88E-08 |
| Difficulty_with_complex_ideas | FP-FN | 2.17E-31 | 4.44E-08 | 7.05E-17 | 0.000547 |
| Difficulty_with_complex_ideas | TN-FN | 2.17E-31 | 0.825398 | 7.05E-17 | 0.459448 |
| Difficulty_with_complex_ideas | TP-FN | 2.17E-31 | 0 | 7.05E-17 | 2.22E-06 |
| Difficulty_with_complex_ideas | TN-FP | 2.17E-31 | 2.96E-12 | 7.05E-17 | 4.36E-08 |
| Difficulty_with_complex_ideas | TP-FP | 2.17E-31 | 0.012272 | 7.05E-17 | 0.262988 |
| Difficulty_with_complex_ideas | TP-TN | 2.17E-31 | 0 | 7.05E-17 | 5.47E-16 |
| Semantic_paraphasias | FP-FN | 5.07E-13 | 0.036766 | 4.06E-10 | 0.021829 |
| Semantic_paraphasias | TN-FN | 5.07E-13 | 0.868117 | 4.06E-10 | 0.369032 |
| Semantic_paraphasias | TP-FN | 5.07E-13 | 5.7E-07 | 4.06E-10 | 0.000493 |
| Semantic_paraphasias | TN-FP | 5.07E-13 | 0.000954 | 4.06E-10 | 5.21E-05 |
| Semantic_paraphasias | TP-FP | 5.07E-13 | 0.163396 | 4.06E-10 | 0.268824 |
| Semantic_paraphasias | TP-TN | 5.07E-13 | 6.53E-13 | 4.06E-10 | 1.67E-10 |
| Lack_of_coherence | FP-FN | 4.17E-25 | 1.05E-07 | 5.15E-15 | 0.000254 |
| Lack_of_coherence | TN-FN | 4.17E-25 | 0.710031 | 5.15E-15 | 0.290075 |
| Lack_of_coherence | TP-FN | 4.17E-25 | 2.89E-13 | 5.15E-15 | 1.6E-07 |
| Lack_of_coherence | TN-FP | 4.17E-25 | 3.61E-12 | 5.15E-15 | 4.11E-06 |
| Lack_of_coherence | TP-FP | 4.17E-25 | 0.708177 | 5.15E-15 | 0.88801 |
| Lack_of_coherence | TP-TN | 4.17E-25 | 0 | 5.15E-15 | 2.59E-13 |
| Consistency_in_errors | FP-FN | 4.64E-20 | 4.66E-07 | 2.07E-13 | 5.71E-05 |
| Consistency_in_errors | TN-FN | 4.64E-20 | 0.928176 | 2.07E-13 | 0.471051 |
| Consistency_in_errors | TP-FN | 4.64E-20 | 3.2E-10 | 2.07E-13 | 7.05E-08 |
| Consistency_in_errors | TN-FP | 4.64E-20 | 1.94E-10 | 2.07E-13 | 1.53E-05 |
| Consistency_in_errors | TP-FP | 4.64E-20 | 0.995746 | 2.07E-13 | 0.618 |
| Consistency_in_errors | TP-TN | 4.64E-20 | 7.22E-15 | 2.07E-13 | 5.62E-11 |
| Lack_of_semantic_fluency | FP-FN | 5.09E-25 | 8.42E-07 | 7.67E-15 | 0.000915 |
| Lack_of_semantic_fluency | TN-FN | 5.09E-25 | 0.286092 | 7.67E-15 | 0.210409 |
| Lack_of_semantic_fluency | TP-FN | 5.09E-25 | 5.57E-12 | 7.67E-15 | 5.78E-07 |
| Lack_of_semantic_fluency | TN-FP | 5.09E-25 | 2.44E-12 | 7.67E-15 | 6.08E-06 |
| Lack_of_semantic_fluency | TP-FP | 5.09E-25 | 0.718379 | 7.67E-15 | 0.664401 |
| Lack_of_semantic_fluency | TP-TN | 5.09E-25 | 0 | 7.67E-15 | 1.1E-13 |
| Disorganized_narrative_structure | FP-FN | 1.05E-27 | 3.47E-10 | 5.91E-16 | 5.71E-05 |
| Disorganized_narrative_structure | TN-FN | 1.05E-27 | 0.668492 | 5.91E-16 | 0.359106 |
| Disorganized_narrative_structure | TP-FN | 1.05E-27 | 4.43E-14 | 5.91E-16 | 2.76E-08 |
| Disorganized_narrative_structure | TN-FP | 1.05E-27 | 1.43E-14 | 5.91E-16 | 1.59E-06 |
| Disorganized_narrative_structure | TP-FP | 1.05E-27 | 0.99793 | 5.91E-16 | 0.935991 |
| Disorganized_narrative_structure | TP-TN | 1.05E-27 | 0 | 5.91E-16 | 1.27E-13 |
| Orientation_issues | FP-FN | 1.67E-13 | 0.016941 | 6.91E-12 | 0.000792 |
| Orientation_issues | TN-FN | 1.67E-13 | 0.964278 | 6.91E-12 | 0.439353 |
| Orientation_issues | TP-FN | 1.67E-13 | 1.09E-07 | 6.91E-12 | 3.48E-06 |
| Orientation_issues | TN-FP | 1.67E-13 | 0.000707 | 6.91E-12 | 2.45E-05 |
| Orientation_issues | TP-FP | 1.67E-13 | 0.158896 | 6.91E-12 | 0.901719 |
| Orientation_issues | TP-TN | 1.67E-13 | 3.27E-13 | 6.91E-12 | 9.48E-11 |
| Decline_in_conversational_engagement_and_responsiveness | FP-FN | 4.33E-17 | 7.85E-06 | 1.31E-12 | 9.18E-05 |
| Decline_in_conversational_engagement_and_responsiveness | TN-FN | 4.33E-17 | 0.998972 | 1.31E-12 | 0.962519 |
| Decline_in_conversational_engagement_and_responsiveness | TP-FN | 4.33E-17 | 5.1E-10 | 1.31E-12 | 4.12E-06 |
| Decline_in_conversational_engagement_and_responsiveness | TN-FP | 4.33E-17 | 1.82E-07 | 1.31E-12 | 6.69E-07 |
| Decline_in_conversational_engagement_and_responsiveness | TP-FP | 4.33E-17 | 0.842846 | 1.31E-12 | 0.849353 |
| Decline_in_conversational_engagement_and_responsiveness | TP-TN | 4.33E-17 | 1.77E-14 | 1.31E-12 | 6.68E-11 |
| Memory_issues | FP-FN | 7.57E-18 | 0.022738 | 3.55E-11 | 0.077295 |
| Memory_issues | TN-FN | 7.57E-18 | 0.4538 | 3.55E-11 | 0.120634 |
| Memory_issues | TP-FN | 7.57E-18 | 5.49E-09 | 3.55E-11 | 0.000642 |
| Memory_issues | TN-FP | 7.57E-18 | 4.22E-05 | 3.55E-11 | 0.000117 |
| Memory_issues | TP-FP | 7.57E-18 | 0.034054 | 3.55E-11 | 0.134181 |
| Memory_issues | TP-TN | 7.57E-18 | 3.77E-15 | 3.55E-11 | 5.84E-12 |
| Lack_of_abstract_thinking_and_synthesis | FP-FN | 2.24E-24 | 2.07E-07 | 3.34E-14 | 0.000124 |
| Lack_of_abstract_thinking_and_synthesis | TN-FN | 2.24E-24 | 0.90943 | 3.34E-14 | 0.569888 |
| Lack_of_abstract_thinking_and_synthesis | TP-FN | 2.24E-24 | 1.18E-13 | 3.34E-14 | 4.68E-07 |
| Lack_of_abstract_thinking_and_synthesis | TN-FP | 2.24E-24 | 5.25E-11 | 3.34E-14 | 1.42E-06 |
| Lack_of_abstract_thinking_and_synthesis | TP-FP | 2.24E-24 | 0.513435 | 3.34E-14 | 0.890096 |
| Lack_of_abstract_thinking_and_synthesis | TP-TN | 2.24E-24 | 0 | 3.34E-14 | 1.95E-12 |
| Abnormal_emotions | FP-FN | 1.87E-08 | 0.002946 | 1.18E-08 | 0.0009 |
| Abnormal_emotions | TN-FN | 1.87E-08 | 0.965796 | 1.18E-08 | 0.532032 |
| Abnormal_emotions | TP-FN | 1.87E-08 | 3.41E-05 | 1.18E-08 | 0.000642 |
| Abnormal_emotions | TN-FP | 1.87E-08 | 0.001156 | 1.18E-08 | 4.41E-06 |
| Abnormal_emotions | TP-FP | 1.87E-08 | 0.982871 | 1.18E-08 | 0.625274 |
| Abnormal_emotions | TP-TN | 1.87E-08 | 4.24E-07 | 1.18E-08 | 8.23E-08 |

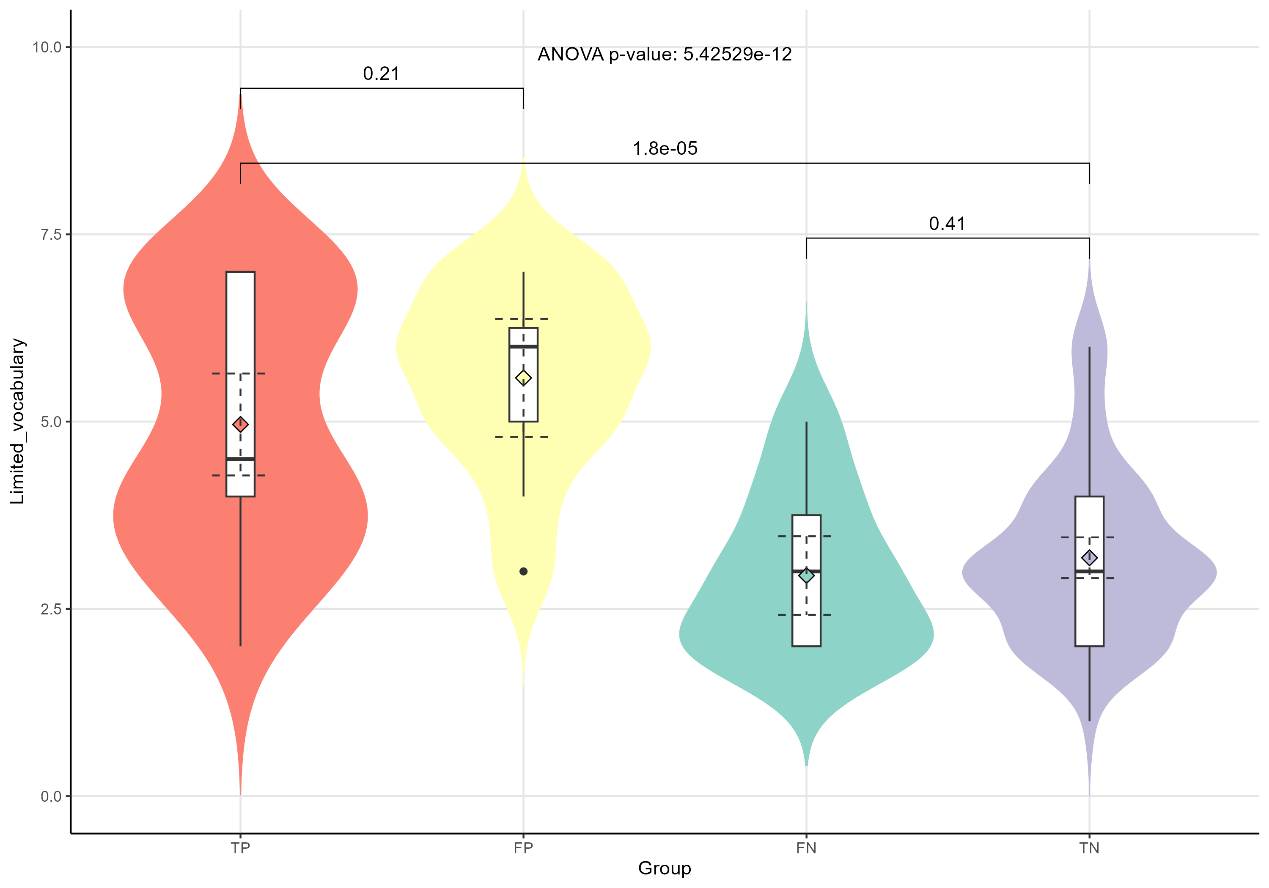

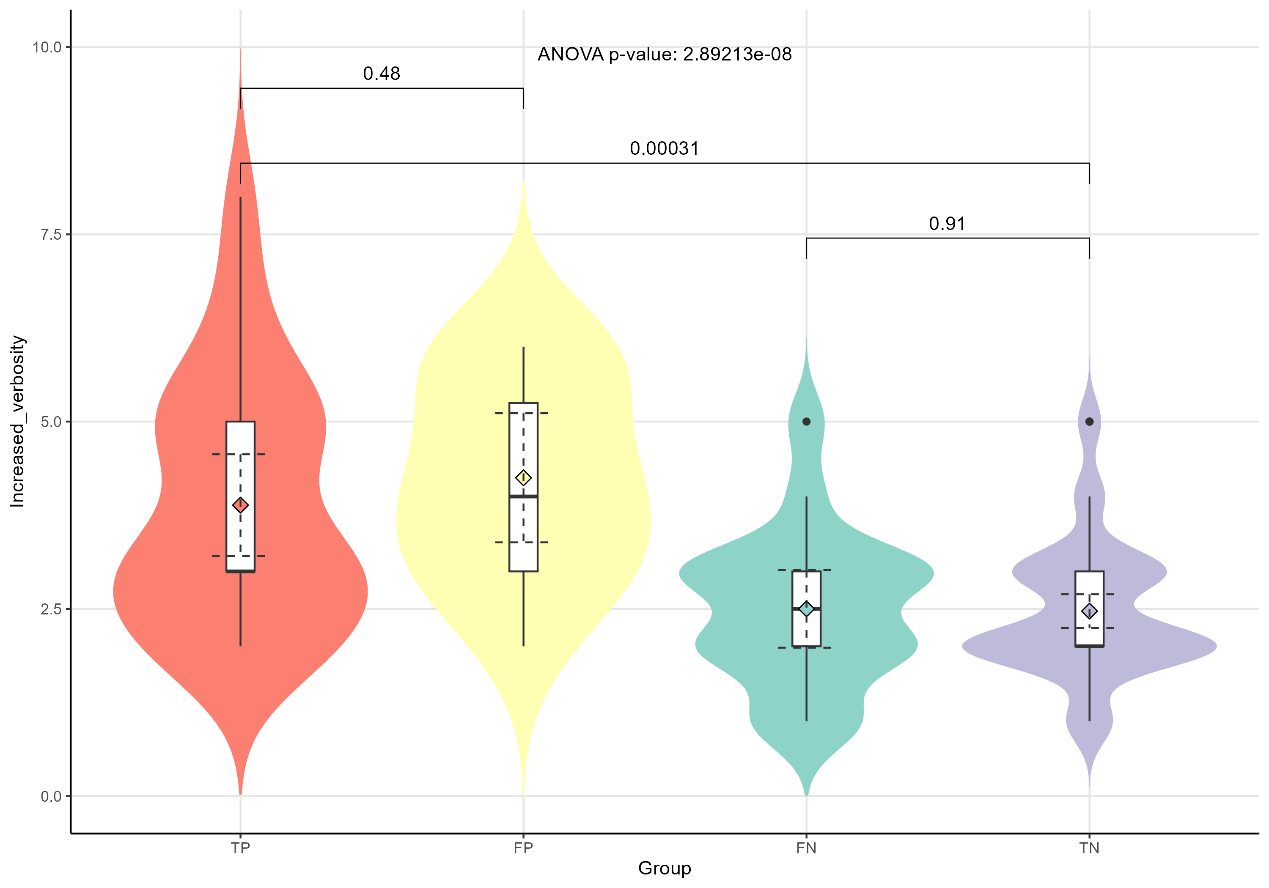

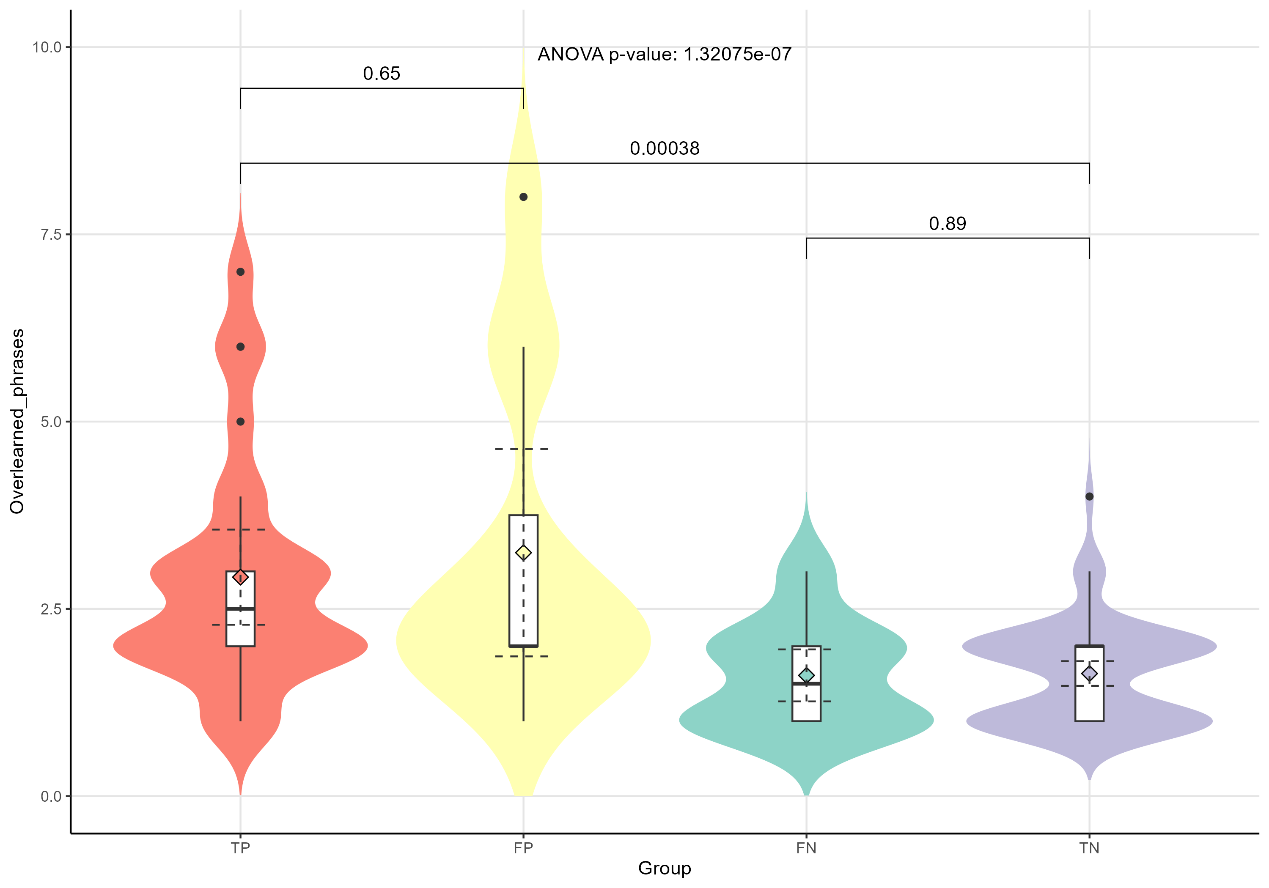

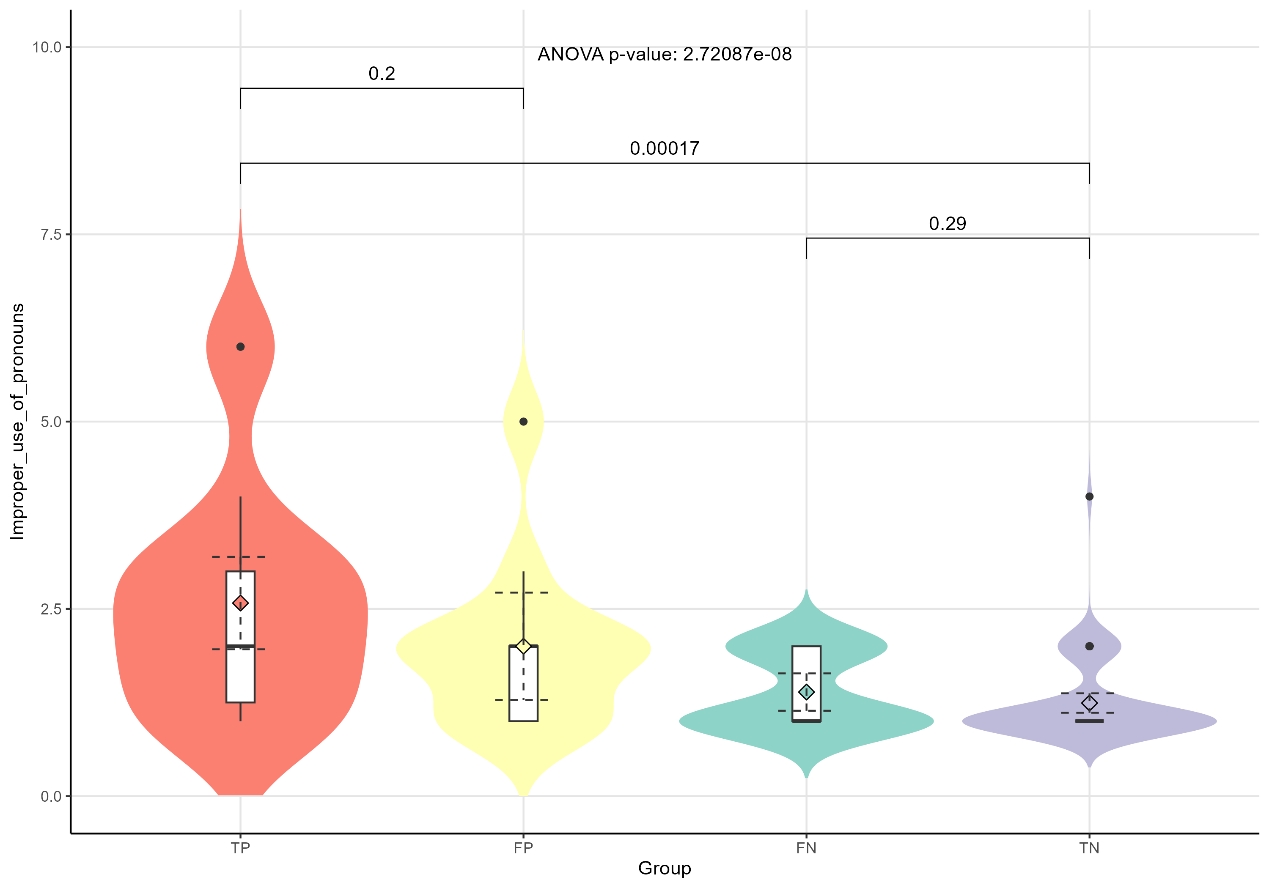

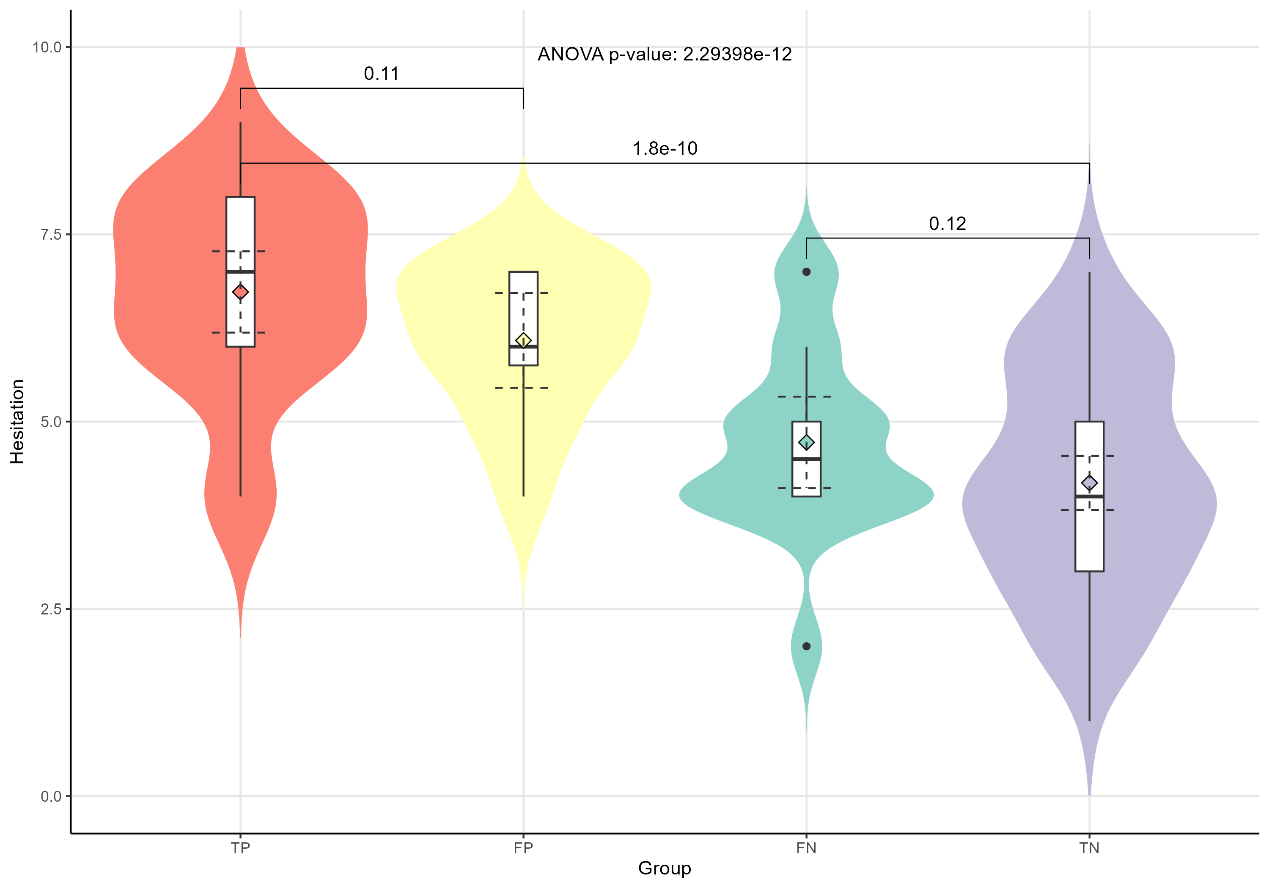

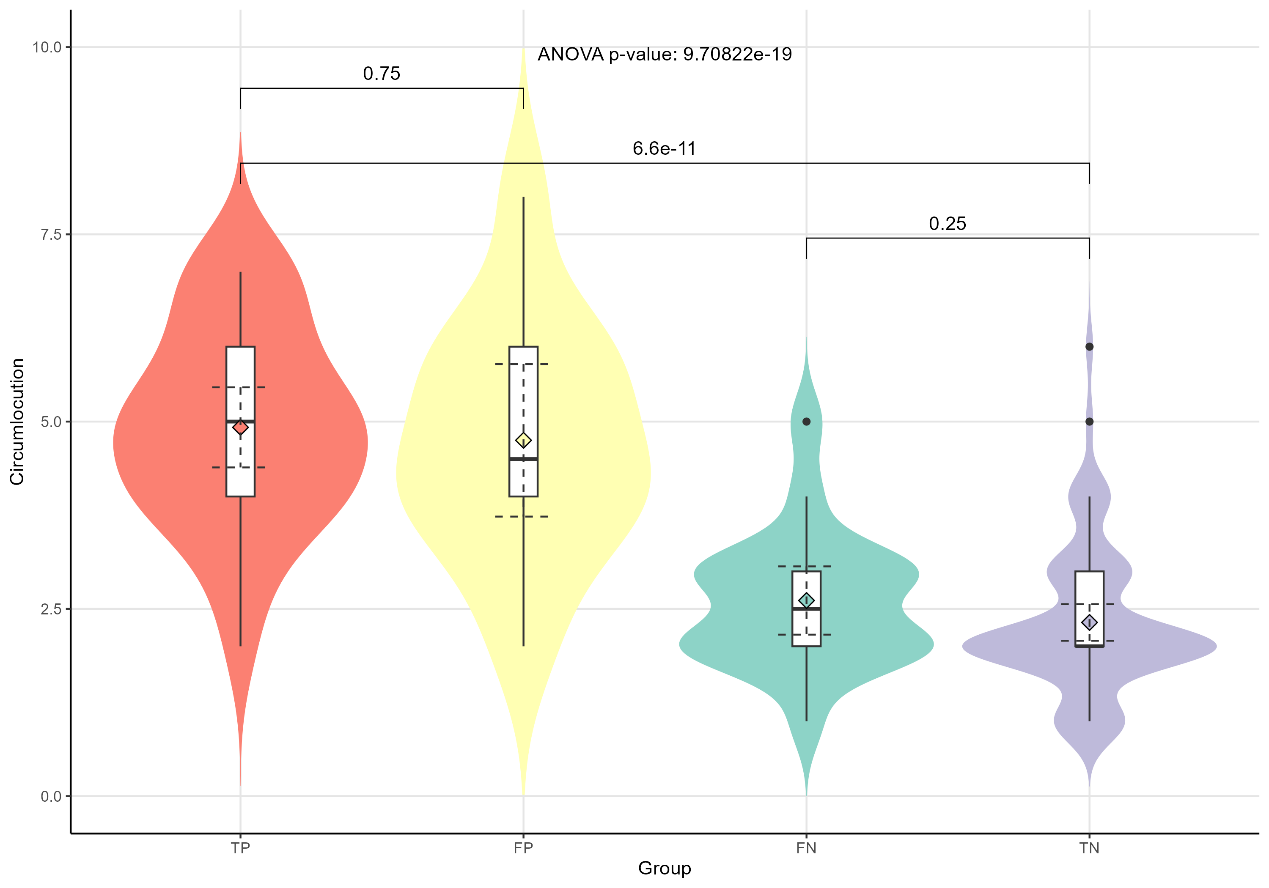

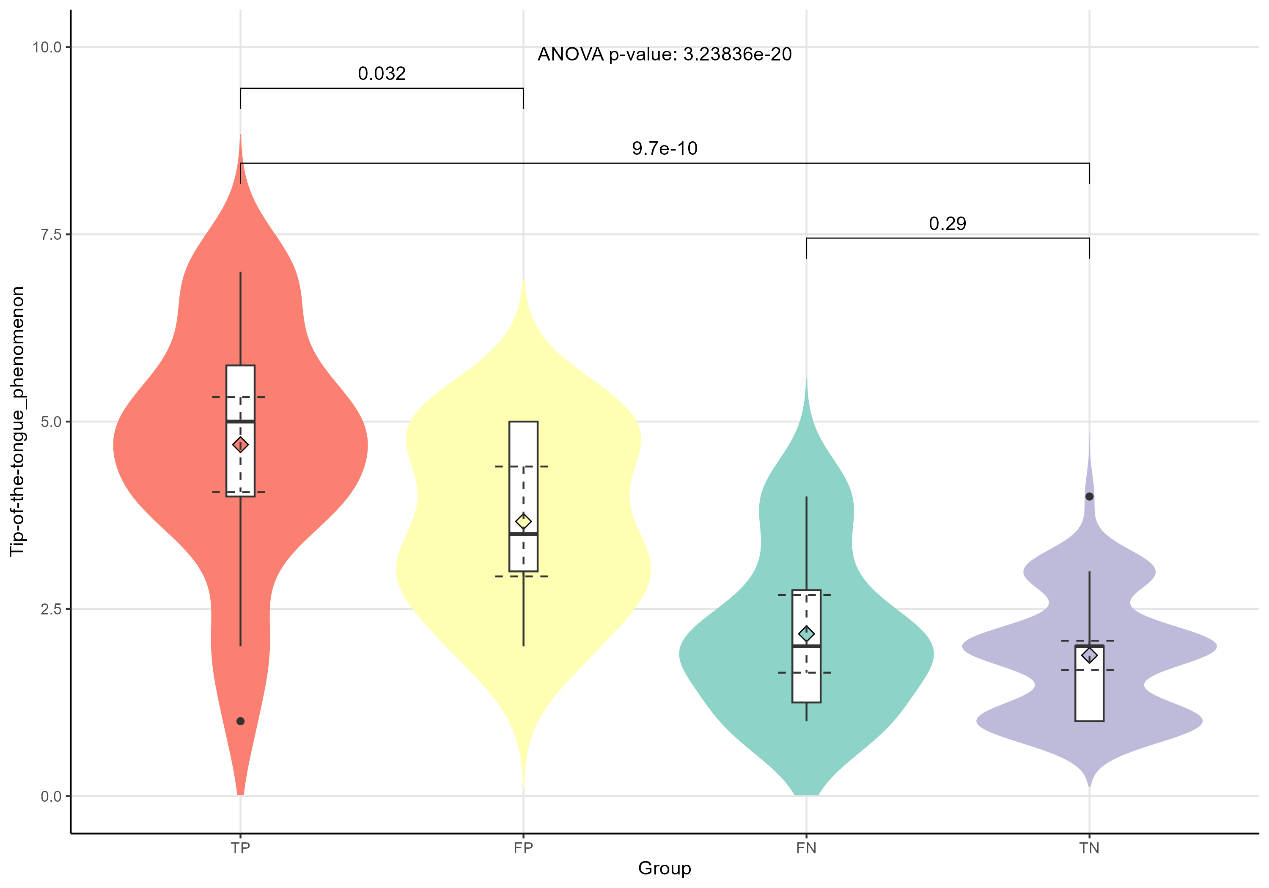

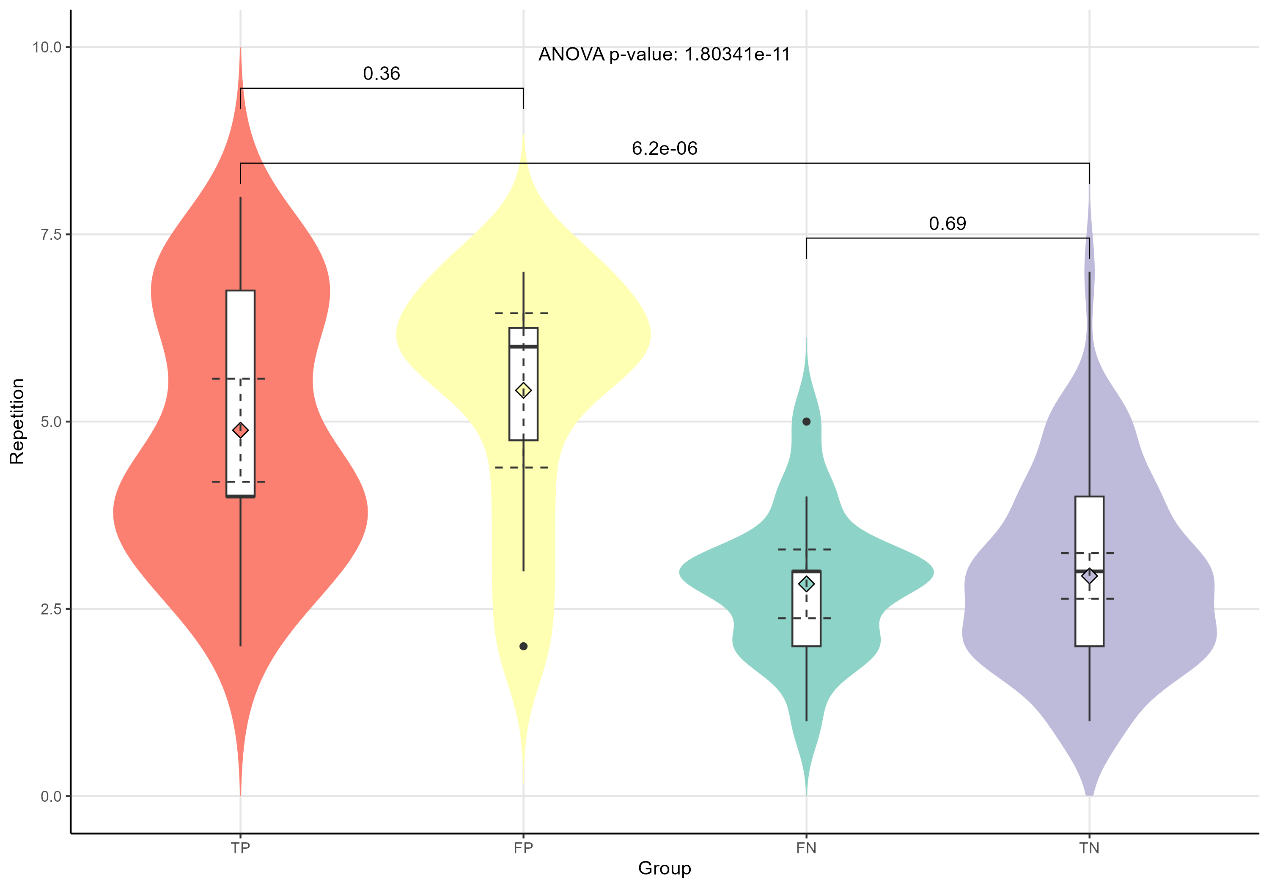

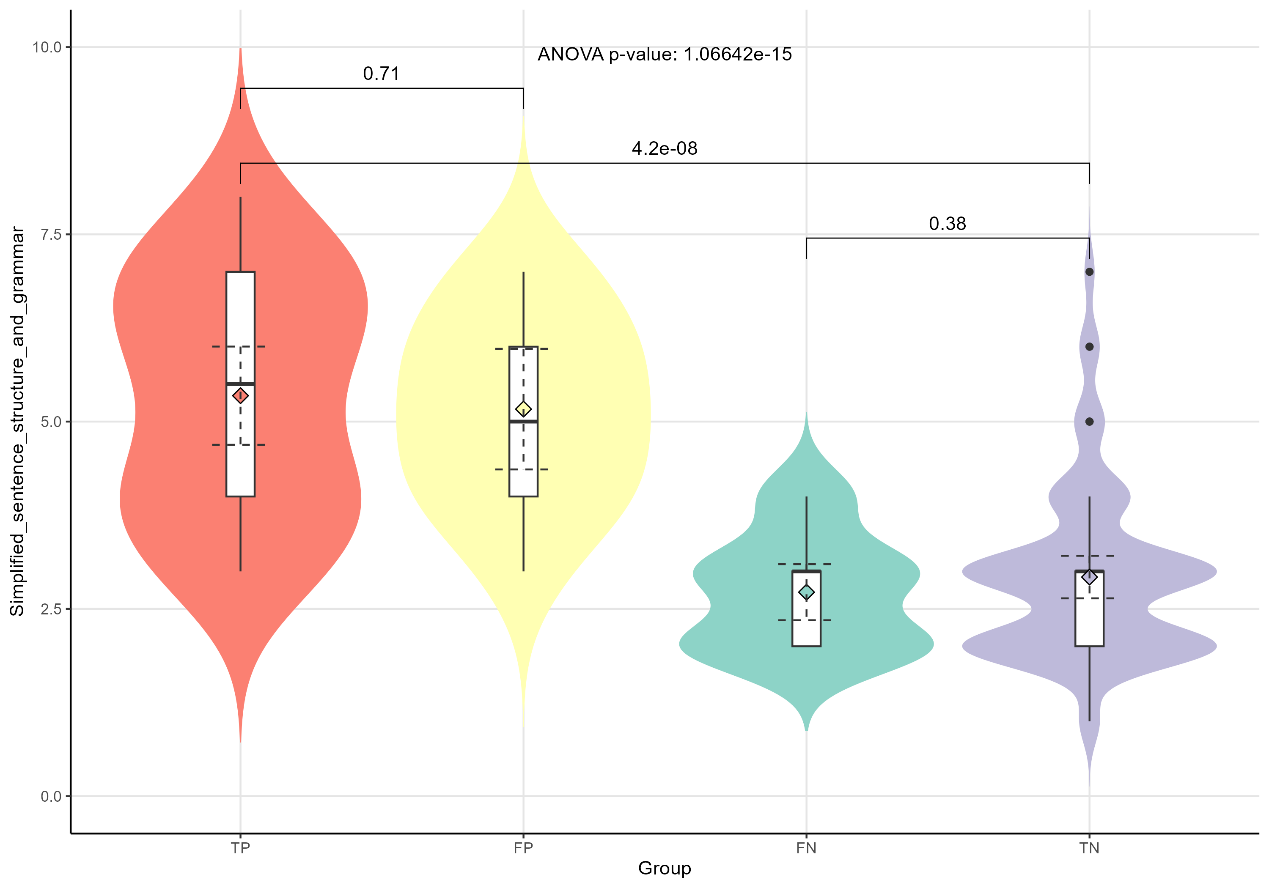

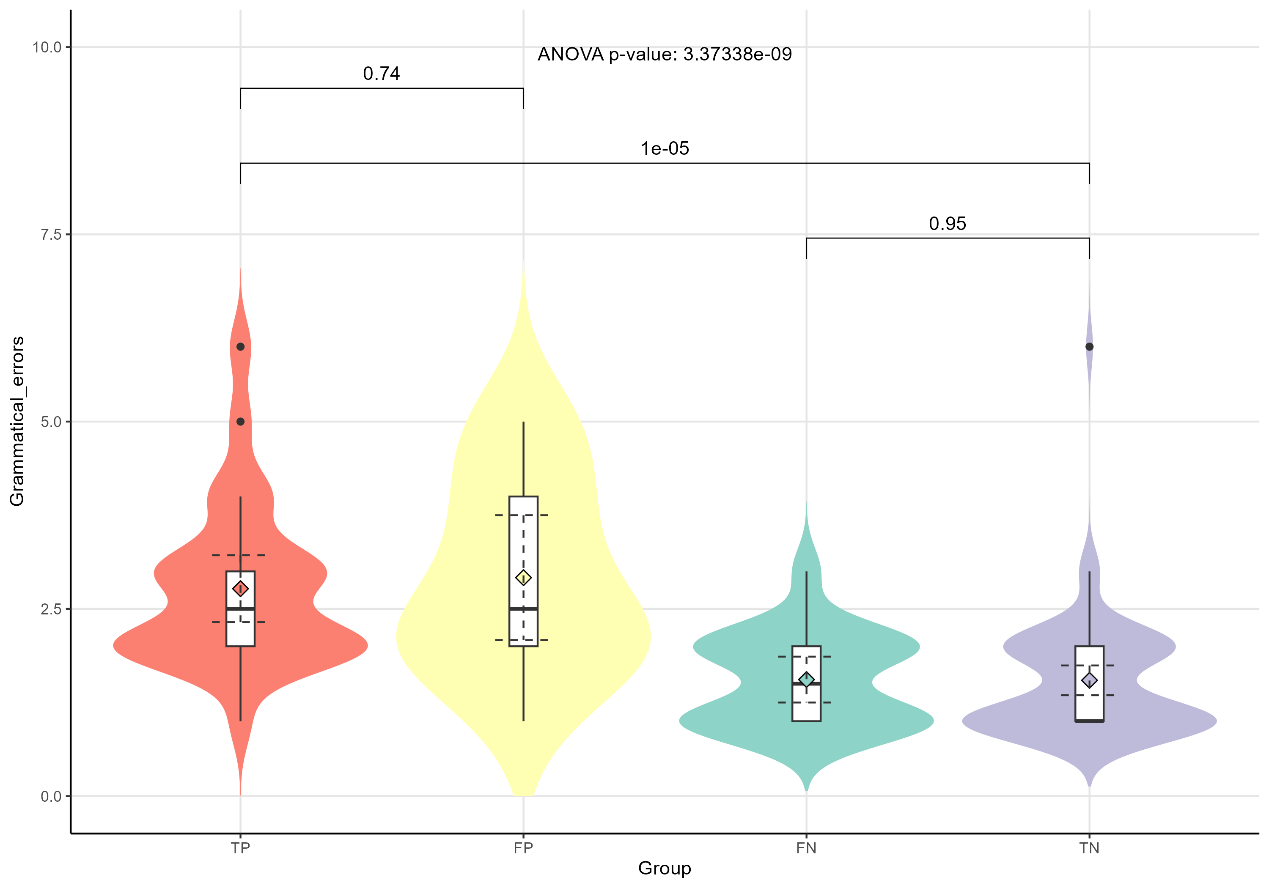

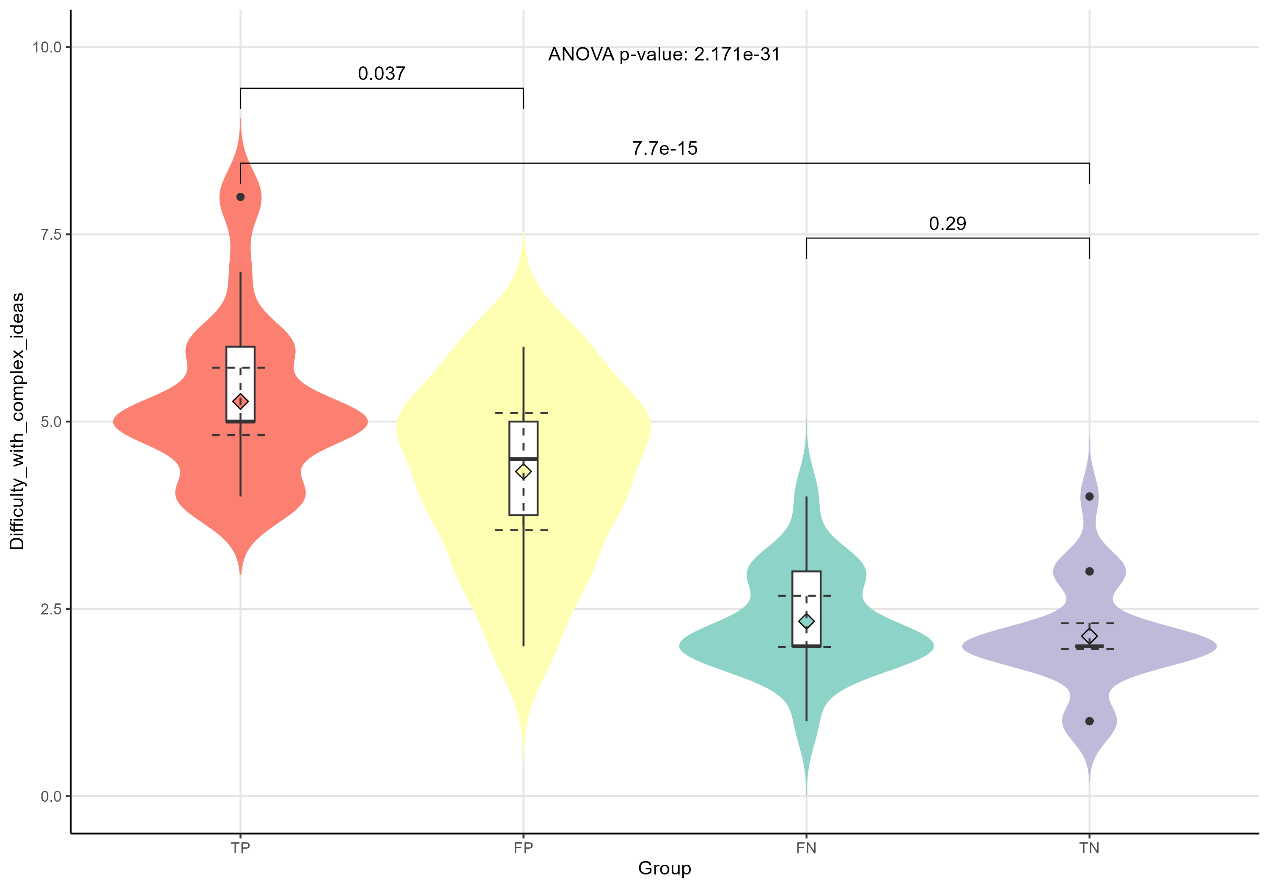

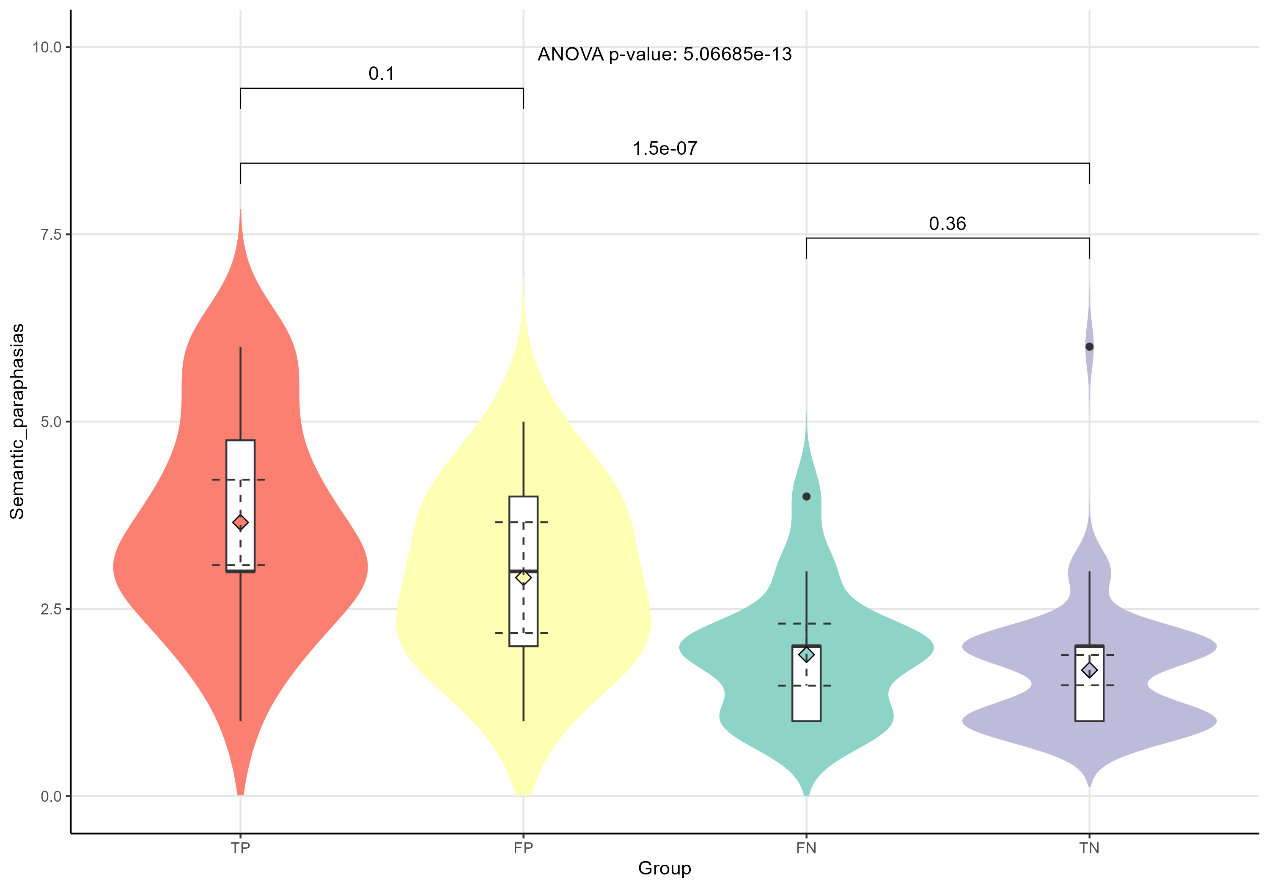

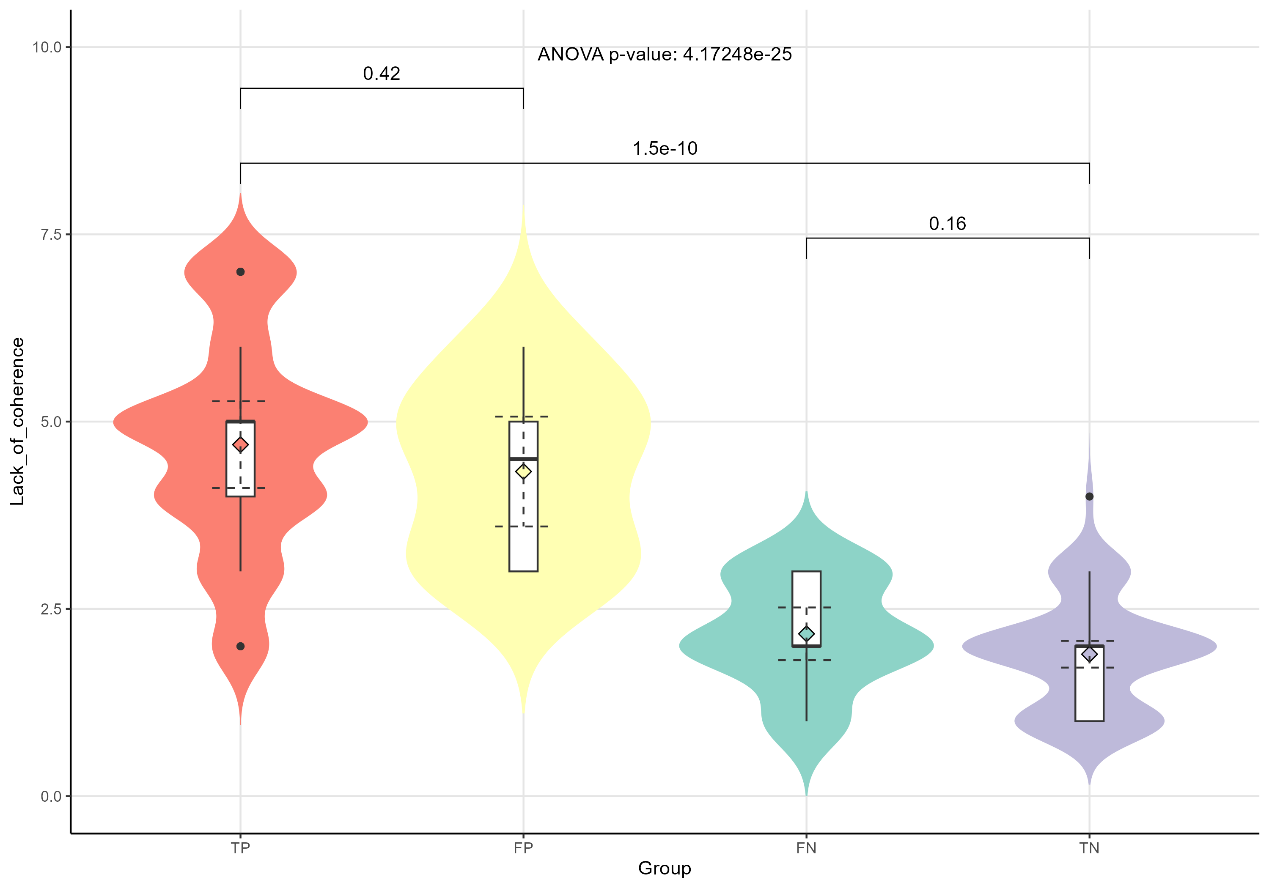

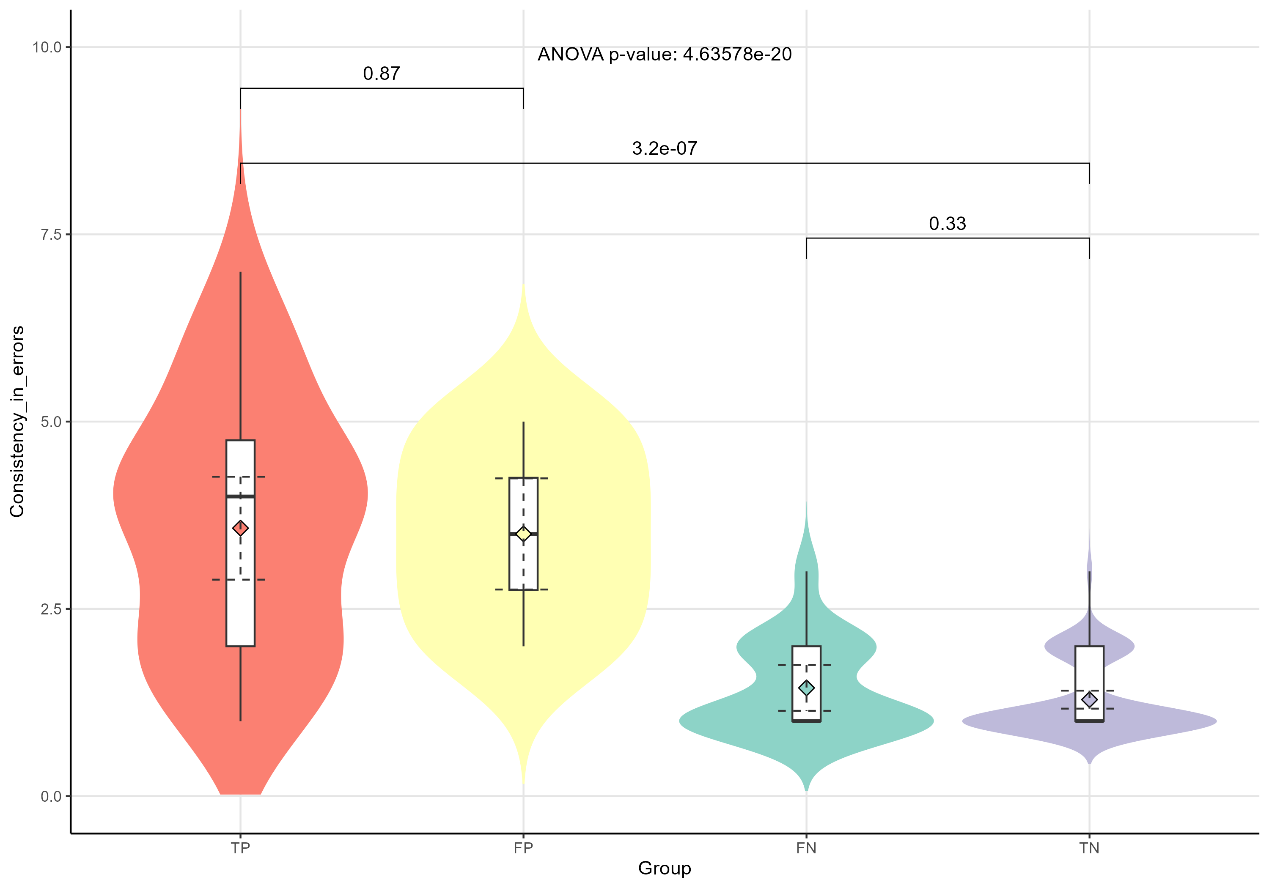

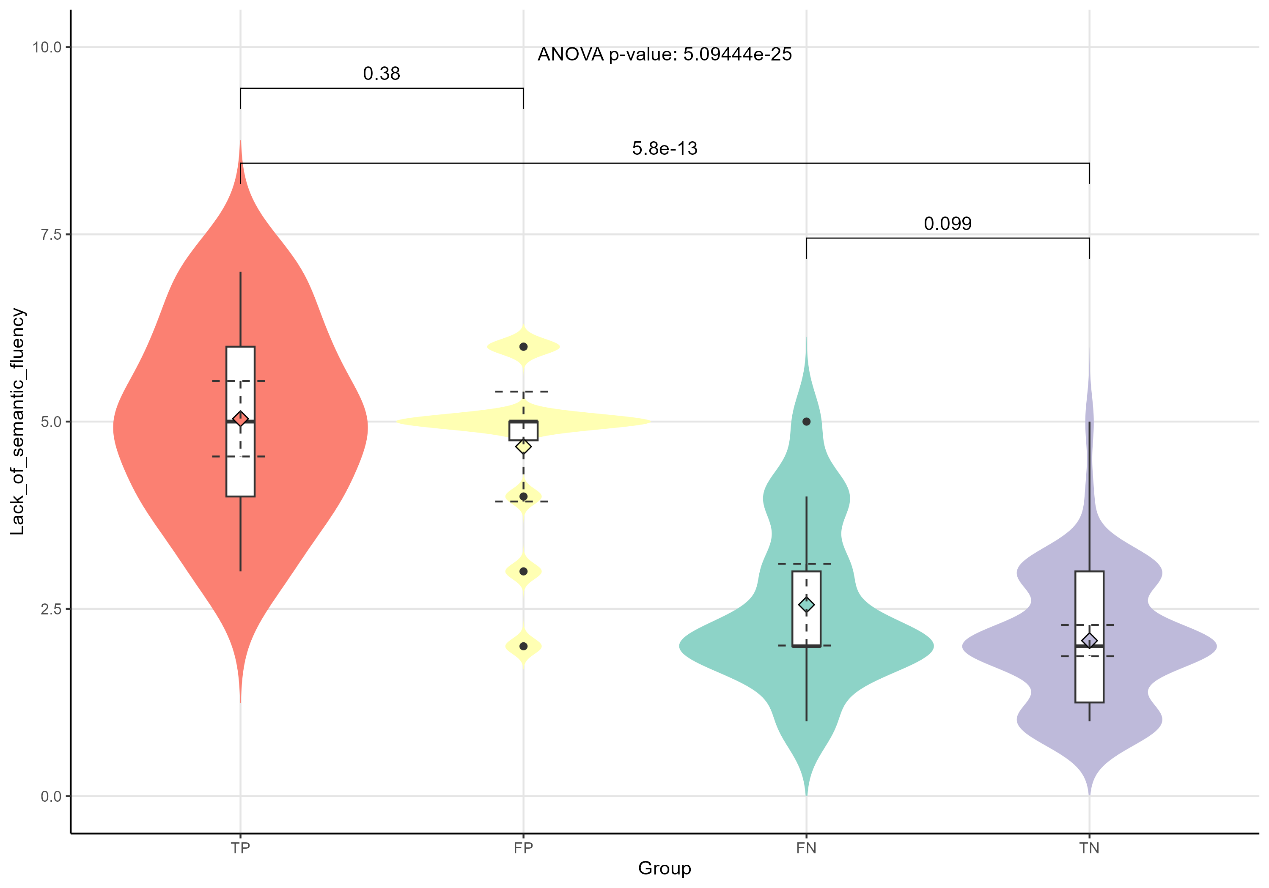

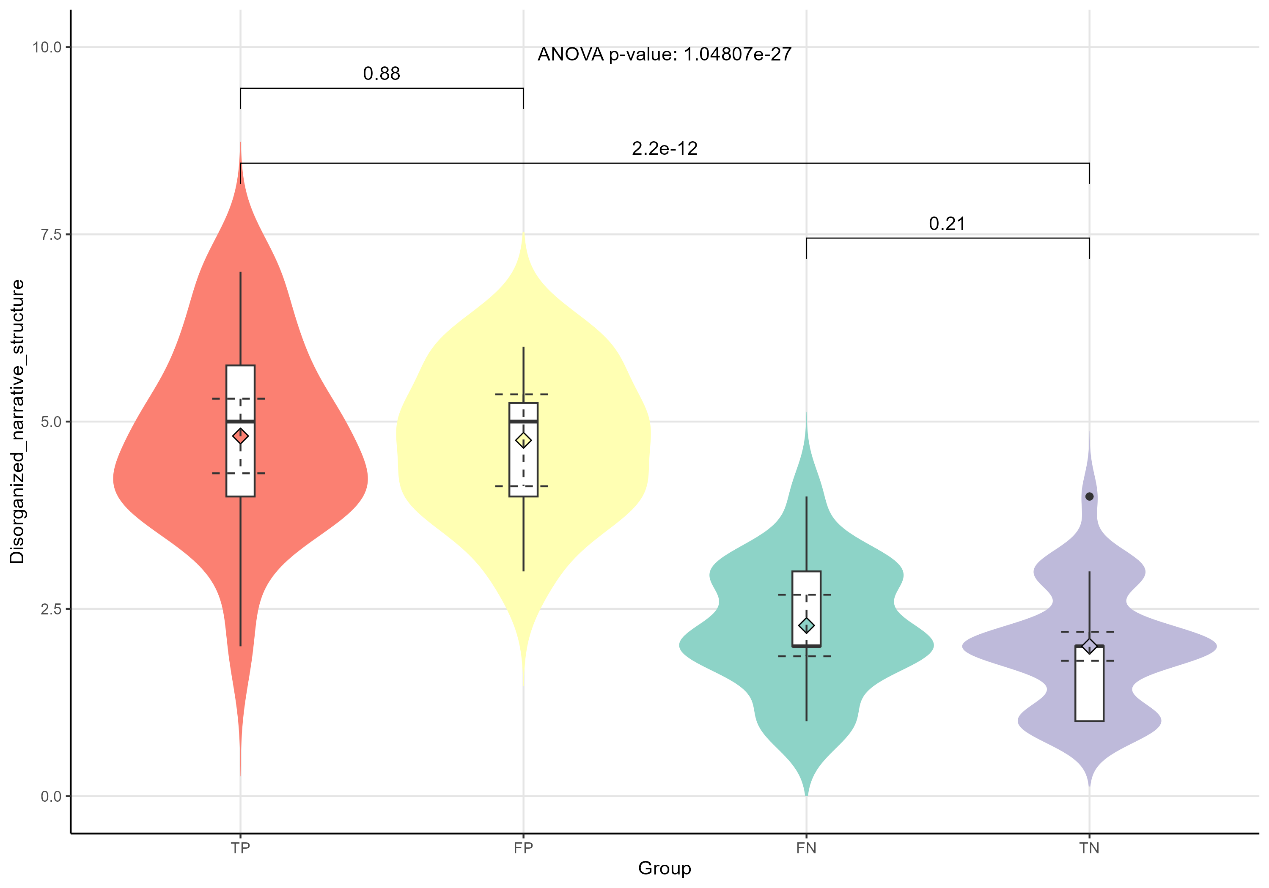

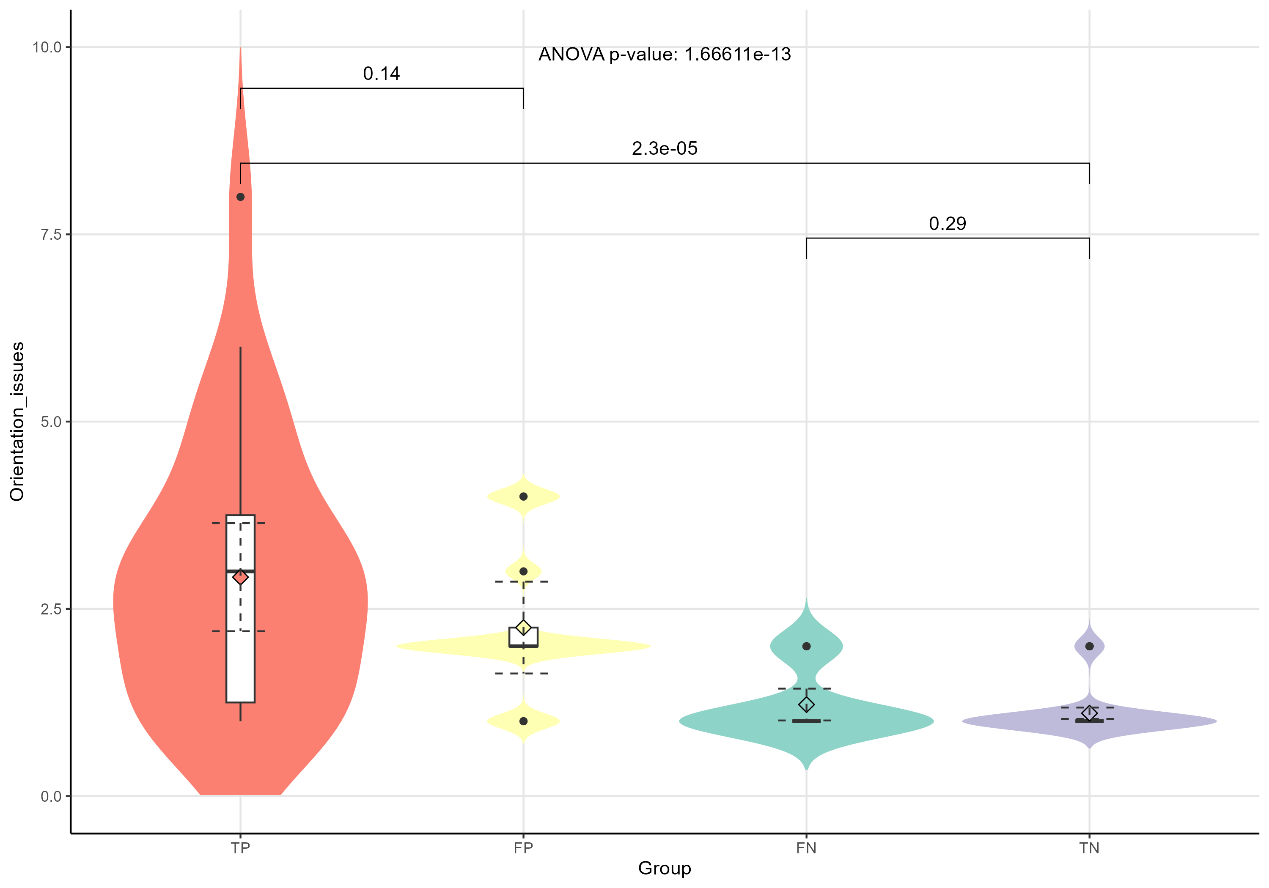

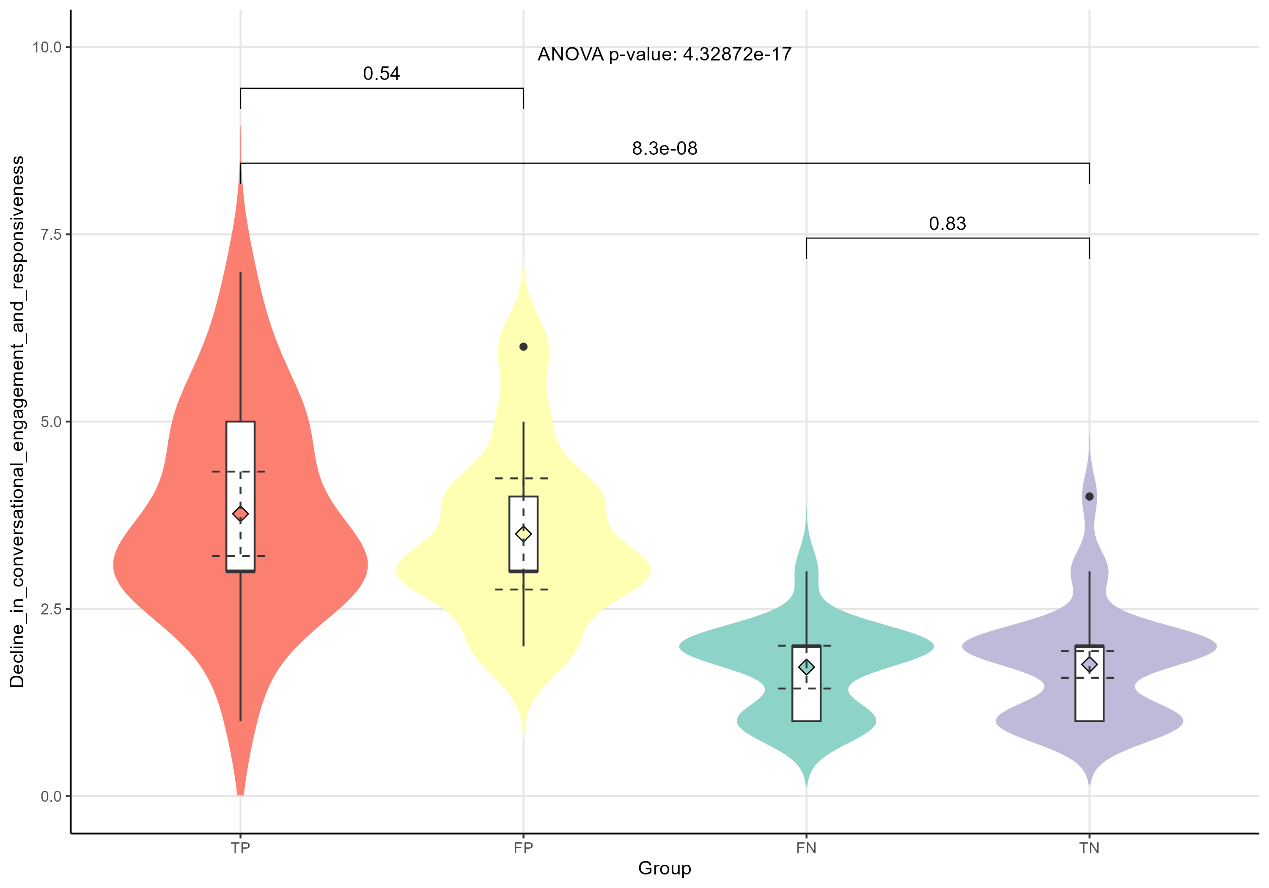

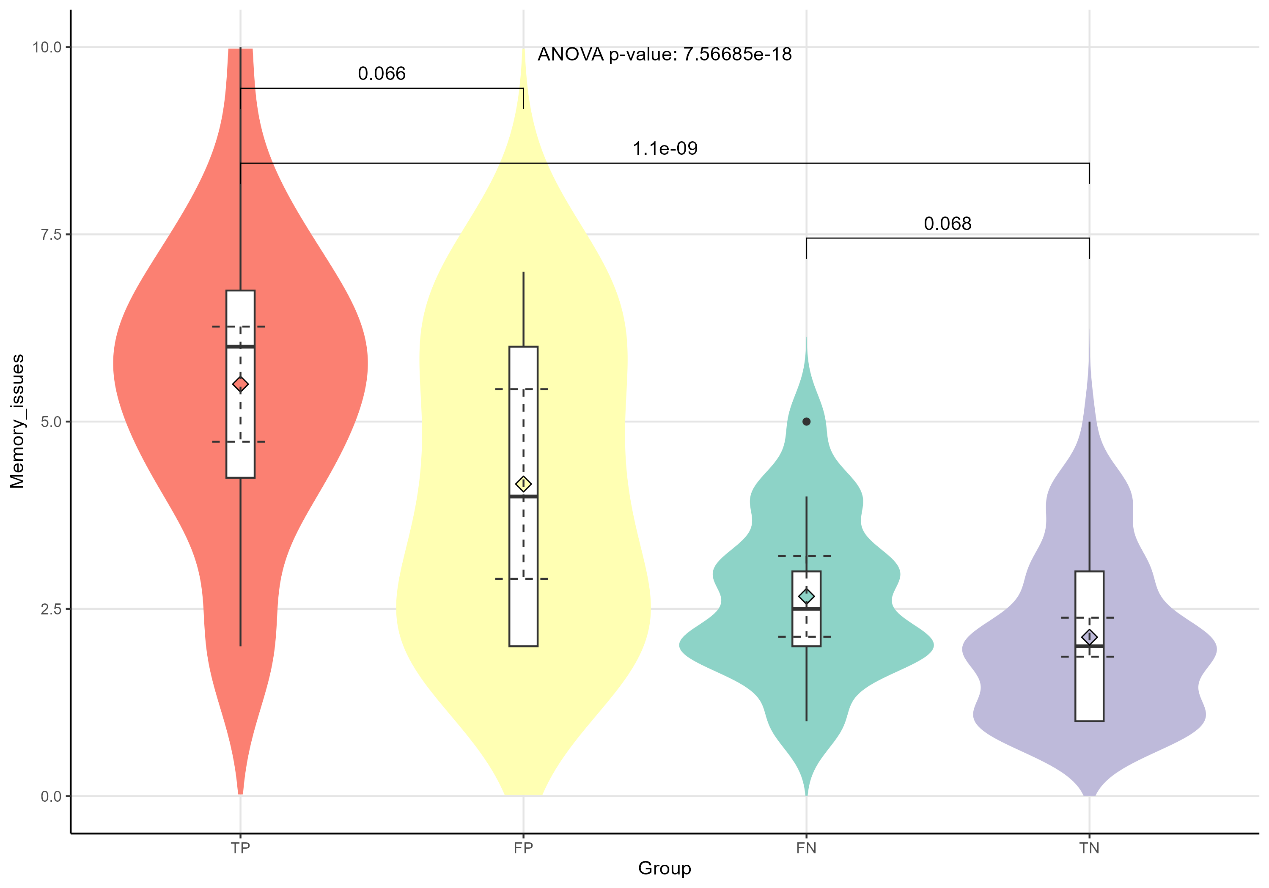

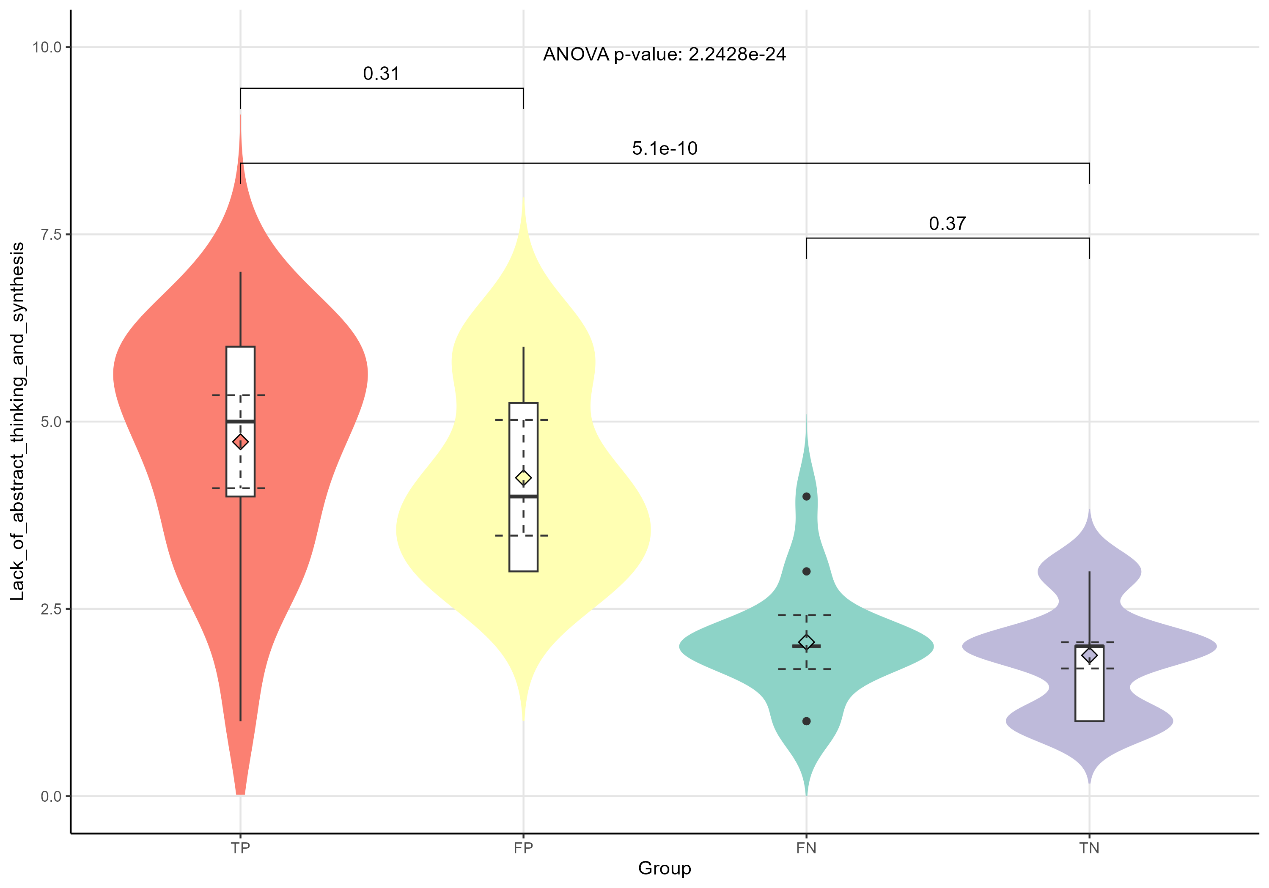

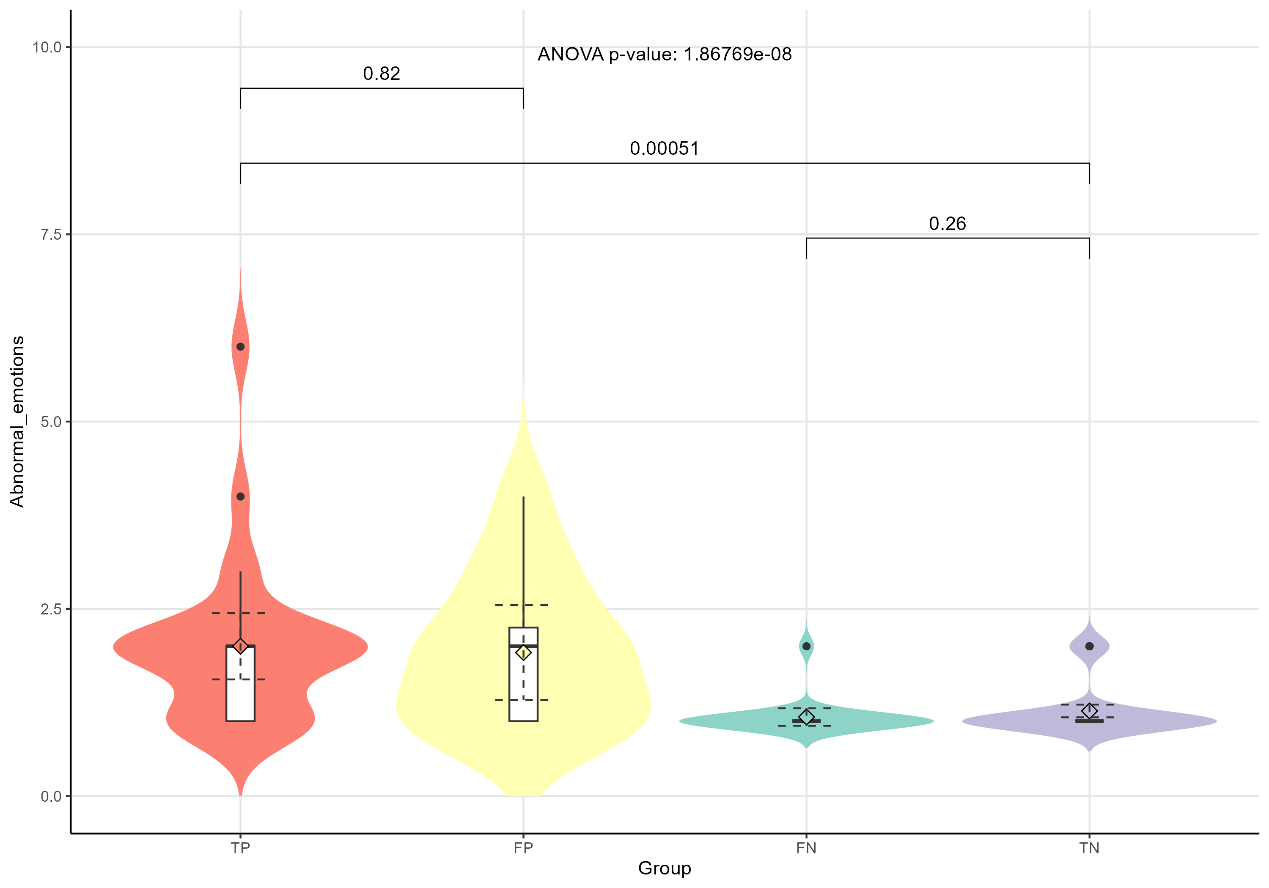
