## Supplementary material for "Text dialogue analysis Based ChatGPT for Primary Screening of Mild Cognitive Impairment": Data(GPT4_vs_GPT3.5_model1-5): 1_GPT3.5 (2).docx

Attention! You do not need to diagnose, just follow the steps below to initially screen for PARTICIPANT with possibly mild cognitive impairment (MCI). Some differences between MCI and normal cognition are also given, please analyse them in detail, step by step. Firstly, please assess the Vocabulary features of PARTICIPANT at the Word frequency and Word ratio, Phrase frequency and Phrase ratio, Word-finding difficulties, Repetition, Lexical complexity, Limited vocabulary. Word frequency: Count the occurrences of each unique word in the text. Unique words are distinct words that appear in the text, not taking into account their repetitions. Each word is counted only once, regardless of how many times it occurs. Word ratio: Calculate the proportion of unique words to the total number of words in the text. Phrase frequency: Count the occurrences of each unique phrases in the text. Unique phrases are distinct combinations of words that appear in the text, not taking into account their repetitions. Phrase ratio: Calculate the proportion of unique phrases to the total number of phrases in the text. Word-finding difficulties: Frequent pauses or use of filler words, like "uh" or "um,". MCI: more frequent instances, longer time to find the right word, or using more general terms. Normal: fewer instances, and better lexical retrieval. Repetition: The participant repeats certain phrases or ideas a few times throughout the conversation. MCI: more frequent repetition. Normal: less frequent repetition (some repetition of words but not excessive) and better memory of what has been mentioned. Lexical complexity: Measure the variety and sophistication of the vocabulary used, considering factors, including word length and frequency of rare words. Limited vocabulary: Repeated use of the same words or simple words. MCI: slightly reduced vocabulary, and more frequent use of simpler words or common terms; Normal: richer vocabulary (low limitation). Secondly, please assess the Semantics features of PARTICIPANT, from Semantic density and Inability to convey meaning, Semantic coherence and Lack of coherence. Semantic density: Measure the amount of meaningful information in the text, and considering factor is the number of content words (nouns, verbs, adjectives, adverbs) relative to function words (prepositions, pronouns, conjunctions, etc.). Inability to convey meaning: Difficulty in expressing ideas clearly, leading to confusion or misunderstanding. Normal: the participant was able to describe the scene clearly. Semantic coherence: Assess the logical consistency and meaningfulness of the text, which can be done by analyzing the relationships between words, phrases, and sentences, and checking for coherence and cohesion. Lack of coherence: Difficulty in maintaining a logical flow of ideas, with thoughts appearing disjointed or unrelated. MCI: struggle with organizing thoughts coherently or maintaining a clear focus in conversation; Normal: greater coherence and organization in their storytelling and conversations. Thirdly, please assess the Syntax and Grammar features of PARTICIPANT, from Syntactic complexity and Simplified sentence structure, Grammatical components and Grammatical errors. Syntactic complexity: Measure the complexity of the sentence structure in the text, considering factors such as the number of clauses per sentence, the average length of sentences, and the use of subordinate clauses. Simplified sentence structure: A lack of complex sentence structures. MCI: simpler sentence structures; Normal: in longer dialogues there is a greater variety of sentence structures (moderately complex), but in short dialogues the sentences may turn out to be simple. Grammatical components: Analyze the text for correct usage of grammar, such as subject-verb agreement, tense consistency, and proper use of pronouns, prepositions, and conjunctions. MCI: avoidance of complex grammatical constructions; Normal: better use of complex grammatical constructions. Grammatical errors: Frequent mistakes in verb tense, subject-verb agreement, or other aspects of grammar. MCI: more frequent grammatical errors, such as mistakes in verb tense and subject-verb agreement; Normal: fewer grammatical errors (a few errors, but not significant). Finally, please assign PARTICIPANT to the Probably Normal group or the Probably Mild Cognitive Impairment group, only. PARTICIPANT with MCI might exhibit more difficulty with vocabulary, sentence structure, repetition, word-finding, coherence, and grammar. However, it is not definitive and requires a comprehensive judgement. INVESTIGATOR: okay why don't you start with the woman again ? INVESTIGATOR: woman's wiping dishes . PARTICIPANT: the woman is wiping the dishes . PARTICIPANT: she's standing in a puddle of water . PARTICIPANT: if that's action I don't know . PARTICIPANT: the sink is overflowing . PARTICIPANT: doesn't seem to be anything going on outside that I can tell in INVESTIGATOR: okay good . PARTICIPANT: everything that I see going on . PARTICIPANT: okay . PARTICIPANT: well a little boy is stealing a cookie from the cookie jar . PARTICIPANT: he's also handing a cookie with his left hand to the girl . PARTICIPANT: he's also falling off a stool or the stool is tipping over . PARTICIPANT: the little girl is reaching for the cookie . PARTICIPANT: at the same time she's picking her nose or something . PARTICIPANT: I don't know what's she's doing . PARTICIPANT: …uh it looks like maybe the lid is falling from the cookie jar . PARTICIPANT: I'm not sure . PARTICIPANT: the lady is wiping a plate while the sink overflows . PARTICIPANT: and she's standing in a puddle of water . PARTICIPANT: am I going too fast ? INVESTIGATOR: no you take xxx . PARTICIPANT: and she's looking out the window . PARTICIPANT: I can't make out anything in particular going on outside . PARTICIPANT: the water … the faucet's turned on of_course which is causing PARTICIPANT: and the water is splashing on the floor . PARTICIPANT: that's really about all I can tell going on so to speak . INVESTIGATOR: okay . INVESTIGATOR: what's going on in the picture . PARTICIPANT: all of the things that I see going on ? INVESTIGATOR: right . PARTICIPANT: well the boy is stealing a cookie . PARTICIPANT: he's standing on the stool which is tipping over . PARTICIPANT: the girl is asking for cookies . PARTICIPANT: the lady is doing the dishes . PARTICIPANT: the sink is flowing over . PARTICIPANT: and as far as action goes I think that's about all . PARTICIPANT: she's standing in a puddle of water . PARTICIPANT: and the window is open . PARTICIPANT: and …uh grass and things are …uh outside . PARTICIPANT: a boy is handing the girl a cookie . PARTICIPANT: girl is picking her nose I guess . PARTICIPANT: I don't know what she's doing .

ChatGPT

Assessment of PARTICIPANT's language characteristics:

Vocabulary Features:

- Word frequency: The participant used a variety of words to describe the scene, including nouns (e.g., woman, dishes, sink), verbs (e.g., wiping, stealing, handing), and adjectives (e.g., overflowing, tipping). There is evidence of a reasonable word repertoire.
- Word ratio: The unique word ratio cannot be accurately determined without the full text, but the participant used a mixture of different words to describe the scene.
- Phrase frequency: The participant provided various phrases to describe the actions in the picture, such as "the woman is wiping the dishes" and "the boy is stealing a cookie from the cookie jar."
- Phrase ratio: The unique phrase ratio cannot be determined without the full text, but the participant used a range of phrases to convey information about the picture.
- Word-finding difficulties: The participant did not exhibit frequent pauses or use filler words like "uh" or "um," suggesting a relatively good ability to find the right words.
- Repetition: The participant repeated certain phrases or ideas a few times throughout the conversation, indicating some repetition but not excessive.
- Lexical complexity: The participant used both common and specific vocabulary related to the scene, indicating a reasonable level of lexical complexity.
- Limited vocabulary: The participant did not exhibit a significant repetition of words and demonstrated a variety of vocabulary.

Semantics Features:

- Semantic density: The participant provided sufficient content words (nouns, verbs, adjectives) relative to function words (prepositions, pronouns, conjunctions), indicating a reasonable semantic density.
- Inability to convey meaning: The participant expressed ideas clearly and effectively, with no apparent difficulty in conveying meaning.
- Semantic coherence: The participant's responses were logically consistent and meaningful, demonstrating coherence and cohesion in describing the scene.
- Lack of coherence: The participant maintained a logical flow of ideas throughout the conversation, without thoughts appearing disjointed or unrelated.

Syntax and Grammar Features:

- Syntactic complexity: The participant used a mixture of sentence structures, including simple and compound sentences, showing some syntactic complexity.
- Simplified sentence structure: The participant did not predominantly use short, simple sentences and displayed variation in sentence structures.
- Grammatical components: The participant demonstrated correct usage of grammar, including subject-verb agreement, tense consistency, and appropriate use of pronouns, prepositions, and conjunctions.
- Grammatical errors: There were no significant grammatical errors or frequent mistakes in verb tense, subject-verb agreement, or other aspects of grammar.

Based on the assessment, the participant's language characteristics align with the Probably Normal group. The participant exhibited a reasonable vocabulary range, coherent semantics, and syntactic complexity. However, it's important to note that a comprehensive evaluation by medical professionals is necessary to accurately determine cognitive status, as language assessment alone cannot provide a definitive diagnosis for mild cognitive impairment.
