## Supplementary material for "Text dialogue analysis Based ChatGPT for Primary Screening of Mild Cognitive Impairment": Data(GPT4_vs_GPT3.5_model1-5): 1_GPT3.5 (3).docx

ChatGPT

Analysis and evaluation indicators for PARTICIPANT:

Vocabulary Features:

- Word frequency and Word ratio: Without the full text, it is not possible to calculate the word frequency and word ratio accurately.
- Phrase frequency and Phrase ratio: Without the full text, it is not possible to calculate the phrase frequency and phrase ratio accurately.
- Lexical complexity: Based on the given dialogue, the participant demonstrated a reasonable variety of vocabulary, including nouns, verbs, and adjectives, indicating moderate lexical complexity.

Evaluation indicators:

1. Limited vocabulary: No evidence of a significantly limited vocabulary. Severity: 1
2. Increased verbosity: No evidence of excessive verbosity. Severity: 1
3. Overlearned phrases: No evidence of overuse of cliches or stock phrases. Severity: 1
4. Improper use of pronouns: No evidence of significant difficulty with pronouns. Severity: 1
5. Hesitation (Word-finding difficulties): No significant pauses or filler words observed. Severity: 1
6. Circumlocution (Word-finding difficulties): No evidence of deliberate vagueness or evasiveness in language use. Severity: 1
7. Tip-of-the-tongue phenomenon: No evidence of partial recall or difficulty in recalling specific words. Severity: 1
8. Repetition: Some repetition of words and phrases, but not excessive. Severity: 2

Syntax and Grammar Features:

- Syntactic complexity: Without the full text, it is not possible to accurately assess the syntactic complexity.
- Grammatical components: No significant grammatical errors observed.

Evaluation indicators:

1. Simplified sentence structure and grammar: No evidence of predominantly simplified sentence structures or avoidance of complex grammatical constructions. Severity: 1
2. Grammatical errors: No significant grammatical errors observed. Severity: 1
3. Difficulty with complex ideas: No evidence of difficulty in handling complex sentences or ideas. Severity: 1

Semantics Features:

- Semantic density: Without the full text, it is not possible to accurately assess the semantic density.
- Semantic coherence: The participant's responses were logically consistent and meaningful.

Evaluation indicators:

1. Semantic paraphasias: No evidence of errors in word choice or odd combinations of words. Severity: 1
2. Lack of coherence: No evidence of significant struggle with maintaining coherence or changing topics abruptly. Severity: 1
3. Consistency in errors: Without the full text, it is not possible to assess consistency in errors.
4. Lack of semantic fluency: No evidence of difficulty in naming items or describing processes. Severity: 1
5. Disorganized narrative structure: No evidence of a disorganized narrative structure. Severity: 1
6. Orientation issues: No evidence of confusion about time, place, or sequence of events. Severity: 1
7. Decline in conversational engagement and responsiveness: No evidence of decreased engagement, difficulties in following a conversation, or steering conversations back to familiar topics. Severity: 1
8. Memory issues: No evidence of significant difficulty in memory recall. Severity: 1
9. Lack of abstract thinking and synthesis: No evidence of difficulty in understanding or expressing abstract concepts. Severity: 1
10. Abnormal emotions: No evidence of changes in tone, lack of initiative, or abnormal emotional responses. Severity: 1

Based on the available information, PARTICIPANT's performance aligns more with the Probably Normal Cognition Group. The participant demonstrated a reasonable vocabulary range, coherence in semantics and syntax, and an overall ability to convey information clearly. However, it is important to note that a comprehensive evaluation by medical professionals is necessary to accurately determine cognitive status, as language assessment alone cannot provide a definitive diagnosis
