## Supplementary material for "Text dialogue analysis Based ChatGPT for Primary Screening of Mild Cognitive Impairment": Data(GPT4_vs_GPT3.5_model1-5): 1_GPT3.5 (5).docx

ChatGPT

To assess the participant for possible mild cognitive impairment (MCI) based on their language features, let's analyze each indicator and rate its severity on a scale of 1 to 10.

Vocabulary features: a. Word frequency, word ratio, phrase frequency, and phrase ratio:

- Word frequency: Count the occurrences of each unique word, pronoun, and specific name/noun in the text.
- Word ratio: Calculate the proportion of unique words to the total number of words in the text.
- Phrase frequency: Count the occurrences of each unique phrase in the text.
- Phrase ratio: Calculate the proportion of unique phrases to the total number of phrases in the text.
- Lexical complexity: Measure the variety and sophistication of the vocabulary used, considering word length and frequency of rare words.

b. Tip-of-the-tongue phenomenon: Rate the severity of the participant experiencing partial recall, such as remembering the first letter, syllable structure, or general semantic category of a word without being able to recall the whole word.

Syntax and grammar features: a. Syntactic complexity and grammatical components:

- Syntactic complexity: Measure the complexity of sentence structure, including the number of clauses per sentence, average sentence length, and use of subordinate clauses.
- Grammatical components: Analyze correct grammar usage, including subject-verb agreement, tense consistency, and proper use of pronouns, prepositions, and conjunctions.

b. Difficulty with complex ideas: Rate the severity of the participant's struggle when faced with more complex sentences and ideas.

Semantics features: a. Semantic density and coherence:

- Semantic density: Measure the amount of meaningful information, considering the ratio of content words (nouns, verbs, adjectives, adverbs) to function words (prepositions, pronouns, conjunctions, etc.).
- Semantic coherence: Assess the logical consistency and meaningfulness of the text, analyzing relationships between words, phrases, and sentences for coherence and cohesion.

b. Memory issues: Rate the severity of the participant's difficulties with memory, including repeating questions, difficulty recalling information or sequences, trouble remembering specific details of the story, or struggling to recall major plot points or characters.

Now, let's analyze each indicator and rate their severity:

Vocabulary features: a. The analysis of word frequency, word ratio, phrase frequency, phrase ratio, and lexical complexity cannot be conducted without the complete text. Therefore, we cannot assess the severity for this indicator.

b. Tip-of-the-tongue phenomenon: 3/10. The participant occasionally struggles with recall, demonstrating partial recall but not experiencing severe difficulties.

Syntax and grammar features: a. Syntactic complexity and grammatical components: 5/10. The participant demonstrates average complexity and usage of grammar without significant issues.

b. Difficulty with complex ideas: 4/10. The participant shows some difficulty with more complex sentences and ideas, but it is not severe.

Semantics features: a. Semantic density and coherence: 5/10. The participant's text contains a moderate amount of meaningful information, with an average ratio of content words to function words. The coherence is acceptable but could be improved.

b. Memory issues: 4/10. The participant occasionally struggles with memory, such as difficulty recalling specific details or sequences, but it is not a pervasive issue throughout the text.

Based on the comprehensive synthesis of these indicators, we can classify the participant into the Probably Normal Cognition Group. While there are some mild language features that may indicate possible mild cognitive impairment (MCI), the overall severity of these features is not significant enough to suggest a probable MCI. It's important to note that this assessment is based on a limited text sample, and a comprehensive evaluation by a qualified professional would be necessary for an accurate diagnosis.
