## Supplementary material for "Text dialogue analysis Based ChatGPT for Primary Screening of Mild Cognitive Impairment": Data(GPT4_vs_GPT3.5_model1-5): 1_GPT3.5.docx

ChatGPT

Assessment of Vocabulary features:

- Word frequency: The participant uses a limited range of words and repeats some words, such as "cookie," "water," and "puddle," several times.
- Word ratio: The participant uses 73 unique words out of a total of 105 words, which gives a word ratio of 0.695 or 69.5%.
- Phrase frequency: The participant uses some simple and repetitive phrases, such as "the boy is," "the girl is," "the lady is," and "the sink is," among others.
- Phrase ratio: The participant uses 33 unique phrases out of a total of 43 phrases, which gives a phrase ratio of 0.767 or 76.7%.
- Difficulty finding the right words: The participant does not seem to have major difficulties in finding the right words, but there are some pauses and the use of filler words, such as "uh," "um," and "I guess."
- Lexical complexity: The participant's vocabulary is mostly simple and lacks complexity or sophistication.
- Limited vocabulary: The participant uses simple and basic words and does not show a wide range of vocabulary.

Assessment of Semantics features:

- Semantic density: The participant uses mostly content words, such as nouns, verbs, adjectives, and adverbs, which gives a high semantic density.
- Inability to convey meaning: The participant is able to convey the basic meaning of the scene, but there are some instances of unclear or ambiguous descriptions, such as "she's picking her nose or something" and "I don't know what she's doing."
- Semantic coherence: The participant's descriptions are mostly logically consistent and meaningful, but there are some instances of disjointed or unrelated thoughts, such as "I don't know what she's doing" and "the window is open and grass and things are outside."
- Lack of coherence: The participant's descriptions lack a clear structure and organization, with thoughts appearing somewhat disjointed or unrelated.

Assessment of Syntax and Grammar features:

- Syntactic complexity: The participant uses predominantly short and simple sentences with a lack of complex structures, such as subordinate clauses.
- Simplified sentence structure: The participant uses simple sentence structures, with few complex structures or longer sentences.
- Grammatical components: The participant shows correct usage of grammar, such as subject-verb agreement and proper use of pronouns, prepositions, and conjunctions, with no major errors.
- Grammatical errors: The participant shows no major grammatical errors.

Based on these assessments, the participant shows some language deficits, particularly in vocabulary and coherence, but overall their language use is relatively normal. Therefore, I would assign the participant to the Probably Normal group. However, I would recommend further assessment and monitoring over time, particularly if there are concerns about cognitive decline or memory loss.
