## Supplementary material for "Text dialogue analysis Based ChatGPT for Primary Screening of Mild Cognitive Impairment": Data(GPT4_vs_GPT3.5_model1-5): 1_GPT4 (2).docx

ChatGPT

Analyzing the participant's responses based on the criteria, here are the findings:

1. **Vocabulary Features**:
   - *Word frequency and Word ratio*: The participant uses a wide range of words with some repetition. They use the words "cookie", "girl", "boy", "water", and "window" frequently. The ratio of unique words to the total number of words used is moderately high.
   - *Phrase frequency and Phrase ratio*: The participant uses a variety of phrases and sentences to describe the scene, and there is not much repetition.
   - *Word-finding difficulties*: There are no significant word-finding difficulties exhibited by the participant.
   - *Repetition*: The participant does not repeat phrases excessively.
   - *Lexical complexity*: The participant uses a variety of words, including more complex terms such as "overflowing", "picking her nose", "stealing".
   - *Limited vocabulary*: The participant doesn't seem to have a limited vocabulary as they use a range of different words and phrases.
2. **Semantics Features**:
   - *Semantic density*: The participant's responses contain a good mix of content words and function words, indicating a high semantic density.
   - *Inability to convey meaning*: The participant seems capable of conveying meaning, as their descriptions of the scene are clear and detailed.
   - *Semantic coherence*: The participant's responses seem logically consistent and meaningful, demonstrating good semantic coherence.
   - *Lack of coherence*: The participant does not struggle with maintaining a logical flow of ideas.
3. **Syntax and Grammar Features**:
   - *Syntactic complexity*: The participant uses a variety of sentence structures, indicating high syntactic complexity.
   - *Simplified sentence structure*: The participant does not consistently use simplified sentence structures.
   - *Grammatical components*: The participant seems to have a good grasp of grammar, as their responses generally follow grammatical rules.
   - *Grammatical errors*: There are few, if any, grammatical errors in the participant's responses.

Based on this analysis, I would place the participant in the "Normal" group. The participant demonstrates a good command of vocabulary, semantic coherence, and grammar, and there are no significant indications of potential cognitive impairment based on the provided responses. However, for a definitive conclusion, further comprehensive assessments would be needed
