## Supplementary material for "Text dialogue analysis Based ChatGPT for Primary Screening of Mild Cognitive Impairment": Data(GPT4_vs_GPT3.5_model1-5): 1_GPT4 (3).docx

ChatGPT

Based on the given text, here's the analysis:

**Vocabulary Features**

1. Limited vocabulary: 3/10. The PARTICIPANT has decent vocabulary, but tends to repeat certain phrases, such as "the girl is" and "the lady is."
2. Increased verbosity: 2/10. The PARTICIPANT mostly stays to the point, though occasionally they include additional, unnecessary details.
3. Overlearned phrases: 1/10. The PARTICIPANT doesn't seem to rely on cliches or stock phrases.
4. Improper use of pronouns: 2/10. The PARTICIPANT uses pronouns appropriately.
5. Hesitation (Word-finding difficulties): 3/10. Occasional hesitation is observed, but it is not frequent.
6. Circumlocution (Word-finding difficulties): 2/10. There's little evidence of circumlocution; the PARTICIPANT generally finds the right words.
7. Tip-of-the-tongue phenomenon: 1/10. There isn't any evidence of this phenomenon in the text.
8. Repetition: 3/10. There is some repetition in the PARTICIPANT's description.

**Syntax and Grammar Features**

1. Simplified sentence structure and grammar: 2/10. The PARTICIPANT uses a mix of simple and complex sentences.
2. Grammatical errors: 1/10. The PARTICIPANT's grammar is mostly correct.
3. Difficulty with complex ideas: 2/10. The PARTICIPANT seems to understand and express complex ideas without much difficulty.

**Semantics Features**

1. Semantic paraphasias: 2/10. The PARTICIPANT generally expresses ideas clearly.
2. Lack of coherence: 2/10. The narrative is coherent, though the PARTICIPANT occasionally loses focus.
3. Consistency in errors: 1/10. The PARTICIPANT makes few errors.
4. Lack of semantic fluency: 2/10. The PARTICIPANT seems to have no trouble naming items or describing a sequence of events.
5. Disorganized narrative structure: 2/10. The PARTICIPANT's narrative occasionally jumps around but is mostly organized.
6. Orientation Issues: 1/10. The PARTICIPANT seems to understand the sequence of events.
7. Decline in conversational engagement and responsiveness: 2/10. The PARTICIPANT responds to the investigator's prompts and maintains the conversation.
8. Memory issues: 1/10. The PARTICIPANT seems to remember details of the scene.
9. Lack of abstract thinking and synthesis: 2/10. The PARTICIPANT synthesizes the overall situation effectively.
10. Abnormal emotions: 1/10. There is no evidence of abnormal emotions.

Based on this analysis, the PARTICIPANT would likely fall into the **Probably Normal Cognition Group**. Despite occasional repetition and slight hesitation, the PARTICIPANT demonstrates a good understanding of the scene, expresses ideas clearly, and maintains a coherent narrative. There are no serious issues with vocabulary, syntax, grammar, or semantics, and there is no evidence of memory problems or abnormal emotions. However, please note that this is an initial screening, and further assessment may be necessary to confirm these findings
