## Supplementary material for "Text dialogue analysis Based ChatGPT for Primary Screening of Mild Cognitive Impairment": Data(GPT4_vs_GPT3.5_model1-5): 1_GPT4 (4).docx

ChatGPT

Based on the text presented, I will assess the participant's language features:

Vocabulary features:

1. Word and phrase frequency: 6/10 - The participant frequently uses phrases like "the sink is overflowing", "she's standing in a puddle of water", showing some repetition.
2. Lexical complexity: 5/10 - The participant uses a variety of words and phrases, but there's a tendency to stick to simple and direct vocabulary.
3. Limited vocabulary: 4/10 - The participant uses a narrow range of vocabulary, often repeating similar phrases.
4. Increased verbosity: 3/10 - The participant is mostly concise in their language use.
5. Overlearned phrases: 2/10 - The participant doesn't appear to rely heavily on overlearned phrases.
6. Improper use of pronouns: 1/10 - The participant uses pronouns appropriately.
7. Hesitation: 5/10 - The participant uses "uh" frequently, indicating hesitancy.
8. Circumlocution: 1/10 - The participant doesn't seem to talk around the point, remaining direct and specific.
9. Tip-of-the-tongue phenomenon: 2/10 - The participant doesn't seem to struggle to recall specific words.
10. Repetition: 6/10 - Some significant repetition is observed in the participant's narration.

Syntax and grammar features:

1. Syntactic complexity: 4/10 - The participant uses mostly simple sentences, with a few complex ones.
2. Simplified sentence structure and grammar: 2/10 - The participant doesn't tend to oversimplify sentences.
3. Grammatical errors: 1/10 - No obvious grammatical errors are noticed.
4. Difficulty with complex ideas: 3/10 - The participant doesn't seem to struggle with complex ideas.

Semantics features:

1. Semantic density and coherence: 6/10 - The participant's statements are generally coherent, but some repetition reduces semantic density.
2. Semantic paraphasias: 1/10 - The participant doesn't seem to use wrong words.
3. Lack of coherence: 2/10 - Most of the participant's statements are coherent.
4. Consistency in errors: 3/10 - Some errors are consistent, such as repetition and frequent usage of "uh".
5. Lack of semantic fluency: 3/10 - The participant seems somewhat semantically fluent but with a narrow range of vocabulary.
6. Disorganized narrative structure: 2/10 - The participant's narratives seem structured.
7. Orientation issues: 2/10 - The participant demonstrates a clear understanding of the visual scene.
8. Decline in conversational engagement and responsiveness: 2/10 - The participant responds to all prompts adequately.
9. Memory issues: 2/10 - No immediate memory issues are detected in this interaction.
10. Lack of abstract thinking and synthesis: 2/10 - The participant seems capable of abstract thinking.
11. Abnormal emotions: 1/10 - No abnormal emotions are detected in the given text.

Based on these assessments, the participant seems to be in the Probably Normal Cognition Group. Some indicators, such as repetition and frequent use of "uh", may suggest a need for further observation. However, these issues might be part of the participant's normal speech patterns. These assessments should be validated with further testing and expert evaluations.
