## Supplementary material for "Text dialogue analysis Based ChatGPT for Primary Screening of Mild Cognitive Impairment": Data(GPT4_vs_GPT3.5_model1-5): 1_GPT4 (5).docx

ChatGPT

Vocabulary Features: a. The participant used a variety of unique words, pronouns, and specific nouns. The lexical complexity appears to be moderate, as the participant doesn't use many sophisticated words or phrases but maintains a certain variety. The word frequency and ratio, as well as the phrase frequency and ratio, show a relatively balanced use of language, with no excessive repetition of particular words or phrases. Severity: 3/10

b. The tip-of-the-tongue phenomenon doesn't seem very pronounced in this transcript. There are occasional moments where the participant seems unsure (e.g., "I don't know what she's doing"), but it's not very frequent or severe. Severity: 2/10

Syntax and Grammar Features: a. The participant's sentences tend to be of moderate complexity, with a good balance of shorter and longer sentences. The grammatical components are generally used correctly, including appropriate use of verbs, tense consistency, pronouns, prepositions, and conjunctions. Severity: 2/10

b. While the participant doesn't express many complex ideas in the text, they do seem to have some difficulty when describing more complex scenes or actions (e.g., "I can't make out anything in particular going on outside"). This suggests a possible issue with handling complex ideas, though it's not severe in this case. Severity: 4/10

Semantic Features: a. The participant's semantic density appears to be relatively high, with many content words used and a fair amount of meaningful information conveyed. The semantic coherence is also good, with the descriptions generally consistent and meaningful, and the relationships between words, phrases, and sentences mostly clear. Severity: 2/10

b. The participant doesn't show strong signs of memory issues in the provided text. They repeat some descriptions ("she's standing in a puddle of water"), but it could be due to the nature of the task. They seem to remember the general sequence of their narrative well. Severity: 3/10

Based on this analysis, the participant would be classified in the "Probably Normal Cognition Group". While there are some minor signs of possible cognitive impairment, they aren't pronounced enough to suggest a likelihood of mild cognitive impairment. Remember, this evaluation is a screening tool and does not replace a formal diagnosis from a healthcare professional.
