## Supplementary material for "Text dialogue analysis Based ChatGPT for Primary Screening of Mild Cognitive Impairment": Data(GPT4_vs_GPT3.5_model1-5): 1_GPT4.docx

ChatGPT

Assessment of PARTICIPANT:

Vocabulary features: Word frequency: 115 unique words Word ratio: 0.348 (115 unique words / 330 total words) Phrase frequency: 96 unique phrases Phrase ratio: 0.291 (96 unique phrases / 330 total words) Difficulty finding the right words: Mild, with some occurrences of "uh" and "I don't know" Lexical complexity: Moderate, with a mix of simple and more sophisticated words Limited vocabulary: Mildly limited, with some repetition of words and phrases

Semantics features: Semantic density: Moderate, with a balance of content and function words Inability to convey meaning: Mild, with some confusion regarding the girl's action Semantic coherence: Moderate, with mostly logical consistency and meaningfulness Lack of coherence: Mild, with some disjointed thoughts and repetition

Syntax and Grammar features: Syntactic complexity: Moderate, with a mix of simple and complex sentence structures Simplified sentence structure: Mild, with some simple sentences and repetitive phrasing Grammatical components: Generally correct, with occasional errors Grammatical errors: Mild, with a few tense inconsistencies and other minor errors

Based on this assessment, I would assign PARTICIPANT to the Probably Normal group. The participant demonstrates some mild difficulties and inconsistencies in language use, but overall, they are able to communicate effectively and demonstrate a moderate level of vocabulary, syntax, and semantic coherence.
