## Supplementary material for "Text dialogue analysis Based ChatGPT for Primary Screening of Mild Cognitive Impairment": Data(GPT4_vs_GPT3.5_model1-5): 10_GPT3.5 (3).docx

ChatGPT

Based on the provided conversation, here is an analysis of the indicators and evaluation for the participant:

Vocabulary Features:

- Word frequency and Word ratio: The participant uses a moderate number of unique words, with some repetition. The ratio of pronouns to specific names (nouns) is relatively balanced. (Severity: 3)
- Phrase frequency and Phrase ratio: The participant uses a moderate number of unique phrases. (Severity: 3)
- Lexical complexity: The participant's vocabulary is relatively simple, with the use of common terms and simple words. (Severity: 4)

Indicators based on Vocabulary Features:

1. Limited vocabulary: MCI (Severity: 3)
2. Increased verbosity: Probably normal cognition (Severity: 1)
3. Overlearned phrases: Probably normal cognition (Severity: 1)
4. Improper use of pronouns: Probably normal cognition (Severity: 1)
5. Hesitation (Word-finding difficulties): Probably normal cognition (Severity: 1)
6. Circumlocution (Word-finding difficulties): Probably normal cognition (Severity: 1)
7. Tip-of-the-tongue phenomenon: Probably normal cognition (Severity: 1)
8. Repetition: Probably normal cognition (Severity: 2)

Syntax and Grammar Features:

- Syntactic complexity: The participant uses relatively simple sentence structures and grammar. (Severity: 3)
- Grammatical components: The participant makes a few grammatical errors, such as missing verb tense agreement. (Severity: 3)

Indicators based on Syntax and Grammar Features:

1. Simplified sentence structure and grammar: MCI (Severity: 3)
2. Grammatical errors: MCI (Severity: 3)
3. Difficulty with complex ideas: Probably normal cognition (Severity: 1)

Semantics Features:

- Semantic density: The participant includes meaningful information but lacks depth in content words. (Severity: 4)
- Semantic coherence: The participant's narrative is relatively coherent and organized. (Severity: 2)

Indicators based on Semantics Features:

1. Semantic paraphasias: Probably normal cognition (Severity: 1)
2. Lack of coherence: Probably normal cognition (Severity: 2)
3. Consistency in errors: Probably normal cognition (Severity: 1)
4. Lack of semantic fluency: Probably normal cognition (Severity: 1)
5. Disorganized narrative structure: Probably normal cognition (Severity: 1)
6. Orientation issues: Probably normal cognition (Severity: 1)
7. Decline in conversational engagement and responsiveness: Probably normal cognition (Severity: 1)
8. Memory issues: Probably normal cognition (Severity: 1)
9. Lack of abstract thinking and synthesis: Probably normal cognition (Severity: 1)
10. Abnormal emotions: Probably normal cognition (Severity: 1)

Based on the comprehensive synthesis of these indicators, the participant is likely in the Probably Normal Cognition Group. However, it is important to note that not all indicators may be present, and further evaluation is necessary to confirm the cognitive status of the participant
