## Supplementary material for "Text dialogue analysis Based ChatGPT for Primary Screening of Mild Cognitive Impairment": Data(GPT4_vs_GPT3.5_model1-5): 10_GPT3.5 (4).docx

ChatGPT

Vocabulary features: a. Word frequency, word ratio, phrase frequency, and phrase ratio: It is challenging to analyze word and phrase frequency without a larger sample of the participant's language. The participant demonstrates the ability to describe various elements in the picture using appropriate vocabulary. b. Lexical complexity: The participant's lexical complexity appears to be moderate, with a mix of simple and descriptive words.

Severity rating for vocabulary features:

- Limited vocabulary: Not observed
- Increased verbosity: Not observed
- Overlearned phrases: Not observed
- Improper use of pronouns: Not observed
- Hesitation: Not observed
- Circumlocution: Not observed
- Tip-of-the-tongue phenomenon: Not observed
- Repetition: Not observed

Syntax and grammar features: a. Syntactic complexity and grammatical components: The participant demonstrates a moderate level of syntactic complexity, using relatively complex sentence structures with proper grammar. b. Assess and rate the severity of the following aspects: The participant does not exhibit simplified sentence structure, grammatical errors, or difficulty with complex ideas.

Severity rating for syntax and grammar features:

- Simplified sentence structure and grammar: Not observed
- Grammatical errors: Not observed
- Difficulty with complex ideas: Not observed

Semantics features: a. Semantic density and coherence: The participant's language exhibits moderate semantic density and coherence. They provide relevant and coherent information related to the picture and accurately describe the actions and elements present. b. Assess and rate the severity of the following aspects: The participant does not display semantic paraphasias, lack of coherence, lack of semantic fluency, disorganized narrative structure, orientation issues, decline in conversational engagement and responsiveness, memory issues, lack of abstract thinking and synthesis, or abnormal emotions.

Severity rating for semantics features:

- Lack of coherence: Not observed
- Consistency in errors: Not observed
- Lack of semantic fluency: Not observed
- Disorganized narrative structure: Not observed
- Orientation issues: Not observed
- Decline in conversational engagement and responsiveness: Not observed
- Memory issues: Not observed
- Lack of abstract thinking and synthesis: Not observed
- Abnormal emotions: Not observed

Based on the comprehensive synthesis of all the indicators, the participant can be classified into the Probably Normal Cognition Group. The observed language features suggest relatively normal vocabulary, syntax, and semantics. However, please note that a formal diagnosis can only be made through a comprehensive evaluation conducted by a qualified healthcare professional.
