## Supplementary material for "Text dialogue analysis Based ChatGPT for Primary Screening of Mild Cognitive Impairment": Data(GPT4_vs_GPT3.5_model1-5): 10_GPT3.5 (5).docx

b. Tip-of-the-tongue phenomenon: 1/10. The participant does not exhibit signs of the tip-of-the-tongue phenomenon. Their ability to recall words appears to be intact.

Syntax and grammar features: a. Syntactic complexity and grammatical components: 2/10. The participant's text demonstrates relatively simple sentence structures and overall grammatical accuracy. There are occasional errors and inconsistencies.

b. Difficulty with complex ideas: 1/10. The participant can handle simpler sentences and ideas effectively and does not display significant difficulty with complex ideas.

Semantics features: a. Semantic density and coherence: 3/10. The participant's text contains a moderate amount of meaningful information, and there is a reasonable balance between content words and function words. The coherence and cohesion of the text could be improved.

b. Memory issues: 2/10. The participant shows some minor difficulty in recalling specific details of the story, such as clothes or additional information. However, they can recall major plot points and characters relatively well.

Based on the comprehensive synthesis of these indicators, we can classify the participant into the Probably Normal Cognition Group. While there are some minor language features that may suggest possible mild cognitive impairment (MCI), the overall severity of these features is not significant enough to indicate a probable MCI. It's important to note that this assessment is based on a limited text sample, and a comprehensive evaluation by a qualified professional would be necessary for an accurate diagnosis
