## Supplementary material for "Text dialogue analysis Based ChatGPT for Primary Screening of Mild Cognitive Impairment": Data(GPT4_vs_GPT3.5_model1-5): 10_GPT3.5.docx

ChatGPT

Based on the conversation, here is my assessment of the Vocabulary, Semantics, Syntax and Grammar features of PARTICIPANT:

Vocabulary:

- Word frequency: Not assessed as it requires a larger text sample.
- Word ratio: 0.45 (low)
- Phrase frequency: Not assessed as it requires a larger text sample.
- Phrase ratio: Not assessed as it requires a larger text sample.
- Difficulty finding the right words: Not observed.
- Lexical complexity: Moderate, with a mix of common and less common words used.
- Limited vocabulary: Not observed, although there is a limited range of words used.

Semantics:

- Semantic density: Low, with a low proportion of content words.
- Inability to convey meaning: Not observed.
- Semantic coherence: Generally coherent, with clear relationships between words, phrases, and sentences.
- Lack of coherence: Not observed.

Syntax and Grammar:

- Syntactic complexity: Low, with mainly simple sentences and some use of subordinate clauses.
- Simplified sentence structure: Predominantly short, simple sentences.
- Grammatical components: Generally correct, with only a few errors observed.
- Grammatical errors: A few errors in verb tense observed.

Based on these assessments, it is difficult to make a definitive determination of whether the participant has mild cognitive impairment or not. However, the language features do not appear to indicate significant impairment. Therefore, the participant is most likely in the Probably Normal group
