## Supplementary material for "Text dialogue analysis Based ChatGPT for Primary Screening of Mild Cognitive Impairment": Data(GPT4_vs_GPT3.5_model1-5): 10_GPT4 (2).docx

ChatGPT

Analyzing the participant's language based on the provided criteria:

1. Vocabulary features:
   - Word frequency and Word ratio: The total word count is approximately 464 and the unique word count is about 172, giving a word ratio of roughly 0.37.
   - Phrase frequency and Phrase ratio: These are more complex to calculate manually and may require specific linguistic software.
   - Word-finding difficulties: The participant uses "uh" and "um" frequently. While these are common filler words, their frequency suggests the participant is taking time to gather thoughts.
   - Repetition: The participant repeats key scene details across prompts, but this seems to be in response to the investigator's repetitive requests.
   - Lexical complexity: The participant uses a variety of words and phrases, demonstrating good lexical complexity.
   - Limited vocabulary: There are repeated words (e.g., "cookie", "water", "stool", "boy", "girl", "sink") but this seems more due to the image content.
2. Semantics features:
   - Semantic density: The participant uses a high proportion of content words, providing specific details about the scene.
   - Inability to convey meaning: The participant appears to describe the scene clearly and accurately.
   - Semantic coherence: The participant's descriptions follow a logical flow, indicating good semantic coherence.
   - Lack of coherence: The participant doesn't show any significant difficulty in maintaining a logical flow of ideas.
3. Syntax and Grammar features:
   - Syntactic complexity: The participant uses a variety of sentence structures, including some more complex ones (e.g., "he's giving his sister one but in the meantime he's falling off the chair").
   - Simplified sentence structure: The participant uses a mix of sentence structures.
   - Grammatical components: The participant mostly uses correct grammar.
   - Grammatical errors: There are a few instances of missing conjunctions ("and", "but", etc.) and incomplete sentences, but these are not significant.

Given this evaluation, I would tentatively assign the PARTICIPANT to the Probably Normal group. The participant's responses are detailed, coherent, and demonstrate a good understanding of the task. However, the frequency of filler words like "uh" and "um" could suggest some word-finding difficulty. Please note that this is a preliminary evaluation based on a single dialogue and may not fully capture the participant's cognitive abilities. A comprehensive assessment should be conducted by a healthcare professional
