## Supplementary material for "Text dialogue analysis Based ChatGPT for Primary Screening of Mild Cognitive Impairment": Data(GPT4_vs_GPT3.5_model1-5): 10_GPT4 (3).docx

ChatGPT

Vocabulary Analysis:

1. Limited Vocabulary: 3/10 - The participant does have some repetition of words, but they also use a variety of words to describe the scene.
2. Increased Verbosity: 3/10 - The participant does repeat certain descriptions, but it doesn't significantly hinder the overall clarity of their narrative.
3. Overlearned Phrases: 1/10 - The participant doesn't seem to overuse cliches or stock phrases.
4. Improper Use of Pronouns: 2/10 - There's no significant misuse of pronouns in the conversation.
5. Hesitation (Word-finding difficulties): 2/10 - The participant demonstrates some hesitation, but this doesn't seem to impact their ability to describe the scene.
6. Circumlocution (Word-finding difficulties): 2/10 - There's no significant evidence of circumlocution.
7. Tip-of-the-tongue Phenomenon: 2/10 - The participant does not exhibit strong evidence of the tip-of-the-tongue phenomenon.
8. Repetition: 3/10 - Some repetition is observed, but it's not excessive.

Syntax and Grammar Analysis:

1. Simplified Sentence Structure and Grammar: 2/10 - The participant uses both simple and complex sentences to describe the scene.
2. Grammatical Errors: 1/10 - The participant makes very few grammatical errors.
3. Difficulty with Complex Ideas: 2/10 - The participant seems to understand the complex narrative of the picture and communicates it well.

Semantics Analysis:

1. Semantic Paraphasias: 2/10 - There's no significant evidence of semantic paraphasias in the conversation.
2. Lack of Coherence: 2/10 - The participant demonstrates good coherence in their narrative.
3. Consistency in Errors: 1/10 - There's no consistent pattern of errors observed.
4. Lack of Semantic Fluency: 1/10 - The participant is fluent in their narration.
5. Disorganized Narrative Structure: 2/10 - The participant's narrative is mostly organized and logical.
6. Orientation Issues: 1/10 - The participant does not show any signs of confusion about the sequence of events in the picture.
7. Decline in Conversational Engagement and Responsiveness: 1/10 - The participant remains engaged and responsive throughout the conversation.
8. Memory Issues: 2/10 - The participant repeats some details but does not demonstrate significant issues with memory.
9. Lack of Abstract Thinking and Synthesis: 1/10 - The participant is able to synthesize the overall situation in the picture.
10. Abnormal Emotions: 2/10 - There's no significant evidence of abnormal emotions in the conversation.

Based on the evaluation of the various cognitive and linguistic indicators, it appears that the participant probably belongs to the Normal Cognition Group. While the participant does exhibit a few minor issues, such as slight repetition and some hesitation, these do not significantly impair their ability to describe the scene or engage in the conversation. Moreover, they do not present any severe or consistent patterns of linguistic or cognitive difficulties. Please note that this analysis is based on the text provided and might not capture all aspects of the participant's cognitive abilities. A formal evaluation by a qualified professional is necessary for an accurate diagnosis
