## Supplementary material for "Text dialogue analysis Based ChatGPT for Primary Screening of Mild Cognitive Impairment": Data(GPT4_vs_GPT3.5_model1-5): 10_GPT4 (4).docx

ChatGPT

Based on the conversation you provided, I will attempt to rate the severity of each evaluation indicator on a scale of 1 to 10 for the participant.

Vocabulary features: a. Word frequency, word ratio, phrase frequency, and phrase ratio: These can't be directly evaluated in the current context, as they require quantitative linguistic analysis software. However, the participant seems to use a variety of words and phrases suggesting a decent vocabulary. b. Lexical complexity: Again, without software this is hard to quantify precisely, but from the presented conversation, the participant demonstrates a good range of vocabulary suggesting a moderate to high lexical complexity. c. Severity of aspects:

- Limited vocabulary: 2
- Increased verbosity: 3
- Overlearned phrases: 1
- Improper use of pronouns: 1
- Hesitation: 3
- Circumlocution: 2
- Tip-of-the-tongue phenomenon: 2
- Repetition: 3

Syntax and grammar features: a. Syntactic complexity and grammatical components: The participant forms complex sentences with multiple clauses and makes use of conjunctions and prepositions, indicating a good command of syntax and grammar. b. Severity of aspects:

- Simplified sentence structure and grammar: 2
- Grammatical errors: 1
- Difficulty with complex ideas: 2

Semantic features: a. Semantic density and coherence: The participant seems to have a coherent narrative with dense semantic content, describing the picture in detail. b. Severity of aspects:

- Semantic paraphasias: 1
- Lack of coherence: 2
- Consistency in errors: 1
- Lack of semantic fluency: 2
- Disorganized narrative structure: 2
- Orientation issues: 1
- Decline in conversational engagement and responsiveness: 2
- Memory issues: 2
- Lack of abstract thinking and synthesis: 2
- Abnormal emotions: 1

Based on the comprehensive synthesis of all the indicators, the participant would likely fall into the Probably Normal Cognition Group. However, please consider this evaluation as a preliminary observation and not a diagnostic conclusion. A more formal evaluation might require further data and more quantitative methods. The assessment seems to show mild indicators of possible MCI, but they are generally low and within the range that could be considered normal. As always, professional judgment and a comprehensive assessment, potentially including additional cognitive and medical evaluations, would be needed for a more definitive understanding of the participant's cognitive status
