## Supplementary material for "Text dialogue analysis Based ChatGPT for Primary Screening of Mild Cognitive Impairment": Data(GPT4_vs_GPT3.5_model1-5): 10_GPT4 (5).docx

ChatGPT

Based on the provided text, here are my observations:

Vocabulary features:

a. Word frequency, word ratio, phrase frequency, and phrase ratio: The participant does use a variety of words, with common words such as "cookie", "dishes", "sink", "boy", and "mother" repeated. In terms of pronouns to specific names (or nouns), the participant does make a fair usage of pronouns, but not excessively. There are multiple phrases repeated, but this is due to the nature of describing a single scene. Overall, these factors seem within the expected range.

Lexical complexity: The participant uses a reasonable mix of simple and complex words, indicating a moderate lexical complexity.

Rating: 3/10

b. Tip-of-the-tongue phenomenon: The participant shows instances of partial recall ("xxx at her dishwasher's or …", "xxx curtains on the windows are tied back"). These instances could potentially indicate some issues with recall but they're not excessively frequent.

Rating: 4/10

Syntax and grammar features:

a. Syntactic complexity and grammatical components: The participant's sentences show a fair amount of complexity, including the use of subordinate clauses. Their grammar seems correct most of the time, albeit with a few minor errors or informalities.

Rating: 3/10

b. Difficulty with complex ideas: The participant appears to understand and articulate complex scenes within the image well. They are able to identify multiple actions and their potential consequences (e.g., the boy falling off the stool while reaching for cookies), indicating an ability to handle complex narratives.

Rating: 2/10

Semantics features:

a. Semantic density and coherence: The participant's responses are semantically dense and coherent. They are able to convey a lot of information about the scene, and their description makes logical sense.

Rating: 2/10

b. Memory issues: The participant does show some signs of repeating themselves, especially when asked to recall what they've said. They might have some difficulty in recalling their exact previous statements.

Rating: 4/10

Based on a comprehensive synthesis of all these indicators, it is challenging to classify the participant definitively. However, considering the relatively low severity scores in most aspects, it seems more likely that the participant falls into the Probably Normal Cognition Group. They exhibit some possible signs of MCI, but these are not pervasive or severe enough to suggest a clear shift from normal aging. Nevertheless, it's important to note that this is an informal analysis and not a diagnosis. Any concerns about cognitive function should be discussed with a healthcare professional.
