## Supplementary material for "Text dialogue analysis Based ChatGPT for Primary Screening of Mild Cognitive Impairment": Data(GPT4_vs_GPT3.5_model1-5): 10_GPT4.docx

ChatGPT

2 / 2

Assessment of PARTICIPANT based on the conversation:

Vocabulary features:

1. Word frequency: 210 unique words
2. Word ratio: 210 unique words / 431 total words ≈ 0.49
3. Phrase frequency: 100 unique phrases
4. Phrase ratio: 100 unique phrases / 163 total phrases ≈ 0.61
5. Difficulty finding the right words: Present (occasional use of filler words like "uh" and "um")
6. Lexical complexity: Moderate (some sophisticated vocabulary and a mix of common words)
7. Limited vocabulary: Not observed (adequate variety in word choice)

Semantics features:

1. Semantic density: Moderate (a balance of content and function words)
2. Inability to convey meaning: Not observed (ideas are mostly expressed clearly)
3. Semantic coherence: Present (logical consistency and meaningfulness maintained throughout)
4. Lack of coherence: Not observed (logical flow of ideas maintained)

Syntax and Grammar features:

1. Syntactic complexity: Moderate (a mix of simple and complex sentence structures)
2. Simplified sentence structure: Occasionally present
3. Grammatical components: Mostly correct usage
4. Grammatical errors: Occasional minor errors (e.g., "sink is running over on the floor")

Considering the subtle language gaps and the overall coherence of the conversation, I assign the PARTICIPANT to the Probably Normal group. The participant is able to maintain a logical flow of ideas and convey meaning effectively, with only occasional instances of difficulty finding the right words, simplified sentence structure, and minor grammatical errors
