## Supplementary material for "Text dialogue analysis Based ChatGPT for Primary Screening of Mild Cognitive Impairment": Data(GPT4_vs_GPT3.5_model1-5): 11_GPT3.5 (2).docx

Attention! You do not need to diagnose, just follow the steps below to initially screen for PARTICIPANT with possibly mild cognitive impairment (MCI). Some differences between MCI and normal cognition are also given, please analyse them in detail, step by step. Firstly, please assess the Vocabulary features of PARTICIPANT at the Word frequency and Word ratio, Phrase frequency and Phrase ratio, Word-finding difficulties, Repetition, Lexical complexity, Limited vocabulary. Word frequency: Count the occurrences of each unique word in the text. Unique words are distinct words that appear in the text, not taking into account their repetitions. Each word is counted only once, regardless of how many times it occurs. Word ratio: Calculate the proportion of unique words to the total number of words in the text. Phrase frequency: Count the occurrences of each unique phrases in the text. Unique phrases are distinct combinations of words that appear in the text, not taking into account their repetitions. Phrase ratio: Calculate the proportion of unique phrases to the total number of phrases in the text. Word-finding difficulties: Frequent pauses or use of filler words, like "uh" or "um,". MCI: more frequent instances, longer time to find the right word, or using more general terms. Normal: fewer instances, and better lexical retrieval. Repetition: The participant repeats certain phrases or ideas a few times throughout the conversation. MCI: more frequent repetition. Normal: less frequent repetition (some repetition of words but not excessive) and better memory of what has been mentioned. Lexical complexity: Measure the variety and sophistication of the vocabulary used, considering factors, including word length and frequency of rare words. Limited vocabulary: Repeated use of the same words or simple words. MCI: slightly reduced vocabulary, and more frequent use of simpler words or common terms; Normal: richer vocabulary (low limitation). Secondly, please assess the Semantics features of PARTICIPANT, from Semantic density and Inability to convey meaning, Semantic coherence and Lack of coherence. Semantic density: Measure the amount of meaningful information in the text, and considering factor is the number of content words (nouns, verbs, adjectives, adverbs) relative to function words (prepositions, pronouns, conjunctions, etc.). Inability to convey meaning: Difficulty in expressing ideas clearly, leading to confusion or misunderstanding. Normal: the participant was able to describe the scene clearly. Semantic coherence: Assess the logical consistency and meaningfulness of the text, which can be done by analyzing the relationships between words, phrases, and sentences, and checking for coherence and cohesion. Lack of coherence: Difficulty in maintaining a logical flow of ideas, with thoughts appearing disjointed or unrelated. MCI: struggle with organizing thoughts coherently or maintaining a clear focus in conversation; Normal: greater coherence and organization in their storytelling and conversations. Thirdly, please assess the Syntax and Grammar features of PARTICIPANT, from Syntactic complexity and Simplified sentence structure, Grammatical components and Grammatical errors. Syntactic complexity: Measure the complexity of the sentence structure in the text, considering factors such as the number of clauses per sentence, the average length of sentences, and the use of subordinate clauses. Simplified sentence structure: A lack of complex sentence structures. MCI: simpler sentence structures; Normal: in longer dialogues there is a greater variety of sentence structures (moderately complex), but in short dialogues the sentences may turn out to be simple. Grammatical components: Analyze the text for correct usage of grammar, such as subject-verb agreement, tense consistency, and proper use of pronouns, prepositions, and conjunctions. MCI: avoidance of complex grammatical constructions; Normal: better use of complex grammatical constructions. Grammatical errors: Frequent mistakes in verb tense, subject-verb agreement, or other aspects of grammar. MCI: more frequent grammatical errors, such as mistakes in verb tense and subject-verb agreement; Normal: fewer grammatical errors (a few errors, but not significant). Finally, Please dichotomise PARTICIPANT into the Probably Normal group or the Probably Mild Cognitive Impairment group. PARTICIPANT with MCI might exhibit more difficulty with vocabulary, sentence structure, repetition, word-finding, coherence, and grammar. However, it is not definitive and requires a comprehensive judgement. INVESTIGATOR: alright can you just tell me everything that you see going on in this picture ? PARTICIPANT: okay . PARTICIPANT: looks like a kitchen . PARTICIPANT: mom …uh washing or drying dishes . PARTICIPANT: the sink overflowing . PARTICIPANT: the …uh little boy trying to get in the cookie jar . PARTICIPANT: the stool tipping over . PARTICIPANT: and the little girl wants a share of the cookie wealth . INVESTIGATOR: perfect . INVESTIGATOR: alright so for the next one , this picture tells a story . INVESTIGATOR: I'd like you to look at everything that's happening and then tell me a story about what you see with a beginning , middle , and end . PARTICIPANT: looks like the … a cat climbed up in the tree . PARTICIPANT: the little girl …uh who was riding her bicycle wants the cat down . PARTICIPANT: she called her daddy . PARTICIPANT: put up the ladder which fell down . PARTICIPANT: the dog was barking . PARTICIPANT: bird's singing . PARTICIPANT: and it looks like somebody called the fire department to rescue the dad and the cat . INVESTIGATOR: perfect . INVESTIGATOR: and then I have one more picture …um that also tells a story . INVESTIGATOR: and if you can look at everything that's happening and tell me a story with a beginning , middle , and end . PARTICIPANT: looks like a family outing . PARTICIPANT: typical Norman_Rockwell the way it looks . PARTICIPANT: …uh dad driving . PARTICIPANT: mother riding shotgun . PARTICIPANT: little boy sticking his head out . PARTICIPANT: one little boy sticking his head out the window . PARTICIPANT: a little girl blowing bubbles . PARTICIPANT: and one giving …uh a …uh nasty sign to somebody behind them . PARTICIPANT: and then there's another picture . PARTICIPANT: dad driving , maybe smoking . PARTICIPANT: little girl riding in between . PARTICIPANT: an:d well they're probably asleep . PARTICIPANT: same little boy looking out the window with a …del dog in the middle . PARTICIPANT: girl blowing bubbles . PARTICIPANT: and grandma in the back with the little kid keeping him quiet . INVESTIGATOR: perfect . INVESTIGATOR: alright and then the next story that you're gonna tell me is the story of Cinderella . INVESTIGATOR: have you heard this story before ? PARTICIPANT: yes but not the new one on tv . INVESTIGATOR: me either . INVESTIGATOR: …um would you like some pictures to jog your memory or would you like to just tell me the story of Cinderella ? PARTICIPANT: well she was …uh I believe . taken in by a … a more wealthy family to do housework . PARTICIPANT: and the family's daughters were quite mean to her and disrespectful . PARTICIPANT: and …uh somehow I don't remember how she got to …uh go to the ball . PARTICIPANT: and had glass slippers . PARTICIPANT: and at midnight she hadta leave because her ride was gonna turn into a pumpkin and she was going to be returned to her housework dress . PARTICIPANT: … I guess she did marry the prince or something . INVESTIGATOR: you got it . INVESTIGATOR: alright so the next thing is gonna be a little bit different . INVESTIGATOR: is … I would just like you to tell me how to make a peanut butter and jelly sandwich . PARTICIPANT: okay . PARTICIPANT: well preferably it takes two slices of bread . PARTICIPANT: we'll put a_lots of peanut butter on one and some jelly on the other slice . PARTICIPANT: and push the two of (th)em together . PARTICIPANT: and cut (th)em in half .
