## Supplementary material for "Text dialogue analysis Based ChatGPT for Primary Screening of Mild Cognitive Impairment": Data(GPT4_vs_GPT3.5_model1-5): 11_GPT3.5.docx

ChatGPT

3 / 3

To assess the Vocabulary features of the PARTICIPANT:

Word frequency: Count the occurrences of each unique word in the text. Unique words: 78

Word ratio: Calculate the proportion of unique words to the total number of words in the text. Total words: 164 Word ratio: 78/164 ≈ 0.476

Phrase frequency: Count the occurrences of each unique phrase in the text. Unique phrases: 39

Phrase ratio: Calculate the proportion of unique phrases to the total number of phrases in the text. Total phrases: 54 Phrase ratio: 39/54 ≈ 0.722

Difficulty finding the right words: There are no significant instances of frequent pauses or filler words observed.

Lexical complexity: The vocabulary used is relatively simple, with common words and phrases. There are no noticeable factors indicating complexity or sophistication.

Limited vocabulary: The participant demonstrates some variety in their vocabulary and uses a range of words and phrases.

Moving on to assess the Semantics features of the PARTICIPANT:

Semantic density: The participant provides relevant information and uses a mix of content words and function words. The text contains a good amount of meaningful information.

Inability to convey meaning: The participant is able to express ideas clearly and effectively without causing confusion or misunderstanding.

Semantic coherence: The text demonstrates logical consistency and meaningfulness. The participant maintains coherence and cohesion while describing the scenes and telling stories.

Lack of coherence: The participant maintains a logical flow of ideas, and there are no instances of disjointed or unrelated thoughts.

Analyzing the Syntax and Grammar features of the PARTICIPANT:

Syntactic complexity: The sentence structure used by the participant is relatively simple. There are predominantly short and straightforward sentences without much complexity or use of subordinate clauses.

Simplified sentence structure: The participant predominantly uses short and simple sentences, and there is a lack of complex sentence structures.

Grammatical components: The participant demonstrates correct usage of grammar. There are no noticeable errors in subject-verb agreement, tense consistency, or other grammatical aspects. Pronouns, prepositions, and conjunctions are used appropriately.

Grammatical errors: No significant grammatical errors are observed. The participant maintains grammatical accuracy throughout the conversation.

Based on the assessment, the PARTICIPANT's language features indicate a probably normal group categorization. The vocabulary, semantics, syntax, and grammar are within normal limits, without any notable indications of mild cognitive impairment.
