## Supplementary material for "Text dialogue analysis Based ChatGPT for Primary Screening of Mild Cognitive Impairment": Data(GPT4_vs_GPT3.5_model1-5): 11_GPT4 (2).docx

ChatGPT

Assessment based on the given parameters:

1. **Vocabulary features:**
   - Word frequency: The participant uses a set of recurring words such as "little", "girl", "boy", "dad", "window", "driving", "bubbles", "story", "riding", "looks".
   - Word ratio: The ratio of unique words to the total number of words is high. There is some repetition but it's not excessive.
   - Phrase frequency: The phrases like "looks like", "little boy", "little girl", "dad driving" are repeated.
   - Phrase ratio: The proportion of unique phrases to the total number of phrases is high.
   - Word-finding difficulties: There are a few instances of word-finding difficulties, with some pauses and filler words ("uh").
   - Repetition: The participant repeats certain phrases or ideas a few times throughout the conversation, but it's not excessive.
   - Lexical complexity: The participant uses a relatively simple vocabulary, but it's varied and appropriate for the context.
   - Limited vocabulary: The participant uses a good variety of words and does not overly rely on a limited set of terms.
2. **Semantic features:**
   - Semantic density: The text contains a lot of meaningful information, with a good balance of content and function words.
   - Inability to convey meaning: The participant is able to express the ideas clearly; there doesn't seem to be any significant difficulty in conveying meaning.
   - Semantic coherence: The participant maintains semantic coherence throughout the description of the pictures and the storytelling.
   - Lack of coherence: No significant lack of coherence is observed.
3. **Syntax and Grammar features:**
   - Syntactic complexity: The sentences used by the participant are simple to moderate in complexity, with a few instances of more complex sentence structures.
   - Simplified sentence structure: The participant uses simple to moderately complex sentences.
   - Grammatical components: There are a few instances where the participant drops the 'g' from words ending in 'ing'. This is more of a dialectic variation than a grammatical error.
   - Grammatical errors: There are no significant grammatical errors in the participant's language production.

Given the observed characteristics, the participant shows some signs of word-finding difficulties, but also maintains a high degree of coherence and is able to convey meaning effectively. The participant's sentence structures are simple to moderate in complexity, and vocabulary is varied and contextually appropriate. There's some repetition, but it's not excessive.

**Overall, it seems likely that this participant falls into the Probably Normal group.** However, it's important to remember that such assessment can't replace a comprehensive clinical evaluation, as language abilities can vary among individuals and can be influenced by many factors not captured in this analysis
