## Supplementary material for "Text dialogue analysis Based ChatGPT for Primary Screening of Mild Cognitive Impairment": Data(GPT4_vs_GPT3.5_model1-5): 11_GPT4 (3).docx

ChatGPT

Based on the provided text, here's the analysis:

**Vocabulary Features:** Word frequency and Word ratio: The participant used a variety of words and pronouns. There was no significant overuse of specific words or phrases. The ratio of pronouns to specific names or nouns seems to be appropriate for normal conversation. Phrase frequency and Phrase ratio: The participant used different phrases without significant repetition. Lexical complexity: The participant used a mix of simple and complex words, and there was no apparent overuse of simple or common terms.

1. Limited vocabulary: 2 (The participant demonstrated a good variety of vocabulary.)
2. Increased verbosity: 1 (The participant was concise and to the point.)
3. Overlearned phrases: 1 (There was no apparent overuse of cliches or stock phrases.)
4. Improper use of pronouns: 1 (The participant used pronouns appropriately.)
5. Hesitation (Word-finding difficulties): 4 (The participant had some hesitations and used filler words like "uh".)
6. Circumlocution (Word-finding difficulties): 2 (The participant was mostly direct and specific in their responses.)
7. Tip-of-the-tongue phenomenon: 2 (The participant had minor difficulty recalling details of the Cinderella story.)
8. Repetition: 3 (The participant repeated some phrases and words but not excessively.)

**Syntax and Grammar Features:** Syntactic complexity: The participant used a mixture of simple and complex sentence structures. Grammatical components: The participant made no significant grammatical errors.

1. Simplified sentence structure and grammar: 2 (The participant used a variety of sentence structures.)
2. Grammatical errors: 1 (The participant made no significant grammatical errors.)
3. Difficulty with complex ideas: 2 (The participant was able to describe complex scenes and tell stories with a beginning, middle, and end.)

**Semantics Features:** Semantic density: The participant's responses included a good amount of meaningful information, with a balance of content and function words. Semantic coherence: The participant's responses were logically consistent and meaningful.

1. Semantic paraphasias: 1 (The participant expressed ideas clearly and correctly.)
2. Lack of coherence: 1 (The participant's thoughts were organized and coherent.)
3. Consistency in errors: 1 (No consistent language errors were noticed.)
4. Lack of semantic fluency: 2 (The participant had some trouble naming items in the Cinderella story.)
5. Disorganized narrative structure: 1 (The participant maintained a coherent line of thought in their narratives.)
6. Orientation Issues: 1 (The participant showed no confusion about time, place, or sequence of events.)
7. Decline in conversational engagement and responsiveness: 1 (The participant actively engaged in the conversation.)
8. Memory issues: 3 (The participant had some difficulty recalling the story of Cinderella.)
9. Lack of abstract thinking and synthesis: 1 (The participant was able to synthesize the overall situations in the pictures.)
10. Abnormal emotions: 1 (No abnormal emotions were noticed.)

Based on this analysis, it seems that the participant falls into the "Probably Normal Cognition Group". The minor hesitations, some repetitions, and slight difficulty in recalling the story of Cinderella are not unusual and may occur in normal cognitive participants. However, it's important to remember that this assessment is based on a single interaction and more comprehensive assessments may be needed for a definitive categorization
