## Supplementary material for "Text dialogue analysis Based ChatGPT for Primary Screening of Mild Cognitive Impairment": Data(GPT4_vs_GPT3.5_model1-5): 11_GPT4 (4).docx

ChatGPT

Based on the participant's responses, here is a preliminary evaluation of their language features:

1. Vocabulary features: a. Word and phrase frequency and ratio: The participant used a diverse set of words and phrases, with no obvious repeats or over-reliance on certain phrases, indicating a rich vocabulary. They also used certain complex expressions like "typical Norman Rockwell the way it looks". Severity Score: 2 b. Lexical complexity: The participant's lexical complexity appears to be fairly typical, using a combination of common and more complex words. Severity Score: 2 c. Limited vocabulary, increased verbosity, overlearned phrases, improper use of pronouns, hesitation, circumlocution, tip-of-the-tongue phenomenon, repetition: The participant did not show signs of limited vocabulary, overlearned phrases, or repetition. However, there were instances of slight hesitation and circumlocution, especially when recalling the Cinderella story. Severity Score: 3
2. Syntax and grammar features: a. Syntactic complexity and grammatical components: The participant used a mix of simple and complex sentence structures and followed appropriate grammatical rules. Severity Score: 2 b. Simplified sentence structure and grammar, grammatical errors, difficulty with complex ideas: The participant did not show signs of simplified sentence structure or grammatical errors, and they were able to communicate complex ideas effectively. Severity Score: 2
3. Semantics features: a. Semantic density and coherence: The participant maintained good coherence throughout their responses and used a variety of words contributing to a high semantic density. Severity Score: 2 b. Semantic paraphasias, lack of coherence, consistency in errors, lack of semantic fluency, disorganized narrative structure, orientation issues, decline in conversational engagement and responsiveness, memory issues, lack of abstract thinking and synthesis, abnormal emotions: The participant did not display semantic paraphasias, lack of coherence, consistency in errors, lack of semantic fluency, or disorganized narrative structure. They had minor difficulty recalling the Cinderella story which might indicate some memory issues. Severity Score: 3

Based on the comprehensive synthesis of all the indicators, the participant would be classified into the Probably Normal Cognition Group. It's worth noting that this evaluation is not meant to provide a formal diagnosis but to serve as an initial screening tool for potential MCI. It would be beneficial for the participant to undergo further examination if there are additional concerns about their cognitive abilities.
