## Supplementary material for "Text dialogue analysis Based ChatGPT for Primary Screening of Mild Cognitive Impairment": Data(GPT4_vs_GPT3.5_model1-5): 11_GPT4 (5).docx

ChatGPT

Vocabulary Features: a. Word frequency, word ratio, phrase frequency, phrase ratio, and lexical complexity are difficult to fully calculate from the given text but based on the participant's usage of a variety of words, repeated use of some words, and use of various phrases, lexical complexity can be estimated as moderate. The participant uses a few different nouns ("girl", "cat", "tree") and verbs ("called", "riding", "climbing"), but the complexity of the language isn't very high, indicating the lexical complexity as approximately a 5 out of 10. b. Regarding the tip-of-the-tongue phenomenon, there are a few instances where the participant seems to struggle with finding the right word (e.g., "...uh" interjections). However, it's not clear whether this is due to a struggle with word retrieval or natural speech patterns. Rating for tip-of-the-tongue phenomenon: 3 out of 10.

Syntax and Grammar Features: a. Syntactic complexity and grammatical components are acceptable. The participant uses fairly simple sentence structures but is able to produce syntactically complete and correct sentences. Some of the participant's responses are more complex than others, indicating some variability in the complexity of their narrative. Rating for syntactic complexity: 4 out of 10. b. The participant appears to handle simple sentences better than complex ones. There's no evidence of a severe difficulty with complex ideas; most of the storylines are conveyed, albeit with simplicity. Rating for difficulty with complex ideas: 4 out of 10.

Semantic Features: a. Semantic density and coherence: The participant provides coherent narratives for the images and story recall, and there's a reasonable amount of content information relative to function words. Rating for semantic density and coherence: 4 out of 10. b. There are several instances where the participant repeats phrases or appears to struggle with recalling certain details (e.g., Cinderella's story), suggesting some possible memory issues. However, the participant still recalls most of the key aspects of each narrative. Rating for memory issues: 4 out of 10.

Based on these evaluations, the participant demonstrates some mild cognitive difficulties, particularly with lexical complexity and memory issues. However, these challenges don't seem to significantly impair their ability to communicate or understand narratives. Therefore, it's plausible that the participant falls under the "Probably Mild Cognitive Impairment Group," but the mildness of these issues means this is not certain without further assessment. Keep in mind that this evaluation is not a formal diagnosis but a screening tool. Consultation with a healthcare professional is essential for a comprehensive assessment.
