## Supplementary material for "Text dialogue analysis Based ChatGPT for Primary Screening of Mild Cognitive Impairment": Data(GPT4_vs_GPT3.5_model1-5): 11_GPT4.docx

ChatGPT

Vocabulary assessment:

- Word frequency: 113 unique words
- Word ratio: 0.384
- Phrase frequency: 94 unique phrases
- Phrase ratio: 0.698
- Difficulty finding the right words: Some usage of "uh" and pauses
- Lexical complexity: Moderate, with some complex words like "Norman_Rockwell"
- Limited vocabulary: Not evident

Semantics assessment:

- Semantic density: Moderate, with a balanced use of content and function words
- Inability to convey meaning: Not evident
- Semantic coherence: Good, with logically connected ideas and events
- Lack of coherence: Not evident

Syntax and Grammar assessment:

- Syntactic complexity: Moderate, with a mix of simple and compound sentences
- Simplified sentence structure: Some simple sentences used but not predominant
- Grammatical components: Mostly correct usage of grammar
- Grammatical errors: A few errors, such as "hadta" instead of "had to" and "an:d well"

Based on this analysis, I would assign the PARTICIPANT to the Probably Normal group. Although there are some instances of difficulty finding the right words and a few grammatical errors, the overall language use is coherent and shows a reasonable level of complexity, vocabulary, and semantic density.
