## Supplementary material for "Text dialogue analysis Based ChatGPT for Primary Screening of Mild Cognitive Impairment": Data(GPT4_vs_GPT3.5_model1-5): 100_GPT3.5 (2).docx

Attention! You do not need to diagnose, just follow the steps below to initially screen for PARTICIPANT with possibly mild cognitive impairment (MCI). Some differences between MCI and normal cognition are also given, please analyse them in detail, step by step. Firstly, please assess the Vocabulary features of PARTICIPANT at the Word frequency and Word ratio, Phrase frequency and Phrase ratio, Word-finding difficulties, Repetition, Lexical complexity, Limited vocabulary. Word frequency: Count the occurrences of each unique word in the text. Unique words are distinct words that appear in the text, not taking into account their repetitions. Each word is counted only once, regardless of how many times it occurs. Word ratio: Calculate the proportion of unique words to the total number of words in the text. Phrase frequency: Count the occurrences of each unique phrases in the text. Unique phrases are distinct combinations of words that appear in the text, not taking into account their repetitions. Phrase ratio: Calculate the proportion of unique phrases to the total number of phrases in the text. Word-finding difficulties: Frequent pauses or use of filler words, like "uh" or "um,". MCI: more frequent instances, longer time to find the right word, or using more general terms. Normal: fewer instances, and better lexical retrieval. Repetition: The participant repeats certain phrases or ideas a few times throughout the conversation. MCI: more frequent repetition. Normal: less frequent repetition (some repetition of words but not excessive) and better memory of what has been mentioned. Lexical complexity: Measure the variety and sophistication of the vocabulary used, considering factors, including word length and frequency of rare words. Limited vocabulary: Repeated use of the same words or simple words. MCI: slightly reduced vocabulary, and more frequent use of simpler words or common terms; Normal: richer vocabulary (low limitation). Secondly, please assess the Semantics features of PARTICIPANT, from Semantic density and Inability to convey meaning, Semantic coherence and Lack of coherence. Semantic density: Measure the amount of meaningful information in the text, and considering factor is the number of content words (nouns, verbs, adjectives, adverbs) relative to function words (prepositions, pronouns, conjunctions, etc.). Inability to convey meaning: Difficulty in expressing ideas clearly, leading to confusion or misunderstanding. Normal: the participant was able to describe the scene clearly. Semantic coherence: Assess the logical consistency and meaningfulness of the text, which can be done by analyzing the relationships between words, phrases, and sentences, and checking for coherence and cohesion. Lack of coherence: Difficulty in maintaining a logical flow of ideas, with thoughts appearing disjointed or unrelated. MCI: struggle with organizing thoughts coherently or maintaining a clear focus in conversation; Normal: greater coherence and organization in their storytelling and conversations. Thirdly, please assess the Syntax and Grammar features of PARTICIPANT, from Syntactic complexity and Simplified sentence structure, Grammatical components and Grammatical errors. Syntactic complexity: Measure the complexity of the sentence structure in the text, considering factors such as the number of clauses per sentence, the average length of sentences, and the use of subordinate clauses. Simplified sentence structure: A lack of complex sentence structures. MCI: simpler sentence structures; Normal: in longer dialogues there is a greater variety of sentence structures (moderately complex), but in short dialogues the sentences may turn out to be simple. Grammatical components: Analyze the text for correct usage of grammar, such as subject-verb agreement, tense consistency, and proper use of pronouns, prepositions, and conjunctions. MCI: avoidance of complex grammatical constructions; Normal: better use of complex grammatical constructions. Grammatical errors: Frequent mistakes in verb tense, subject-verb agreement, or other aspects of grammar. MCI: more frequent grammatical errors, such as mistakes in verb tense and subject-verb agreement; Normal: fewer grammatical errors (a few errors, but not significant). Finally, Please dichotomise PARTICIPANT into the Probably Normal group or the Probably Mild Cognitive Impairment group. PARTICIPANT with MCI might exhibit more difficulty with vocabulary, sentence structure, repetition, word-finding, coherence, and grammar. However, it is not definitive and requires a comprehensive judgement. Now, you do not have to respond to this dialogue. When I have entered THE DIALOGUE BETWEEN PARTICIPANT AND INVESTIGATOR, evaluate PARTICIPANT according to the evaluation rules above. INVESTIGATOR: so I'd just like you to tell me everything that you see going on in this picture . PARTICIPANT: I see that … the … their overflowing sink . PARTICIPANT: … …c the boy is …ge …s …gi stealing cookies from the cookie jar which is up in the cabinet . PARTICIPANT: the mother is standing there unaware of what's going on , busy …uh washing dishes or at least drying it , with something or other with the dish . PARTICIPANT: …uh outside they … it looks like they have some playground stuff outside . PARTICIPANT: I don't know what that thing is down there . PARTICIPANT: …um yeah …n and there's a tree outside . PARTICIPANT: and they got the curtains . PARTICIPANT: and let's see . PARTICIPANT: what else do we want to know ? PARTICIPANT: little girl's got …uh socks and shoes on . PARTICIPANT: the … so has the kid … the boy . PARTICIPANT: he's got shorts on . PARTICIPANT: …um yeah look … I … I see a face out there …uh in the ground … what looks like in the ground of the yard . PARTICIPANT: but I don't know whether that's sposta be a reflection off the glass or … or whether it's somebody out there in the yard , a reflection in the yard , or something they've made out_of the lawn . PARTICIPANT: I don't know . PARTICIPANT: …um there's some dishes on the counter . PARTICIPANT: let's see . PARTICIPANT: what else there would be xxx . PARTICIPANT: she has gotten wet feet , by the way , the lady . PARTICIPANT: …um I can't tell what's … what else that stuff is out . PARTICIPANT: there's a tree out there and I think I said that already . PARTICIPANT: and I don't know whether it looks like a jungle gym or what . PARTICIPANT: …um I don't see anything else . PARTICIPANT: well wait … wait a minute . PARTICIPANT: the girl's . PARTICIPANT: no . PARTICIPANT: no I … I guess that's her hair but I'm not sure whether …sh she's holding something or not . PARTICIPANT: can't tell . PARTICIPANT: she may be holding a dollar or something . PARTICIPANT: it's not clear . PARTICIPANT: the lady has …uh an apron on . PARTICIPANT: I don't know . PARTICIPANT: that's about it , I think . INVESTIGATOR: okay that works . INVESTIGATOR: so this picture tells a story . PARTICIPANT: yes it does . INVESTIGATOR: I want you to look at everything that's happening and then tell me a story about what you see . INVESTIGATOR: and the story should have a beginning a middle and an end . PARTICIPANT: oh_my_goodness . PARTICIPANT: okay well let's see . PARTICIPANT: the little girl was out with … on her bike . PARTICIPANT: and …um I don't know whether it's her cat or not . PARTICIPANT: but this dog came along and chased the cat up the tree . PARTICIPANT: and she wanted to get the … …ki the cat down . PARTICIPANT: so her dad , I assume to be her dad , or at least some guy , came along , got … went and got a ladder . PARTICIPANT: but once he got up in the tree …s some reason or another the ladder fell down . PARTICIPANT: and he didn't get the cat out_of the tree . PARTICIPANT: now he was stuck up in the tree too . PARTICIPANT: so somebody who saw what was going on . PARTICIPANT: or maybe it was the little bird in the tree that called the fire department . PARTICIPANT: and the fire department brought their truck . PARTICIPANT: and two firemen got out . PARTICIPANT: and they've got the ladder . PARTICIPANT: and hopefully they … they get both the man and the cat down and find out whose dog it is . INVESTIGATOR: so I want you to look at both of these pictures . INVESTIGATOR: …um and this whole thing tells a story . INVESTIGATOR: and so I'd like you to look at everything that's happening and tell me a story about what you see . INVESTIGATOR: and the story should have a beginning a middle and an end . PARTICIPANT: okay hm alright . PARTICIPANT: well I think what happened was that they , …um in the top picture , were ridin(g) along , going on …uh …uh a trip . PARTICIPANT: a:nd they had …uh . PARTICIPANT: that's the family with their three children , two … …bo two boys and a girl , and whoop … and a baby up front too . PARTICIPANT: I didn't see that before , okay . PARTICIPANT: and in the back seat is grandma . PARTICIPANT: and grandma is …uh oh … not even as old as I am . PARTICIPANT: …um okay . PARTICIPANT: and then there's something on the side of the car here that I can't figure out what the heck that is . PARTICIPANT: oh it's a … …no oh okay so this is a Norman Rockwell painting . PARTICIPANT: aha that explains the …e error . PARTICIPANT: okay …um … well let's see . PARTICIPANT: okay everybody's excited because they're going on vacation . PARTICIPANT: and they got … it's gonna on the way to the … to …uh the shore or someplace with their whole family , and with grandma and …uh mother and father , and …uh the little baby or youngest one upfront , the …uh son , the daughter , and a dog . PARTICIPANT: and the dog is looking out the … …win the window while they're moving along and so_forth . PARTICIPANT: now …aft they went down to the beach , had a good day at the beach , and had a wonderful time running around and … with the dog and everything else . PARTICIPANT: and then they came back . PARTICIPANT: and this … the bottom picture is what's happening as they're driving home that evening . PARTICIPANT: and the … the kids are looking tired . PARTICIPANT: his somewhat red faced . PARTICIPANT: mom is sound asleep there but still with the kid on a thing . PARTICIPANT: papa is looking a_lot more beat too . PARTICIPANT: he's leaning forward with smokin(g) his cigar . PARTICIPANT: …uh well if … well if that's the case I wouldn't want to be in his car .

ChatGPT

Based on the evaluation rules provided, let's assess the PARTICIPANT's performance in the different aspects mentioned.

1. Vocabulary Features:

- Word Frequency: Counting the occurrences of each unique word is not possible in this dialogue format.
- Word Ratio: Similarly, calculating the word ratio is not feasible without the complete text.
- Phrase Frequency: Counting the occurrences of each unique phrase is not possible in this dialogue format.
- Phrase Ratio: Calculating the phrase ratio is also not feasible without the complete text.
- Word-Finding Difficulties: There are a few instances where the PARTICIPANT pauses or uses filler words like "uh" and "um." This could indicate some mild word-finding difficulties.
- Repetition: The PARTICIPANT repeats certain phrases, such as mentioning the tree multiple times. This suggests a tendency towards repetition.
- Lexical Complexity: Assessing the lexical complexity based on word length and frequency of rare words is not possible in this dialogue format.
- Limited Vocabulary: There is no significant evidence of a limited vocabulary in this dialogue.

2. Semantics Features:

- Semantic Density: Measuring the amount of meaningful information in the text is not feasible without the complete text.
- Inability to Convey Meaning: The PARTICIPANT expresses ideas and describes the scene, but there are instances where they struggle to convey meaning clearly, leading to some confusion or uncertainty.
- Semantic Coherence: The overall coherence and meaningfulness of the text are reasonable. The PARTICIPANT provides descriptions and attempts to form a coherent narrative based on the given picture.
- Lack of Coherence: While there are some disjointed thoughts and moments where the PARTICIPANT gets sidetracked, the overall flow of ideas remains relatively intact.

3. Syntax and Grammar Features:

- Syntactic Complexity: It's challenging to assess the complexity of sentence structures without the complete text.
- Simplified Sentence Structure: The sentences used by the PARTICIPANT are generally simple, with a lack of complex sentence structures.
- Grammatical Components: There are no obvious errors in terms of subject-verb agreement, tense consistency, pronoun usage, etc.
- Grammatical Errors: There are a few minor grammatical errors, such as missing verb endings (e.g., "ridin(g)"). However, the number of errors is not significant.

Based on the given dialogue, the PARTICIPANT shows some indications of mild cognitive impairment (MCI) in terms of word-finding difficulties, repetition, and occasional difficulty conveying meaning clearly. However, it's important to note that this is a preliminary assessment and a comprehensive judgment would require further evaluation.
