## Supplementary material for "Text dialogue analysis Based ChatGPT for Primary Screening of Mild Cognitive Impairment": Data(GPT4_vs_GPT3.5_model1-5): 100_GPT3.5 (3).docx

ChatGPT

Based on the analysis of the provided conversation, here are the evaluations of the indicators for mild cognitive impairment (MCI) for the participant:

Vocabulary features:

1. Limited vocabulary: 2
2. Increased verbosity: 3
3. Overlearned phrases: 3
4. Improper use of pronouns: 4
5. Hesitation (Word-finding difficulties): 6
6. Circumlocution (Word-finding difficulties): 5
7. Tip-of-the-tongue phenomenon: 6
8. Repetition: 3

Syntax and Grammar features:

1. Simplified sentence structure and grammar: 4
2. Grammatical errors: 3
3. Difficulty with complex ideas: 4

Semantics features:

1. Semantic paraphasias: 2
2. Lack of coherence: 3
3. Consistency in errors: 2
4. Lack of semantic fluency: 4
5. Disorganized narrative structure: 4
6. Orientation issues: 3
7. Decline in conversational engagement and responsiveness: 5
8. Memory issues: 4
9. Lack of abstract thinking and synthesis: 3
10. Abnormal emotions: 2

Based on these indicators, the participant shows some signs of mild cognitive impairment (MCI). However, it is important to note that not all indicators are present, and some of these issues can also be seen in individuals with normal cognitive function. A comprehensive evaluation should be conducted to make a conclusive determination.
