## Supplementary material for "Text dialogue analysis Based ChatGPT for Primary Screening of Mild Cognitive Impairment": Data(GPT4_vs_GPT3.5_model1-5): 100_GPT3.5 (4).docx

ChatGPT

Based on the language features exhibited by the participant, we can assess and rate the severity of each evaluation indicator:

Vocabulary features: a. Word frequency, word ratio, phrase frequency, and phrase ratio: Not enough information is provided to analyze these features.

b. Lexical complexity: Not enough information is provided to calculate lexical complexity.

c. Severity assessment:

- Limited vocabulary: 3
- Increased verbosity: 4
- Overlearned phrases: 2
- Improper use of pronouns: 2
- Hesitation: 2
- Circumlocution: 3
- Tip-of-the-tongue phenomenon: 2
- Repetition: 3

Syntax and grammar features: a. Syntactic complexity and grammatical components: The participant's sentences are generally coherent and grammatically correct, with occasional pauses and hesitations.

b. Severity assessment:

- Simplified sentence structure and grammar: 2
- Grammatical errors: 1
- Difficulty with complex ideas: 2

Semantics features: a. Semantic density and coherence: The participant's responses lack coherence at times, with some unrelated or unclear statements.

b. Severity assessment:

- Semantic paraphasias: Not observed
- Lack of coherence: 4
- Consistency in errors: 2
- Lack of semantic fluency: 3
- Disorganized narrative structure: 3
- Orientation issues: Not observed
- Decline in conversational engagement and responsiveness: Not observed
- Memory issues: Not observed
- Lack of abstract thinking and synthesis: 2
- Abnormal emotions: Not observed

Based on the assessment of the indicators, we can classify the participant into the Probably Mild Cognitive Impairment Group. The participant exhibits some language features associated with mild cognitive impairment, such as limited vocabulary, increased verbosity, overlearned phrases, improper use of pronouns, hesitation, circumlocution, tip-of-the-tongue phenomenon, repetition, lack of coherence, and some difficulty with abstract thinking and synthesis. It's important to note that this classification is based solely on the language features observed and does not provide a formal diagnosis. Further assessment and evaluation by a medical professional would be necessary to confirm the presence of mild cognitive impairment.
