## Supplementary material for "Text dialogue analysis Based ChatGPT for Primary Screening of Mild Cognitive Impairment": Data(GPT4_vs_GPT3.5_model1-5): 100_GPT3.5 (5).docx

ChatGPT

Based on the provided conversation, we will assess the participant's language features related to vocabulary, syntax and grammar, and semantics. We will rate the severity of each evaluation indicator on a scale of 1 to 10, with 1 being the least severe and 10 being the most severe.

Vocabulary Features: a. Word frequency and word ratio: The participant's word usage seems relatively simple, with limited diversity and frequency of unique words. They repeat certain words and phrases, such as "tree," "don't know," and "little girl." The ratio of unique words to the total number of words appears low. Additionally, they use pronouns more frequently than specific names or nouns. Rating: 7

b. Tip-of-the-tongue phenomenon: The participant exhibits instances of the tip-of-the-tongue phenomenon, where they struggle to recall specific words and rely on partial recall or general descriptions. For example, they remember certain characteristics or semantic categories of words but have difficulty retrieving the complete word. Rating: 6

Syntax and Grammar Features: a. Syntactic complexity and grammatical components: The participant's sentences generally follow a simple structure with straightforward clauses. They do not demonstrate a high level of syntactic complexity or sophisticated use of subordinate clauses. Grammatical components, such as subject-verb agreement, tense consistency, and proper use of pronouns, are generally correct. Rating: 4

b. Difficulty with complex ideas: The participant shows some difficulty when faced with more complex narratives or questions. They express uncertainty and struggle to provide detailed explanations or elaborate on certain aspects. Rating: 5

Semantics Features: a. Semantic density and coherence: The participant's speech lacks semantic density, with a limited use of content words (nouns, verbs, adjectives, adverbs) relative to function words (prepositions, pronouns, conjunctions). However, the text generally maintains coherence and cohesion, with logical relationships between words, phrases, and sentences. Rating: 6

b. Memory issues: The participant demonstrates some memory issues, including difficulty recalling specific information, sequences, and details from the provided conversation. They repeat questions and exhibit challenges in remembering plot points or characters. However, memory issues should be evaluated in the context of overall cognitive function. Rating: 7

Based on the comprehensive synthesis of the indicators, we can classify the participant into the Probably Mild Cognitive Impairment Group. The participant shows several language features commonly associated with mild cognitive impairment, such as limited vocabulary diversity, the tip-of-the-tongue phenomenon, difficulty with complex ideas, and memory issues. However, it is important to note that this evaluation is not a formal diagnosis, but rather an initial screening tool for potential MCI. A comprehensive assessment by a healthcare professional is necessary for an accurate diagnosis.
