## Supplementary material for "Text dialogue analysis Based ChatGPT for Primary Screening of Mild Cognitive Impairment": Data(GPT4_vs_GPT3.5_model1-5): 100_GPT3.5.docx

ChatGPT

Assessment of Vocabulary Features:

Word frequency: The participant's vocabulary appears to be limited, as they frequently repeat certain words such as "and," "uh," "um," "is," "the," and "okay." They also struggle to find the right words at times, resulting in pauses and the use of filler words.

Word ratio: It is difficult to calculate the word ratio without the exact word count of the text. However, based on the given conversation, the participant's use of unique words seems relatively low compared to the repetition of common words.

Phrase frequency: The participant tends to use simple and repetitive phrases throughout the conversation.

Phrase ratio: Without the total number of phrases in the text, it is challenging to determine the exact phrase ratio. However, similar to the word ratio, the participant's use of unique phrases appears to be limited.

Difficulty finding the right words: The participant frequently experiences difficulty finding the right words, resulting in pauses and the use of filler words such as "uh" and "um."

Lexical complexity: The participant's vocabulary lacks variety and sophistication. They rely on simple and commonly used words, with a limited range of more complex or rare vocabulary.

Limited vocabulary: The participant exhibits a limited vocabulary, characterized by repeated use of the same words and a lack of variety or sophistication in their word choice.

Assessment of Semantics Features:

Semantic density: The participant's speech lacks semantic density. They use a high proportion of non-content words (filler words, repetitions, etc.) compared to meaningful content words.

Inability to convey meaning: The participant struggles at times to express their ideas clearly, leading to confusion and potential misunderstanding.

Semantic coherence: The participant's responses generally exhibit logical consistency and meaningfulness. However, their use of filler words and occasional disjointed thoughts may impact the overall coherence of the conversation.

Lack of coherence: The participant occasionally struggles to maintain a logical flow of ideas, resulting in thoughts that appear disjointed or unrelated.

Assessment of Syntax and Grammar Features:

Syntactic complexity: The participant predominantly uses simple sentence structures with minimal use of complex or subordinate clauses. The sentences are relatively short in length.

Simplified sentence structure: The participant tends to use short, simple sentences, avoiding more complex sentence structures.

Grammatical components: The participant demonstrates some grammatical knowledge, using appropriate subject-verb agreement and pronouns. However, there are instances of incomplete or incorrect sentence structures.

Grammatical errors: The participant makes occasional grammatical errors, such as tense inconsistencies and incorrect verb forms.

Final Assessment:

Based on the conversation analysis, the participant shows signs of language difficulties and limitations in vocabulary, syntax, and grammar. They frequently struggle to find the right words, exhibit a limited vocabulary, and rely on simple sentence structures. Their speech lacks semantic density at times and may appear disjointed or unclear. Considering the language gaps observed, the participant is likely part of the Probably Mild Cognitive Impairment group. However, it is important to note that a comprehensive evaluation conducted by a healthcare professional is necessary for a definitive diagnosis
