## Supplementary material for "Text dialogue analysis Based ChatGPT for Primary Screening of Mild Cognitive Impairment": Data(GPT4_vs_GPT3.5_model1-5): 100_GPT4 (2).docx

INVESTIGATOR: so I'd just like you to tell me everything that you see going on in this picture . PARTICIPANT: I see that … the … their overflowing sink . PARTICIPANT: … …c the boy is …ge …s …gi stealing cookies from the cookie jar which is up in the cabinet . PARTICIPANT: the mother is standing there unaware of what's going on , busy …uh washing dishes or at least drying it , with something or other with the dish . PARTICIPANT: …uh outside they … it looks like they have some playground stuff outside . PARTICIPANT: I don't know what that thing is down there . PARTICIPANT: …um yeah …n and there's a tree outside . PARTICIPANT: and they got the curtains . PARTICIPANT: and let's see . PARTICIPANT: what else do we want to know ? PARTICIPANT: little girl's got …uh socks and shoes on . PARTICIPANT: the … so has the kid … the boy . PARTICIPANT: he's got shorts on . PARTICIPANT: …um yeah look … I … I see a face out there …uh in the ground … what looks like in the ground of the yard . PARTICIPANT: but I don't know whether that's sposta be a reflection off the glass or … or whether it's somebody out there in the yard , a reflection in the yard , or something they've made out_of the lawn . PARTICIPANT: I don't know . PARTICIPANT: …um there's some dishes on the counter . PARTICIPANT: let's see . PARTICIPANT: what else there would be xxx . PARTICIPANT: she has gotten wet feet , by the way , the lady . PARTICIPANT: …um I can't tell what's … what else that stuff is out . PARTICIPANT: there's a tree out there and I think I said that already . PARTICIPANT: and I don't know whether it looks like a jungle gym or what . PARTICIPANT: …um I don't see anything else . PARTICIPANT: well wait … wait a minute . PARTICIPANT: the girl's . PARTICIPANT: no . PARTICIPANT: no I … I guess that's her hair but I'm not sure whether …sh she's holding something or not . PARTICIPANT: can't tell . PARTICIPANT: she may be holding a dollar or something . PARTICIPANT: it's not clear . PARTICIPANT: the lady has …uh an apron on . PARTICIPANT: I don't know . PARTICIPANT: that's about it , I think . INVESTIGATOR: okay that works . INVESTIGATOR: so this picture tells a story . PARTICIPANT: yes it does . INVESTIGATOR: I want you to look at everything that's happening and then tell me a story about what you see . INVESTIGATOR: and the story should have a beginning a middle and an end . PARTICIPANT: oh_my_goodness . PARTICIPANT: okay well let's see . PARTICIPANT: the little girl was out with … on her bike . PARTICIPANT: and …um I don't know whether it's her cat or not . PARTICIPANT: but this dog came along and chased the cat up the tree . PARTICIPANT: and she wanted to get the … …ki the cat down . PARTICIPANT: so her dad , I assume to be her dad , or at least some guy , came along , got … went and got a ladder . PARTICIPANT: but once he got up in the tree …s some reason or another the ladder fell down . PARTICIPANT: and he didn't get the cat out_of the tree . PARTICIPANT: now he was stuck up in the tree too . PARTICIPANT: so somebody who saw what was going on . PARTICIPANT: or maybe it was the little bird in the tree that called the fire department . PARTICIPANT: and the fire department brought their truck . PARTICIPANT: and two firemen got out . PARTICIPANT: and they've got the ladder . PARTICIPANT: and hopefully they … they get both the man and the cat down and find out whose dog it is . INVESTIGATOR: so I want you to look at both of these pictures . INVESTIGATOR: …um and this whole thing tells a story . INVESTIGATOR: and so I'd like you to look at everything that's happening and tell me a story about what you see . INVESTIGATOR: and the story should have a beginning a middle and an end . PARTICIPANT: okay hm alright . PARTICIPANT: well I think what happened was that they , …um in the top picture , were ridin(g) along , going on …uh …uh a trip . PARTICIPANT: a:nd they had …uh . PARTICIPANT: that's the family with their three children , two … …bo two boys and a girl , and whoop … and a baby up front too . PARTICIPANT: I didn't see that before , okay . PARTICIPANT: and in the back seat is grandma . PARTICIPANT: and grandma is …uh oh … not even as old as I am . PARTICIPANT: …um okay . PARTICIPANT: and then there's something on the side of the car here that I can't figure out what the heck that is . PARTICIPANT: oh it's a … …no oh okay so this is a Norman Rockwell painting . PARTICIPANT: aha that explains the …e error . PARTICIPANT: okay …um … well let's see . PARTICIPANT: okay everybody's excited because they're going on vacation . PARTICIPANT: and they got … it's gonna on the way to the … to …uh the shore or someplace with their whole family , and with grandma and …uh mother and father , and …uh the little baby or youngest one upfront , the …uh son , the daughter , and a dog . PARTICIPANT: and the dog is looking out the … …win the window while they're moving along and so_forth . PARTICIPANT: now …aft they went down to the beach , had a good day at the beach , and had a wonderful time running around and … with the dog and everything else . PARTICIPANT: and then they came back . PARTICIPANT: and this … the bottom picture is what's happening as they're driving home that evening . PARTICIPANT: and the … the kids are looking tired . PARTICIPANT: his somewhat red faced . PARTICIPANT: mom is sound asleep there but still with the kid on a thing . PARTICIPANT: papa is looking a_lot more beat too . PARTICIPANT: he's leaning forward with smokin(g) his cigar . PARTICIPANT: …uh well if … well if that's the case I wouldn't want to be in his car . PARTICIPANT: …um and so_forth . PARTICIPANT: and all this is a painting by Norman Rockwell . PARTICIPANT: they got the … the boat on the top , by the way , is … is …uh . PARTICIPANT: the boat … they have a boat on the … …uh a rowboat on the top of the car and a beach umbrella . PARTICIPANT: so …uh they were going to the beach . PARTICIPANT: and then they came home dead tired because they were worn out from their whole day that they were hustling around at the beach . PARTICIPANT: the end . INVESTIGATOR: are you familiar with the story of Cinderella ? PARTICIPANT: yeah somewhat „ mhm . INVESTIGATOR: so I have two options . INVESTIGATOR: you can either tell me the story of Cinderella from what you remember or I can show you some pictures to jog your memory and then have you tell me the story . PARTICIPANT: ah let's see . PARTICIPANT: what do I remember about Cinderella ? PARTICIPANT: she had sisters . PARTICIPANT: …um and she was the one who …um ... PARTICIPANT: am I thinking right … of the right story ? PARTICIPANT: …um yeah that's a good question . PARTICIPANT: …um and she had three sisters . PARTICIPANT: and she …um ... PARTICIPANT: …um that … …uh that may not be Cinderella . PARTICIPANT: anyway she was busy . PARTICIPANT: …uh they kept … …uh she cleaned the house , did all kinds of things for the sisters and so_forth . PARTICIPANT: and then …uh the good fairy came along and set her up . PARTICIPANT: so she went to the ball . PARTICIPANT: is … is that right ? PARTICIPANT: I don't remember . PARTICIPANT: I haven't read that in .

ChatGPT

Based on the provided dialogue and the guidelines, let's evaluate the PARTICIPANT's language features:

1. Vocabulary:

- Word frequency and Word ratio: A detailed statistical analysis can't be conducted in this format, but anecdotally, the participant seems to have a fairly wide range of unique words.
- Phrase frequency and Phrase ratio: Similar to word frequency, a thorough analysis isn't possible here, but the participant uses various phrases to convey their thoughts.
- Word-finding difficulties: The participant often hesitates ("...uh", "...um") and has some difficulties finding the right words.
- Repetition: There's noticeable repetition of phrases (e.g., "I don't know" and "let's see") and ideas (e.g., mentioning the tree outside multiple times).
- Lexical complexity: The participant uses a mix of common and more complex words.
- Limited vocabulary: The participant does use some simple and common words frequently, but doesn't demonstrate a significantly reduced vocabulary.

2. Semantics:

- Semantic density: The participant uses a good number of content words relative to function words.
- Inability to convey meaning: The participant generally conveys their thoughts clearly, but there are instances of confusion or difficulty (e.g., forgetting the Cinderella story).
- Semantic coherence: The participant generally maintains semantic coherence, but there are moments of disconnection or disjointed thoughts.
- Lack of coherence: As mentioned, some moments of disconnection exist, especially when recalling the Cinderella story.

3. Syntax and Grammar:

- Syntactic complexity: The participant uses both simple and complex sentence structures.
- Simplified sentence structure: While the participant uses simple sentences often, there's still a fair use of complex sentences.
- Grammatical components: The participant mostly uses correct grammar, but there's occasional avoidance of complex grammatical constructions.
- Grammatical errors: There are very few grammatical errors.

Overall, the participant does show some features of Mild Cognitive Impairment (MCI), such as word-finding difficulties, some repetition, and moments of lack of coherence. However, they also demonstrate many elements of normal cognitive function, such as maintaining a good level of vocabulary and semantic density, relatively complex syntax, and proper use of grammar.

Based on this dialogue analysis, I would assign the participant to the "Probably Mild Cognitive Impairment" group due to the observable difficulties, but it's crucial to remember that a definitive diagnosis would require a comprehensive evaluation by healthcare professionals.
