## Supplementary material for "Text dialogue analysis Based ChatGPT for Primary Screening of Mild Cognitive Impairment": Data(GPT4_vs_GPT3.5_model1-5): 100_GPT4 (3).docx

ChatGPT

Given the extensive data provided, I'll attempt to provide a thorough analysis based on the stated aspects. It's important to remember that this is an initial screening, not a clinical diagnosis.

**Vocabulary Features Analysis:**

1. Limited Vocabulary: The participant does seem to have a reduced vocabulary, with a tendency to use simple words and phrases repetitively. Rating: 6/10
2. Increased Verbosity: The participant is quite verbose and sometimes uses more words than necessary. Rating: 7/10
3. Overlearned Phrases: There aren't many cliches or stock phrases observed. Rating: 3/10
4. Improper Use of Pronouns: The participant tends to use pronouns appropriately. Rating: 2/10
5. Hesitation: The participant shows signs of hesitation and word-finding difficulties, with noticeable pauses and use of filler words. Rating: 8/10
6. Circumlocution: The participant occasionally speaks in a roundabout way, especially when finding specific words or concepts challenging. Rating: 7/10
7. Tip-of-the-tongue phenomenon: The participant doesn't demonstrate this phenomenon explicitly in the given text. Rating: 2/10
8. Repetition: There is a noticeable amount of repetition in the participant's speech. Rating: 7/10

**Syntax and Grammar Features Analysis:**

1. Simplified Sentence Structure and Grammar: The participant prefers simpler sentence structures and doesn't use complex grammatical constructions often. Rating: 7/10
2. Grammatical Errors: There are minimal grammatical errors observed in the participant's speech. Rating: 2/10
3. Difficulty with Complex Ideas: The participant sometimes struggles to present complex ideas coherently. Rating: 5/10

**Semantics Features Analysis:**

1. Semantic Paraphasias: The participant occasionally has difficulty expressing ideas clearly, especially when the narrative becomes complex. Rating: 6/10
2. Lack of Coherence: The participant struggles to maintain coherence in their narratives. Rating: 7/10
3. Consistency in Errors: The participant seems consistent in certain language errors. Rating: 6/10
4. Lack of Semantic Fluency: The participant demonstrates some difficulty in naming items or describing sequences of events. Rating: 6/10
5. Disorganized Narrative Structure: The participant's narrative structure is somewhat disorganized, often lacking in key elements or coherence. Rating: 7/10
6. Orientation Issues: The participant does not demonstrate significant orientation issues in the given text. Rating: 2/10
7. Decline in Conversational Engagement and Responsiveness: The participant remains relatively engaged in the conversation, though at times they exhibit difficulty following or responding appropriately. Rating: 5/10
8. Memory Issues: The participant shows mild memory issues, such as repetition of certain phrases. Rating: 6/10
9. Lack of Abstract Thinking and Synthesis: The participant has some difficulty in understanding or expressing abstract concepts. Rating: 6/10
10. Abnormal Emotions: The participant doesn't exhibit noticeable abnormal emotions in the given text. Rating: 2/10

Based on the comprehensive synthesis of all the indicators, the participant may fall into the Probably Mild Cognitive Impairment Group. There are significant signs of hesitation, circumlocution, and repetition in their vocabulary features, along with a tendency to prefer simpler sentence structures. Semantically, they display some difficulty in expressing complex ideas clearly and maintaining coherence in their narratives. As a reminder, this evaluation is not a definitive diagnosis and any suspected cognitive impairment should be further assessed by a healthcare professional.
