## Supplementary material for "Text dialogue analysis Based ChatGPT for Primary Screening of Mild Cognitive Impairment": Data(GPT4_vs_GPT3.5_model1-5): 100_GPT4 (4).docx

ChatGPT

**Vocabulary features:** a. Word frequency is generally high, and phrase frequency is moderate. The participant uses many simple, commonly used words and phrases. b. Lexical complexity seems to be moderate, with a mix of simple and more complex words. c. Assessments:

- Limited vocabulary: 5/10 (Moderate)
- Increased verbosity: 4/10 (Moderate)
- Overlearned phrases: 3/10 (Low)
- Improper use of pronouns: 1/10 (Low)
- Hesitation: 7/10 (High)
- Circumlocution: 3/10 (Low)
- Tip-of-the-tongue phenomenon: 5/10 (Moderate)
- Repetition: 6/10 (Moderate)

**Syntax and grammar features:** a. Syntactic complexity and grammatical components are largely preserved, but some sentences show simplification. b. Assessments:

- Simplified sentence structure and grammar: 5/10 (Moderate)
- Grammatical errors: 2/10 (Low)
- Difficulty with complex ideas: 3/10 (Low)

**Semantics features:** a. Semantic density is generally high, but coherence sometimes breaks down with more complex descriptions. b. Assessments:

- Semantic paraphasias: 3/10 (Low)
- Lack of coherence: 6/10 (Moderate)
- Consistency in errors: 3/10 (Low)
- Lack of semantic fluency: 4/10 (Moderate)
- Disorganized narrative structure: 5/10 (Moderate)
- Orientation issues: 4/10 (Moderate)
- Decline in conversational engagement and responsiveness: 3/10 (Low)
- Memory issues: 7/10 (High)
- Lack of abstract thinking and synthesis: 5/10 (Moderate)
- Abnormal emotions: 1/10 (Low)

Based on the comprehensive synthesis of all the indicators, the participant would probably be classified as the **Probably Mild Cognitive Impairment Group** due to moderate-to-high levels of repetition, hesitation, limited vocabulary, simplified sentence structure, lack of coherence, and memory issues. However, further cognitive testing would be needed to make a definitive classification. The participant's ability to engage in conversation and respond to prompts remains generally good, but there are noticeable struggles with recall and narrative coherence. It's important to emphasize that this is not a diagnostic tool but an initial assessment based on language features.
