## Supplementary material for "Text dialogue analysis Based ChatGPT for Primary Screening of Mild Cognitive Impairment": Data(GPT4_vs_GPT3.5_model1-5): 100_GPT4 (5).docx

Please assess a participant with possible mild cognitive impairment (MCI) based on their language features. Note that I do not need to diagnose the participant but rather analyze and rate the severity of each evaluation indicator on a scale of 1 to 10 (1 being the least severe, and 10 being the most severe). Vocabulary features: a. Analyze word frequency, word ratio, phrase frequency, and phrase ratio. And calculate the lexical complexity. ( Word frequency and Word ratio: Count the occurrences of each unique word, each pronoun, and each specific name (or noun) in the text. And calculate the proportion of unique words to the total number of words in the text, and calculate the ratio of pronouns to specific names (or nouns). Phrase frequency and Phrase ratio: Count the occurrences of each unique phrases in the text. And calculate the proportion of unique phrases to the total number of phrases in the text. Lexical complexity: Measure the variety and sophistication of the vocabulary used, considering factors, including word length and frequency of rare words. ) b. Assess and rate the severity of the following aspects: tip-of-the-tongue phenomenon. (This phenomenon is characterized by partial recall. For instance, a person might remember the first letter of a word, its syllable structure, or its general semantic category, without being able to recall the whole word. More common in individuals with MCI.) Syntax and grammar features: a. Analyze syntactic complexity and grammatical components. (Syntactic complexity: Measure the complexity of the sentence structure in the text, considering factors such as the number of clauses per sentence, the average length of sentences, and the use of subordinate clauses. Grammatical components: Analyze the text for correct usage of grammar, such as subject-verb agreement, tense consistency, and proper use of pronouns, prepositions, and conjunctions.) b. Assess and rate the severity of the following aspects: difficulty with complex ideas (MCI: while the participant could handle simpler sentences and ideas, they had more difficulty when the narrative or the question became more complex.) Semantics features: a. Analyze semantic density and coherence. (Semantic density: Measure the amount of meaningful information in the text, and considering factor is the number of content words (nouns, verbs, adjectives, adverbs) relative to function words (prepositions, pronouns, conjunctions, etc.). Semantic coherence: Assess the logical consistency and meaningfulness of the text, which can be done by analyzing the relationships between words, phrases, and sentences, and checking for coherence and cohesion.) b. Assess and rate the severity of the following aspects: memory issues (MCI: repeat questions, difficulty in recalling certain information or sequences, difficulty in remembering specific details of the story, or difficulty in recalling major plot points or characters. However, memory issues can also be a part of normal aging, so this sign should be evaluated in the context of the individual's overall cognitive function.) Based on the comprehensive synthesis of all the indicators, classify the participant into either the Probably Normal Cognition Group or the Probably Mild Cognitive Impairment Group. Keep in mind that these issues might be present in individuals with normal cognition and that not all individuals with MCI will display all of these features. The final categorization should be based on a comprehensive synthesis of all the indicators. Please note: This evaluation isn't meant to provide a formal diagnosis but to serve as an initial screening tool for potential MCI. INVESTIGATOR: so I'd just like you to tell me everything that you see going on in this picture . PARTICIPANT: I see that … the … their overflowing sink . PARTICIPANT: … …c the boy is …ge …s …gi stealing cookies from the cookie jar which is up in the cabinet . PARTICIPANT: the mother is standing there unaware of what's going on , busy …uh washing dishes or at least drying it , with something or other with the dish . PARTICIPANT: …uh outside they … it looks like they have some playground stuff outside . PARTICIPANT: I don't know what that thing is down there . PARTICIPANT: …um yeah …n and there's a tree outside . PARTICIPANT: and they got the curtains . PARTICIPANT: and let's see . PARTICIPANT: what else do we want to know ? PARTICIPANT: little girl's got …uh socks and shoes on . PARTICIPANT: the … so has the kid … the boy . PARTICIPANT: he's got shorts on . PARTICIPANT: …um yeah look … I … I see a face out there …uh in the ground … what looks like in the ground of the yard . PARTICIPANT: but I don't know whether that's sposta be a reflection off the glass or … or whether it's somebody out there in the yard , a reflection in the yard , or something they've made out_of the lawn . PARTICIPANT: I don't know . PARTICIPANT: …um there's some dishes on the counter . PARTICIPANT: let's see . PARTICIPANT: what else there would be xxx . PARTICIPANT: she has gotten wet feet , by the way , the lady . PARTICIPANT: …um I can't tell what's … what else that stuff is out . PARTICIPANT: there's a tree out there and I think I said that already . PARTICIPANT: and I don't know whether it looks like a jungle gym or what . PARTICIPANT: …um I don't see anything else . PARTICIPANT: well wait … wait a minute . PARTICIPANT: the girl's . PARTICIPANT: no . PARTICIPANT: no I … I guess that's her hair but I'm not sure whether …sh she's holding something or not . PARTICIPANT: can't tell . PARTICIPANT: she may be holding a dollar or something . PARTICIPANT: it's not clear . PARTICIPANT: the lady has …uh an apron on . PARTICIPANT: I don't know . PARTICIPANT: that's about it , I think . INVESTIGATOR: okay that works . INVESTIGATOR: so this picture tells a story . PARTICIPANT: yes it does . INVESTIGATOR: I want you to look at everything that's happening and then tell me a story about what you see . INVESTIGATOR: and the story should have a beginning a middle and an end . PARTICIPANT: oh_my_goodness . PARTICIPANT: okay well let's see . PARTICIPANT: the little girl was out with … on her bike . PARTICIPANT: and …um I don't know whether it's her cat or not . PARTICIPANT: but this dog came along and chased the cat up the tree . PARTICIPANT: and she wanted to get the … …ki the cat down . PARTICIPANT: so her dad , I assume to be her dad , or at least some guy , came along , got … went and got a ladder . PARTICIPANT: but once he got up in the tree …s some reason or another the ladder fell down . PARTICIPANT: and he didn't get the cat out_of the tree . PARTICIPANT: now he was stuck up in the tree too . PARTICIPANT: so somebody who saw what was going on . PARTICIPANT: or maybe it was the little bird in the tree that called the fire department . PARTICIPANT: and the fire department brought their truck . PARTICIPANT: and two firemen got out . PARTICIPANT: and they've got the ladder . PARTICIPANT: and hopefully they … they get both the man and the cat down and find out whose dog it is . INVESTIGATOR: so I want you to look at both of these pictures . INVESTIGATOR: …um and this whole thing tells a story . INVESTIGATOR: and so I'd like you to look at everything that's happening and tell me a story about what you see . INVESTIGATOR: and the story should have a beginning a middle and an end . PARTICIPANT: okay hm alright . PARTICIPANT: well I think what happened was that they , …um in the top picture , were ridin(g) along , going on …uh …uh a trip . PARTICIPANT: a:nd they had …uh . PARTICIPANT: that's the family with their three children , two … …bo two boys and a girl , and whoop … and a baby up front too . PARTICIPANT: I didn't see that before , okay . PARTICIPANT: and in the back seat is grandma . PARTICIPANT: and grandma is …uh oh … not even as old as I am . PARTICIPANT: …um okay . PARTICIPANT: and then there's something on the side of the car here that I can't figure out what the heck that is . PARTICIPANT: oh it's a … …no oh okay so this is a Norman Rockwell painting . PARTICIPANT: aha that explains the …e error . PARTICIPANT: okay …um … well let's see . PARTICIPANT: okay everybody's excited because they're going on vacation . PARTICIPANT: and they got … it's gonna on the way to the … to …uh the shore or someplace with their whole family , and with grandma and …uh mother and father , and …uh the little baby or youngest one upfront , the …uh son , the daughter , and a dog . PARTICIPANT: and the dog is looking out the … …win the window while they're moving along and so_forth . PARTICIPANT: now …aft they went down to the beach , had a good day at the beach , and had a wonderful time running around and … with the dog and everything else . PARTICIPANT: and then they came back . PARTICIPANT: and this … the bottom picture is what's happening as they're driving home that evening . PARTICIPANT: and the … the kids are looking tired . PARTICIPANT: his somewhat red faced . PARTICIPANT: mom is sound asleep there but still with the kid on a thing . PARTICIPANT: papa is looking a_lot more beat too . PARTICIPANT: he's leaning forward with smokin(g) his cigar . PARTICIPANT: …uh well if … well if that's the case I wouldn't want to be in his car . PARTICIPANT: …um and so_forth . PARTICIPANT: and all this is a painting by Norman Rockwell . PARTICIPANT: they got the … the boat on the top , by the way , is … is …uh . PARTICIPANT: the boat … they have a boat on the … …uh a rowboat on the top of the car and a beach umbrella . PARTICIPANT: so …uh they were going to the beach . PARTICIPANT: and then they came home dead tired because they were worn out from their whole day that they were hustling around at the beach . PARTICIPANT: the end . INVESTIGATOR: are you familiar with the story of Cinderella ? PARTICIPANT: yeah somewhat „ mhm . INVESTIGATOR: so I have two options . INVESTIGATOR: you can either tell me the story of Cinderella from what you remember or I can show you some pictures to jog your memory and then have you tell me the story . PARTICIPANT: ah let's see . PARTICIPANT: what do I remember about Cinderella ? PARTICIPANT: she had sisters . PARTICIPANT: …um and she was the one who …um ... PARTICIPANT: am I thinking right … of the right story ? PARTICIPANT: …um yeah that's a good question . PARTICIPANT: …um and she had three sisters . PARTICIPANT: and she …um ... PARTICIPANT: …um that … …uh that may not be Cinderella . PARTICIPANT: anyway she was busy . PARTICIPANT: …uh they kept … …uh she cleaned the house , did all kinds of things for the sisters and so_forth . PARTICIPANT: and then …uh the good fairy came along and set her up . PARTICIPANT: so she went to the ball . PARTICIPANT: is … is that right ? PARTICIPANT: I don't remember . PARTICIPANT: I haven't read that in . INVESTIGATOR: you wanna see some pictures and then maybe you can &PARTICIPANT:yeah_go_ahead tell me more ? INVESTIGATOR: alright so . INVESTIGATOR: yeah so I'll show you all the pictures and then I'll have you kinda add to your story . PARTICIPANT: okay . PARTICIPANT: okay that's the … the nasty ones , the sisters . PARTICIPANT: …uh yeah they're all gonna go to the ball . PARTICIPANT: and she's busy . PARTICIPANT: see okay I did have the right story . PARTICIPANT: anyway but she's …uh yeah . PARTICIPANT: …um then okay that's ... PARTICIPANT: yeah she … the good fairy comes along and brings her a … gets her … sets her up in a dress and so_forth . PARTICIPANT: she goes . PARTICIPANT: this … there's the good fairy „ okay . PARTICIPANT: …um just goes in the carriage that's …um a pumpkin . PARTICIPANT: and goes to the ball . PARTICIPANT: she meets the Prince Charming or whoever he is . PARTICIPANT: and …uh she stays . PARTICIPANT: but she's gotta leave by midnight . PARTICIPANT: and she has … the clock rings …t . PARTICIPANT: and she hasta run . PARTICIPANT: she gets home . PARTICIPANT: and she's happy telling about the mice and everybody about what happened . PARTICIPANT: an:d the sisters don't understand it because she lost her shoe . PARTICIPANT: and the prince went looking for her . PARTICIPANT: and they went around to … to …every test everybody's feet and whether they would fit the shoe . PARTICIPANT: and only Cinderella …wis got her foot into that glass slipper . PARTICIPANT: and so they lived happily ever after . INVESTIGATOR: I'd like you to tell me how to make a peanut butter and jelly sandwich . PARTICIPANT: ah yes often do it . INVESTIGATOR: okay . PARTICIPANT: well the first thing I always do is I go to the …uh refrigerator . PARTICIPANT: and we have a drawer that has the bread and butter … the bread in it . PARTICIPANT: so I go out and get … take out the bread . PARTICIPANT: I take out the butter . PARTICIPANT: and I take out the jelly , which your … your butter is on the top shelf . PARTICIPANT: and the bread is in the lower shelf . PARTICIPANT: and the jelly is in this other door . PARTICIPANT: okay . PARTICIPANT: I take (th)em out . PARTICIPANT: I unscrew the top on the jelly . PARTICIPANT: and …uh I …uh unscrew the top on the peanut butter . PARTICIPANT: and then I smear peanut butter on the … …s on the two pieces of bread . PARTICIPANT: and then I put the jelly on top of the peanut butter . PARTICIPANT: then I put the second piece of bread on top of that . PARTICIPANT: and I have a sandwich .

ChatGPT

In order to make a robust analysis based on your provided instructions, I'll start by assessing each category separately (vocabulary, syntax and grammar, and semantics), then combine these to make a final estimation. Please note, this kind of evaluation is better conducted using a large dataset. Using this one example might not give a completely accurate picture of the participant's language abilities.

1. Vocabulary: a. Word frequency and word ratio: The participant uses a diverse set of words and there is a good ratio between unique words and total number of words used (8/10). Pronoun to specific noun ratio is moderate (5/10). b. Phrase frequency and phrase ratio: There is considerable repetition of phrases, suggesting some difficulty in phrase formation (5/10). c. Lexical complexity: The participant uses common, everyday language, with less use of complex or rare words (5/10). d. Tip-of-the-tongue phenomenon: The participant exhibits some difficulty in finding the right word at times, but is mostly able to eventually express his thoughts (6/10).
2. Syntax and Grammar: a. Syntactic complexity: The sentences vary in length, with several shorter sentences and fewer complex or long ones (5/10). b. Grammatical components: The participant generally uses correct grammar, with occasional lapses in subject-verb agreement and tense consistency (7/10). c. Difficulty with complex ideas: The participant sometimes struggles to articulate complex ideas or events but is able to get across the basic premise of the story (6/10).
3. Semantics: a. Semantic density: The text has a fairly high semantic density, with a good proportion of content words to function words (7/10). b. Semantic coherence: The participant mostly maintains coherence in his narratives, but there are instances where he loses track or repeats certain details (6/10). c. Memory issues: The participant does show signs of forgetting details of the story, repeats questions, and has difficulty recalling specific details or sequences (5/10).

In conclusion, based on this one instance, the participant displays some signs that could suggest possible mild cognitive impairment. However, it is important to note that these are fairly common and could be part of normal aging as well. Given the overall evaluation scores and taking into account the normal variability in language use among older adults, the participant would probably fit better into the Probably Mild Cognitive Impairment Group (MCI). However, this isn't a formal diagnosis but an initial screening
