## Supplementary material for "Text dialogue analysis Based ChatGPT for Primary Screening of Mild Cognitive Impairment": Data(GPT4_vs_GPT3.5_model1-5): 100_GPT4.docx

ChatGPT

2 / 2

Assessment of PARTICIPANT's language features:

Vocabulary features:

- Word frequency: 335 unique words
- Word ratio: 0.43 (335 unique words / 778 total words)
- Phrase frequency: Not applicable for this analysis
- Phrase ratio: Not applicable for this analysis
- Difficulty finding the right words: Frequent pauses and use of filler words like "uh" and "um"
- Lexical complexity: Moderate, with a mix of simple and more sophisticated words
- Limited vocabulary: Some repetition, but an overall variety of words

Semantics features:

- Semantic density: Moderate, with a balance of content and function words
- Inability to convey meaning: Occasional difficulty, but generally clear expression of ideas
- Semantic coherence: Mostly coherent, with some occasional disjointed thoughts
- Lack of coherence: Occasional difficulty maintaining a logical flow of ideas

Syntax and Grammar features:

- Syntactic complexity: Moderate complexity, with a mix of simple and more complex sentence structures
- Simplified sentence structure: Some simple sentences, but also more complex ones
- Grammatical components: Generally correct grammar, with occasional errors
- Grammatical errors: A few mistakes, but not frequent

Based on this analysis, I would assign PARTICIPANT to the Probably Normal group. While there are some signs of difficulty in finding the right words and occasional lack of coherence, the overall language features suggest normal cognitive functioning.
