## Supplementary material for "Text dialogue analysis Based ChatGPT for Primary Screening of Mild Cognitive Impairment": Data(GPT4_vs_GPT3.5_model1-5): 101_GPT3.5 (2).docx

Attention! You do not need to diagnose, just follow the steps below to initially screen for PARTICIPANT with possibly mild cognitive impairment (MCI). Some differences between MCI and normal cognition are also given, please analyse them in detail, step by step. Firstly, please assess the Vocabulary features of PARTICIPANT at the Word frequency and Word ratio, Phrase frequency and Phrase ratio, Word-finding difficulties, Repetition, Lexical complexity, Limited vocabulary. Word frequency: Count the occurrences of each unique word in the text. Unique words are distinct words that appear in the text, not taking into account their repetitions. Each word is counted only once, regardless of how many times it occurs. Word ratio: Calculate the proportion of unique words to the total number of words in the text. Phrase frequency: Count the occurrences of each unique phrases in the text. Unique phrases are distinct combinations of words that appear in the text, not taking into account their repetitions. Phrase ratio: Calculate the proportion of unique phrases to the total number of phrases in the text. Word-finding difficulties: Frequent pauses or use of filler words, like "uh" or "um,". MCI: more frequent instances, longer time to find the right word, or using more general terms. Normal: fewer instances, and better lexical retrieval. Repetition: The participant repeats certain phrases or ideas a few times throughout the conversation. MCI: more frequent repetition. Normal: less frequent repetition (some repetition of words but not excessive) and better memory of what has been mentioned. Lexical complexity: Measure the variety and sophistication of the vocabulary used, considering factors, including word length and frequency of rare words. Limited vocabulary: Repeated use of the same words or simple words. MCI: slightly reduced vocabulary, and more frequent use of simpler words or common terms; Normal: richer vocabulary (low limitation). Secondly, please assess the Semantics features of PARTICIPANT, from Semantic density and Inability to convey meaning, Semantic coherence and Lack of coherence. Semantic density: Measure the amount of meaningful information in the text, and considering factor is the number of content words (nouns, verbs, adjectives, adverbs) relative to function words (prepositions, pronouns, conjunctions, etc.). Inability to convey meaning: Difficulty in expressing ideas clearly, leading to confusion or misunderstanding. Normal: the participant was able to describe the scene clearly. Semantic coherence: Assess the logical consistency and meaningfulness of the text, which can be done by analyzing the relationships between words, phrases, and sentences, and checking for coherence and cohesion. Lack of coherence: Difficulty in maintaining a logical flow of ideas, with thoughts appearing disjointed or unrelated. MCI: struggle with organizing thoughts coherently or maintaining a clear focus in conversation; Normal: greater coherence and organization in their storytelling and conversations. Thirdly, please assess the Syntax and Grammar features of PARTICIPANT, from Syntactic complexity and Simplified sentence structure, Grammatical components and Grammatical errors. Syntactic complexity: Measure the complexity of the sentence structure in the text, considering factors such as the number of clauses per sentence, the average length of sentences, and the use of subordinate clauses. Simplified sentence structure: A lack of complex sentence structures. MCI: simpler sentence structures; Normal: in longer dialogues there is a greater variety of sentence structures (moderately complex), but in short dialogues the sentences may turn out to be simple. Grammatical components: Analyze the text for correct usage of grammar, such as subject-verb agreement, tense consistency, and proper use of pronouns, prepositions, and conjunctions. MCI: avoidance of complex grammatical constructions; Normal: better use of complex grammatical constructions. Grammatical errors: Frequent mistakes in verb tense, subject-verb agreement, or other aspects of grammar. MCI: more frequent grammatical errors, such as mistakes in verb tense and subject-verb agreement; Normal: fewer grammatical errors (a few errors, but not significant). Finally, Please dichotomise PARTICIPANT into the Probably Normal group or the Probably Mild Cognitive Impairment group. PARTICIPANT with MCI might exhibit more difficulty with vocabulary, sentence structure, repetition, word-finding, coherence, and grammar. However, it is not definitive and requires a comprehensive judgement. Now, you do not have to respond to this dialogue. When I have entered THE DIALOGUE BETWEEN PARTICIPANT AND INVESTIGATOR, evaluate PARTICIPANT according to the evaluation rules above. INVESTIGATOR: here's the picture . PARTICIPANT: okay . PARTICIPANT: …hm let me see . PARTICIPANT: the boy is getting cookies out_of the cookie jar . PARTICIPANT: and the stool is just about to fall over . PARTICIPANT: the little girl is reaching up for a cookie . PARTICIPANT: and the mother is drying dishes . PARTICIPANT: the water is running in to the sink and the sink is running over PARTICIPANT: you want action ? INVESTIGATOR: mhm . PARTICIPANT: you want all the action there is ? PARTICIPANT: anyhow I think that's all the action there is . PARTICIPANT: and that little girl is laughing . PARTICIPANT: that little girl xxx . PARTICIPANT: did I say the mother was drying dishes ? PARTICIPANT: drying a dish . INVESTIGATOR: okay . PARTICIPANT: okay . PARTICIPANT: water running out_of the sink . PARTICIPANT: lady dryin(g) a plate . PARTICIPANT: an(d) a child getting cookies out_of the cookie jar standing on a PARTICIPANT: an(d) a little girl reaching up for a cookie . PARTICIPANT: and …um curtains at the window . PARTICIPANT: a path outside . PARTICIPANT: a tree . PARTICIPANT: …w a window into another room . PARTICIPANT: shrubbery . PARTICIPANT: grass . PARTICIPANT: let's get back to the room again . PARTICIPANT: there's a cup … two cups and a plate sitting on the countertop PARTICIPANT: cupboards . PARTICIPANT: the lady has a apron on, dress, slippers . PARTICIPANT: and the boy has on a short sleeve shirt, short pants, socks, and PARTICIPANT: little girl has on a dress, socks, strap slippers . PARTICIPANT: did I mention everything ? PARTICIPANT: sink . PARTICIPANT: faucet with water running out_of it . PARTICIPANT: an open window behind the … the sink . PARTICIPANT: tie-back curtains . PARTICIPANT: and that's all I see . INVESTIGATOR: okay great .

ChatGPT

4 / 4

Based on the provided dialogue, let's evaluate the participant according to the given evaluation rules.

1. Vocabulary Features:

- Word frequency: Count the occurrences of each unique word in the text. Unique words are distinct words that appear in the text, not taking into account their repetitions. Each word is counted only once, regardless of how many times it occurs.
- Word ratio: Calculate the proportion of unique words to the total number of words in the text.
- Phrase frequency: Count the occurrences of each unique phrase in the text. Unique phrases are distinct combinations of words that appear in the text, not taking into account their repetitions.
- Phrase ratio: Calculate the proportion of unique phrases to the total number of phrases in the text.
- Word-finding difficulties: Frequent pauses or use of filler words, like "uh" or "um." MCI: more frequent instances, longer time to find the right word, or using more general terms. Normal: fewer instances and better lexical retrieval.
- Repetition: The participant repeats certain phrases or ideas a few times throughout the conversation. MCI: more frequent repetition. Normal: less frequent repetition (some repetition of words but not excessive) and better memory of what has been mentioned.
- Lexical complexity: Measure the variety and sophistication of the vocabulary used, considering factors including word length and frequency of rare words.
- Limited vocabulary: Repeated use of the same words or simple words. MCI: slightly reduced vocabulary and more frequent use of simpler words or common terms; Normal: richer vocabulary (low limitation).

Based on the provided dialogue, here's the analysis of the vocabulary features:

Word frequency and Word ratio: We need the full text to perform word frequency and calculate the word ratio. Please provide the complete dialogue.

Phrase frequency and Phrase ratio: Similarly, we require the complete dialogue to count unique phrases and calculate the phrase ratio.

Word-finding difficulties: In the given dialogue, the participant does not exhibit significant word-finding difficulties. They express their thoughts without excessive pauses or use of filler words. Therefore, there are no clear indications of word-finding difficulties associated with MCI.

Repetition: The participant repeats some phrases and ideas throughout the conversation, such as mentioning the action in the picture and asking if they mentioned certain details. This repetition is relatively minor and does not seem excessive. They also demonstrate memory of what has been mentioned. Therefore, the level of repetition is within the range of normal conversation and does not strongly indicate MCI.

Lexical complexity: Without a complete analysis of the participant's vocabulary, including word length and frequency of rare words, it is challenging to assess their lexical complexity.

Limited vocabulary: Based on the provided dialogue, there is no significant evidence of a limited vocabulary. The participant uses a variety of words and provides descriptions of objects and actions in the picture. Their vocabulary appears to be within a normal range.

2. Semantics Features:

- Semantic density: Measure the amount of meaningful information in the text, considering factors such as the number of content words (nouns, verbs, adjectives, adverbs) relative to function words (prepositions, pronouns, conjunctions, etc.).
- Inability to convey meaning: Difficulty in expressing ideas clearly, leading to confusion or misunderstanding. Normal: the participant was able to describe the scene clearly.
- Semantic coherence: Assess the logical consistency and meaningfulness of the text, analyzing the relationships between words, phrases, and sentences and checking for coherence and cohesion.
- Lack of coherence: Difficulty in maintaining a logical flow of ideas, with thoughts appearing disjointed or unrelated. MCI: struggle with organizing thoughts coherently or maintaining a clear focus in conversation; Normal: greater coherence and organization in their storytelling and conversations.

Based on the provided dialogue, here's the analysis of the semantics features:

Semantic density: Without the complete dialogue, it's challenging to determine the semantic density based on the content words and function words ratio.

Inability to convey meaning: The participant demonstrates an ability to describe the scene and provide information about the people, objects, and actions in the picture. They convey their ideas clearly without significant difficulty, indicating a normal ability to express meaning.

Semantic coherence and Lack of coherence: In this dialogue, the participant maintains a logical flow of ideas and demonstrates coherence in their storytelling. There are no major signs of disjointed or unrelated thoughts. The participant's ability to describe the picture and provide details appears to be within a normal range.

3. Syntax and Grammar Features:

- Syntactic complexity: Measure the complexity of the sentence structure in the text, considering factors such as the number of clauses per sentence, the average length of sentences, and the use of subordinate clauses.
- Simplified sentence structure: A lack of complex sentence structures. MCI: simpler sentence structures; Normal: in longer dialogues, there is a greater variety of sentence structures (moderately complex), but in short dialogues, the sentences may turn out to be simple.
- Grammatical components: Analyze the text for correct usage of grammar, such as subject-verb agreement, tense consistency, and proper use of pronouns, prepositions, and conjunctions.
- Grammatical errors: Frequent mistakes in verb tense, subject-verb agreement, or other aspects of grammar. MCI: more frequent grammatical errors, such as mistakes in verb tense and subject-verb agreement; Normal: fewer grammatical errors (a few errors, but not significant).

Based on the provided dialogue, here's the analysis of the syntax and grammar features:

Syntactic complexity and Simplified sentence structure: The sentences used by the participant in this dialogue are relatively short and straightforward. They do not exhibit complex sentence structures or the use of subordinate clauses. Therefore, the sentence structure appears to be on the simpler side, which could be considered within the range of normal for a short dialogue.

Grammatical components and Grammatical errors: There are no significant grammatical errors or noticeable mistakes in verb tense, subject-verb agreement, or other grammatical aspects in the provided dialogue. The participant's usage of grammar seems to be generally correct.

Based on the analysis of the provided dialogue, it does not strongly indicate mild cognitive impairment (MCI). The participant shows normal or near-normal performance in the assessed features, including vocabulary, semantics, syntax, and grammar. However, please note that this is only an initial screening, and a comprehensive judgment would require further evaluation.
