## Supplementary material for "Text dialogue analysis Based ChatGPT for Primary Screening of Mild Cognitive Impairment": Data(GPT4_vs_GPT3.5_model1-5): 101_GPT3.5 (3).docx

ChatGPT

Based on the provided text, here is an analysis of the indicators and evaluation of their severity on a scale of 1 to 10 for the PARTICIPANT:

Vocabulary features:

- Limited vocabulary: 3/10 (MCI): There is some repetition of words, such as "cookie," "little girl," and "drying dishes."
- Increased verbosity: 2/10 (MCI): The PARTICIPANT tends to provide more information than necessary, such as saying "you want all the action there is?" when asked for action.
- Overlearned phrases: 1/10 (Normal): There is no evidence of overusing cliches or stock phrases.
- Improper use of pronouns: 1/10 (Normal): Pronoun usage is appropriate in the text.
- Hesitation (Word-finding difficulties): 1/10 (Normal): There are no frequent pauses or filler words in the text.
- Circumlocution (Word-finding difficulties): 2/10 (MCI): The PARTICIPANT uses more words than necessary and describes the scene in a roundabout way, mentioning various details before providing the intended answer.
- Tip-of-the-tongue phenomenon: Not applicable based on the given text.
- Repetition: 1/10 (Normal): There is minimal repetition of words or phrases.

Syntax and Grammar features:

- Simplified sentence structure and grammar: 2/10 (MCI): The PARTICIPANT tends to use simpler sentence structures and avoids complex grammatical constructions.
- Grammatical errors: 1/10 (Normal): There are no significant grammatical errors in the text.
- Difficulty with complex ideas: 1/10 (Normal): The text does not involve complex ideas.

Semantics features:

- Semantic paraphasias: 1/10 (Normal): The PARTICIPANT expresses ideas clearly without errors in word choice.
- Lack of coherence: 2/10 (MCI): The PARTICIPANT struggles with organizing thoughts coherently and changes topics frequently.
- Consistency in errors: 1/10 (Normal): There are no consistent language errors.
- Lack of semantic fluency: 1/10 (Normal): The PARTICIPANT does not exhibit difficulty in naming items or describing processes.
- Disorganized narrative structure: 3/10 (MCI): The PARTICIPANT struggles to maintain a coherent line of thought and jumps around chronologically.
- Orientation issues: Not applicable based on the given text.
- Decline in conversational engagement and responsiveness: 2/10 (MCI): The PARTICIPANT exhibits decreased initiation, difficulties in following the conversation, and steering back to familiar topics.
- Memory issues: 2/10 (MCI): The PARTICIPANT repeats questions and struggles to recall specific details.
- Lack of abstract thinking and synthesis: 2/10 (MCI): The PARTICIPANT has difficulty in synthesizing the overall situation and understanding abstract concepts.
- Abnormal emotions: 1/10 (Normal): There are no evident abnormal emotions in the text.

Based on the analysis of these indicators, it is likely that the PARTICIPANT falls into the Probably Mild Cognitive Impairment Group. The presence of limited vocabulary, increased verbosity, circumlocution, disorganized narrative structure, memory issues, and difficulties with abstract thinking and synthesis are suggestive of mild cognitive impairment. However, further evaluation and assessment by a qualified healthcare professional are necessary for a definitive diagnosis.
