## Supplementary material for "Text dialogue analysis Based ChatGPT for Primary Screening of Mild Cognitive Impairment": Data(GPT4_vs_GPT3.5_model1-5): 101_GPT3.5 (4).docx

ChatGPT

Vocabulary features: a. Word frequency, word ratio, phrase frequency, and phrase ratio:

- It is difficult to assess word frequency, word ratio, phrase frequency, and phrase ratio without a larger sample of the participant's language. Based on the provided conversation, it is not possible to make an accurate evaluation.

b. Lexical complexity:

- Lexical complexity cannot be accurately assessed based on the provided conversation.

c. Evaluation of severity:

- Limited vocabulary: The participant demonstrates a relatively limited vocabulary, using simple and repetitive language. Severity rating: 6
- Increased verbosity: The participant tends to repeat and overexplain certain aspects, leading to verbosity. Severity rating: 5
- Overlearned phrases: There is no evidence of overlearned phrases in the conversation.
- Improper use of pronouns: There is no evidence of improper use of pronouns in the conversation.
- Hesitation: The participant shows some hesitation and pauses while speaking. Severity rating: 4
- Circumlocution: There is no evidence of significant circumlocution in the conversation.
- Tip-of-the-tongue phenomenon: There is no evidence of the tip-of-the-tongue phenomenon in the conversation.
- Repetition: The participant repeats certain phrases and information. Severity rating: 5

Syntax and grammar features: a. Syntactic complexity and grammatical components:

- It is difficult to assess syntactic complexity and grammatical components without a larger sample of the participant's language. Based on the provided conversation, it is not possible to make an accurate evaluation.

b. Evaluation of severity:

- Simplified sentence structure and grammar: The participant predominantly uses simple sentence structures and grammar. Severity rating: 7
- Grammatical errors: No noticeable grammatical errors were observed in the conversation.
- Difficulty with complex ideas: There is no evidence of difficulty with complex ideas in the conversation.

Semantics features: a. Semantic density and coherence:

- It is difficult to assess semantic density and coherence without a larger sample of the participant's language. Based on the provided conversation, it is not possible to make an accurate evaluation.

b. Evaluation of severity:

- Semantic paraphasias: There is no evidence of semantic paraphasias in the conversation.
- Lack of coherence: The participant's narrative lacks coherence and jumps between unrelated topics. Severity rating: 8
- Consistency in errors: No consistent errors were observed in the conversation.
- Lack of semantic fluency: The participant demonstrates some difficulty in expressing their thoughts fluently. Severity rating: 4
- Disorganized narrative structure: The participant's narrative structure lacks organization and cohesion. Severity rating: 7
- Orientation issues: There is no evidence of orientation issues in the conversation.
- Decline in conversational engagement and responsiveness: The participant appears engaged and responsive during the conversation.
- Memory issues: There is no direct evidence of memory issues in the conversation.
- Lack of abstract thinking and synthesis: There is no evidence of a lack of abstract thinking and synthesis in the conversation.
- Abnormal emotions: There is no evidence of abnormal emotions in the conversation.

Based on the limited sample of conversation and the assessment of the language features, the participant can be tentatively classified into the Probably Mild Cognitive Impairment Group. However, it is important to note that a comprehensive evaluation involving other cognitive domains and additional language samples would be necessary for a more accurate assessment. It is also crucial to consider that some of the observed language features might be present in individuals with normal cognition as well. Therefore, further evaluation by a qualified healthcare professional is essential for a definitive diagnosis.
