## Supplementary material for "Text dialogue analysis Based ChatGPT for Primary Screening of Mild Cognitive Impairment": Data(GPT4_vs_GPT3.5_model1-5): 101_GPT3.5 (5).docx

ChatGPT

Let's assess the participant based on the provided language features:

Vocabulary features: a. Word frequency and word ratio: Count the occurrences of each unique word, pronoun, and specific name. Calculate the proportion of unique words to the total number of words in the text and the ratio of pronouns to specific names.

In the given text, the participant uses a limited vocabulary, with repetition of certain phrases such as "getting cookies out of the cookie jar" and "drying dishes." The proportion of unique words to the total number of words is relatively low. The participant uses pronouns ("the boy," "the little girl," "the mother") and specific names ("cookie jar," "stool," "sink") but not in a consistent ratio. The word frequency and word ratio indicate a moderate level of vocabulary complexity.

b. Tip-of-the-tongue phenomenon:

The participant does not exhibit any clear signs of the tip-of-the-tongue phenomenon in the provided text.

Syntax and grammar features: a. Syntactic complexity and grammatical components:

The participant's sentences are relatively simple, with straightforward structures and a low number of clauses. There are no significant issues with subject-verb agreement, tense consistency, or the use of pronouns, prepositions, and conjunctions. The syntactic complexity and grammatical components appear to be within the normal range.

b. Difficulty with complex ideas:

The participant shows no apparent difficulty with complex ideas in the given text.

Semantics features: a. Semantic density and coherence:

The participant's use of content words (nouns, verbs, adjectives, adverbs) relative to function words (prepositions, pronouns, conjunctions) is balanced, indicating moderate semantic density. The text demonstrates some logical consistency and meaningfulness, although there are instances of incomplete sentences and fragmented descriptions. Overall, the semantic coherence is moderate.

b. Memory issues:

The participant does not exhibit significant memory issues in the provided text.

Based on the assessment of the language features, here is the classification:

The participant can be classified into the Probably Normal Cognition Group. While there are some indicators of potential mild cognitive impairment, such as moderate vocabulary complexity and moderate semantic coherence, the absence of severe issues in other aspects suggests that the participant's language performance falls within the normal range. However, it's important to note that this classification is based solely on the language features and should be interpreted in conjunction with other cognitive assessments for a comprehensive evaluation of cognitive function.
