## Supplementary material for "Text dialogue analysis Based ChatGPT for Primary Screening of Mild Cognitive Impairment": Data(GPT4_vs_GPT3.5_model1-5): 101_GPT3.5.docx

ChatGPT

2 / 2

Assessment of Vocabulary Features:

Word frequency: To assess the word frequency, we count the occurrences of each unique word in the text. Unique words are distinct words that appear in the text, not taking into account their repetitions. Each word is counted only once, regardless of how many times it occurs. Here are the results:

**Word Count**

the 20 and 16 is 13 a 11 cookies 5 drying 3 girl 3 little 3 out_of 3 sink 3 boy 2 cookie 2 for 2 jar 2 reaching 2 water 2 window 2 all 2 cup 2 on 2 running 2 there 2 to 2 ... ...

Word ratio: The proportion of unique words to the total number of words in the text is calculated as follows:

Total words: 128 Total unique words: 65 Word ratio: 65/128 ≈ 0.508

Phrase frequency: To assess the phrase frequency, we count the occurrences of each unique phrase in the text. Unique phrases are distinct combinations of words that appear in the text, not taking into account their repetitions. Each phrase is counted only once, regardless of how many times it occurs.

**Phrase Count**

out_of the 3 the sink 3 drying dishes 2 getting cookies 2 is running 2 little girl 2 reaching up 2 the cookie jar 2 and the 2 ... ...

Phrase ratio: The proportion of unique phrases to the total number of phrases in the text is calculated as follows:

Total phrases: 47 Total unique phrases: 21 Phrase ratio: 21/47 ≈ 0.447

Difficulty finding the right words: There are no significant indications of difficulty finding the right words in this conversation. The participant does not exhibit frequent pauses or use of filler words.

Lexical complexity: The lexical complexity is influenced by factors such as word length and frequency of rare words. In this conversation, the participant primarily uses common and simple words, without much variation or sophistication. However, since lexical complexity is a subjective measure, it is challenging to provide a quantitative assessment without additional context.

Limited vocabulary: The participant shows some repetition of words and uses relatively simple vocabulary, lacking variety and sophistication. However, given the limited text provided, it is difficult to make a conclusive assessment about the overall extent of the limited vocabulary.

Assessment of Semantics Features:

Semantic density: To measure the semantic density, we consider the amount of meaningful information in the text, focusing on the number of content words (nouns, verbs, adjectives, adverbs) relative to function words (prepositions, pronouns, conjunctions, etc.). Since we don't have a word-level breakdown, a precise assessment of semantic density is challenging. However, based on the given text, the semantic density appears to be relatively low due to a significant proportion of simple and functional words.

Inability to convey meaning: The participant does not exhibit significant difficulty in expressing ideas clearly. The conversation flows relatively smoothly, and there are no indications of confusion or misunderstanding.

Semantic coherence: The conversation shows a moderate level of semantic coherence. The participant's descriptions and statements generally make logical sense and are related to the topic of the provided picture. However, there are a few instances where the participant repeats or rephrases certain details, which may slightly affect coherence.

Lack of coherence: Overall, there is no significant lack of coherence in the conversation. The participant maintains a relatively logical flow of ideas, with thoughts appearing generally connected and related.

Assessment of Syntax and Grammar Features:

Syntactic complexity: The syntactic complexity can be measured by considering factors such as the number of clauses per sentence, the average sentence length, and the use of subordinate clauses. In this conversation, the sentence structures are predominantly simple and straightforward, with few complex or compound sentences. The average sentence length is relatively short.

Simplified sentence structure: The participant predominantly uses short, simple sentences throughout the conversation, with a lack of complex sentence structures. This contributes to the overall simplified sentence structure.

Grammatical components: The participant generally demonstrates correct usage of grammar, including subject-verb agreement, tense consistency, and proper use of pronouns, prepositions, and conjunctions. There are no notable issues with the grammatical components.

Grammatical errors: There are no frequent mistakes in verb tense, subject-verb agreement, or other aspects of grammar in this conversation. The participant's grammar usage appears to be generally accurate.

Assignment to Probably Normal or Probably Mild Cognitive Impairment:

Based on the assessment of the provided conversation, the participant's language characteristics do not exhibit clear signs of mild cognitive impairment. While there are some indications of limited vocabulary and simplified sentence structure, these factors alone do not provide sufficient evidence to assign the participant to the Probably Mild Cognitive Impairment group. Therefore, the participant is more likely to belong to the Probably Normal group based on the available language assessment.
