## Supplementary material for "Text dialogue analysis Based ChatGPT for Primary Screening of Mild Cognitive Impairment": Data(GPT4_vs_GPT3.5_model1-5): 101_GPT4 (2).docx

INVESTIGATOR: here's the picture . PARTICIPANT: okay . PARTICIPANT: …hm let me see . PARTICIPANT: the boy is getting cookies out_of the cookie jar . PARTICIPANT: and the stool is just about to fall over . PARTICIPANT: the little girl is reaching up for a cookie . PARTICIPANT: and the mother is drying dishes . PARTICIPANT: the water is running in to the sink and the sink is running over PARTICIPANT: you want action ? INVESTIGATOR: mhm . PARTICIPANT: you want all the action there is ? PARTICIPANT: anyhow I think that's all the action there is . PARTICIPANT: and that little girl is laughing . PARTICIPANT: that little girl xxx . PARTICIPANT: did I say the mother was drying dishes ? PARTICIPANT: drying a dish . INVESTIGATOR: okay . PARTICIPANT: okay . PARTICIPANT: water running out_of the sink . PARTICIPANT: lady dryin(g) a plate . PARTICIPANT: an(d) a child getting cookies out_of the cookie jar standing on a PARTICIPANT: an(d) a little girl reaching up for a cookie . PARTICIPANT: and …um curtains at the window . PARTICIPANT: a path outside . PARTICIPANT: a tree . PARTICIPANT: …w a window into another room . PARTICIPANT: shrubbery . PARTICIPANT: grass . PARTICIPANT: let's get back to the room again . PARTICIPANT: there's a cup … two cups and a plate sitting on the countertop PARTICIPANT: cupboards . PARTICIPANT: the lady has a apron on, dress, slippers . PARTICIPANT: and the boy has on a short sleeve shirt, short pants, socks, and PARTICIPANT: little girl has on a dress, socks, strap slippers . PARTICIPANT: did I mention everything ? PARTICIPANT: sink . PARTICIPANT: faucet with water running out_of it . PARTICIPANT: an open window behind the … the sink . PARTICIPANT: tie-back curtains . PARTICIPANT: and that's all I see . INVESTIGATOR: okay great .

ChatGPT

Vocabulary features:

- Word frequency: 77 unique words
- Word ratio: 77/133 = 0.58
- Phrase frequency: Not applicable in this context
- Phrase ratio: Not applicable in this context
- Word-finding difficulties: Few instances of "um" and "hm", some repetition
- Repetition: Moderate repetition observed
- Lexical complexity: Moderate
- Limited vocabulary: No significant limitation observed

Semantics features:

- Semantic density: Good content word usage relative to function words
- Inability to convey meaning: The participant was able to describe the scene clearly
- Semantic coherence: The participant's description is coherent and logically connected
- Lack of coherence: No significant lack of coherence observed

Syntax and Grammar features:

- Syntactic complexity: Moderate complexity in sentence structures
- Simplified sentence structure: The sentences are mostly simple
- Grammatical components: Proper use of grammatical components
- Grammatical errors: No significant errors observed

Based on this evaluation, I would assign the PARTICIPANT to the Probably Normal group. The participant demonstrates a good understanding of the picture and can describe it in detail. Their sentences are grammatically correct, and they maintain coherence throughout their response. The minor instances of word-finding difficulties and repetition do not significantly impact the overall clarity of their description
