## Supplementary material for "Text dialogue analysis Based ChatGPT for Primary Screening of Mild Cognitive Impairment": Data(GPT4_vs_GPT3.5_model1-5): 101_GPT4 (3).docx

ChatGPT

Here's the evaluation based on your criteria:

1. **Vocabulary Features:**
   - Word frequency and Word ratio: The participant has a rich vocabulary and uses a variety of words, showing no limitation (1/10). The pronoun to specific noun ratio appears normal (2/10).
   - Phrase frequency and Phrase ratio: The participant uses diverse phrases, with no overuse of cliches or stock phrases (2/10).
   - Lexical complexity: The participant uses simple words mostly, but also some complex words and phrases (4/10).
   - Increased verbosity: The participant is slightly verbose (4/10).
   - Overlearned phrases: The participant does not overuse cliches or stock phrases (1/10).
   - Improper use of pronouns: The participant uses pronouns correctly (1/10).
   - Hesitation: The participant shows occasional hesitation, but not excessive (3/10).
   - Circumlocution: The participant describes the scene in detail, not using too many words unnecessarily (3/10).
   - Tip-of-the-tongue phenomenon: The participant does not show clear signs of this (2/10).
   - Repetition: The participant does repeat some information, but not excessively (3/10).
2. **Syntax and Grammar Features:**
   - Syntactic complexity: The participant uses a variety of sentence structures, indicating normal complexity (2/10).
   - Grammatical components: The participant uses grammar correctly, with no notable errors (1/10).
   - Simplified sentence structure and grammar: The participant does not overly simplify sentences or grammar (1/10).
   - Grammatical errors: The participant does not make grammatical errors (1/10).
   - Difficulty with complex ideas: The participant seems to handle complex ideas adequately (2/10).
3. **Semantics Features:**
   - Semantic density: The participant provides a dense and detailed description of the scene, indicating good semantic density (2/10).
   - Semantic coherence: The participant maintains coherence throughout the dialogue (2/10).
   - Semantic paraphasias: The participant does not demonstrate semantic paraphasias (1/10).
   - Lack of coherence: The participant's narrative is generally coherent (2/10).
   - Consistency in errors: The participant does not demonstrate consistent errors (1/10).
   - Lack of semantic fluency: The participant does not show lack of semantic fluency (1/10).
   - Disorganized narrative structure: The participant's narrative is organized (2/10).
   - Orientation Issues: The participant does not demonstrate orientation issues (1/10).
   - Decline in conversational engagement and responsiveness: The participant remains engaged and responsive throughout the dialogue (2/10).
   - Memory issues: The participant repeats some information, but not significantly enough to indicate memory issues (3/10).
   - Lack of abstract thinking and synthesis: The participant synthesizes the overall situation effectively (2/10).
   - Abnormal emotions: The participant does not display any abnormal emotions (1/10).

Based on these evaluations, the participant would be categorized into the **Probably Normal Cognition Group**. While some mild issues (slight verbosity, occasional repetition, and slight hesitations) are observed, these are not pronounced or persistent enough to suggest MCI, especially considering the overall coherence, variety, and complexity of the participant's narrative.
