## Supplementary material for "Text dialogue analysis Based ChatGPT for Primary Screening of Mild Cognitive Impairment": Data(GPT4_vs_GPT3.5_model1-5): 101_GPT4 (4).docx

ChatGPT

**Vocabulary Features**

a. The participant demonstrated good word and phrase frequency. They used a diverse range of words and phrases to describe the scene, and the ratios are within the normal range.

b. The lexical complexity is moderate. There's a good mix of basic and more specific terms.

c. Severity assessment:

- Limited Vocabulary: 2/10 (The participant uses a decent range of vocabulary to describe the scene)
- Increased Verbosity: 5/10 (The participant goes into a fair amount of detail, possibly more than necessary)
- Overlearned Phrases: 2/10 (The participant doesn't seem to rely heavily on common or clichéd phrases)
- Improper Use of Pronouns: 1/10 (The participant correctly uses pronouns)
- Hesitation: 4/10 (Some hesitation is noticed, but it's not severe)
- Circumlocution: 4/10 (The participant occasionally goes around the point before reaching it)
- Tip-of-the-Tongue Phenomenon: 3/10 (Slightly noticeable but not pronounced)
- Repetition: 5/10 (There are several instances where the participant repeats themselves, especially when confirming previous descriptions)

**Syntax and Grammar Features**

a. Syntactic complexity is moderate, with the participant forming complete and complex sentences.

b. Severity assessment:

- Simplified Sentence Structure and Grammar: 3/10 (The participant is able to formulate complex sentences)
- Grammatical Errors: 2/10 (Grammatical mistakes are rare)
- Difficulty with Complex Ideas: 3/10 (The participant seems to understand and relay complex ideas)

**Semantics Features**

a. Semantic density and coherence are decent. The participant's descriptions of the scene are clear and logically connected.

b. Severity assessment:

- Semantic Paraphasias: 1/10 (There are no noticeable instances of incorrect word use)
- Lack of Coherence: 2/10 (The participant's responses generally make sense and follow a logical order)
- Consistency in Errors: 2/10 (Errors are not frequent and don't follow a consistent pattern)
- Lack of Semantic Fluency: 3/10 (The participant generally speaks fluently, with a few hesitations)
- Disorganized Narrative Structure: 3/10 (The participant's descriptions are generally organized)
- Orientation Issues: 1/10 (The participant seems to be oriented to the situation)
- Decline in Conversational Engagement and Responsiveness: 2/10 (The participant stays engaged and responsive)
- Memory Issues: 3/10 (A few instances of repetition suggest potential mild memory issues)
- Lack of Abstract Thinking and Synthesis: 2/10 (The participant doesn't display significant issues with abstract thinking or synthesis)
- Abnormal Emotions: 1/10 (The participant doesn't display any abnormal emotions)

Based on this analysis, I would suggest the participant is in the **Probably Normal Cognition Group**. However, slight issues, such as repetition and increased verbosity, are worth keeping an eye on. Remember that this evaluation is not a definitive diagnosis. Any significant concerns should be discussed with a healthcare provider.
