## Supplementary material for "Text dialogue analysis Based ChatGPT for Primary Screening of Mild Cognitive Impairment": Data(GPT4_vs_GPT3.5_model1-5): 101_GPT4 (5).docx

ChatGPT

Vocabulary features: a. The participant used a wide range of unique words, pronouns, and specific names. Their word frequency and word ratio is generally high, suggesting good lexical diversity and a balance between nouns and pronouns. Their phrase frequency and phrase ratio suggest a moderate variety of different phrases used. Overall, their lexical complexity is rather high, indicating a robust and sophisticated vocabulary. I would rate the severity of issues here as 2/10. b. The participant does not appear to experience the tip-of-the-tongue phenomenon severely, as they generally seem to recall specific words without noticeable struggle. I would rate the severity of issues here as 1/10.

Syntax and grammar features: a. The syntactic complexity of the participant's language is moderate. They mostly use simple sentences, but they also use some more complex structures. Their grammatical components are largely correct, with good use of subject-verb agreement, tense consistency, and proper use of pronouns, prepositions, and conjunctions. I would rate the severity of issues here as 2/10. b. The participant seems to handle complex ideas relatively well, although there are moments when they repeat information or struggle to recall something they have previously stated. This suggests some minor difficulty with complex ideas. I would rate the severity of issues here as 3/10.

Semantics features: a. The participant's semantic density is high, as they use a significant number of content words relative to function words. Their semantic coherence is also good, as the text is logically consistent and meaningful, with clear relationships between words, phrases, and sentences. I would rate the severity of issues here as 1/10. b. The participant does display some memory issues, such as repeating questions and difficulty recalling certain details or sequences (for example, the participant asked, "did I say the mother was drying dishes?"). However, this could also be a part of normal aging. I would rate the severity of issues here as 4/10.

Based on the comprehensive synthesis of all the indicators, I would classify the participant into the Probably Normal Cognition Group. The participant displayed minor issues with handling complex ideas and some potential memory issues, but their vocabulary, syntax, grammar, and semantic features are generally strong. It's important to note that these issues might be present in individuals with normal cognition, and not all individuals with MCI will display all of these features. Also, further assessment would be needed for a formal diagnosis
