## Supplementary material for "Text dialogue analysis Based ChatGPT for Primary Screening of Mild Cognitive Impairment": Data(GPT4_vs_GPT3.5_model1-5): 101_GPT4.docx

ChatGPT

After analyzing the conversation between the PARTICIPANT and INVESTIGATOR, I have assessed the following:

Vocabulary features:

- Word frequency: 86 unique words
- Word ratio: 86/179 = 0.48
- Phrase frequency: 46 unique phrases
- Phrase ratio: 46/63 = 0.73
- Difficulty finding the right words: Minimal difficulty; only a few instances of filler words and pauses.
- Lexical complexity: Moderate complexity with some variety in vocabulary.
- Limited vocabulary: Some repetition of words, but not excessive.

Semantics features:

- Semantic density: Moderate density; a balance of content words and function words.
- Inability to convey meaning: Minimal difficulty; the participant generally conveys the scene clearly.
- Semantic coherence: The text appears coherent and meaningful.
- Lack of coherence: Minimal; the participant mostly maintains a logical flow of ideas.

Syntax and Grammar features:

- Syntactic complexity: Moderate complexity; a mix of simple and compound sentences.
- Simplified sentence structure: Some simple sentences, but not overly simplified.
- Grammatical components: Generally correct grammar usage with only a few errors.
- Grammatical errors: A few instances of errors, but not frequent.

Based on the language analysis, I would assign the PARTICIPANT to the Probably Normal group. Keep in mind, however, that this is only an initial screening based on language usage and not a definitive diagnosis. A proper evaluation should be conducted by a healthcare professional.
