## Supplementary material for "Text dialogue analysis Based ChatGPT for Primary Screening of Mild Cognitive Impairment": Data(GPT4_vs_GPT3.5_model1-5): 102_GPT3.5 (2).docx

Attention! You do not need to diagnose, just follow the steps below to initially screen for PARTICIPANT with possibly mild cognitive impairment (MCI). Some differences between MCI and normal cognition are also given, please analyse them in detail, step by step. Firstly, please assess the Vocabulary features of PARTICIPANT at the Word frequency and Word ratio, Phrase frequency and Phrase ratio, Word-finding difficulties, Repetition, Lexical complexity, Limited vocabulary. Word frequency: Count the occurrences of each unique word in the text. Unique words are distinct words that appear in the text, not taking into account their repetitions. Each word is counted only once, regardless of how many times it occurs. Word ratio: Calculate the proportion of unique words to the total number of words in the text. Phrase frequency: Count the occurrences of each unique phrases in the text. Unique phrases are distinct combinations of words that appear in the text, not taking into account their repetitions. Phrase ratio: Calculate the proportion of unique phrases to the total number of phrases in the text. Word-finding difficulties: Frequent pauses or use of filler words, like "uh" or "um,". MCI: more frequent instances, longer time to find the right word, or using more general terms. Normal: fewer instances, and better lexical retrieval. Repetition: The participant repeats certain phrases or ideas a few times throughout the conversation. MCI: more frequent repetition. Normal: less frequent repetition (some repetition of words but not excessive) and better memory of what has been mentioned. Lexical complexity: Measure the variety and sophistication of the vocabulary used, considering factors, including word length and frequency of rare words. Limited vocabulary: Repeated use of the same words or simple words. MCI: slightly reduced vocabulary, and more frequent use of simpler words or common terms; Normal: richer vocabulary (low limitation). Secondly, please assess the Semantics features of PARTICIPANT, from Semantic density and Inability to convey meaning, Semantic coherence and Lack of coherence. Semantic density: Measure the amount of meaningful information in the text, and considering factor is the number of content words (nouns, verbs, adjectives, adverbs) relative to function words (prepositions, pronouns, conjunctions, etc.). Inability to convey meaning: Difficulty in expressing ideas clearly, leading to confusion or misunderstanding. Normal: the participant was able to describe the scene clearly. Semantic coherence: Assess the logical consistency and meaningfulness of the text, which can be done by analyzing the relationships between words, phrases, and sentences, and checking for coherence and cohesion. Lack of coherence: Difficulty in maintaining a logical flow of ideas, with thoughts appearing disjointed or unrelated. MCI: struggle with organizing thoughts coherently or maintaining a clear focus in conversation; Normal: greater coherence and organization in their storytelling and conversations. Thirdly, please assess the Syntax and Grammar features of PARTICIPANT, from Syntactic complexity and Simplified sentence structure, Grammatical components and Grammatical errors. Syntactic complexity: Measure the complexity of the sentence structure in the text, considering factors such as the number of clauses per sentence, the average length of sentences, and the use of subordinate clauses. Simplified sentence structure: A lack of complex sentence structures. MCI: simpler sentence structures; Normal: in longer dialogues there is a greater variety of sentence structures (moderately complex), but in short dialogues the sentences may turn out to be simple. Grammatical components: Analyze the text for correct usage of grammar, such as subject-verb agreement, tense consistency, and proper use of pronouns, prepositions, and conjunctions. MCI: avoidance of complex grammatical constructions; Normal: better use of complex grammatical constructions. Grammatical errors: Frequent mistakes in verb tense, subject-verb agreement, or other aspects of grammar. MCI: more frequent grammatical errors, such as mistakes in verb tense and subject-verb agreement; Normal: fewer grammatical errors (a few errors, but not significant). Finally, Please dichotomise PARTICIPANT into the Probably Normal group or the Probably Mild Cognitive Impairment group. PARTICIPANT with MCI might exhibit more difficulty with vocabulary, sentence structure, repetition, word-finding, coherence, and grammar. However, it is not definitive and requires a comprehensive judgement. Now, you do not have to respond to this dialogue. When I have entered THE DIALOGUE BETWEEN PARTICIPANT AND INVESTIGATOR, evaluate PARTICIPANT according to the evaluation rules above.| INVESTIGATOR: okay . PARTICIPANT: okay . INVESTIGATOR: what do you see goin(g) on there ? PARTICIPANT: the woman fouled up and left the spigot running and then it's running onto the floor . PARTICIPANT: …uh the kid's reachin(g) in for cookies which is correct but he's …d …uh …uh …sl falling off the stool . PARTICIPANT: …uh she's standing in water . PARTICIPANT: …hm I'd say that's about it . INVESTIGATOR: anything else ? PARTICIPANT: anything else . PARTICIPANT: I don't see nothin(g) else . PARTICIPANT: right now (there)'s snow outside . PARTICIPANT: she's got the window open naturally . PARTICIPANT: no I don't see nothing else . INVESTIGATOR: okay . PARTICIPANT: that's out of the … …or out_of the ordinary . INVESTIGATOR: okay . PARTICIPANT: right . INVESTIGATOR: alright . INVESTIGATOR: going on, that you see happening . PARTICIPANT: okay . PARTICIPANT: the sink's running over . PARTICIPANT: the kid's on a stool and is falling over . PARTICIPANT: …mm she's standin(g) in the water . PARTICIPANT: …um I'd say that's about it . PARTICIPANT: oh_no there's … there's somethin(g) ... PARTICIPANT: no … no it ain't … it ain't writing upside down here . PARTICIPANT: it looks like it . PARTICIPANT: …uh no that's … that's all . INVESTIGATOR: okay . INVESTIGATOR: on to the picture . PARTICIPANT: the boy's … …uh the girl's makin(g) fun of the boy . PARTICIPANT: she made fun of him so much while he was stealin(g) a cookie out o(f) the cookie jar . PARTICIPANT: and it made him trip . PARTICIPANT: and he's going to fall . PARTICIPANT: the mother's listening to the kids . PARTICIPANT: she's drying dishes . PARTICIPANT: she got the window open against a mirror . PARTICIPANT: she's isn't payin(g) attention to the sink . PARTICIPANT: the sink's spillin(g) water onto the floor . PARTICIPANT: it's gettin(g) her feet wet . PARTICIPANT: the curtains are open to allow …uh fresh air to come in the house . PARTICIPANT: there's two …uh cups on the … …st a dish on the sink . PARTICIPANT: and the faucets are open and the cupboard door (i)s open . PARTICIPANT: …uh looks like it might be …su spring or summer out because things look like they're growin(g) outside . PARTICIPANT: …uh she has shoes on . PARTICIPANT: she has shoes on . PARTICIPANT: he has shoes on . PARTICIPANT: she has shoes on . PARTICIPANT: she has a dress, dress, apron, shirt, shorts, socks, socks . PARTICIPANT: …uh the lid (i)s off the cookie jar . PARTICIPANT: there's …uh …s a fancy sink cabinet over here and a fancy wall cabinet over there . PARTICIPANT: that's about it . INVESTIGATOR: okay good .

ChatGPT

2 / 2

Based on the provided dialogue between the participant and the investigator, let's analyze the participant's performance according to the evaluation rules mentioned earlier.

1. Vocabulary features:

- Word frequency: Count the occurrences of each unique word in the text. Since the complete text is not provided, it's not possible to calculate word frequency accurately.
- Word ratio: Calculate the proportion of unique words to the total number of words in the text. Since the complete text is not provided, it's not possible to calculate word ratio accurately.
- Phrase frequency: Count the occurrences of each unique phrase in the text. Since the complete text is not provided, it's not possible to calculate phrase frequency accurately.
- Phrase ratio: Calculate the proportion of unique phrases to the total number of phrases in the text. Since the complete text is not provided, it's not possible to calculate phrase ratio accurately.
- Word-finding difficulties: The participant exhibits some word-finding difficulties, such as frequent pauses and the use of filler words like "uh" and "um." This could indicate mild cognitive impairment (MCI).
- Repetition: The participant repeats certain phrases or ideas multiple times, which suggests some difficulty with memory or retrieval. This could be indicative of MCI.
- Lexical complexity: It's challenging to assess lexical complexity based on the given dialogue. The participant's use of vocabulary seems to be simple and repetitive, which may suggest limited vocabulary.

2. Semantics features:

- Semantic density: It's not possible to accurately measure semantic density without the complete text. The number of content words relative to function words cannot be determined.
- Inability to convey meaning: The participant does not demonstrate significant difficulty in expressing ideas clearly in the given dialogue.
- Semantic coherence: The participant's responses seem to be logically consistent and meaningful, maintaining a reasonable level of coherence in the conversation.

3. Syntax and Grammar features:

- Syntactic complexity: The complexity of sentence structures is limited due to the relatively short and straightforward nature of the dialogue. It's difficult to assess the participant's syntactic complexity without a more extensive sample.
- Simplified sentence structure: The participant tends to use simpler sentence structures in the given dialogue, which could be indicative of MCI.
- Grammatical components: Based on the given dialogue, it's not possible to assess the participant's usage of complex grammatical constructions accurately.
- Grammatical errors: No significant grammatical errors are apparent in the participant's responses.

Considering the observations made above, it appears that the participant shows some signs that align with mild cognitive impairment (MCI). These signs include word-finding difficulties, repetition, limited vocabulary, simplified sentence structures, and potentially reduced lexical complexity. However, a comprehensive judgment would require a more extensive evaluation and assessment.
