## Supplementary material for "Text dialogue analysis Based ChatGPT for Primary Screening of Mild Cognitive Impairment": Data(GPT4_vs_GPT3.5_model1-5): 102_GPT3.5 (3).docx

ChatGPT

Based on the provided conversation, here is an analysis of the indicators and evaluation of their severity for the participant:

Vocabulary features:

- Word frequency and Word ratio: The participant uses a limited vocabulary with a tendency to repeat words. The severity of limited vocabulary is 6 out of 10.
- Phrase frequency and Phrase ratio: The participant uses a few unique phrases, but the proportion of unique phrases to the total number of phrases is low. The severity of limited phrase variety is 4 out of 10.
- Lexical complexity: The participant's vocabulary consists of simple words and common terms. The severity of lexical complexity reduction is 7 out of 10.

Syntax and Grammar features:

- Syntactic complexity: The participant's sentence structures are mostly simple, with a preference for shorter sentences. The severity of simplified sentence structure is 5 out of 10.
- Grammatical components: The participant makes a few grammatical errors, such as subject-verb agreement errors. The severity of grammatical errors is 3 out of 10.
- Difficulty with complex ideas: The participant has some difficulty expressing and understanding complex ideas. The severity of difficulty with complex ideas is 6 out of 10.

Semantics features:

- Semantic density: The participant's text contains a reasonable amount of meaningful information. The severity of semantic density reduction is 4 out of 10.
- Semantic coherence: The participant's responses show some logical consistency and meaningfulness. The severity of lack of coherence is 3 out of 10.
- Semantic paraphasias: The participant doesn't exhibit clear semantic paraphasias. The severity of semantic paraphasias is 1 out of 10.
- Lack of coherence: The participant struggles to maintain a clear focus and frequently changes topics. The severity of lack of coherence is 7 out of 10.
- Consistency in errors: There are no consistent language errors observed. The severity of consistency in errors is 1 out of 10.
- Lack of semantic fluency: The participant has some difficulty in describing and naming specific items or processes. The severity of lack of semantic fluency is 5 out of 10.
- Disorganized narrative structure: The participant's narrative lacks a clear structure and jumps around chronologically. The severity of disorganized narrative structure is 7 out of 10.
- Orientation issues: There is no evidence of orientation issues. The severity of orientation issues is 1 out of 10.
- Decline in conversational engagement and responsiveness: The participant shows a decreased level of engagement and responsiveness, often steering conversations back to familiar topics or ignoring/responding inappropriately. The severity of decline in conversational engagement and responsiveness is 8 out of 10.
- Memory issues: There are no significant memory issues observed in the conversation. The severity of memory issues is 2 out of 10.
- Lack of abstract thinking and synthesis: The participant struggles to synthesize the overall situation and abstract concepts. The severity of lack of abstract thinking and synthesis is 6 out of 10.
- Abnormal emotions: No abnormal emotions are evident in the conversation. The severity of abnormal emotions is 1 out of 10.

Based on the comprehensive synthesis of these indicators, the participant's cognitive impairment is likely indicative of mild cognitive impairment (MCI).
