## Supplementary material for "Text dialogue analysis Based ChatGPT for Primary Screening of Mild Cognitive Impairment": Data(GPT4_vs_GPT3.5_model1-5): 102_GPT3.5 (4).docx

ChatGPT

Let's assess the participant based on the provided language features.

Vocabulary features: a. Word frequency, word ratio, phrase frequency, and phrase ratio: Based on the provided conversation, it is difficult to analyze word and phrase frequency accurately. b. Lexical complexity: It is challenging to determine the lexical complexity without a larger sample of the participant's language.

Severity rating of vocabulary features:

1. Limited vocabulary: Not evident in the given conversation.
2. Increased verbosity: Mild (3) - Some instances of unnecessary repetition and wordiness.
3. Overlearned phrases: Not evident in the given conversation.
4. Improper use of pronouns: Mild (3) - Occasional pronoun confusion.
5. Hesitation: Moderate (5) - Frequent hesitations observed.
6. Circumlocution: Mild (4) - Some instances of using longer phrases to express simple ideas.
7. Tip-of-the-tongue phenomenon: Not evident in the given conversation.
8. Repetition: Mild (3) - Occasional repetition of words and phrases.

Syntax and grammar features: a. Syntactic complexity and grammatical components: The participant generally uses simple sentence structures and shows occasional grammatical errors.

Severity rating of syntax and grammar features:

1. Simplified sentence structure and grammar: Mild (4) - Mostly simple sentence structures with occasional complexity.
2. Grammatical errors: Mild (3) - Occasional grammatical errors.
3. Difficulty with complex ideas: Not evident in the given conversation.

Semantics features: a. Semantic density and coherence: The participant's responses contain relevant information but lack organization and coherence at times.

Severity rating of semantics features:

1. Semantic paraphasias: Not evident in the given conversation.
2. Lack of coherence: Mild (4) - Some lack of organization and coherence observed.
3. Consistency in errors: Not evident in the given conversation.
4. Lack of semantic fluency: Not evident in the given conversation.
5. Disorganized narrative structure: Mild (4) - Some lack of organization in the responses.
6. Orientation issues: Not evident in the given conversation.
7. Decline in conversational engagement and responsiveness: Not evident in the given conversation.
8. Memory issues: Not evident in the given conversation.
9. Lack of abstract thinking and synthesis: Not evident in the given conversation.
10. Abnormal emotions: Not evident in the given conversation.

Based on the comprehensive synthesis of the indicators, the participant can be classified into the Probably Normal Cognition Group. However, it is important to note that this evaluation is based on a limited sample and should not be considered a formal diagnosis. A comprehensive evaluation by a healthcare professional is necessary for an accurate assessment of cognitive impairment.
