## Supplementary material for "Text dialogue analysis Based ChatGPT for Primary Screening of Mild Cognitive Impairment": Data(GPT4_vs_GPT3.5_model1-5): 102_GPT3.5 (5).docx

ChatGPT

Based on the provided conversation, here is an assessment of the participant's language features and the severity rating for each evaluation indicator:

Vocabulary Features: a. Word Frequency and Word Ratio: The participant's use of vocabulary appears to be relatively normal. They are able to generate appropriate words to describe the scene, although there are some instances of repetition and limited word variety. Severity Rating: 4

b. Tip-of-the-tongue Phenomenon: There is no evidence of the tip-of-the-tongue phenomenon in the conversation. Severity Rating: 1

Syntax and Grammar Features: a. Syntactic Complexity and Grammatical Components: The participant demonstrates relatively simple sentence structures with occasional errors in subject-verb agreement and verb tense consistency. The use of subordinate clauses is limited. Severity Rating: 5

b. Difficulty with Complex Ideas: The participant shows some difficulty when the narrative becomes more complex. They struggle to provide detailed descriptions and may exhibit hesitations. Severity Rating: 6

Semantics Features: a. Semantic Density and Coherence: The participant's conversation contains meaningful information, but there are instances of redundant phrases and repetitions. The logical consistency and coherence of the text are generally maintained. Severity Rating: 4

b. Memory Issues: There is no clear evidence of memory issues in the conversation. The participant can recall details from the scene and provide relevant information. Severity Rating: 2

Based on the comprehensive synthesis of all the indicators, the participant can be classified as belonging to the Probably Normal Cognition Group. Although there are some language features that suggest mild impairments, such as limited word variety, difficulty with complex ideas, and occasional grammatical errors, they do not exhibit a sufficient number of indicators or severe impairments to warrant classification into the Probably Mild Cognitive Impairment Group. It's important to note that this assessment is based on a single conversation and should be interpreted cautiously. A comprehensive evaluation by a qualified healthcare professional would be necessary for a more accurate diagnosis.
