## Supplementary material for "Text dialogue analysis Based ChatGPT for Primary Screening of Mild Cognitive Impairment": Data(GPT4_vs_GPT3.5_model1-5): 102_GPT3.5.docx

Word ratio: The word ratio is calculated by dividing the number of unique words by the total number of words in the text.

Phrase frequency: To assess the phrase frequency, we count the occurrences of each unique phrase in the text. Unique phrases are distinct combinations of words that appear in the text, not taking into account their repetitions. Each phrase is counted only once, regardless of how many times it occurs.

Phrase ratio: The phrase ratio is calculated by dividing the number of unique phrases by the total number of phrases in the text.

Difficulty finding the right words: We look for frequent pauses or use of filler words, like "uh" or "um," as the participant searches for the desired word.

Lexical complexity: We measure the variety and sophistication of the vocabulary used, considering factors such as word length and frequency of rare words.

Limited vocabulary: We look for repeated use of the same words or simple words, with a lack of variety or sophistication.

Let's perform the assessment based on the provided dialogue:

Word frequency: The assessment requires counting the occurrences of each unique word in the text. Unique words are distinct words that appear in the text, not taking into account their repetitions. Each word is counted only once, regardless of how many times it occurs.

Word ratio: The word ratio is calculated by dividing the number of unique words by the total number of words in the text.

Phrase frequency: The assessment requires counting the occurrences of each unique phrase in the text. Unique phrases are distinct combinations of words that appear in the text, not taking into account their repetitions. Each phrase is counted only once, regardless of how many times it occurs.

Phrase ratio: The phrase ratio is calculated by dividing the number of unique phrases by the total number of phrases in the text.

Difficulty finding the right words: We will look for frequent pauses or use of filler words, like "uh" or "um," as the participant searches for the desired word.

Lexical complexity: We will measure the variety and sophistication of the vocabulary used, considering factors such as word length and frequency of rare words.

Limited vocabulary: We will look for repeated use of the same words or simple words, with a lack of variety or sophistication.

Let's perform the assessment based on the provided dialogue:

Word frequency: The assessment requires counting the occurrences of each unique word in the text. Unique words are distinct words that appear in the text, not taking into account their repetitions. Each word is counted only once, regardless of how many times it occurs.

Here are the unique words and their frequencies in the text:

Word: Count okay: 8 running: 3 floor: 1 uh: 14 um: 3 falling: 2 stool: 2 standing: 2 water: 4 see: 6 nothing: 3 right: 3 snow: 1 outside: 1 naturally: 1 ordinary: 1 going: 3 sink: 4 kid: 3 mm: 1 makin: 1 fun: 2 girl: 1 boy: 2 cookie: 2 jar: 2 trip: 1 fall: 1 mother: 1 listening: 1 kids: 1 drying: 1 dishes: 1 window: 2 open: 4 mirror: 1 paying: 1 attention: 1 spilling: 1 feet: 1 wet: 1 curtains: 1 allow: 1 fresh: 1 air: 1 come: 1 house: 1 two: 1 cups: 1 dish: 1 faucets: 1 cupboard: 1 door: 1 looks: 1 spring: 1 summer: 1 shoes: 4 dress: 3 apron: 1 shirt: 1 shorts: 1 socks: 2 lid: 1 fancy: 2 cabinet: 2 wall: 1 about: 2 good: 1

Total unique words: 60

Word ratio: The word ratio is calculated by dividing the number of unique words by the total number of words in the text.

Total words in the text: 146

Word ratio = 60/146 = 0.41 (rounded to two decimal places)

Phrase frequency: The assessment requires counting the occurrences of each unique phrase in the text. Unique phrases are distinct combinations of words that appear in the text, not taking into account their repetitions. Each phrase is counted only once, regardless of how many times it occurs.

Here are the unique phrases and their frequencies in the text:

Phrase: Count okay: 2 sink's running over: 1 kid's on a stool: 1 falling over: 1 she's standing in the water: 1 that's about it: 2 it looks like it: 1 no it ain't writing upside down here: 1 that's all: 1 the girl's making fun of the boy: 1 so much while he was stealing a cookie out of the cookie jar: 1 and it made him trip: 1 he's going to fall: 1 the mother's listening to the kids: 1 she's drying dishes: 1 she got the window open against a mirror: 1 she isn't paying attention to the sink: 1 the sink's spilling water onto the floor: 1 it's getting her feet wet: 1 the curtains are open to allow fresh air to come in the house: 1 there's two cups on the sink: 1 a dish on the sink: 1 the faucets are open and the cupboard door is open: 1 looks like it might be spring or summer out because things look like they're growing outside: 1 she has shoes on: 1 she has a dress: 1 dress: 2 apron: 1 shirt: 1 shorts: 1 socks: 2 the lid is off the cookie jar: 1 a fancy sink cabinet over here and a fancy wall cabinet over there: 1 that's about it: 1 good: 1

Total unique phrases: 34

Phrase ratio: The phrase ratio is calculated by dividing the number of unique phrases by the total number of phrases in the text.

Total phrases in the text: 35

Phrase ratio = 34/35 = 0.97 (rounded to two decimal places)

Difficulty finding the right words: There are several instances of the participant pausing or using filler words like "uh" and "um" as they search for the desired words.

Lexical complexity: The vocabulary used by the participant is relatively simple and lacks variety. There is a repetition of words such as "shoes," "dress," "socks," and "okay." There are no significant indications of lexical complexity or the use of rare words.

Limited vocabulary: The participant demonstrates limited vocabulary by repeatedly using simple words and phrases. There is a lack of variety or sophistication in their choice of words.

Assessment of Semantics features:

Semantic density: The assessment requires measuring the amount of meaningful information in the text, considering factors such as the number of content words (nouns, verbs, adjectives, adverbs) relative to function words (prepositions, pronouns, conjunctions, etc.).

Inability to convey meaning: There are no significant difficulties observed in the participant's ability to express ideas clearly. However, there are instances where the participant hesitates or repeats words.

Semantic coherence: The text demonstrates some level of logical consistency and meaningfulness. The participant provides descriptions and explanations of the depicted scene, even though there are occasional repetitions and hesitations.

Lack of coherence: The participant maintains a reasonable flow of ideas, with thoughts generally appearing related and connected.

Assessment of Syntax and Grammar features:

Syntactic complexity: The sentence structure in the text is relatively simple, with predominantly short and straightforward sentences. There is a lack of complex sentence structures or the use of subordinate clauses.

Simplified sentence structure: The participant tends to use short, simple sentences.

Grammatical components: The participant demonstrates a reasonable usage of grammar with correct subject-verb agreement, tense consistency, and the proper use of pronouns, prepositions, and conjunctions.

Grammatical errors: There are no frequent mistakes observed in verb tense, subject-verb agreement, or other aspects of grammar.

Based on the assessment of Vocabulary, Semantics, and Syntax and Grammar features, the participant's language shows some indications of limited vocabulary, hesitation in finding the right words, and a relatively simple sentence structure. However, there are no significant semantic or grammatical errors observed.

Given the subtle language gaps, the participant is likely part of the Probably Normal group rather than the Probably Mild Cognitive Impairment group. However, it is important to note that a conclusive determination of cognitive impairment cannot be made based solely on a conversation, and a comprehensive evaluation by a medical professional is necessary for an accurate diagnosis.
