## Supplementary material for "Text dialogue analysis Based ChatGPT for Primary Screening of Mild Cognitive Impairment": Data(GPT4_vs_GPT3.5_model1-5): 102_GPT4 (2).docx

ChatGPT

Based on the text provided, let's analyze the participant's responses:

1. **Vocabulary Features**:
   - *Word frequency and Word ratio*: The participant uses a variety of words in their responses. However, there are some repetitions such as "she's", "uh", "it's", and "the". The ratio of unique words to the total number of words is relatively high, indicating a good vocabulary range.
   - *Phrase frequency and Phrase ratio*: The participant uses a diverse range of phrases to describe the scene. However, there are repetitions such as "she's standing in water" and "the sink's running over". The ratio of unique phrases to the total phrases seems high, pointing towards a diverse phrase usage.
   - *Word-finding difficulties*: The participant frequently uses "uh", which might be indicative of some word-finding difficulties.
   - *Repetition*: The participant does repeat certain phrases and ideas a few times throughout the conversation.
   - *Lexical complexity*: The participant uses a variety of words, including more complex terms such as "spigot" and "fancy sink cabinet".
   - *Limited vocabulary*: The participant doesn't seem to have a limited vocabulary as they use a variety of different words and phrases.
2. **Semantics Features**:
   - *Semantic density*: The participant's responses contain a mix of content words and function words, indicating a fair semantic density.
   - *Inability to convey meaning*: The participant seems capable of conveying meaning, as their descriptions of the scene are clear.
   - *Semantic coherence*: The participant's responses seem logically consistent and meaningful, demonstrating semantic coherence.
   - *Lack of coherence*: The participant doesn't seem to struggle with maintaining a logical flow of ideas.
3. **Syntax and Grammar Features**:
   - *Syntactic complexity*: The participant uses a variety of sentence structures, indicating moderate syntactic complexity.
   - *Simplified sentence structure*: The participant doesn't consistently use simplified sentence structures.
   - *Grammatical components*: The participant seems to have a good grasp of grammar, as their responses generally follow grammatical rules.
   - *Grammatical errors*: There are some minor grammatical errors, such as "she's isn't paying attention to the sink".

Based on this analysis, it's difficult to definitively assign the participant to a group. There are some minor indications of potential cognitive impairment, such as the frequent use of "uh" and some repetitions. However, these could also be a part of normal conversational patterns. Therefore, I would tentatively place the participant in the "Probably Normal" group, but recommend further assessments for a more definitive conclusion.
