## Supplementary material for "Text dialogue analysis Based ChatGPT for Primary Screening of Mild Cognitive Impairment": Data(GPT4_vs_GPT3.5_model1-5): 102_GPT4 (3).docx

ChatGPT

Analysis based on the provided dialogue:

**Vocabulary features:**

1. Limited vocabulary: The participant appears to have slightly reduced vocabulary with more frequent use of the same words and common terms. For example, words like "uh", "okay", and "running" are repeated often. Rating: 6/10
2. Increased verbosity: The participant does seem to use more words than necessary, often repeating the same ideas. Rating: 5/10
3. Overlearned phrases: There isn't much evidence of the participant overusing cliches or stock phrases. Rating: 3/10
4. Improper use of pronouns: There doesn't seem to be a problem with pronouns in this dialogue. Rating: 2/10
5. Hesitation (Word-finding difficulties): Frequent use of filler words like "uh" and "um", indicating some word-finding difficulties. Rating: 6/10
6. Circumlocution (Word-finding difficulties): There is some evidence of circumlocution, with the participant using many words where fewer would do. Rating: 6/10
7. Tip-of-the-tongue phenomenon: This phenomenon isn't particularly evident in the provided dialogue. Rating: 3/10
8. Repetition: There is a degree of repetition in the participant's responses, repeating phrases like "she's standing in water", "she has shoes on". Rating: 7/10

**Syntax and Grammar features:**

1. Simplified sentence structure and grammar: The participant uses simpler sentence structures and seems to avoid complex grammatical constructions. Rating: 5/10
2. Grammatical errors: The participant makes a few grammatical errors, like "I don't see nothin(g) else". Rating: 5/10
3. Difficulty with complex ideas: The participant does not seem to have difficulty when the narrative becomes complex. Rating: 2/10

**Semantics features:**

1. Semantic paraphasias: There is no clear evidence of semantic paraphasias. Rating: 2/10
2. Lack of coherence: The participant sometimes struggles with organizing thoughts coherently, often repeating the same information or going off on tangents. Rating: 5/10
3. Consistency in errors: There doesn't seem to be consistent language errors. Rating: 2/10
4. Lack of semantic fluency: The participant seems to have trouble describing a sequence of events and tends to repeat the same details. Rating: 5/10
5. Disorganized narrative structure: The participant struggles to maintain a coherent line of thought at times. Rating: 5/10
6. Orientation Issues: No obvious orientation issues are evident in this dialogue. Rating: 1/10
7. Decline in conversational engagement and responsiveness: There is some evidence of this, with the participant repeating questions and steering conversations back to familiar topics. Rating: 6/10
8. Memory issues: The participant doesn't show clear memory issues. Rating: 2/10
9. Lack of abstract thinking and synthesis: The participant seems to have a difficulty in synthesizing the overall situation. Rating: 4/10
10. Abnormal emotions: There are no clear signs of abnormal emotions in the participant's responses. Rating: 1/10

**Final Evaluation:**

The participant shows some signs of Mild Cognitive Impairment (MCI), such as a limited vocabulary, increased verbosity, word-finding difficulties, repetition, simplified sentence structure and grammar, lack of semantic fluency, and some decline in conversational engagement and responsiveness. However, it's important to remember that this is just an initial screen and further cognitive tests would be required for an accurate diagnosis. Based on the dialogue provided, the participant is classified as "Probably Mild Cognitive Impairment Group"
