## Supplementary material for "Text dialogue analysis Based ChatGPT for Primary Screening of Mild Cognitive Impairment": Data(GPT4_vs_GPT3.5_model1-5): 102_GPT4 (4).docx

ChatGPT

**Vocabulary Features**

a. Word frequency, word ratio, phrase frequency, and phrase ratio are generally acceptable, though some repetition is noticed.

b. Lexical complexity is moderate, with some use of specific terms and elaboration in descriptions.

c. Severity assessment:

- Limited Vocabulary: 2/10 (The participant uses an adequate range of vocabulary)
- Increased Verbosity: 6/10 (The participant gives detailed descriptions and tends to over-clarify)
- Overlearned Phrases: 2/10 (Few clichéd phrases were used)
- Improper Use of Pronouns: 1/10 (Pronouns are used appropriately)
- Hesitation: 6/10 (Notable pauses and filler words are used)
- Circumlocution: 5/10 (Participant occasionally provides roundabout descriptions)
- Tip-of-the-Tongue Phenomenon: 5/10 (Some apparent difficulty in immediately accessing certain words)
- Repetition: 7/10 (Several repeated phrases and descriptions are present)

**Syntax and Grammar Features**

a. Syntactic complexity is moderate, with some complex sentence formations.

b. Severity assessment:

- Simplified Sentence Structure and Grammar: 3/10 (Participant can formulate complex sentences)
- Grammatical Errors: 2/10 (Few grammatical mistakes are present)
- Difficulty with Complex Ideas: 4/10 (Some struggle with expressing complex concepts smoothly)

**Semantics Features**

a. Semantic density and coherence are generally good, with most descriptions logically connected and contextually appropriate.

b. Severity assessment:

- Semantic Paraphasias: 2/10 (Few, if any, instances of incorrect word use)
- Lack of Coherence: 4/10 (Participant occasionally has issues with logical sequencing)
- Consistency in Errors: 3/10 (Errors are not overly frequent nor consistent)
- Lack of Semantic Fluency: 4/10 (Participant's flow of speech is somewhat disrupted by hesitations)
- Disorganized Narrative Structure: 5/10 (Some degree of disorganization in the narrative structure)
- Orientation Issues: 2/10 (Participant appears aware of the situation)
- Decline in Conversational Engagement and Responsiveness: 3/10 (Participant remains engaged and responsive)
- Memory Issues: 5/10 (Repetition could suggest memory issues)
- Lack of Abstract Thinking and Synthesis: 3/10 (Participant can draw inferences and abstract ideas)
- Abnormal Emotions: 1/10 (No abnormal emotions detected)

Based on this analysis, I would suggest that the participant falls into the **Probably Mild Cognitive Impairment Group**. Although this assessment is based on language features, it does not constitute a diagnosis, and any concerns should be followed up with a healthcare provider. Notably, the participant's hesitation, circumlocution, and repetition might point towards potential cognitive impairment.
