## Supplementary material for "Text dialogue analysis Based ChatGPT for Primary Screening of Mild Cognitive Impairment": Data(GPT4_vs_GPT3.5_model1-5): 102_GPT4 (5).docx

ChatGPT

Vocabulary Features: a. The participant uses a variety of unique words and phrases, demonstrating some lexical complexity. Repeated words such as "cookie," "sink," and "water" are relevant to the narrative, contributing to the theme of the story. There is a relatively balanced usage of pronouns and specific nouns. The word frequency and phrase frequency may rate around a 5 out of 10, showing average vocabulary utilization. b. The tip-of-the-tongue phenomenon is somewhat present, with the participant displaying hesitation and partial recall in some instances, such as "…uh …uh …sl falling off the stool." and "... it ain't writing upside down here." I would rate this aspect as a 6 out of 10.

Syntax and Grammar Features: a. The participant uses a mix of simple and complex sentences, demonstrating some syntactic complexity. However, there are instances where sentences are fragmented or incorrect, such as "…uh the kid's reachin(g) in for cookies which is correct but he's …d …uh …uh …sl falling off the stool." I would rate the syntactic complexity as a 5 out of 10, and the grammatical component as a 5 out of 10 due to some grammatical errors. b. Difficulty with complex ideas is visible but not too severe. For example, the participant is able to maintain a coherent narrative about the picture and describe events happening in it, but sometimes struggles with elaboration or complex phrases. This could be rated a 4 out of 10.

Semantics Features: a. The semantic density of the text is moderate, with a good balance of content and function words. The participant provides meaningful information relevant to the context. The semantic coherence is also good, with logical consistency in the narrative. I would rate both aspects at around a 5 out of 10. b. The participant does show some signs of memory issues, repeating certain information such as "she has shoes on." multiple times, and seems to ask for reassurance with phrases like "did I mention everything?". I would rate this aspect as a 6 out of 10.

Comprehensive synthesis: The participant does show some signs that could be associated with Mild Cognitive Impairment, such as the tip-of-the-tongue phenomenon and minor memory issues. However, overall, their language features suggest that their cognitive function might still be in the normal range. Therefore, based on this assessment, the participant could be placed in the "Probably Normal Cognition Group." It's important to note that this is just an initial screen and further evaluation would be needed for a more definitive understanding of the participant's cognitive status.
