## Supplementary material for "Text dialogue analysis Based ChatGPT for Primary Screening of Mild Cognitive Impairment": Data(GPT4_vs_GPT3.5_model1-5): 103.txt

Cookie Theft Picture Description
Recall 1: George and Melanie: Just after Thanksgiving George Miller moved in with his son's family. His granddaughter, Melanie, enjoyed listening to the wonderful stories about his childhood in the city. She asked if they could visit the house where he had lived as a child. One Friday, George and Melanie set off on their trip. The streets were crowded and George held Melanie's hand tightly. The little girl was frightened of the noisy streets and, although she did not tell her grandfather, she was pleased to return to the country.
Recall 2: Uncle Bill and Johnny: Bill returned to the USA on 4 July. As with other relatives, his nephew Johnny welcomed him back very much. They used to spend happy times in the summer, like camping and fishing.
Sentence Construction task

INVESTIGATOR:	what I want you to do here is take a look a that picture . 
INVESTIGATOR:	I want you to tell me everything that you see happening in that picture . 
INVESTIGATOR:	everything that you see going on in there . 
PARTICIPANT:	well  the kids are raiding the cookie jar . 
PARTICIPANT:	and he's about to fall off the stool . 
PARTICIPANT:	mom's washing the dishes but the …uh sink's overflowing . 
PARTICIPANT:	and …um …ts there's a view out the window . 
PARTICIPANT:	get some dishes here some cups . 
PARTICIPANT:	and the kitchen cabinets .  
PARTICIPANT:	what … do you mean what's goin(g) on ?  
INVESTIGATOR:	mhm . 
PARTICIPANT:	that's about it .  
INVESTIGATOR:	okay . 
PARTICIPANT:	the window's open . 
INVESTIGATOR:	okay . 
INVESTIGATOR:	alright thank_you .

"
12:56
"
INVESTIGATOR:	can you tell me everything you can remember from that ? 
PARTICIPANT:	well  George moved in with his son ,  his granddaughter …um Melanie . 
PARTICIPANT:	very interested in his stories about his … …um where he's brought up . 
PARTICIPANT:	and then they went and took her … he took her out to visit the city . 
PARTICIPANT:	and she was frightened but he grabbed her hand . 
PARTICIPANT:	and it was xxx (be)cause the streets were very crowded . 
PARTICIPANT:	that's about it .  
INVESTIGATOR:	okay  good . 
"
20:36
"
INVESTIGATOR:	do you remember the short story I read to you a while ago ? 
INVESTIGATOR:	and then as_soon_as I was done reading the short story I asked you to recall as much as you could from it . 
INVESTIGATOR:	tell me as much as you can remember about that story now . 
PARTICIPANT:	well … well  let's see .  
PARTICIPANT:	George went to live with his son . 
INVESTIGATOR:	right . 
PARTICIPANT:	and told the granddaughter about the … …uh where he'd been brought up  in the city . 
PARTICIPANT:	grand … then he took his granddaughter . 
PARTICIPANT:	she was interest . 
PARTICIPANT:	she wants to visit the city . 
PARTICIPANT:	granddaughter was scared so he held her hand . 
PARTICIPANT:	and there was lots_of crowds . 
PARTICIPANT:	so …  
INVESTIGATOR:	nice . 
INVESTIGATOR:	good  very good . 

INVESTIGATOR:	pencil . 
PARTICIPANT:	write with the pencil . 
INVESTIGATOR:	tree . 
PARTICIPANT:	cut the tree down . 
INVESTIGATOR:	… now I'm going to give you two words . 
INVESTIGATOR:	I still want you to use them in a sentence and it doesn't matter their order . 
INVESTIGATOR:	child and hospital . 
PARTICIPANT:	child's in the hospital . 
INVESTIGATOR:	cold and winter . 
PARTICIPANT:	winter is cold . 
INVESTIGATOR:	now  I'm going to give you three words so I still want you to use them in a sentence and it doesn't matter their order . 
INVESTIGATOR:	chair , doctor and sit . 
PARTICIPANT:	doctor sits in the chair . 
PARTICIPANT:	sit in it .  
PARTICIPANT:	oh brother .  
INVESTIGATOR:	it doesn't matter . 
INVESTIGATOR:	that's fine . 
INVESTIGATOR:	it's any form of the word . 
INVESTIGATOR:	that's okay . 
INVESTIGATOR:	bureau , open and drawer . 
PARTICIPANT:	the drawer in the bureau was open . 
INVESTIGATOR:	… okay . 
