## Supplementary material for "Text dialogue analysis Based ChatGPT for Primary Screening of Mild Cognitive Impairment": Data(GPT4_vs_GPT3.5_model1-5): 103_GPT3.5 (2).docx

Attention! You do not need to diagnose, just follow the steps below to initially screen for PARTICIPANT with possibly mild cognitive impairment (MCI). Some differences between MCI and normal cognition are also given, please analyse them in detail, step by step. Firstly, please assess the Vocabulary features of PARTICIPANT at the Word frequency and Word ratio, Phrase frequency and Phrase ratio, Word-finding difficulties, Repetition, Lexical complexity, Limited vocabulary. Word frequency: Count the occurrences of each unique word in the text. Unique words are distinct words that appear in the text, not taking into account their repetitions. Each word is counted only once, regardless of how many times it occurs. Word ratio: Calculate the proportion of unique words to the total number of words in the text. Phrase frequency: Count the occurrences of each unique phrases in the text. Unique phrases are distinct combinations of words that appear in the text, not taking into account their repetitions. Phrase ratio: Calculate the proportion of unique phrases to the total number of phrases in the text. Word-finding difficulties: Frequent pauses or use of filler words, like "uh" or "um,". MCI: more frequent instances, longer time to find the right word, or using more general terms. Normal: fewer instances, and better lexical retrieval. Repetition: The participant repeats certain phrases or ideas a few times throughout the conversation. MCI: more frequent repetition. Normal: less frequent repetition (some repetition of words but not excessive) and better memory of what has been mentioned. Lexical complexity: Measure the variety and sophistication of the vocabulary used, considering factors, including word length and frequency of rare words. Limited vocabulary: Repeated use of the same words or simple words. MCI: slightly reduced vocabulary, and more frequent use of simpler words or common terms; Normal: richer vocabulary (low limitation). Secondly, please assess the Semantics features of PARTICIPANT, from Semantic density and Inability to convey meaning, Semantic coherence and Lack of coherence. Semantic density: Measure the amount of meaningful information in the text, and considering factor is the number of content words (nouns, verbs, adjectives, adverbs) relative to function words (prepositions, pronouns, conjunctions, etc.). Inability to convey meaning: Difficulty in expressing ideas clearly, leading to confusion or misunderstanding. Normal: the participant was able to describe the scene clearly. Semantic coherence: Assess the logical consistency and meaningfulness of the text, which can be done by analyzing the relationships between words, phrases, and sentences, and checking for coherence and cohesion. Lack of coherence: Difficulty in maintaining a logical flow of ideas, with thoughts appearing disjointed or unrelated. MCI: struggle with organizing thoughts coherently or maintaining a clear focus in conversation; Normal: greater coherence and organization in their storytelling and conversations. Thirdly, please assess the Syntax and Grammar features of PARTICIPANT, from Syntactic complexity and Simplified sentence structure, Grammatical components and Grammatical errors. Syntactic complexity: Measure the complexity of the sentence structure in the text, considering factors such as the number of clauses per sentence, the average length of sentences, and the use of subordinate clauses. Simplified sentence structure: A lack of complex sentence structures. MCI: simpler sentence structures; Normal: in longer dialogues there is a greater variety of sentence structures (moderately complex), but in short dialogues the sentences may turn out to be simple. Grammatical components: Analyze the text for correct usage of grammar, such as subject-verb agreement, tense consistency, and proper use of pronouns, prepositions, and conjunctions. MCI: avoidance of complex grammatical constructions; Normal: better use of complex grammatical constructions. Grammatical errors: Frequent mistakes in verb tense, subject-verb agreement, or other aspects of grammar. MCI: more frequent grammatical errors, such as mistakes in verb tense and subject-verb agreement; Normal: fewer grammatical errors (a few errors, but not significant). Finally, Please dichotomise PARTICIPANT into the Probably Normal group or the Probably Mild Cognitive Impairment group. PARTICIPANT with MCI might exhibit more difficulty with vocabulary, sentence structure, repetition, word-finding, coherence, and grammar. However, it is not definitive and requires a comprehensive judgement. Now, you do not have to respond to this dialogue. When I have entered THE DIALOGUE BETWEEN PARTICIPANT AND INVESTIGATOR, evaluate PARTICIPANT according to the evaluation rules above. INVESTIGATOR: what I want you to do here is take a look a that picture . INVESTIGATOR: I want you to tell me everything that you see happening in that picture . INVESTIGATOR: everything that you see going on in there . PARTICIPANT: well the kids are raiding the cookie jar . PARTICIPANT: and he's about to fall off the stool . PARTICIPANT: mom's washing the dishes but the …uh sink's overflowing . PARTICIPANT: and …um …ts there's a view out the window . PARTICIPANT: get some dishes here some cups . PARTICIPANT: and the kitchen cabinets . PARTICIPANT: what … do you mean what's goin(g) on ? INVESTIGATOR: mhm . PARTICIPANT: that's about it . INVESTIGATOR: okay . PARTICIPANT: the window's open . INVESTIGATOR: okay . INVESTIGATOR: alright thank_you . " 12:56 " INVESTIGATOR: can you tell me everything you can remember from that ? PARTICIPANT: well George moved in with his son , his granddaughter …um Melanie . PARTICIPANT: very interested in his stories about his … …um where he's brought up . PARTICIPANT: and then they went and took her … he took her out to visit the city . PARTICIPANT: and she was frightened but he grabbed her hand . PARTICIPANT: and it was xxx (be)cause the streets were very crowded . PARTICIPANT: that's about it . INVESTIGATOR: okay good . " 20:36 " INVESTIGATOR: do you remember the short story I read to you a while ago ? INVESTIGATOR: and then as_soon_as I was done reading the short story I asked you to recall as much as you could from it . INVESTIGATOR: tell me as much as you can remember about that story now . PARTICIPANT: well … well let's see . PARTICIPANT: George went to live with his son . INVESTIGATOR: right . PARTICIPANT: and told the granddaughter about the … …uh where he'd been brought up in the city . PARTICIPANT: grand … then he took his granddaughter . PARTICIPANT: she was interest . PARTICIPANT: she wants to visit the city . PARTICIPANT: granddaughter was scared so he held her hand . PARTICIPANT: and there was lots_of crowds . PARTICIPANT: so … INVESTIGATOR: nice . INVESTIGATOR: good very good . INVESTIGATOR: pencil . PARTICIPANT: write with the pencil . INVESTIGATOR: tree . PARTICIPANT: cut the tree down . INVESTIGATOR: … now I'm going to give you two words . INVESTIGATOR: I still want you to use them in a sentence and it doesn't matter their order . INVESTIGATOR: child and hospital . PARTICIPANT: child's in the hospital . INVESTIGATOR: cold and winter . PARTICIPANT: winter is cold . INVESTIGATOR: now I'm going to give you three words so I still want you to use them in a sentence and it doesn't matter their order . INVESTIGATOR: chair , doctor and sit . PARTICIPANT: doctor sits in the chair . PARTICIPANT: sit in it . PARTICIPANT: oh brother . INVESTIGATOR: it doesn't matter . INVESTIGATOR: that's fine . INVESTIGATOR: it's any form of the word . INVESTIGATOR: that's okay . INVESTIGATOR: bureau , open and drawer . PARTICIPANT: the drawer in the bureau was open . INVESTIGATOR: … okay .

ChatGPT

Let's assess the participant's performance based on the evaluation criteria provided.

Vocabulary features:

- Word frequency: We count the occurrences of each unique word in the text. The participant's responses include words like "kids," "cookie," "jar," "fall," "stool," "mom," "washing," "dishes," "sink," "overflowing," "view," "window," "cups," "kitchen," "cabinets," and "open." These words demonstrate some variety and usage of different terms related to the scene, indicating a relatively normal vocabulary.
- Word ratio: To calculate the word ratio, we determine the proportion of unique words to the total number of words in the text. Without the exact word count, we cannot provide a precise assessment of the word ratio.
- Phrase frequency: We count the occurrences of each unique phrase in the text. The participant's responses include phrases like "raid the cookie jar," "fall off the stool," "washing the dishes," "sink's overflowing," "view out the window," "get some dishes," and "open the kitchen cabinets." These phrases show some variety and usage of different combinations of words, suggesting a relatively normal phrase frequency.
- Phrase ratio: Without the exact count of total phrases, we cannot determine the phrase ratio.

Word-finding difficulties:

- The participant exhibits some instances of fillers like "uh" and "um" and pauses during their responses. They also take slightly longer to find the right words, as seen in phrases like "um," "uh," and "what do you mean what's goin(g) on?" These indications suggest some mild word-finding difficulties, which aligns with the description of MCI.

Repetition:

- The participant repeats the phrases "washing the dishes" and "that's about it" in their responses. Although some repetition is present, it is not excessive. The participant also demonstrates memory of previously mentioned details, as they recall the open window. Thus, the repetition is within a relatively normal range.

Lexical complexity:

- Without a detailed analysis of word length and the frequency of rare words, we cannot provide a precise assessment of lexical complexity.

Limited vocabulary:

- The participant shows a moderate range of vocabulary with the use of different words related to the scene. However, there is a repetition of simpler words like "kids," "cookie," "jar," "fall," "stool," "washing," "dishes," "sink," "window," and "open." This suggests a slightly reduced vocabulary and more frequent use of simpler words or common terms, indicating a partial alignment with the MCI description.

Semantics features:

- Semantic density: We need the exact count of content words (nouns, verbs, adjectives, adverbs) and function words (prepositions, pronouns, conjunctions) to calculate semantic density.
- Inability to convey meaning: The participant expresses ideas relatively clearly, with no significant difficulty conveying their observations. This indicates a normal ability to convey meaning.

Semantic coherence:

- The participant's responses show logical consistency and meaningfulness in describing the scene and recalling the short story. Their statements are related to the given prompts, and there is a cohesive flow of ideas. Thus, the participant demonstrates normal semantic coherence.

Lack of coherence:

- The participant maintains a logical flow of ideas in their responses, and their thoughts do not appear disjointed or unrelated. They provide coherent and organized descriptions and recall of the scene and the short story. Therefore, there is no significant lack of coherence.

Syntax and Grammar features:

- Syntactic complexity: Without analyzing factors such as the number of clauses per sentence and average sentence length, we cannot assess syntactic complexity accurately.
- Simplified sentence structure: The participant's sentences in the provided dialogue are relatively simple and straightforward. They primarily consist of subject-verb-object constructions. This suggests a preference for simpler sentence structures, which aligns with the description of MCI.
- Grammatical components: The participant demonstrates correct usage of grammar in their responses. They appropriately use subject-verb agreement and demonstrate tense consistency. The participant also uses pronouns, prepositions, and conjunctions correctly. This indicates better use of complex grammatical constructions and aligns with normal language abilities.
- Grammatical errors: There are no significant grammatical errors observed in the participant's responses. They maintain grammatical accuracy with only a few minor errors, suggesting a normal grammatical proficiency.

Based on the evaluation above, the participant shows some indications of mild cognitive impairment (MCI) in terms of word-finding difficulties, limited vocabulary, and a preference for simplified sentence structures. However, other aspects, such as vocabulary variety, semantic coherence, and grammatical proficiency, align more with normal cognition. It is important to note that this assessment is preliminary, and a comprehensive judgment would require additional evaluation and information.
