## Supplementary material for "Text dialogue analysis Based ChatGPT for Primary Screening of Mild Cognitive Impairment": Data(GPT4_vs_GPT3.5_model1-5): 103_GPT3.5 (3).docx

ChatGPT

Based on the provided conversation, here is an analysis of the indicators and evaluation indicators for screening for possible mild cognitive impairment (MCI) in the participant:

1. Vocabulary Features:

- Limited vocabulary: The participant demonstrates a slightly reduced vocabulary and more frequent use of the same words or common terms. (Severity: 3)
- Increased verbosity: The participant tends to be more verbose and uses more words than necessary. (Severity: 4)
- Overlearned phrases: The participant does not exhibit significant overuse of cliches or stock phrases. (Severity: 1)
- Improper use of pronouns: The participant does not show specific difficulties with pronouns. (Severity: 1)
- Hesitation (Word-finding difficulties): The participant shows mild word-finding difficulties with some pauses and filler words. (Severity: 3)
- Circumlocution (Word-finding difficulties): The participant does not exhibit significant circumlocution or deliberate vagueness in their speech. (Severity: 1)
- Tip-of-the-tongue phenomenon: There is no evidence of tip-of-the-tongue phenomenon in the conversation. (Severity: 1)
- Repetition: The participant does not excessively repeat stories, questions, or statements. (Severity: 1)

2. Syntax and Grammar Features:

- Simplified sentence structure and grammar: The participant prefers simpler sentence structures, even in longer dialogues. (Severity: 4)
- Grammatical errors: The participant makes a few grammatical errors. (Severity: 2)
- Difficulty with complex ideas: The participant shows some difficulty when the narrative or questions become more complex. (Severity: 3)

3. Semantics Features:

- Semantic paraphasias: The participant does not exhibit significant errors in word choice or odd combinations of words. (Severity: 1)
- Lack of coherence: The participant demonstrates some struggle with maintaining coherence and focus in the conversation. (Severity: 3)
- Consistency in errors: There is no evidence of consistent language errors. (Severity: 1)
- Lack of semantic fluency: The participant does not have significant difficulty naming items or describing processes. (Severity: 1)
- Disorganized narrative structure: The participant shows some difficulty in maintaining a coherent line of thought and chronology. (Severity: 3)
- Orientation issues: There is no evidence of orientation issues in the conversation. (Severity: 1)
- Decline in conversational engagement and responsiveness: The participant exhibits decreased spontaneity and engagement in the conversation. (Severity: 4)
- Memory issues: The participant shows some difficulty in recalling specific details or plot points. (Severity: 3)
- Lack of abstract thinking and synthesis: The participant demonstrates the ability to describe details but struggles with synthesizing the overall situation or expressing abstract concepts. (Severity: 3)
- Abnormal emotions: There is no evidence of abnormal emotions in the conversation. (Severity: 1)

Based on the comprehensive synthesis of the indicators, the participant shows several indicators that suggest the possibility of mild cognitive impairment (MCI), including limited vocabulary, increased verbosity, hesitation in word-finding, simplified sentence structure and grammar, difficulty with complex ideas, lack of coherence, disorganized narrative structure, decline in conversational engagement, memory issues, and lack of abstract thinking and synthesis. However, it's important to note that not all indicators are present, and some of these issues can also be observed in individuals with normal cognitive function. Therefore, further evaluation and assessment are necessary to make a conclusive determination.

Dichotomization: Based on the analysis, the participant is more likely to belong to the Probably Mild Cognitive Impairment Group.
