## Supplementary material for "Text dialogue analysis Based ChatGPT for Primary Screening of Mild Cognitive Impairment": Data(GPT4_vs_GPT3.5_model1-5): 103_GPT3.5 (4).docx

ChatGPT

Let's assess the participant's language features based on the provided conversation. Please note that the severity ratings are subjective and based on the information provided.

Vocabulary features: a. Analyze word frequency, word ratio, phrase frequency, and phrase ratio. Since we don't have the complete transcript of the conversation, it's difficult to analyze word frequency, word ratio, phrase frequency, and phrase ratio accurately.

b. Calculate the lexical complexity. The participant's lexical complexity appears to be relatively normal based on the given conversation.

c. Assess and rate the severity of the following aspects: limited vocabulary, increased verbosity, overlearned phrases, improper use of pronouns, hesitation, circumlocution, tip-of-the-tongue phenomenon, and repetition.

- Limited vocabulary: No evidence of limited vocabulary.
- Increased verbosity: Mild to moderate verbosity is observed, with the participant occasionally providing unnecessary details or repeating information. Severity rating: 3
- Overlearned phrases: No evidence of overlearned phrases.
- Improper use of pronouns: No evidence of improper use of pronouns.
- Hesitation: Some hesitation is observed, such as fillers (e.g., "uh," "um") and pauses. Severity rating: 2
- Circumlocution: The participant occasionally uses circumlocution, struggling to find the right words or descriptions. Severity rating: 3
- Tip-of-the-tongue phenomenon: No evidence of the tip-of-the-tongue phenomenon.
- Repetition: The participant repeats certain phrases and ideas during the conversation. Severity rating: 2

Syntax and grammar features: a. Analyze syntactic complexity and grammatical components. Since the conversation is relatively short, it's challenging to assess syntactic complexity accurately.

b. Assess and rate the severity of the following aspects: simplified sentence structure and grammar, grammatical errors, and difficulty with complex ideas.

- Simplified sentence structure and grammar: The participant's sentence structure and grammar are generally appropriate, with some occasional errors. Severity rating: 2
- Grammatical errors: Some minor grammatical errors are present in the conversation. Severity rating: 2
- Difficulty with complex ideas: The participant appears to understand and convey complex ideas without significant difficulty.

Semantics features: a. Analyze semantic density and coherence. It's challenging to analyze semantic density and coherence without a complete transcript of the conversation.

b. Assess and rate the severity of the following aspects: semantic paraphasias, lack of coherence, consistency in errors, lack of semantic fluency, disorganized narrative structure, orientation issues, decline in conversational engagement and responsiveness, memory issues, lack of abstract thinking and synthesis, and abnormal emotions.

- Semantic paraphasias: No evidence of semantic paraphasias.
- Lack of coherence: The participant's responses generally maintain coherence and relevance to the topics discussed. Severity rating: 1
- Consistency in errors: Some consistent errors, such as hesitation and repetition, are present but not significantly impacting overall communication.
- Lack of semantic fluency: The participant demonstrates a relatively good level of semantic fluency, with occasional circumlocution.
- Disorganized narrative structure: No evidence of a significantly disorganized narrative structure.
- Orientation issues: No evidence of orientation issues.
- Decline in conversational engagement and responsiveness: No evidence of a significant decline in conversational engagement and responsiveness.
- Memory issues: The participant's memory appears to be intact, as they were able to recall details from previous discussions. Severity rating: 1
- Lack of abstract thinking and synthesis: The participant demonstrates the ability to think abstractly and synthesize information.
- Abnormal emotions: No evidence of abnormal emotions.

Based on the comprehensive synthesis of all the indicators, the participant can be classified into the Probably Normal Cognition Group. Although there are some mild language features that suggest possible mild cognitive impairment (e.g., verbosity, hesitation, circumlocution), these features are not severe enough or consistently present to definitively indicate mild cognitive impairment.
