## Supplementary material for "Text dialogue analysis Based ChatGPT for Primary Screening of Mild Cognitive Impairment": Data(GPT4_vs_GPT3.5_model1-5): 103_GPT3.5 (5).docx

ChatGPT

To assess the participant's language features for possible mild cognitive impairment (MCI), we'll evaluate the indicators mentioned in the request and rate their severity on a scale of 1 to 10. Please note that the categorization into "Probably Normal Cognition Group" or "Probably Mild Cognitive Impairment Group" will be based on a comprehensive synthesis of all the indicators.

Vocabulary Features: a. Analyzing word frequency, word ratio, phrase frequency, and phrase ratio:

- Word frequency and word ratio: Not evaluated in the provided conversation.
- Phrase frequency and phrase ratio: Not evaluated in the provided conversation.
- Lexical complexity: Not evaluated in the provided conversation.

b. Assessing tip-of-the-tongue phenomenon:

- Severity rating: 3
- Explanation: The participant experiences partial recall, mentioning instances where they couldn't recall certain words fully but had some information about them, such as the first letter or syllable structure. This is a mild symptom often seen in individuals with MCI.

Syntax and Grammar Features: a. Analyzing syntactic complexity and grammatical components:

- Syntactic complexity: Not evaluated in the provided conversation.
- Grammatical components: Not evaluated in the provided conversation.

b. Assessing difficulty with complex ideas:

- Severity rating: 2
- Explanation: The participant appears to have some difficulty with more complex questions and narratives. While they can handle simpler sentences and ideas, they struggle when the information becomes more intricate.

Semantics Features: a. Analyzing semantic density and coherence:

- Semantic density: Not evaluated in the provided conversation.
- Semantic coherence: Not evaluated in the provided conversation.

b. Assessing memory issues:

- Severity rating: 3
- Explanation: The participant displays mild memory issues. While they were able to recall some details from the picture and short story, they omitted certain information and had difficulty recalling specific details, such as the overflowing sink in the picture or additional plot points from the short story.

Based on the indicators assessed, the participant's severity ratings for each aspect, and the comprehensive synthesis of these indicators, it's likely that the participant falls into the Probably Mild Cognitive Impairment Group. However, it's important to note that a formal diagnosis should be conducted by a healthcare professional specializing in cognitive assessment to confirm the presence of mild cognitive impairment or any other cognitive condition. This initial screening tool can serve as a starting point for further evaluation and monitoring.
