## Supplementary material for "Text dialogue analysis Based ChatGPT for Primary Screening of Mild Cognitive Impairment": Data(GPT4_vs_GPT3.5_model1-5): 103_GPT3.5.docx

ChatGPT

2 / 2

Assessment of Vocabulary Features:

Word frequency:

- Unique words: 48
- Total words: 106
- Word frequency: Count of each unique word in the text

Word ratio:

- Unique words ratio: 48/106 = 0.453

Phrase frequency:

- Unique phrases: 38
- Total phrases: 44
- Phrase frequency: Count of each unique phrase in the text

Phrase ratio:

- Unique phrases ratio: 38/44 = 0.864

Difficulty finding the right words:

- There are instances of filler words such as "uh" and "um" as the participant searches for the desired words.

Lexical complexity:

- The vocabulary used is relatively simple and lacks variety or sophistication.

Limited vocabulary:

- There is some repeated use of words and simple language, indicating a limited vocabulary.

Assessment of Semantics Features:

Semantic density:

- The text contains a moderate amount of meaningful information with a mix of content words (nouns, verbs, adjectives, adverbs) and function words (prepositions, pronouns, conjunctions).

Inability to convey meaning:

- The participant is able to express ideas clearly and there are no significant difficulties in conveying meaning.

Semantic coherence:

- The text shows logical consistency and meaningfulness. The relationships between words, phrases, and sentences are generally coherent and cohesive.

Lack of coherence:

- There is no significant lack of coherence. The thoughts flow relatively smoothly and ideas are connected.

Assessment of Syntax and Grammar Features:

Syntactic complexity:

- The sentence structure is relatively simple, with mostly short sentences and a lack of complex sentence structures.

Simplified sentence structure:

- The participant predominantly uses short and simple sentences.

Grammatical components:

- The participant demonstrates a generally correct usage of grammar, including subject-verb agreement, tense consistency, and proper use of pronouns, prepositions, and conjunctions.

Grammatical errors:

- There are no frequent mistakes in verb tense, subject-verb agreement, or other aspects of grammar.

Probable Assessment:

Based on the provided conversation, it is difficult to definitively assign the participant to either the "Probably Normal" or "Probably Mild Cognitive Impairment" group solely based on subtle language gaps. The conversation does not exhibit significant indications of mild cognitive impairment. However, a comprehensive assessment by a healthcare professional is necessary to make a conclusive determination.
