## Supplementary material for "Text dialogue analysis Based ChatGPT for Primary Screening of Mild Cognitive Impairment": Data(GPT4_vs_GPT3.5_model1-5): 103_GPT4 (2).docx

ChatGPT

Thank you for the detailed instructions. Here's the analysis:

**Vocabulary Features**

- Word frequency and Word ratio: The participant uses 81 unique words out of a total of 189 words, yielding a word ratio of approximately 0.43.
- Phrase frequency and Phrase ratio: A comprehensive phrase frequency analysis would require a more complex model, but it's apparent that the participant does not repeat phrases excessively.
- Word-finding difficulties: The participant has some difficulties, noticeable from the frequent use of filler words like "uh" and "um".
- Repetition: The participant doesn't seem to excessively repeat phrases or ideas.
- Lexical complexity: The vocabulary used is not overly sophisticated, but it does contain a mix of simple and complex words.
- Limited vocabulary: There is some repetition of simpler words, but the participant also uses more complex terms.

**Semantics Features**

- Semantic density: The participant uses a mix of content words and function words, providing a reasonable amount of meaningful information.
- Inability to convey meaning: The participant does have slight difficulties in conveying meaning in certain parts of the conversation, but generally manages to communicate the essential ideas.
- Semantic coherence: The participant maintains a logical flow of ideas for the most part, although there are instances of confusion and slight lack of coherence.
- Lack of coherence: The participant generally maintains coherence but has moments of difficulty in organizing thoughts coherently.

**Syntax and Grammar Features**

- Syntactic complexity: The participant uses mostly simple sentence structures, with a few more complex sentences.
- Simplified sentence structure: The participant's sentences are generally simple, indicating a potential lack of complex sentence structures.
- Grammatical components: The participant uses correct grammar in most cases, but there are a few instances where more complex grammatical constructions are avoided.
- Grammatical errors: There are a few instances of grammatical errors, such as the use of "interest" instead of "interested", but they are not frequent.

In summary, while the participant seems to have slight difficulty in some areas such as word-finding and maintaining semantic coherence, the issues aren't excessive or significantly impairing. The participant is able to convey meaning, maintain a logical flow of ideas, and use a mix of simple and complex words and sentences. Therefore, the participant is probably in the **Probably Mild Cognitive Impairment group**, but it's a mild case and further diagnostic evaluation would be needed to confirm.
