## Supplementary material for "Text dialogue analysis Based ChatGPT for Primary Screening of Mild Cognitive Impairment": Data(GPT4_vs_GPT3.5_model1-5): 103_GPT4 (3).docx

ChatGPT

Given the conversation excerpts provided, we can infer the following. Please note that this analysis is preliminary and does not constitute a formal diagnosis of cognitive ability or cognitive impairment:

Vocabulary Features:

1. Limited vocabulary: The participant seems to have a sufficient range of vocabulary. Rating: 3/10
2. Increased verbosity: The participant uses a reasonable amount of words to express ideas, not too verbose. Rating: 2/10
3. Overlearned phrases: The participant doesn't appear to overuse cliches or stock phrases. Rating: 2/10
4. Improper use of pronouns: The participant appears to use pronouns appropriately. Rating: 1/10
5. Hesitation (Word-finding difficulties): The participant does seem to pause or use filler words frequently. Rating: 6/10
6. Circumlocution (Word-finding difficulties): There's not enough evidence to suggest a high level of circumlocution. Rating: 3/10
7. Tip-of-the-tongue phenomenon: Some instances are present, but not excessive. Rating: 4/10
8. Repetition: The participant does repeat some information but not excessively. Rating: 4/10

Syntax and Grammar Features:

1. Simplified sentence structure and grammar: The participant seems to use a mix of sentence structures. Rating: 3/10
2. Grammatical errors: The participant makes few grammatical errors. Rating: 2/10
3. Difficulty with complex ideas: The participant doesn't seem to struggle excessively with complex ideas. Rating: 4/10

Semantics Features:

1. Semantic paraphasias: The participant seems able to express ideas relatively clearly. Rating: 3/10
2. Lack of coherence: The participant maintains a reasonable level of coherence. Rating: 3/10
3. Consistency in errors: There don't seem to be consistent unusual language errors. Rating: 2/10
4. Lack of semantic fluency: The participant shows some difficulties in describing sequences of events. Rating: 4/10
5. Disorganized narrative structure: Some slight difficulties with maintaining a coherent narrative are present. Rating: 4/10
6. Orientation Issues: No clear orientation issues are present. Rating: 2/10
7. Decline in conversational engagement and responsiveness: The participant is responsive and engaged in conversation. Rating: 3/10
8. Memory issues: The participant seems to have some difficulty recalling specific details. Rating: 5/10
9. Lack of abstract thinking and synthesis: The participant demonstrates some ability to synthesize information, but has some difficulty. Rating: 4/10
10. Abnormal emotions: No clear abnormal emotions or changes in tone are evident. Rating: 2/10

Based on these findings, I would categorize the participant as "Probably Normal Cognition Group". The participant shows some signs that could be consistent with mild cognitive impairment (MCI), such as word-finding difficulties and some memory issues, but overall, the language and cognitive abilities demonstrated here do not show significant impairment. This analysis should be taken with caution and a healthcare professional should be consulted for a definitive diagnosis.
