## Supplementary material for "Text dialogue analysis Based ChatGPT for Primary Screening of Mild Cognitive Impairment": Data(GPT4_vs_GPT3.5_model1-5): 103_GPT4 (4).docx

ChatGPT

**Vocabulary Features**

a. Word frequency, word ratio, phrase frequency, and phrase ratio appear normal, with repetition primarily in response to investigator queries.

b. Lexical complexity is moderate with a decent range of different words and phrases.

c. Severity assessment:

- Limited Vocabulary: 2/10 (The participant uses an adequate range of vocabulary)
- Increased Verbosity: 3/10 (The participant does not tend to over-elaborate)
- Overlearned Phrases: 2/10 (Few clichéd phrases were used)
- Improper Use of Pronouns: 1/10 (Pronouns are used appropriately)
- Hesitation: 3/10 (Some hesitations but not pronounced)
- Circumlocution: 2/10 (There is little evidence of roundabout speech)
- Tip-of-the-Tongue Phenomenon: 3/10 (Some minor struggle with word recall)
- Repetition: 3/10 (Some repetitions are apparent)

**Syntax and Grammar Features**

a. Syntactic complexity and grammatical components are good; the participant uses compound sentences and a range of different grammatical structures.

b. Severity assessment:

- Simplified Sentence Structure and Grammar: 2/10 (The participant formulates complex sentences)
- Grammatical Errors: 2/10 (Few grammatical errors are observed)
- Difficulty with Complex Ideas: 3/10 (The participant appears to handle complex ideas relatively well)

**Semantics Features**

a. Semantic density and coherence are strong with most descriptions logically connected and contextually appropriate.

b. Severity assessment:

- Semantic Paraphasias: 1/10 (Very few incorrect word uses)
- Lack of Coherence: 2/10 (Participant presents a largely coherent narrative)
- Consistency in Errors: 2/10 (Few consistent errors)
- Lack of Semantic Fluency: 2/10 (The participant maintains a smooth flow of speech)
- Disorganized Narrative Structure: 2/10 (The narrative structure is relatively organized)
- Orientation Issues: 2/10 (The participant appears aware of the situation)
- Decline in Conversational Engagement and Responsiveness: 1/10 (Participant remains engaged and responsive)
- Memory Issues: 4/10 (Participant demonstrates some memory recall difficulty)
- Lack of Abstract Thinking and Synthesis: 2/10 (Participant can draw inferences and abstract ideas)
- Abnormal Emotions: 1/10 (No abnormal emotions are detected)

Based on this analysis, the participant could be classified into the **Probably Normal Cognition Group**. While there are minor issues such as hesitation, some repetition, and small memory recall difficulties, they do not appear to significantly impair communication or cognition. As always, this is not a formal diagnosis, and any concerns should be followed up with a healthcare provider
