## Supplementary material for "Text dialogue analysis Based ChatGPT for Primary Screening of Mild Cognitive Impairment": Data(GPT4_vs_GPT3.5_model1-5): 103_GPT4 (5).docx

ChatGPT

Assessing the participant based on their language features yields the following evaluations:

Vocabulary features: a. The participant’s vocabulary is relatively varied, but many unique words and phrases are repeated throughout the conversation. They frequently use filler words like "uh" and "um," which are common but could suggest some difficulty with word retrieval. The ratio of pronouns to specific names seems balanced. Lexical complexity seems to be moderate, without many complex or rare words. Severity Rating: 5/10. b. There are instances of the tip-of-the-tongue phenomenon where the participant stumbles over words or takes a moment to recall the appropriate term. This is evident from the use of fillers ("uh," "um") and pauses. Severity Rating: 6/10.

Syntax and grammar features: a. The syntactic complexity is relatively low. The participant typically constructs simple sentences with few clauses, and there are minimal uses of subordinate clauses. Their grammar usage is generally correct, with proper subject-verb agreement, tense consistency, and usage of prepositions and conjunctions. Severity Rating: 4/10. b. There seems to be no significant difficulty with complex ideas. The participant can articulate reasonably complex events like the story of George and his granddaughter. Still, they have some trouble when the narrative becomes more complex. Severity Rating: 5/10.

Semantics features: a. Semantic density is moderate. The participant frequently uses content words over function words. Their semantic coherence is mostly good. They seem to understand the stories they're telling or recounting and stay on topic. Severity Rating: 4/10. b. There is some indication of memory issues, with slight repetitions and small inconsistencies in story recall. However, the participant does remember significant plot points and characters. Severity Rating: 5/10.

In a comprehensive synthesis of these indicators, it's reasonable to categorize this participant in the "Probably Mild Cognitive Impairment Group." These issues might be present in individuals with normal cognition, and not all individuals with MCI will display all of these features. However, considering the occasional word-finding difficulties, slight repetitions, and moderate difficulty with complex ideas, this participant may have mild cognitive impairment. Please note that this is not a formal diagnosis and further investigation is needed to confirm this
