## Supplementary material for "Text dialogue analysis Based ChatGPT for Primary Screening of Mild Cognitive Impairment": Data(GPT4_vs_GPT3.5_model1-5): 103_GPT4.docx

ChatGPT

Vocabulary features:

- Word frequency: 160 unique words
- Word ratio: 160 unique words / 257 total words = 0.62257
- Phrase ratio: difficult to assess without repetitive text
- Difficulty finding the right words: Participant occasionally used filler words like "uh" and "um" but not excessively.
- Lexical complexity: Moderate; the text includes a mix of simple and more sophisticated words.
- Limited vocabulary: Some repetition and simple words, but overall vocabulary seems sufficient.

Semantics features:

- Semantic density: Moderate; the text contains a mix of content and function words.
- Inability to convey meaning: Participant expresses ideas clearly, with no significant confusion or misunderstanding.
- Semantic coherence: Generally coherent, with relationships between words, phrases, and sentences evident.
- Lack of coherence: Participant maintains a logical flow of ideas, with only minor disjointedness.

Syntax and Grammar features:

- Syntactic complexity: Moderate complexity, with a mix of simple and complex sentence structures.
- Simplified sentence structure: A mix of short, simple sentences and more complex structures.
- Grammatical components: Generally correct usage of grammar, with proper subject-verb agreement, tense consistency, and proper use of pronouns, prepositions, and conjunctions.
- Grammatical errors: Minimal errors, mainly in the form of dropped "g"s in words like "going" and "standing."

Based on the analysis above, I would assign PARTICIPANT to the Probably Normal group. The participant's language is generally coherent and clear, with only minor difficulties in finding the right words and some simplification of vocabulary and sentence structure
