## Supplementary material for "Text dialogue analysis Based ChatGPT for Primary Screening of Mild Cognitive Impairment": Data(GPT4_vs_GPT3.5_model1-5): 104_GPT3.5 (2).docx

Attention! You do not need to diagnose, just follow the steps below to initially screen for PARTICIPANT with possibly mild cognitive impairment (MCI). Some differences between MCI and normal cognition are also given, please analyse them in detail, step by step. Firstly, please assess the Vocabulary features of PARTICIPANT at the Word frequency and Word ratio, Phrase frequency and Phrase ratio, Word-finding difficulties, Repetition, Lexical complexity, Limited vocabulary. Word frequency: Count the occurrences of each unique word in the text. Unique words are distinct words that appear in the text, not taking into account their repetitions. Each word is counted only once, regardless of how many times it occurs. Word ratio: Calculate the proportion of unique words to the total number of words in the text. Phrase frequency: Count the occurrences of each unique phrases in the text. Unique phrases are distinct combinations of words that appear in the text, not taking into account their repetitions. Phrase ratio: Calculate the proportion of unique phrases to the total number of phrases in the text. Word-finding difficulties: Frequent pauses or use of filler words, like "uh" or "um,". MCI: more frequent instances, longer time to find the right word, or using more general terms. Normal: fewer instances, and better lexical retrieval. Repetition: The participant repeats certain phrases or ideas a few times throughout the conversation. MCI: more frequent repetition. Normal: less frequent repetition (some repetition of words but not excessive) and better memory of what has been mentioned. Lexical complexity: Measure the variety and sophistication of the vocabulary used, considering factors, including word length and frequency of rare words. Limited vocabulary: Repeated use of the same words or simple words. MCI: slightly reduced vocabulary, and more frequent use of simpler words or common terms; Normal: richer vocabulary (low limitation). Secondly, please assess the Semantics features of PARTICIPANT, from Semantic density and Inability to convey meaning, Semantic coherence and Lack of coherence. Semantic density: Measure the amount of meaningful information in the text, and considering factor is the number of content words (nouns, verbs, adjectives, adverbs) relative to function words (prepositions, pronouns, conjunctions, etc.). Inability to convey meaning: Difficulty in expressing ideas clearly, leading to confusion or misunderstanding. Normal: the participant was able to describe the scene clearly. Semantic coherence: Assess the logical consistency and meaningfulness of the text, which can be done by analyzing the relationships between words, phrases, and sentences, and checking for coherence and cohesion. Lack of coherence: Difficulty in maintaining a logical flow of ideas, with thoughts appearing disjointed or unrelated. MCI: struggle with organizing thoughts coherently or maintaining a clear focus in conversation; Normal: greater coherence and organization in their storytelling and conversations. Thirdly, please assess the Syntax and Grammar features of PARTICIPANT, from Syntactic complexity and Simplified sentence structure, Grammatical components and Grammatical errors. Syntactic complexity: Measure the complexity of the sentence structure in the text, considering factors such as the number of clauses per sentence, the average length of sentences, and the use of subordinate clauses. Simplified sentence structure: A lack of complex sentence structures. MCI: simpler sentence structures; Normal: in longer dialogues there is a greater variety of sentence structures (moderately complex), but in short dialogues the sentences may turn out to be simple. Grammatical components: Analyze the text for correct usage of grammar, such as subject-verb agreement, tense consistency, and proper use of pronouns, prepositions, and conjunctions. MCI: avoidance of complex grammatical constructions; Normal: better use of complex grammatical constructions. Grammatical errors: Frequent mistakes in verb tense, subject-verb agreement, or other aspects of grammar. MCI: more frequent grammatical errors, such as mistakes in verb tense and subject-verb agreement; Normal: fewer grammatical errors (a few errors, but not significant). Finally, Please dichotomise PARTICIPANT into the Probably Normal group or the Probably Mild Cognitive Impairment group. PARTICIPANT with MCI might exhibit more difficulty with vocabulary, sentence structure, repetition, word-finding, coherence, and grammar. However, it is not definitive and requires a comprehensive judgement. Now, you do not have to respond to this dialogue. When I have entered THE DIALOGUE BETWEEN PARTICIPANT AND INVESTIGATOR, evaluate PARTICIPANT according to the evaluation rules above. INVESTIGATOR: okay can you just tell me everything that you see going on in this picture ? PARTICIPANT: …um there's a woman standing at the sink , drying a dish . PARTICIPANT: and the water's overflowing onto the floor . PARTICIPANT: and there's two children , a boy and a girl „ it appears to be . PARTICIPANT: of_course I guess you're not allowed to talk about what sex people are anymore but anyway … PARTICIPANT: …um I don't know what they're getting into . PARTICIPANT: …um looks like a dish is falling . PARTICIPANT: oh it's probably the lid . PARTICIPANT: the lid … they took the lid off . PARTICIPANT: and it's … they're getting into the cookie jar . PARTICIPANT: and the …uh boy has a cookie in his hand . PARTICIPANT: and the …uh girl is reaching up to get the cookie . PARTICIPANT: I don't know . PARTICIPANT: does she have something in her hand ? PARTICIPANT: trying to see . PARTICIPANT: it looks . PARTICIPANT: I don't know what's in her hand but she's got her one hand …uh under her chin . PARTICIPANT: …uh what else is going on , anything ? PARTICIPANT: …uh mom is looking out the window and not paying attention to anything apparently . PARTICIPANT: …uh and there's a cup and saucer on the counter , and a plate . PARTICIPANT: that's all I can see for now . INVESTIGATOR: okay . INVESTIGATOR: so this picture tells a story . INVESTIGATOR: so what I'd like you to do is tell me a story about what you see . INVESTIGATOR: and that story should have a beginning a middle and an end . PARTICIPANT: okay . PARTICIPANT: …um looks like it's a beautiful summer day . PARTICIPANT: and …uh the little girl looks like she's lost her cat that's up in the tree . PARTICIPANT: and a gentleman went up the tree . PARTICIPANT: …uh I don't know if the dog chased him up into the tree or the dog chased the cat up in the tree . PARTICIPANT: but …ei either one , dad is stuck in the tree , it appears . PARTICIPANT: and the firemen are coming to help get the cat and the man out_of the tree . PARTICIPANT: and there's a bird in the tree . PARTICIPANT: so …um that's … that's all I can think of to say about that picture . INVESTIGATOR: yeah so can you tell me a story about what you see that has a beginning a middle and an end ? PARTICIPANT: …um well obviously this is …uh a story probably from the fifties from the looks of this car . PARTICIPANT: and …uh there are no seat belts or car seats for the kids . PARTICIPANT: and they're going somewhere in the vehicle . PARTICIPANT: …um then they have three kids . PARTICIPANT: looks like grandma's next_to the one kid in the back seat . PARTICIPANT: and the … you_know presumably the …pa …uh parents . PARTICIPANT: and …uh the little boy looks excited about the trip and so does the dog . PARTICIPANT: and he's got … both of them have their heads out the window . PARTICIPANT: …um and …uh everybody looks pretty happy in that first picture . PARTICIPANT: but then when you go to the one down below , it looks like somebody mighta told the kids to behave and get theirselves ( themselves ) back in the window . PARTICIPANT: because … and dad doesn't look real thrilled in that picture . PARTICIPANT: and mom's sleeping . PARTICIPANT: so maybe that's the way back . PARTICIPANT: or they turned around and said . PARTICIPANT: forget this trip with these brats . PARTICIPANT: that's all I can say . INVESTIGATOR: thank_you . INVESTIGATOR: are you familiar with the story of Cinderella ? PARTICIPANT: of_course . INVESTIGATOR: can you tell me the story of Cinderella ? INVESTIGATOR: or I can show you some pictures to jog your memory . PARTICIPANT: well INVESTIGATOR: whichever one you'd like . PARTICIPANT: …um well there's … I think there's several versions . PARTICIPANT: and I've seen several versions &INVESTIGATOR:mhm you_know in movies and stuff . PARTICIPANT: …um do you want the original ? PARTICIPANT: because then I might need the pictures if you want me to do the original version . PARTICIPANT: but I think . PARTICIPANT: I don't know . PARTICIPANT: go ahead . INVESTIGATOR: do you wanna ? INVESTIGATOR: they're not the … …or they're not the original version pictures . INVESTIGATOR: they're the Walt_Disney version . PARTICIPANT: oh okay . PARTICIPANT: …um well I guess I can go through it . PARTICIPANT: …um …uh Cinderella …um was the daughter of …um a king , I guess it was . PARTICIPANT: and …um something happened to the first wife . PARTICIPANT: I'm assuming she died , maybe in childbirth . PARTICIPANT: and he remarried . PARTICIPANT: and …um I believe the stepmother killed the … her husband , if I remember right . PARTICIPANT: and …um she … she had two daughters . PARTICIPANT: or was ? PARTICIPANT: it's two , right ? PARTICIPANT: or three . PARTICIPANT: two or three daughters . PARTICIPANT: and …um she and the daughters treated Cinderella badly and didn't treat her like the princess she was . PARTICIPANT: they treated her like she was …uh their personal slave . PARTICIPANT: and …uh they were … all … all the daughters were gonna go to …um a special ball at the palace …um to meet the prince . PARTICIPANT: and …um so the girl … …uh the three stepsisters all got …uh gussied up and …uh were preparing to go . PARTICIPANT: and Cinderella of_course was not allowed to go .

ChatGPT

3 / 3

Based on the provided dialogue, let's analyze the participant's performance according to the evaluation criteria:

Vocabulary Features:

- Word frequency: The participant uses a variety of words, but there are instances of repetition (e.g., "um," "and," "the"). There are also some filler words and pauses.
- Word ratio: It is difficult to determine the word ratio without the exact word count, but the participant shows some repetition, which may affect the ratio.
- Phrase frequency: The participant uses a mix of phrases, but there are instances of repetition (e.g., "and the," "I don't know").
- Phrase ratio: Similar to the word ratio, the phrase ratio is challenging to determine without the exact count.

Word-finding Difficulties and Repetition:

- The participant exhibits some word-finding difficulties, as seen in the frequent pauses, filler words, and use of general terms.
- There is repetition of phrases and ideas throughout the conversation, such as mentioning the lid, the cookie jar, and the girl's hand.

Lexical Complexity and Limited Vocabulary:

- The participant's vocabulary is adequate, but there are instances of using simpler words and common terms. Some phrases are repeated, indicating a limited vocabulary.

Semantics Features:

- Semantic density: The dialogue contains a mix of content words (nouns, verbs, adjectives, adverbs) and function words (prepositions, pronouns, conjunctions). However, without an exact count, it is challenging to evaluate the semantic density accurately.
- Inability to convey meaning: The participant sometimes struggles to express ideas clearly, resulting in confusion or uncertainty.
- Semantic coherence: The participant's storytelling lacks logical consistency and meaningfulness. Thoughts appear disjointed and unrelated.

Syntax and Grammar Features:

- Syntactic complexity: The sentence structure used by the participant is mostly simple, with straightforward constructions and a lack of complex sentence structures.
- Simplified sentence structure: The participant tends to use simpler sentence structures, avoiding complex constructions.
- Grammatical components: The participant generally demonstrates a reasonable use of grammar, but there are occasional errors, such as subject-verb agreement and pronoun usage.
- Grammatical errors: Some grammatical errors occur in the dialogue, such as mistakes in verb tense and subject-verb agreement.

Based on the evaluation of the participant's performance, the observations suggest possible signs of mild cognitive impairment (MCI). The participant shows difficulties with vocabulary, word-finding, coherence, and grammar, as well as exhibiting repetition and limited vocabulary. However, it is important to note that this evaluation is preliminary, and a comprehensive judgment would require further assessment and professional expertise.
