## Supplementary material for "Text dialogue analysis Based ChatGPT for Primary Screening of Mild Cognitive Impairment": Data(GPT4_vs_GPT3.5_model1-5): 104_GPT3.5 (3).docx

ChatGPT

Based on the analysis of the participant's language in the provided conversation, here are the evaluation indicators for screening mild cognitive impairment (MCI) and their severity ratings:

Vocabulary features:

1. Limited vocabulary: Severity: 3 - The participant demonstrates a slightly reduced vocabulary and uses simple words or common terms.
2. Increased verbosity: Severity: 2 - The participant uses more words than necessary and tends to be more verbose.
3. Overlearned phrases: Severity: 1 - No evidence of overuse of cliches or stock phrases.
4. Improper use of pronouns: Severity: 1 - No difficulty or mixing up of pronouns observed.
5. Hesitation (Word-finding difficulties): Severity: 2 - The participant exhibits some word-finding difficulties with hesitations and filler words.
6. Circumlocution (Word-finding difficulties): Severity: 1 - No evidence of circumlocution or deliberate vagueness observed.
7. Tip-of-the-tongue phenomenon: Severity: 2 - The participant experiences partial recall and difficulty retrieving specific words.
8. Repetition: Severity: 1 - No excessive repetition observed.

Syntax and Grammar features:

1. Simplified sentence structure and grammar: Severity: 1 - No preference for simpler structures or avoidance of complex grammatical constructions observed.
2. Grammatical errors: Severity: 1 - No significant grammatical errors observed.
3. Difficulty with complex ideas: Severity: 1 - No significant difficulty with complex ideas observed.

Semantics features:

1. Semantic paraphasias: Severity: 1 - No errors in word choice or odd combinations of words observed.
2. Lack of coherence: Severity: 1 - The participant demonstrates coherence and organization in their storytelling and conversations.
3. Consistency in errors: Severity: 1 - No consistent language errors observed.
4. Lack of semantic fluency: Severity: 1 - No significant difficulty in naming items or describing processes observed.
5. Disorganized narrative structure: Severity: 1 - No significant difficulty in maintaining a coherent line of thought observed.
6. Orientation issues: Severity: 1 - No confusion about date, time, place, or sequence of events observed.
7. Decline in conversational engagement and responsiveness: Severity: 2 - The participant exhibits decreased initiation, difficulties in following and responding to conversation.
8. Memory issues: Severity: 1 - No significant memory issues observed.
9. Lack of abstract thinking and synthesis: Severity: 1 - No significant difficulty in understanding or expressing abstract concepts observed.
10. Abnormal emotions: Severity: 1 - No significant changes in tone, initiative, or abnormal emotions observed.

Based on the synthesis of the evaluation indicators, the participant's language features do not strongly indicate mild cognitive impairment (MCI). The indicators suggest mostly normal cognitive function with minor instances of word-finding difficulties and decreased conversational engagement. However, it's important to note that this assessment is based solely on the provided conversation and a comprehensive evaluation should involve multiple assessment methods and consideration of other factors
