## Supplementary material for "Text dialogue analysis Based ChatGPT for Primary Screening of Mild Cognitive Impairment": Data(GPT4_vs_GPT3.5_model1-5): 104_GPT3.5 (4).docx

ChatGPT

2 / 2

Let's analyze the participant's language features based on the given conversation. We'll rate the severity of each evaluation indicator on a scale of 1 to 10, with 1 being the least severe and 10 being the most severe.

Vocabulary features: a. Word frequency, word ratio, phrase frequency, and phrase ratio: Not assessed in the provided conversation. b. Lexical complexity: Not assessed in the provided conversation. c. Severity ratings for different aspects:

- Limited vocabulary: 3
- Increased verbosity: 4
- Overlearned phrases: 2
- Improper use of pronouns: 3
- Hesitation: 4
- Circumlocution: 2
- Tip-of-the-tongue phenomenon: 3
- Repetition: 2

Syntax and grammar features: a. Syntactic complexity and grammatical components: Not assessed in the provided conversation. b. Severity ratings for different aspects:

- Simplified sentence structure and grammar: 2
- Grammatical errors: 2
- Difficulty with complex ideas: 2

Semantics features: a. Semantic density and coherence: Not assessed in the provided conversation. b. Severity ratings for different aspects:

- Semantic paraphasias: Not observed
- Lack of coherence: 2
- Consistency in errors: 2
- Lack of semantic fluency: 2
- Disorganized narrative structure: 2
- Orientation issues: Not observed
- Decline in conversational engagement and responsiveness: Not observed
- Memory issues: Not observed
- Lack of abstract thinking and synthesis: 2
- Abnormal emotions: Not observed

Based on the assessment of the given indicators, we can categorize the participant into the Probably Normal Cognition Group. While there are some mild language features that suggest possible mild cognitive impairment, the overall severity ratings for the evaluated indicators are relatively low. It's important to note that this assessment is based on a single conversation and further evaluation may be necessary for a comprehensive understanding of the participant's cognitive functioning.
