## Supplementary material for "Text dialogue analysis Based ChatGPT for Primary Screening of Mild Cognitive Impairment": Data(GPT4_vs_GPT3.5_model1-5): 104_GPT3.5 (5).docx

ChatGPT

Based on the language features exhibited by the participant, we can evaluate the severity of indicators related to possible mild cognitive impairment (MCI). Please note that this assessment is based solely on the provided conversation and is not a definitive diagnosis. It is important to consider other factors and conduct a thorough evaluation for a comprehensive assessment. Here is the analysis and severity rating for each evaluation indicator:

Vocabulary Features: a. Word Frequency and Word Ratio: Not provided in the conversation. b. Tip-of-the-Tongue Phenomenon: 4/10 - The participant exhibits some instances of partial recall, such as not being able to recall specific details and struggling to find the right words.

Syntax and Grammar Features: a. Syntactic Complexity and Grammatical Components: Not provided in the conversation. b. Difficulty with Complex Ideas: 3/10 - The participant demonstrates occasional difficulty with more complex ideas, as seen when the narrative or question becomes more intricate.

Semantics Features: a. Semantic Density and Coherence: Not provided in the conversation. b. Memory Issues: 5/10 - The participant shows some signs of memory issues, including repeating questions and having difficulty recalling specific details of the story or plot points.

Overall Assessment:

Based on the limited conversation provided, it is difficult to make a definitive classification. However, considering the severity ratings of the indicators, the participant shows mild impairments in vocabulary (tip-of-the-tongue phenomenon), difficulty with complex ideas, and memory issues. There is insufficient information to evaluate the remaining indicators.

Given the preliminary evaluation, the participant can be tentatively categorized into the Probably Mild Cognitive Impairment Group. However, it is crucial to conduct a comprehensive assessment involving other cognitive domains and evaluations to make a more accurate determination.
