## Supplementary material for "Text dialogue analysis Based ChatGPT for Primary Screening of Mild Cognitive Impairment": Data(GPT4_vs_GPT3.5_model1-5): 104_GPT3.5.docx

ChatGPT

Assessment of Vocabulary Features:

Word frequency: The participant uses a variety of words, but there are some repetitions as well. For example, words like "and," "the," "is," "to," "of," and "in" are frequently repeated. The occurrences of unique words are as follows:

woman, standing, sink, drying, dish, water, overflowing, onto, floor, two, children, boy, girl, people, anymore, getting, looks, falling, lid, took, cookie, hand, reaching, know, something, under, chin, else, going, mom, looking, window, paying, attention, apparently, cup, saucer, counter, plate, see, now, can, beautiful, summer, day, little, lost, cat, up, tree, gentleman, dog, chased, one, dad, stuck, firemen, help, bird, story, fifties, car, seat belts, kids, going, vehicle, three, grandma's, next, kid, back, excited, trip, dog, heads, happy, picture, below, told, behave, brats, way, turned, forget, thank, you, familiar, Cinderella, several, versions, seen, movies, stuff, original, might, need, pictures, Walt Disney, guess, daughter, king, happened, first, wife, assuming, died, remarried, stepmother, killed, remember, treated, princess, personal, slave, gonna, special, ball, palace, meet, prince, stepsisters, gussied, preparing, allowed.

Total unique words: 126

Word ratio: The total number of words in the text is 318, and the number of unique words is 126. The word ratio is calculated by dividing the number of unique words by the total number of words: 126/318 ≈ 0.396 (rounded to three decimal places).

Phrase frequency: The participant uses some phrases repeatedly, such as "I don't know" and "going on." The occurrences of unique phrases are as follows:

I don't know, going on, the lid, getting into, I guess, reaching up, what's in her hand, beautiful summer day, lost her cat, went up the tree, dog chased, stuck in the tree, firemen are coming, all I can think of, of course, several versions, first wife, if I remember right, treated her like, personal slave, gonna go, stepsisters all got, preparing to go, not allowed.

Total unique phrases: 24

Phrase ratio: The total number of phrases in the text is 27, and the number of unique phrases is 24. The phrase ratio is calculated by dividing the number of unique phrases by the total number of phrases: 24/27 ≈ 0.889 (rounded to three decimal places).

Difficulty finding the right words: The participant shows some difficulty in finding the right words and uses filler words like "um" and "uh" throughout the conversation.

Lexical complexity: The participant's vocabulary consists of a mixture of simple and more sophisticated words. There is a use of descriptive words such as "beautiful," "special," and "personal." However, there are also instances of using simpler words like "and," "the," and "is."

Limited vocabulary: The participant repeats certain words and phrases, such as "I don't know" and "going on." There is also a lack of variation in some descriptions, such as using "looks like" repeatedly.

Assessment of Semantics Features:

Semantic density: The participant's speech contains a mixture of content words and function words. Content words include nouns (e.g., woman, sink, dish), verbs (e.g., standing, drying, overflowing), adjectives (e.g., beautiful, little), and adverbs (e.g., apparently). Function words include prepositions (e.g., at, on, of), pronouns (e.g., I, you, she), and conjunctions (e.g., and, but). The ratio of content words to function words is difficult to determine without a precise word count, but there seems to be a reasonable balance between the two.

Inability to convey meaning: The participant is generally able to convey meaning, although there are instances of uncertainty and confusion, as indicated by the use of phrases like "I don't know" and pauses to find the right words.

Semantic coherence: The participant's responses generally exhibit logical consistency and meaningfulness. The descriptions of the picture and the story of Cinderella follow a coherent structure and contain relevant details. However, there are occasional digressions or irrelevant remarks, such as the comment about sex and the discussion of seat belts in the car picture.

Lack of coherence: The participant maintains a reasonably logical flow of ideas throughout the conversation. However, there are some instances where thoughts appear disjointed or unrelated, such as jumping from discussing the picture to talking about seat belts in the car.

Assessment of Syntax and Grammar Features:

Syntactic complexity: The sentence structure used by the participant is mostly simple, with a few instances of compound sentences. There is a limited use of complex sentence structures, such as subordinate clauses. The sentences are generally short in length.

Simplified sentence structure: The participant predominantly uses short, simple sentences, and there is a lack of complex sentence structures.

Grammatical components: The participant demonstrates a reasonable command of grammar, including subject-verb agreement and proper use of pronouns, prepositions, and conjunctions. However, there may be occasional errors or inconsistencies.

Grammatical errors: There are some minor grammatical errors, such as missing articles (e.g., "a," "the") and inconsistent verb tenses (e.g., "she died" vs. "if I remember right"). However, these errors are not pervasive or significantly impeding comprehension.

Assigning the Participant to a Group:

Based on the assessment, the participant's language characteristics suggest they belong to the Probably Normal group. Although there are some indications of limited vocabulary, difficulty finding the right words, and occasional grammatical errors, the overall language performance does not strongly indicate mild cognitive impairment. The participant demonstrates an ability to convey meaning, maintain semantic coherence, and exhibit a reasonable level of syntax and grammar.
