## Supplementary material for "Text dialogue analysis Based ChatGPT for Primary Screening of Mild Cognitive Impairment": Data(GPT4_vs_GPT3.5_model1-5): 104_GPT4 (2).docx

INVESTIGATOR: okay can you just tell me everything that you see going on in this picture ? PARTICIPANT: …um there's a woman standing at the sink , drying a dish . PARTICIPANT: and the water's overflowing onto the floor . PARTICIPANT: and there's two children , a boy and a girl „ it appears to be . PARTICIPANT: of_course I guess you're not allowed to talk about what sex people are anymore but anyway … PARTICIPANT: …um I don't know what they're getting into . PARTICIPANT: …um looks like a dish is falling . PARTICIPANT: oh it's probably the lid . PARTICIPANT: the lid … they took the lid off . PARTICIPANT: and it's … they're getting into the cookie jar . PARTICIPANT: and the …uh boy has a cookie in his hand . PARTICIPANT: and the …uh girl is reaching up to get the cookie . PARTICIPANT: I don't know . PARTICIPANT: does she have something in her hand ? PARTICIPANT: trying to see . PARTICIPANT: it looks . PARTICIPANT: I don't know what's in her hand but she's got her one hand …uh under her chin . PARTICIPANT: …uh what else is going on , anything ? PARTICIPANT: …uh mom is looking out the window and not paying attention to anything apparently . PARTICIPANT: …uh and there's a cup and saucer on the counter , and a plate . PARTICIPANT: that's all I can see for now . INVESTIGATOR: okay . INVESTIGATOR: so this picture tells a story . INVESTIGATOR: so what I'd like you to do is tell me a story about what you see . INVESTIGATOR: and that story should have a beginning a middle and an end . PARTICIPANT: okay . PARTICIPANT: …um looks like it's a beautiful summer day . PARTICIPANT: and …uh the little girl looks like she's lost her cat that's up in the tree . PARTICIPANT: and a gentleman went up the tree . PARTICIPANT: …uh I don't know if the dog chased him up into the tree or the dog chased the cat up in the tree . PARTICIPANT: but …ei either one , dad is stuck in the tree , it appears . PARTICIPANT: and the firemen are coming to help get the cat and the man out_of the tree . PARTICIPANT: and there's a bird in the tree . PARTICIPANT: so …um that's … that's all I can think of to say about that picture . INVESTIGATOR: yeah so can you tell me a story about what you see that has a beginning a middle and an end ? PARTICIPANT: …um well obviously this is …uh a story probably from the fifties from the looks of this car . PARTICIPANT: and …uh there are no seat belts or car seats for the kids . PARTICIPANT: and they're going somewhere in the vehicle . PARTICIPANT: …um then they have three kids . PARTICIPANT: looks like grandma's next_to the one kid in the back seat . PARTICIPANT: and the … you_know presumably the …pa …uh parents . PARTICIPANT: and …uh the little boy looks excited about the trip and so does the dog . PARTICIPANT: and he's got … both of them have their heads out the window . PARTICIPANT: …um and …uh everybody looks pretty happy in that first picture . PARTICIPANT: but then when you go to the one down below , it looks like somebody mighta told the kids to behave and get theirselves ( themselves ) back in the window . PARTICIPANT: because … and dad doesn't look real thrilled in that picture . PARTICIPANT: and mom's sleeping . PARTICIPANT: so maybe that's the way back . PARTICIPANT: or they turned around and said . PARTICIPANT: forget this trip with these brats . PARTICIPANT: that's all I can say . INVESTIGATOR: thank_you . INVESTIGATOR: are you familiar with the story of Cinderella ? PARTICIPANT: of_course . INVESTIGATOR: can you tell me the story of Cinderella ? INVESTIGATOR: or I can show you some pictures to jog your memory . PARTICIPANT: well INVESTIGATOR: whichever one you'd like . PARTICIPANT: …um well there's … I think there's several versions . PARTICIPANT: and I've seen several versions &INVESTIGATOR:mhm you_know in movies and stuff . PARTICIPANT: …um do you want the original ? PARTICIPANT: because then I might need the pictures if you want me to do the original version . PARTICIPANT: but I think . PARTICIPANT: I don't know . PARTICIPANT: go ahead . INVESTIGATOR: do you wanna ? INVESTIGATOR: they're not the … …or they're not the original version pictures . INVESTIGATOR: they're the Walt_Disney version . PARTICIPANT: oh okay . PARTICIPANT: …um well I guess I can go through it . PARTICIPANT: …um …uh Cinderella …um was the daughter of …um a king , I guess it was . PARTICIPANT: and …um something happened to the first wife . PARTICIPANT: I'm assuming she died , maybe in childbirth . PARTICIPANT: and he remarried . PARTICIPANT: and …um I believe the stepmother killed the … her husband , if I remember right . PARTICIPANT: and …um she … she had two daughters . PARTICIPANT: or was ? PARTICIPANT: it's two , right ? PARTICIPANT: or three . PARTICIPANT: two or three daughters . PARTICIPANT: and …um she and the daughters treated Cinderella badly and didn't treat her like the princess she was . PARTICIPANT: they treated her like she was …uh their personal slave . PARTICIPANT: and …uh they were … all … all the daughters were gonna go to …um a special ball at the palace …um to meet the prince . PARTICIPANT: and …um so the girl … …uh the three stepsisters all got …uh gussied up and …uh were preparing to go . PARTICIPANT: and Cinderella of_course was not allowed to go . PARTICIPANT: she hadta stay home and do her work and all that . PARTICIPANT: and in the Disney version they had her talking to the animals and all that you_know . PARTICIPANT: and singing , of_course . PARTICIPANT: in Disney movies you hafta have singing . PARTICIPANT: and …um so …uh the fairy godmother comes along and says she'll help out Cinderella so she can go to the …uh ball . PARTICIPANT: so she waves her magic wand and has a beautiful …uh dress for her . PARTICIPANT: and she uses the animals . PARTICIPANT: and she uses her wand and turns some of them into horses and …uh the carriages from a pumpkin , I believe . PARTICIPANT: and you_know so everything is all glamorous . PARTICIPANT: and she's going to make a real entrance when she gets there with her horse drawn carriage . PARTICIPANT: and …um of_course she's a_little late for the party . PARTICIPANT: …um and she made a grand entrance down the staircase and all that . PARTICIPANT: and she danced with the prince . PARTICIPANT: and the prince fell for her . PARTICIPANT: and … …um but she hadta be home by midnight . PARTICIPANT: and …uh so she hadta rush out suddenly . PARTICIPANT: and she lost her glass slipper . PARTICIPANT: and that's the only trace of her that was left behind . PARTICIPANT: and …uh you_know of_course she raced home .

ChatGPT

This is an interesting task, and the participant seems to have some areas where they might show signs of Mild Cognitive Impairment (MCI), but it's also possible they are simply demonstrating a normal range of cognitive function. Please note that only a professional can make a final diagnosis.

1. Vocabulary:
   - Word frequency: The participant uses a wide variety of words, showing a good command of vocabulary. But there are repetitive words like "um", "uh", "and", etc.
   - Word ratio: The proportion of unique words to total words appears to be within a normal range, although this would require a more exact calculation to confirm.
   - Phrase frequency: There are repetitive phrases like "and um".
   - Phrase ratio: The phrase ratio also seems normal, with a good variety of phrases used.
   - Word-finding difficulties: The participant frequently uses "um" and "uh", which might indicate some word-finding difficulties.
   - Repetition: There are instances of repetition but not excessively so.
   - Lexical complexity: The participant uses a diverse range of words and complex sentences, indicating good lexical complexity.
   - Limited vocabulary: The participant does not seem to have a limited vocabulary.
2. Semantics:
   - Semantic density: The participant's speech is rich in content words, indicating high semantic density.
   - Inability to convey meaning: Despite some pauses and filler words, the participant generally conveys their ideas clearly.
   - Semantic coherence: The participant's speech is coherent and logically consistent.
   - Lack of coherence: There are no significant indications of a lack of coherence.
3. Syntax and Grammar:
   - Syntactic complexity: The participant uses complex sentence structures with multiple clauses and different types of conjunctions.
   - Simplified sentence structure: There is no clear evidence of overly simplified sentence structure.
   - Grammatical components: The participant correctly uses various grammatical structures.
   - Grammatical errors: There are minor grammatical errors, but they are not significant or frequent.

In conclusion, based on this dialogue, the participant seems to fall into the "Probably Normal" category. They show some signs of possible MCI, such as word-finding difficulties and minor grammatical errors, but overall, their speech is coherent, logically consistent, and shows good use of vocabulary, syntax, and grammar. However, a more comprehensive examination would be necessary for a definitive conclusion.
