## Supplementary material for "Text dialogue analysis Based ChatGPT for Primary Screening of Mild Cognitive Impairment": Data(GPT4_vs_GPT3.5_model1-5): 104_GPT4 (3).docx

ChatGPT

Based on the conversation provided, here are my preliminary analysis and evaluation indicators:

Vocabulary Analysis:

- Word Frequency and Word Ratio: The participant uses a range of vocabulary, although there are some commonly repeated words such as "and", "um", "the", "looks like", "I don't know". Some pronouns and specific names are present in normal ratios.
- Phrase Frequency and Phrase Ratio: The phrases are varied and do not seem to be repeated excessively.
- Lexical Complexity: The participant uses a mix of simple and complex words, showing a balanced lexical complexity.

Vocabulary Evaluation:

1. Limited Vocabulary: 3/10 - A few instances of repeated words and phrases, but overall vocabulary seems fairly diverse.
2. Increased Verbosity: 5/10 - The participant provides thorough descriptions, but occasionally includes more information than necessary.
3. Overlearned Phrases: 2/10 - There are a few phrases that are repeated, but not to an excessive extent.
4. Improper Use of Pronouns: 2/10 - Mostly correct use of pronouns.
5. Hesitation (Word-finding difficulties): 7/10 - Frequent use of filler words such as "um" and "uh".
6. Circumlocution (Word-finding difficulties): 4/10 - The participant occasionally uses more words than necessary.
7. Tip-of-the-tongue phenomenon: 4/10 - Occasional difficulty finding the right word.
8. Repetition: 3/10 - Some repetition of words and phrases, but not excessively.

Syntax and Grammar Analysis:

- Syntactic Complexity: The participant uses a mix of simple and complex sentences.
- Grammatical Components: The participant generally uses correct grammar, with occasional errors.

Syntax and Grammar Evaluation:

1. Simplified Sentence Structure and Grammar: 4/10 - Occasional preference for simpler structures, but also uses complex structures.
2. Grammatical Errors: 2/10 - Few grammatical errors.
3. Difficulty with Complex Ideas: 3/10 - The participant generally seems capable of handling complex ideas, although there are occasional difficulties.

Semantics Analysis:

- Semantic Density: The participant uses a mixture of content and function words, showing a balanced semantic density.
- Semantic Coherence: The participant maintains a generally coherent narrative, although there are occasional lapses.

Semantics Evaluation:

1. Semantic Paraphasias: 3/10 - Few instances of difficulty in expressing ideas clearly.
2. Lack of Coherence: 3/10 - Occasionally struggles with maintaining a clear focus.
3. Consistency in Errors: 2/10 - Few consistent language errors.
4. Lack of Semantic Fluency: 3/10 - Occasional difficulties in describing processes or sequences.
5. Disorganized Narrative Structure: 3/10 - Occasionally struggles with maintaining a coherent narrative structure.
6. Orientation Issues: 2/10 - Few issues with orientation.
7. Decline in Conversational Engagement and Responsiveness: 3/10 - Some instances of reduced spontaneity and engagement.
8. Memory Issues: 2/10 - Few noticeable memory issues.
9. Lack of Abstract Thinking and Synthesis: 3/10 - Occasional difficulties with abstract concepts.
10. Abnormal Emotions: 2/10 - Few signs of abnormal emotions.

Based on this preliminary analysis, the participant might fall into the Probably Normal Cognition Group, with some indications of possible Mild Cognitive Impairment, particularly in terms of frequent hesitation and occasional difficulties with word-finding, maintaining coherence, and handling complex ideas.
