## Supplementary material for "Text dialogue analysis Based ChatGPT for Primary Screening of Mild Cognitive Impairment": Data(GPT4_vs_GPT3.5_model1-5): 104_GPT4 (4).docx

ChatGPT

**Vocabulary features:**

a. The participant uses a variety of words and phrases, suggesting good vocabulary diversity. Word and phrase frequencies do not seem overly high. b. Lexical complexity appears average; the participant uses a mix of simple and complex words. c. Severity of issues:

- Limited vocabulary: 2/10
- Increased verbosity: 3/10
- Overlearned phrases: 2/10
- Improper use of pronouns: 1/10
- Hesitation: 4/10
- Circumlocution: 3/10
- Tip-of-the-tongue phenomenon: 2/10
- Repetition: 2/10

**Syntax and grammar features:**

a. The syntactic complexity is average. The participant uses a mix of simple and complex sentences. b. Severity of issues:

- Simplified sentence structure and grammar: 2/10
- Grammatical errors: 1/10
- Difficulty with complex ideas: 3/10

**Semantics features:**

a. The participant maintains good semantic density and coherence. b. Severity of issues:

- Semantic paraphasias: 2/10
- Lack of coherence: 2/10
- Consistency in errors: 2/10
- Lack of semantic fluency: 3/10
- Disorganized narrative structure: 2/10
- Orientation issues: 2/10
- Decline in conversational engagement and responsiveness: 2/10
- Memory issues: 2/10
- Lack of abstract thinking and synthesis: 2/10
- Abnormal emotions: 1/10

Based on the comprehensive synthesis of all the indicators, this participant would likely fall into the **Probably Normal Cognition Group**. It's crucial to remember that this analysis does not diagnose MCI but provides an initial evaluation of cognitive function based on linguistic features. It's recommended that the participant consults a healthcare provider if cognitive issues persist or worsen.
