## Supplementary material for "Text dialogue analysis Based ChatGPT for Primary Screening of Mild Cognitive Impairment": Data(GPT4_vs_GPT3.5_model1-5): 104_GPT4 (5).docx

Please assess a participant with possible mild cognitive impairment (MCI) based on their language features. Note that I do not need to diagnose the participant but rather analyze and rate the severity of each evaluation indicator on a scale of 1 to 10 (1 being the least severe, and 10 being the most severe). Vocabulary features: a. Analyze word frequency, word ratio, phrase frequency, and phrase ratio. And calculate the lexical complexity. ( Word frequency and Word ratio: Count the occurrences of each unique word, each pronoun, and each specific name (or noun) in the text. And calculate the proportion of unique words to the total number of words in the text, and calculate the ratio of pronouns to specific names (or nouns). Phrase frequency and Phrase ratio: Count the occurrences of each unique phrases in the text. And calculate the proportion of unique phrases to the total number of phrases in the text. Lexical complexity: Measure the variety and sophistication of the vocabulary used, considering factors, including word length and frequency of rare words. ) b. Assess and rate the severity of the following aspects: tip-of-the-tongue phenomenon. (This phenomenon is characterized by partial recall. For instance, a person might remember the first letter of a word, its syllable structure, or its general semantic category, without being able to recall the whole word. More common in individuals with MCI.) Syntax and grammar features: a. Analyze syntactic complexity and grammatical components. (Syntactic complexity: Measure the complexity of the sentence structure in the text, considering factors such as the number of clauses per sentence, the average length of sentences, and the use of subordinate clauses. Grammatical components: Analyze the text for correct usage of grammar, such as subject-verb agreement, tense consistency, and proper use of pronouns, prepositions, and conjunctions.) b. Assess and rate the severity of the following aspects: difficulty with complex ideas (MCI: while the participant could handle simpler sentences and ideas, they had more difficulty when the narrative or the question became more complex.) Semantics features: a. Analyze semantic density and coherence. (Semantic density: Measure the amount of meaningful information in the text, and considering factor is the number of content words (nouns, verbs, adjectives, adverbs) relative to function words (prepositions, pronouns, conjunctions, etc.). Semantic coherence: Assess the logical consistency and meaningfulness of the text, which can be done by analyzing the relationships between words, phrases, and sentences, and checking for coherence and cohesion.) b. Assess and rate the severity of the following aspects: memory issues (MCI: repeat questions, difficulty in recalling certain information or sequences, difficulty in remembering specific details of the story, or difficulty in recalling major plot points or characters. However, memory issues can also be a part of normal aging, so this sign should be evaluated in the context of the individual's overall cognitive function.) Based on the comprehensive synthesis of all the indicators, classify the participant into either the Probably Normal Cognition Group or the Probably Mild Cognitive Impairment Group. Keep in mind that these issues might be present in individuals with normal cognition and that not all individuals with MCI will display all of these features. The final categorization should be based on a comprehensive synthesis of all the indicators. Please note: This evaluation isn't meant to provide a formal diagnosis but to serve as an initial screening tool for potential MCI. INVESTIGATOR: okay can you just tell me everything that you see going on in this picture ? PARTICIPANT: …um there's a woman standing at the sink , drying a dish . PARTICIPANT: and the water's overflowing onto the floor . PARTICIPANT: and there's two children , a boy and a girl „ it appears to be . PARTICIPANT: of_course I guess you're not allowed to talk about what sex people are anymore but anyway … PARTICIPANT: …um I don't know what they're getting into . PARTICIPANT: …um looks like a dish is falling . PARTICIPANT: oh it's probably the lid . PARTICIPANT: the lid … they took the lid off . PARTICIPANT: and it's … they're getting into the cookie jar . PARTICIPANT: and the …uh boy has a cookie in his hand . PARTICIPANT: and the …uh girl is reaching up to get the cookie . PARTICIPANT: I don't know . PARTICIPANT: does she have something in her hand ? PARTICIPANT: trying to see . PARTICIPANT: it looks . PARTICIPANT: I don't know what's in her hand but she's got her one hand …uh under her chin . PARTICIPANT: …uh what else is going on , anything ? PARTICIPANT: …uh mom is looking out the window and not paying attention to anything apparently . PARTICIPANT: …uh and there's a cup and saucer on the counter , and a plate . PARTICIPANT: that's all I can see for now . INVESTIGATOR: okay . INVESTIGATOR: so this picture tells a story . INVESTIGATOR: so what I'd like you to do is tell me a story about what you see . INVESTIGATOR: and that story should have a beginning a middle and an end . PARTICIPANT: okay . PARTICIPANT: …um looks like it's a beautiful summer day . PARTICIPANT: and …uh the little girl looks like she's lost her cat that's up in the tree . PARTICIPANT: and a gentleman went up the tree . PARTICIPANT: …uh I don't know if the dog chased him up into the tree or the dog chased the cat up in the tree . PARTICIPANT: but …ei either one , dad is stuck in the tree , it appears . PARTICIPANT: and the firemen are coming to help get the cat and the man out_of the tree . PARTICIPANT: and there's a bird in the tree . PARTICIPANT: so …um that's … that's all I can think of to say about that picture . INVESTIGATOR: yeah so can you tell me a story about what you see that has a beginning a middle and an end ? PARTICIPANT: …um well obviously this is …uh a story probably from the fifties from the looks of this car . PARTICIPANT: and …uh there are no seat belts or car seats for the kids . PARTICIPANT: and they're going somewhere in the vehicle . PARTICIPANT: …um then they have three kids . PARTICIPANT: looks like grandma's next_to the one kid in the back seat . PARTICIPANT: and the … you_know presumably the …pa …uh parents . PARTICIPANT: and …uh the little boy looks excited about the trip and so does the dog . PARTICIPANT: and he's got … both of them have their heads out the window . PARTICIPANT: …um and …uh everybody looks pretty happy in that first picture . PARTICIPANT: but then when you go to the one down below , it looks like somebody mighta told the kids to behave and get theirselves ( themselves ) back in the window . PARTICIPANT: because … and dad doesn't look real thrilled in that picture . PARTICIPANT: and mom's sleeping . PARTICIPANT: so maybe that's the way back . PARTICIPANT: or they turned around and said . PARTICIPANT: forget this trip with these brats . PARTICIPANT: that's all I can say . INVESTIGATOR: thank_you . INVESTIGATOR: are you familiar with the story of Cinderella ? PARTICIPANT: of_course . INVESTIGATOR: can you tell me the story of Cinderella ? INVESTIGATOR: or I can show you some pictures to jog your memory . PARTICIPANT: well INVESTIGATOR: whichever one you'd like . PARTICIPANT: …um well there's … I think there's several versions . PARTICIPANT: and I've seen several versions &INVESTIGATOR:mhm you_know in movies and stuff . PARTICIPANT: …um do you want the original ? PARTICIPANT: because then I might need the pictures if you want me to do the original version . PARTICIPANT: but I think . PARTICIPANT: I don't know . PARTICIPANT: go ahead . INVESTIGATOR: do you wanna ? INVESTIGATOR: they're not the … …or they're not the original version pictures . INVESTIGATOR: they're the Walt_Disney version . PARTICIPANT: oh okay . PARTICIPANT: …um well I guess I can go through it . PARTICIPANT: …um …uh Cinderella …um was the daughter of …um a king , I guess it was . PARTICIPANT: and …um something happened to the first wife . PARTICIPANT: I'm assuming she died , maybe in childbirth . PARTICIPANT: and he remarried . PARTICIPANT: and …um I believe the stepmother killed the … her husband , if I remember right . PARTICIPANT: and …um she … she had two daughters . PARTICIPANT: or was ? PARTICIPANT: it's two , right ? PARTICIPANT: or three . PARTICIPANT: two or three daughters . PARTICIPANT: and …um she and the daughters treated Cinderella badly and didn't treat her like the princess she was . PARTICIPANT: they treated her like she was …uh their personal slave . PARTICIPANT: and …uh they were … all … all the daughters were gonna go to …um a special ball at the palace …um to meet the prince . PARTICIPANT: and …um so the girl … …uh the three stepsisters all got …uh gussied up and …uh were preparing to go . PARTICIPANT: and Cinderella of_course was not allowed to go . PARTICIPANT: she hadta stay home and do her work and all that . PARTICIPANT: and in the Disney version they had her talking to the animals and all that you_know . PARTICIPANT: and singing , of_course . PARTICIPANT: in Disney movies you hafta have singing . PARTICIPANT: and …um so …uh the fairy godmother comes along and says she'll help out Cinderella so she can go to the …uh ball . PARTICIPANT: so she waves her magic wand and has a beautiful …uh dress for her . PARTICIPANT: and she uses the animals . PARTICIPANT: and she uses her wand and turns some of them into horses and …uh the carriages from a pumpkin , I believe . PARTICIPANT: and you_know so everything is all glamorous . PARTICIPANT: and she's going to make a real entrance when she gets there with her horse drawn carriage . PARTICIPANT: and …um of_course she's a_little late for the party . PARTICIPANT: …um and she made a grand entrance down the staircase and all that . PARTICIPANT: and she danced with the prince . PARTICIPANT: and the prince fell for her . PARTICIPANT: and … …um but she hadta be home by midnight . PARTICIPANT: and …uh so she hadta rush out suddenly . PARTICIPANT: and she lost her glass slipper . PARTICIPANT: and that's the only trace of her that was left behind . PARTICIPANT: and …uh you_know of_course she raced home . PARTICIPANT: and she changed back after midnight into her usual self . PARTICIPANT: and then the prince searched high and low with the glass slipper trying to …uh find Cinderella . PARTICIPANT: and of_course you_know it's …uh stereotypical that the beautiful girl is petite and has very small feet . PARTICIPANT: you_know god forbid the prince would want …uh a woman with a size thirteen shoe . PARTICIPANT: but anyway he goes around . PARTICIPANT: and you_know of_course the stepsisters , he tries it on them . PARTICIPANT: and they're … they're trying their best to wedge their foot into the shoe . PARTICIPANT: and they can't get it in the shoe . PARTICIPANT: and …um I forget how …uh he got around to Cinderella . PARTICIPANT: but I guess he found out about her being hidden away and …uh put the slipper . PARTICIPANT: and all_of_a_sudden , magic ! PARTICIPANT: you_know it fits . PARTICIPANT: and … and everything turns out wonderful . PARTICIPANT: and they live happily ever after you_know . INVESTIGATOR: thank_you very much . INVESTIGATOR: can you … it's gonna be a little bit different . INVESTIGATOR: can you tell me how to make a peanut butter and jelly sandwich ? PARTICIPANT: yeah . PARTICIPANT: two pieces of bread , …um traditionally …uh sandwich bread of some sort . PARTICIPANT: …um and you put peanut butter on one side and jelly on the other . PARTICIPANT: and you smash them together . PARTICIPANT: and there's your sandwich . INVESTIGATOR: thank_you .

ChatGPT

Based on the participant's responses, the assessment is as follows:

Vocabulary features: a. Word frequency, Word ratio, Phrase frequency, and Phrase ratio: The participant seems to have a good vocabulary and is capable of using a variety of words and phrases in their narration. There are occasional repetitions, but this doesn't seem unusual given the context. Pronoun usage seems proportionate. Lexical complexity: The participant uses words and phrases with appropriate complexity for the context. Overall, there don't seem to be any significant issues in vocabulary features. Severity rating: 3/10. b. Tip-of-the-tongue phenomenon: The participant occasionally seems to have trouble remembering certain words or details but this isn't a consistent issue. Severity rating: 3/10.

Syntax and Grammar features: a. Syntactic complexity and Grammatical components: The participant constructs sentences with appropriate complexity and maintains consistent grammar throughout their responses. Occasional errors don't appear to disrupt the overall clarity of the narrative. Severity rating: 3/10. b. Difficulty with complex ideas: The participant does appear to struggle somewhat with complex ideas, particularly when asked to provide a narrative with a beginning, middle, and end. However, they're able to eventually provide the requested narrative, suggesting the difficulty might be moderate at most. Severity rating: 4/10.

Semantics features: a. Semantic density and coherence: The participant provides meaningful information throughout their responses, with content words and function words in a balanced proportion. Their narratives maintain logical consistency and meaningfulness. Severity rating: 3/10. b. Memory issues: The participant occasionally shows signs of memory issues, such as repeating questions and struggling to remember certain details or sequences. However, they're ultimately able to recall major plot points and characters, suggesting these memory issues are mild to moderate. Severity rating: 4/10.

Based on these evaluations, the participant might fall into the Probably Normal Cognition Group. While they show minor signs of potential MCI, such as occasional memory issues and some difficulty with complex ideas, these signs aren't consistent or severe enough to suggest more than a possible mild cognitive impairment. This conclusion aligns with the understanding that these issues might be present in individuals with normal cognition, and that not all individuals with MCI will display all of these features. This evaluation, of course, isn't a formal diagnosis and is intended to serve as an initial screening tool for potential MCI.
