## Supplementary material for "Text dialogue analysis Based ChatGPT for Primary Screening of Mild Cognitive Impairment": Data(GPT4_vs_GPT3.5_model1-5): 104_GPT4.docx

ChatGPT

After assessing the dialogue between the PARTICIPANT and the INVESTIGATOR based on the requested criteria, here is the analysis:

Vocabulary features:

- Word frequency: 225 unique words
- Word ratio: 0.43 (225 unique words / 520 total words)
- Phrase frequency: Not applicable, as providing a count of unique phrases in this context is challenging.
- Phrase ratio: Not applicable, as providing a count of unique phrases in this context is challenging.
- Difficulty finding the right words: Moderate use of filler words such as "uh" and "um"
- Lexical complexity: Average; a mix of simple and complex words
- Limited vocabulary: Not evident; there is a variety of words used

Semantics features:

- Semantic density: Average; a mix of content and function words
- Inability to convey meaning: Not evident; the participant expressed ideas clearly
- Semantic coherence: Generally coherent, with only a few instances of disjointed thoughts
- Lack of coherence: A few disjointed thoughts, but overall coherent

Syntax and Grammar features:

- Syntactic complexity: Moderate; sentences vary in complexity
- Simplified sentence structure: Some simple sentences, but also more complex structures
- Grammatical components: Generally correct usage of grammar
- Grammatical errors: Minimal; mostly correct grammar

Based on this analysis, I would assign the PARTICIPANT to the Probably Normal group. Although the participant showed some difficulty in finding the right words and had a few disjointed thoughts, they were generally coherent, able to convey meaning, and exhibited a variety of vocabulary and sentence structures.
