## Supplementary material for "Text dialogue analysis Based ChatGPT for Primary Screening of Mild Cognitive Impairment": Data(GPT4_vs_GPT3.5_model1-5): 105_GPT3.5 (2).docx

Attention! You do not need to diagnose, just follow the steps below to initially screen for PARTICIPANT with possibly mild cognitive impairment (MCI). Some differences between MCI and normal cognition are also given, please analyse them in detail, step by step. Firstly, please assess the Vocabulary features of PARTICIPANT at the Word frequency and Word ratio, Phrase frequency and Phrase ratio, Word-finding difficulties, Repetition, Lexical complexity, Limited vocabulary. Word frequency: Count the occurrences of each unique word in the text. Unique words are distinct words that appear in the text, not taking into account their repetitions. Each word is counted only once, regardless of how many times it occurs. Word ratio: Calculate the proportion of unique words to the total number of words in the text. Phrase frequency: Count the occurrences of each unique phrases in the text. Unique phrases are distinct combinations of words that appear in the text, not taking into account their repetitions. Phrase ratio: Calculate the proportion of unique phrases to the total number of phrases in the text. Word-finding difficulties: Frequent pauses or use of filler words, like "uh" or "um,". MCI: more frequent instances, longer time to find the right word, or using more general terms. Normal: fewer instances, and better lexical retrieval. Repetition: The participant repeats certain phrases or ideas a few times throughout the conversation. MCI: more frequent repetition. Normal: less frequent repetition (some repetition of words but not excessive) and better memory of what has been mentioned. Lexical complexity: Measure the variety and sophistication of the vocabulary used, considering factors, including word length and frequency of rare words. Limited vocabulary: Repeated use of the same words or simple words. MCI: slightly reduced vocabulary, and more frequent use of simpler words or common terms; Normal: richer vocabulary (low limitation). Secondly, please assess the Semantics features of PARTICIPANT, from Semantic density and Inability to convey meaning, Semantic coherence and Lack of coherence. Semantic density: Measure the amount of meaningful information in the text, and considering factor is the number of content words (nouns, verbs, adjectives, adverbs) relative to function words (prepositions, pronouns, conjunctions, etc.). Inability to convey meaning: Difficulty in expressing ideas clearly, leading to confusion or misunderstanding. Normal: the participant was able to describe the scene clearly. Semantic coherence: Assess the logical consistency and meaningfulness of the text, which can be done by analyzing the relationships between words, phrases, and sentences, and checking for coherence and cohesion. Lack of coherence: Difficulty in maintaining a logical flow of ideas, with thoughts appearing disjointed or unrelated. MCI: struggle with organizing thoughts coherently or maintaining a clear focus in conversation; Normal: greater coherence and organization in their storytelling and conversations. Thirdly, please assess the Syntax and Grammar features of PARTICIPANT, from Syntactic complexity and Simplified sentence structure, Grammatical components and Grammatical errors. Syntactic complexity: Measure the complexity of the sentence structure in the text, considering factors such as the number of clauses per sentence, the average length of sentences, and the use of subordinate clauses. Simplified sentence structure: A lack of complex sentence structures. MCI: simpler sentence structures; Normal: in longer dialogues there is a greater variety of sentence structures (moderately complex), but in short dialogues the sentences may turn out to be simple. Grammatical components: Analyze the text for correct usage of grammar, such as subject-verb agreement, tense consistency, and proper use of pronouns, prepositions, and conjunctions. MCI: avoidance of complex grammatical constructions; Normal: better use of complex grammatical constructions. Grammatical errors: Frequent mistakes in verb tense, subject-verb agreement, or other aspects of grammar. MCI: more frequent grammatical errors, such as mistakes in verb tense and subject-verb agreement; Normal: fewer grammatical errors (a few errors, but not significant). Finally, Please dichotomise PARTICIPANT into the Probably Normal group or the Probably Mild Cognitive Impairment group. PARTICIPANT with MCI might exhibit more difficulty with vocabulary, sentence structure, repetition, word-finding, coherence, and grammar. However, it is not definitive and requires a comprehensive judgement. Now, you do not have to respond to this dialogue. When I have entered THE DIALOGUE BETWEEN PARTICIPANT AND INVESTIGATOR, evaluate PARTICIPANT according to the evaluation rules above. INVESTIGATOR: picture PARTICIPANT: okay there's a little girl …uh who's reaching up to get some cookies from her brother who's standing on a stool that's starting to tip as he reaches into the cookie jar . PARTICIPANT: the lid's off the cookie jar already . PARTICIPANT: and the cookies are pretty full . PARTICIPANT: and then not paying attention is the mother standing there . PARTICIPANT: and she's really not paying attention (be)cause her sink's overflowing with water . PARTICIPANT: and she's just standing there drying a dish . PARTICIPANT: …um they're …uh appear to be in the kitchen . PARTICIPANT: and there's a nice big window in front of the kitchen sink where you can see outdoors a_bit and see what's going on with the grass and the trees and bushes and stuff . PARTICIPANT: and then there's …um a_couple_of … it looks like a saucer and a dish and a plate to the right of the kitchen sink …uh which again is overflowing and spilling water on the floor . INVESTIGATOR: this picture tells a story . INVESTIGATOR: look at everything that's happening and then tell me a story about what you see . INVESTIGATOR: and tell me a story with a beginning a middle and an end . PARTICIPANT: okay …um little girl was outside playing . PARTICIPANT: and her cat went up the tree . PARTICIPANT: the … …um she was very distressed at the thought that her cat was up the tree , and was trying to encourage it to come down , as was her dog who was barking &=clears:throat as_well . PARTICIPANT: but part of the issue was that dad was trying to help get the cat down . PARTICIPANT: and he got stuck up there himself . PARTICIPANT: he wasn't very comfortable being up the tree . PARTICIPANT: so the dog was barking at the dad and was trying to help get the cat down . PARTICIPANT: so somebody had called the fireman so_that they could come and help . PARTICIPANT: and it appeared as_though the firemen were rushing with their ladders in to try to help get dad down and also get the cat down . PARTICIPANT: because the father's ladder that he had used to get up had fallen off the tree . PARTICIPANT: and so the father had no way of getting down as_well . INVESTIGATOR: thank_you . INVESTIGATOR: (pic)tures …um and both pictures are gonna tell a story . PARTICIPANT: mhm INVESTIGATOR: so look at everything that's happening and then tell me a story about what you see . INVESTIGATOR: tell me the story with a beginning a middle and an end . PARTICIPANT: hm well there's two … two …uh lovely Norman_Rockwell photos here that are depicting a family going on a trip . PARTICIPANT: and …uh in the bottom photo the family looks a_bit tired . PARTICIPANT: so I'd say they're probably on their way home from the trip that they had just taken . PARTICIPANT: and based on what's on top of the …um car , the family had been going for a day out . PARTICIPANT: …um and they were going on the water . PARTICIPANT: they were gonna go …uh rowboating . PARTICIPANT: so as they're getting ready to go , they're all excited and happy . PARTICIPANT: the children are looking out the window . PARTICIPANT: they've got the dog with them . PARTICIPANT: and they're very excited about what's gonna happen for the day . PARTICIPANT: and …uh typical of a long trip , dad's got his cigar hangin(g) out_of his mouth . PARTICIPANT: …um one of the boys is not very happy that …uh dad's smoking a cigar . PARTICIPANT: it's a_bit stinky for him . PARTICIPANT: but the other two children are quite happy . PARTICIPANT: and they're looking forward to their day out . PARTICIPANT: by the time they're finished and they're on the road home everybody looks a_bit tired and …uh ready to fall asleep . PARTICIPANT: mom has actually fallen asleep in the front . PARTICIPANT: and dad's got his eyes fixed on the road …uh trying to get home as quickly as possible , because …uh it looks quite dark out …uh after the long day at the lake . INVESTIGATOR: thank_you . INVESTIGATOR: …um have you heard the story of Cinderella before ? PARTICIPANT: yeah . INVESTIGATOR: okay so I'm gonna have you tell me the story of Cinderella . INVESTIGATOR: I do have some pictures that I can show you to jog your memory if you'd like . INVESTIGATOR: …um so I will show you the pictures …um and then I'll have you tell me the story of Cinderella at the end . INVESTIGATOR: alright can you tell me the story of Cinderella ? PARTICIPANT: I'll give it a shot (be)cause there's a_lot more in there than I ever remember . INVESTIGATOR: that's okay , just from what you remember . PARTICIPANT: yeah exactly . PARTICIPANT: …um …s Cinderella ended up living with …uh her stepmother . PARTICIPANT: and there was two stepsisters that she had . PARTICIPANT: and they did not treat Cinderella the same as they did the stepsisters . PARTICIPANT: so Cinderella was a very sweet girl . PARTICIPANT: and she was compliant with what was asked of her . PARTICIPANT: there was a special ball coming up . PARTICIPANT: and the … …s the stepsisters , who were really rather wicked , were getting themselves all prepared for the ball . PARTICIPANT: and Cinderella was wondering if she'd be able to go . PARTICIPANT: and the fairy godmother came along and …uh made sure she had a dress and was able to go off to the ball . PARTICIPANT: so she went separately off to the ball . PARTICIPANT: and when she got there , there was a requirement that she hadta be home by midnight . PARTICIPANT: and she was given the opportunity to dance with the prince . PARTICIPANT: and the prince was looking for …uh someone that he might be able to live with forever after . PARTICIPANT: and he enjoyed dancing with Cinderella . PARTICIPANT: but as it started to approach midnight , Cinderella hadta run out because she knew she hadta be home . PARTICIPANT: that was the condition in which she was able to go to the ball . PARTICIPANT: and as she left , she ran so fast that she lost her shoe . PARTICIPANT: so when she got home , it turns out that the prince was really enthralled with her and wanted to find her and … and meet her again and had no way of knowing who she was and where she came from except_for the red slipper that she had lost on her way out . PARTICIPANT: so slowly he began to figure out how he could find Cinderella by using that slipper . PARTICIPANT: and he was able to match the slipper with Cinderella . PARTICIPANT: and throughout all this the wicked stepsisters were not happy that it appeared that Cinderella was favored by the prince and she … they were not . PARTICIPANT: …uh in the end Cinderella and the prince didn't meet up . PARTICIPANT: they married and they lived …ev happily ever after much to the chagrin of the two stepsisters who weren't happy at all .

ChatGPT

3 / 3

Let's analyze the dialogue between the participant and the investigator based on the evaluation rules provided.

1. Vocabulary features:

- Word frequency: The participant uses a variety of words throughout the conversation. There are no repeated words, indicating a good word frequency.
- Word ratio: Since the total number of words is not provided, we cannot calculate the word ratio.
- Phrase frequency: The participant uses distinct combinations of words and phrases to describe the scene, indicating a good phrase frequency.
- Phrase ratio: Since the total number of phrases is not provided, we cannot calculate the phrase ratio.
- Word-finding difficulties: The participant does not exhibit frequent pauses or use filler words, demonstrating good lexical retrieval and fewer instances of word-finding difficulties.
- Repetition: The participant does not excessively repeat phrases or ideas, indicating normal repetition and memory recall.
- Lexical complexity: The participant uses a variety of vocabulary and demonstrates sophistication in word choice, indicating good lexical complexity.
- Limited vocabulary: The participant does not exhibit repeated use of the same words or simple words, indicating a rich vocabulary.

2. Semantics features:

- Semantic density: Without the exact count of content words and function words, we cannot determine the semantic density.
- Inability to convey meaning: The participant expresses ideas clearly, providing a coherent description of the scene, indicating normal ability to convey meaning.
- Semantic coherence: The participant maintains logical consistency and meaningfulness throughout the conversation, demonstrating good semantic coherence.
- Lack of coherence: The participant organizes thoughts coherently, maintains a clear focus, and provides a logical flow of ideas, indicating normal coherence.

3. Syntax and Grammar features:

- Syntactic complexity: The sentence structures used by the participant are moderately complex, with multiple clauses and varying lengths, indicating good syntactic complexity.
- Simplified sentence structure: The participant uses a mix of simple and complex sentence structures, indicating normal sentence structure complexity.
- Grammatical components: The participant demonstrates correct usage of grammar, including subject-verb agreement, tense consistency, and proper use of pronouns, prepositions, and conjunctions, indicating good grammatical components.
- Grammatical errors: The participant makes very few grammatical errors, such as minor slips in verb tense or subject-verb agreement, indicating normal grammar usage.

Based on the evaluation, the participant shows characteristics of normal cognition rather than mild cognitive impairment (MCI). The participant demonstrates good vocabulary usage, semantic coherence, syntactic complexity, and grammatical components. There are no significant indicators of vocabulary difficulties, incoherence, simplified sentence structures, or frequent grammatical errors, which are often associated with MCI. However, please note that this evaluation is based on the provided dialogue and should not be considered as a definitive diagnosis. A comprehensive assessment by a medical professional is required for an accurate judgment.
