## Supplementary material for "Text dialogue analysis Based ChatGPT for Primary Screening of Mild Cognitive Impairment": Data(GPT4_vs_GPT3.5_model1-5): 105_GPT3.5 (3).docx

ChatGPT

Based on the provided conversation, let's analyze the indicators and evaluate the severity for the participant.

Vocabulary features:

- Word frequency and Word ratio: It is not possible to count the occurrences of each unique word, pronoun, or specific name/noun in the given conversation. Therefore, it is not feasible to calculate the proportion of unique words or the ratio of pronouns to specific names.
- Phrase frequency and Phrase ratio: It is not possible to count the occurrences of each unique phrase in the given conversation. Therefore, it is not feasible to calculate the proportion of unique phrases.
- Lexical complexity: It is not possible to measure the variety and sophistication of the vocabulary used based on the provided conversation.

Evaluation Indicators:

1. Limited vocabulary: Not applicable.
2. Increased verbosity: The participant tends to provide detailed descriptions and includes additional information not directly related to the main story. Severity: 4
3. Overlearned phrases: Not applicable.
4. Improper use of pronouns: There are no clear difficulties or errors in pronoun usage in the conversation. Not applicable.
5. Hesitation (Word-finding difficulties): The participant occasionally uses filler words like "uh" and "um." Severity: 3
6. Circumlocution (Word-finding difficulties): The participant does not exhibit significant difficulty finding specific words or using more general words in place of specific ones. Not applicable.
7. Tip-of-the-tongue phenomenon: There is no clear evidence of the tip-of-the-tongue phenomenon. Not applicable.
8. Repetition: The participant does not repeat the same stories, questions, or statements within a short span of time. Not applicable.

Syntax and Grammar features:

- Syntactic complexity: The sentence structure in the conversation is generally simple and straightforward, with few complex sentence constructions. Severity: 2
- Grammatical components: There are no significant grammatical errors in the conversation. Severity: 1

Evaluation Indicators:

1. Simplified sentence structure and grammar: The participant tends to use simpler sentence structures and avoids complex grammatical constructions. Severity: 3
2. Grammatical errors: There are no significant grammatical errors observed in the conversation. Severity: 1
3. Difficulty with complex ideas: The participant demonstrates some difficulty when the narrative becomes more complex, but overall handles simpler sentences and ideas. Severity: 3

Semantics features:

- Semantic density: It is not possible to measure the semantic density based on the provided conversation.
- Semantic coherence: The conversation is logically consistent and meaningful.

Evaluation Indicators:

1. Semantic paraphasias: The participant does not exhibit significant errors in word choice or odd combinations of words. Not applicable.
2. Lack of coherence: The participant maintains a coherent line of thought throughout the conversation. Not applicable.
3. Consistency in errors: There is no evidence of consistent language errors. Not applicable.
4. Lack of semantic fluency: The participant demonstrates fluency in describing the scenes and events without significant difficulty in naming items or describing processes. Not applicable.
5. Disorganized narrative structure: The participant maintains a coherent narrative structure with a beginning, middle, and end. Not applicable.
6. Orientation issues: There is no evidence of confusion about date, time, place, or sequence of events. Not applicable.
7. Decline in conversational engagement and responsiveness: The participant actively engages in conversation, responds appropriately, and initiates topics. Not applicable.
8. Memory issues: There is no evidence of significant memory issues, such as repeating questions or difficulty recalling information. Not applicable.
9. Lack of abstract thinking and synthesis: The participant demonstrates the ability to synthesize the overall situation and understand abstract concepts. Not applicable.
10. Abnormal emotions: There are no clear changes in tone or style of speech, lack of initiative, or abnormal emotions observed. Not applicable.

Based on the analysis of the indicators, it is difficult to dichotomize the participant into either the Probably Normal Cognition Group or the Probably Mild Cognitive Impairment Group. The participant shows some minor indicators of verbosity, hesitation (word-finding difficulties), simplified sentence structure and grammar, and difficulty with complex ideas. However, these indicators are not severe or consistent enough to conclusively suggest mild cognitive impairment. It is important to note that not all assessment indicators are present, and a comprehensive evaluation of cognitive function would require additional information and tests conducted by a qualified healthcare professional.
