## Supplementary material for "Text dialogue analysis Based ChatGPT for Primary Screening of Mild Cognitive Impairment": Data(GPT4_vs_GPT3.5_model1-5): 105_GPT3.5 (4).docx

ChatGPT

Based on the participant's language features, we can assess and rate the severity of the indicators as follows:

Vocabulary features: a. Word frequency, word ratio, phrase frequency, and phrase ratio: Not assessed in the given conversation. b. Lexical complexity: Not assessed in the given conversation. c. Severity assessment:

- Limited vocabulary: 2
- Increased verbosity: 2
- Overlearned phrases: 1
- Improper use of pronouns: 1
- Hesitation: 2
- Circumlocution: 1
- Tip-of-the-tongue phenomenon: 1
- Repetition: 1

Syntax and grammar features: a. Syntactic complexity and grammatical components: Not assessed in the given conversation. b. Severity assessment:

- Simplified sentence structure and grammar: 2
- Grammatical errors: 1
- Difficulty with complex ideas: 1

Semantics features: a. Semantic density and coherence: Not assessed in the given conversation. b. Severity assessment:

- Semantic paraphasias: 1
- Lack of coherence: 1
- Consistency in errors: 1
- Lack of semantic fluency: 1
- Disorganized narrative structure: 1
- Orientation issues: 1
- Decline in conversational engagement and responsiveness: 1
- Memory issues: 1
- Lack of abstract thinking and synthesis: 1
- Abnormal emotions: 1

Based on the comprehensive synthesis of all the indicators, the participant can be classified into the Probably Normal Cognition Group. The language features mentioned in the conversation do not strongly indicate the presence of mild cognitive impairment (MCI). However, it's important to note that this assessment is based solely on the language features and a comprehensive evaluation by a healthcare professional is necessary to confirm the cognitive status of an individual
