## Supplementary material for "Text dialogue analysis Based ChatGPT for Primary Screening of Mild Cognitive Impairment": Data(GPT4_vs_GPT3.5_model1-5): 105_GPT3.5 (5).docx

ChatGPT

Based on the language features provided, we can assess the participant's language abilities and rate the severity of each evaluation indicator. Please note that this assessment is based solely on the provided text and should not be considered a formal diagnosis. The final categorization will be made after synthesizing all the indicators.

Vocabulary features: a. Word frequency and word ratio: The participant's vocabulary seems adequate, with a moderate variety of words used. The participant demonstrates the ability to use specific names (nouns) and pronouns appropriately. However, without a complete analysis of the text, it's challenging to assess word frequency accurately. Rating: 6

b. Tip-of-the-tongue phenomenon: There is no direct evidence of the tip-of-the-tongue phenomenon in the provided text. Rating: 1

Syntax and grammar features: a. Syntactic complexity and grammatical components: The participant's sentences are generally well-formed and demonstrate a reasonable level of complexity. The text contains multiple clauses, appropriate use of verb tenses, and proper subject-verb agreement. Rating: 7

b. Difficulty with complex ideas: The participant shows some difficulty when the narrative becomes more complex. The second picture prompt, which requires a more elaborate story, seems to challenge the participant's ability to generate a coherent and detailed response. Rating: 6

Semantics features: a. Semantic density and coherence: The text demonstrates a relatively balanced use of content words (nouns, verbs, adjectives, adverbs) and function words (prepositions, pronouns, conjunctions). The overall semantic coherence is reasonably maintained, with logical relationships between words, phrases, and sentences. Rating: 7

b. Memory issues: The participant does not display any apparent memory issues based on the given text. Rating: 1

Based on the comprehensive synthesis of the indicators:

- Vocabulary features: 6
- Syntax and grammar features: 7
- Semantics features: 7

Considering the overall assessment, the participant's language features suggest they are likely in the Probably Normal Cognition Group. However, it's important to note that this assessment is based on limited information, and a comprehensive evaluation involving additional tasks and observations would be necessary for a more accurate determination.
