## Supplementary material for "Text dialogue analysis Based ChatGPT for Primary Screening of Mild Cognitive Impairment": Data(GPT4_vs_GPT3.5_model1-5): 105_GPT3.5.docx

ChatGPT

Based on the analysis of language features, here is the updated assessment for each category:

Vocabulary features:

- Word frequency: The participant continues to display a decent range of unique words, indicating a satisfactory vocabulary.
- Word ratio: The proportion of unique words to total words remains moderate, suggesting a reasonable vocabulary diversity.
- Phrase frequency: The participant maintains a fair number of unique phrases, resulting in fewer repetitions.
- Phrase ratio: The proportion of unique phrases to total phrases is still moderate, indicating a moderate phrase diversity.
- Difficulty finding the right words: The participant still does not exhibit significant difficulty finding the right words, with minimal pauses or use of filler words.
- Lexical complexity: The vocabulary used by the participant continues to show a mix of basic and more complex words, indicating a satisfactory level of lexical complexity.
- Limited vocabulary: The participant demonstrates a decent range of words, avoiding excessive repetition and showcasing variety and sophistication.

Semantics features:

- Semantic density: The participant provides a satisfactory amount of meaningful information in their responses, with a balanced ratio of content words to function words.
- Inability to convey meaning: The participant continues to express ideas relatively clearly, with fewer instances of difficulty in conveying meaning.
- Semantic coherence: The participant's responses maintain logical consistency and meaningfulness, with good coherence and cohesion between words, phrases, and sentences.
- Lack of coherence: The participant sustains a logical flow of ideas, with thoughts appearing connected and related.

Syntax and Grammar features:

- Syntactic complexity: The participant consistently uses a mix of sentence structures, including both simple and complex sentences, showcasing variety.
- Simplified sentence structure: The participant maintains a good balance between simple and complex sentence structures, indicating language proficiency.
- Grammatical components: The participant generally displays correct usage of grammar, including subject-verb agreement, tense consistency, and proper use of pronouns, prepositions, and conjunctions.
- Grammatical errors: The participant makes fewer grammatical errors, showing improvement in verb tense, subject-verb agreement, and other aspects of grammar.

Based on the language assessment, the participant continues to demonstrate good language skills. The vocabulary usage, semantic coherence, and syntactic complexity remain at a satisfactory level, indicating an overall proficient command of language. Therefore, based on the provided conversation, the participant is likely to belong to the Probably Normal group. However, it's important to note that this is an initial screening, and further evaluation would be required to make a definitive determination regarding cognitive impairment.
