## Supplementary material for "Text dialogue analysis Based ChatGPT for Primary Screening of Mild Cognitive Impairment": Data(GPT4_vs_GPT3.5_model1-5): 105_GPT4 (2).docx

ChatGPT

In order to evaluate this participant's cognitive state, let's examine their performance according to the given categories:

1. **Vocabulary Features:**

- Word frequency and Word ratio: An in-depth analysis is required to calculate the word frequency and word ratio. This involves counting the occurrences of each unique word and calculating the proportion of unique words to the total words in the text.
- Phrase frequency and Phrase ratio: Similarly, a more detailed analysis is required to calculate the phrase frequency and phrase ratio. This involves counting the occurrences of each unique phrase and calculating the proportion of unique phrases to the total phrases in the text.
- Word-finding difficulties: There were several instances where the participant used filler words like "um" and "uh". However, the participant didn't take an exceedingly long time to find the right word and was generally successful in finding the appropriate terms.
- Repetition: The participant repeated certain phrases or ideas a few times throughout the conversation, but this was not excessive.
- Lexical complexity: The participant used a variety of words of different lengths and complexity, and the frequency of rare words was not exceptionally high or low.
- Limited vocabulary: The participant showed a good range of vocabulary, using both simple and more complex words.

2. **Semantics Features:**

- Semantic density: The participant used a good mix of content words and function words, indicating a relatively high semantic density.
- Inability to convey meaning: The participant was able to express ideas clearly and there were no instances of confusion or misunderstanding.
- Semantic coherence: The participant demonstrated good semantic coherence, with clear relationships between words, phrases, and sentences.
- Lack of coherence: The participant maintained a logical flow of ideas, with no evidence of disjointed or unrelated thoughts.

3. **Syntax and Grammar Features:**

- Syntactic complexity: The participant demonstrated moderate syntactic complexity, using both simple and complex sentence structures.
- Simplified sentence structure: The participant did not overuse simple sentence structures and demonstrated a good understanding of complex sentence structures.
- Grammatical components: The participant used correct grammar in most cases, with consistent tense and proper use of pronouns, prepositions, and conjunctions.
- Grammatical errors: There were very few grammatical errors in the participant's speech, with correct verb tense and subject-verb agreement in most cases.

Based on this analysis, the participant demonstrates linguistic characteristics more aligned with the **Probably Normal** group. They show a good command of vocabulary, maintain semantic coherence, and use complex sentence structures correctly. However, it is important to note that a full linguistic analysis would require more detailed quantitative data on word and phrase frequency and ratios. This determination should not be used as a diagnosis, and any concerns about cognitive impairment should be evaluated by a healthcare professional.
