## Supplementary material for "Text dialogue analysis Based ChatGPT for Primary Screening of Mild Cognitive Impairment": Data(GPT4_vs_GPT3.5_model1-5): 105_GPT4 (3).docx

ChatGPT

Based on the participant's response, let's analyze the vocabulary, syntax, grammar, and semantic features, and rate the severity of possible issues in those areas.

Vocabulary Features: Word Frequency and Ratio: The participant has used a variety of unique words and pronouns. The ratio of unique words to the total number of words and the ratio of pronouns to specific names (or nouns) seem balanced and appropriate. Phrase Frequency and Ratio: The participant has also used a good variety of unique phrases and the ratio of unique phrases to the total number of phrases seems appropriate. Lexical Complexity: The vocabulary used seems quite diverse, with a mix of simple and more complex words, indicating a satisfactory lexical complexity.

1. Limited vocabulary: 2/10
2. Increased verbosity: 3/10
3. Overlearned phrases: 2/10
4. Improper use of pronouns: 2/10
5. Hesitation (Word-finding difficulties): 4/10
6. Circumlocution (Word-finding difficulties): 3/10
7. Tip-of-the-tongue phenomenon: 3/10
8. Repetition: 2/10

Syntax and Grammar Features: Syntactic Complexity: The sentences appear to be reasonably complex with a balanced mix of short and long sentences. Grammatical Components: The grammar usage seems correct and appropriate.

1. Simplified sentence structure and grammar: 3/10
2. Grammatical errors: 2/10
3. Difficulty with complex ideas: 3/10

Semantics Features: Semantic Density: The text seems rich with meaningful information and content words. Semantic Coherence: The participant's responses are logically consistent and meaningful.

1. Semantic paraphasias: 2/10
2. Lack of coherence: 2/10
3. Consistency in errors: 2/10
4. Lack of semantic fluency: 3/10
5. Disorganized narrative structure: 2/10
6. Orientation Issues: 2/10
7. Decline in conversational engagement and responsiveness: 2/10
8. Memory issues: 3/10
9. Lack of abstract thinking and synthesis: 3/10
10. Abnormal emotions: 2/10

Taking all the factors into consideration, it appears that the participant's cognitive function is more aligned with the Probably Normal Cognition Group. This is a preliminary assessment and not a diagnosis. It's recommended to seek professional help for a comprehensive evaluation
