## Supplementary material for "Text dialogue analysis Based ChatGPT for Primary Screening of Mild Cognitive Impairment": Data(GPT4_vs_GPT3.5_model1-5): 105_GPT4 (4).docx

ChatGPT

Based on the provided transcript, here's an assessment of the participant's language, syntax, and semantic features:

**Vocabulary features:** a. Word frequency, word ratio, phrase frequency, and phrase ratio: Given the available data, a numerical analysis cannot be done, however, the participant uses a reasonable variety of words and phrases, with no clear overuse of any single term or phrase. b. Lexical complexity: The participant's vocabulary seems to be of moderate complexity, with the use of a few multisyllabic words and more complex phrases. c.

- Limited vocabulary: 2/10. The participant uses a variety of words to describe scenes and to tell stories.
- Increased verbosity: 2/10. The participant provides details without being excessively verbose.
- Overlearned phrases: 2/10. The participant doesn’t seem to rely heavily on overlearned phrases.
- Improper use of pronouns: 1/10. The participant seems to use pronouns correctly.
- Hesitation: 4/10. There are some instances of hesitation marked by "uh" and "um".
- Circumlocution: 2/10. The participant seems quite direct in their responses, with little evidence of talking around a subject.
- Tip-of-the-tongue phenomenon: 2/10. The participant appears to be able to recall most words they want to use.
- Repetition: 2/10. There is little to no repetition in the participant's speech.

**Syntax and grammar features:** a. Syntactic complexity and grammatical components: The participant uses complex sentences with proper structure, indicating good understanding of syntax and grammar. b.

- Simplified sentence structure and grammar: 1/10. The participant doesn't simplify sentence structure, and uses complex sentences accurately.
- Grammatical errors: 1/10. The participant doesn’t make noticeable grammatical errors.
- Difficulty with complex ideas: 2/10. The participant seems capable of understanding and expressing complex ideas.

**Semantics features:** a. Semantic density and coherence: The participant provides detailed responses and maintains a clear topic throughout their speech. They are coherent and follow the narrative well. b.

- Semantic paraphasias: 1/10. There is no clear instance of semantic paraphasias.
- Lack of coherence: 1/10. The participant’s stories are coherent and follow a logical narrative.
- Consistency in errors: 1/10. There are not many errors to establish a consistency.
- Lack of semantic fluency: 1/10. The participant is quite fluent and able to express complex ideas.
- Disorganized narrative structure: 1/10. The participant’s narratives are well organized.
- Orientation issues: 1/10. The participant doesn't seem to have orientation issues.
- Decline in conversational engagement and responsiveness: 1/10. The participant stays engaged in the conversation.
- Memory issues: 2/10. There doesn't seem to be any substantial memory issues.
- Lack of abstract thinking and synthesis: 1/10. The participant shows capability of abstract thinking and synthesis.
- Abnormal emotions: 1/10. There doesn't appear to be any abnormal emotional expression.

Based on the comprehensive synthesis of all the indicators, the participant would likely fall into the Probably Normal Cognition Group. The participant shows minor hesitation but no other substantial issues related to cognitive function as assessed from their language, syntax, and semantic features. It's important to note, however, that more thorough testing should be performed for a definitive assessment, as this assessment only serves as an initial screening tool for potential MCI
