## Supplementary material for "Text dialogue analysis Based ChatGPT for Primary Screening of Mild Cognitive Impairment": Data(GPT4_vs_GPT3.5_model1-5): 105_GPT4 (5).docx

ChatGPT

Based on the transcript provided, here are my assessments for the participant's language, syntax, and semantic features. However, this should be used only as a guide and not a formal diagnostic tool. The individual may perform differently in different situations or on different days.

Vocabulary features: a. Word frequency, word ratio, phrase frequency, and phrase ratio: The participant uses a diverse set of words and phrases, repeating certain words in natural conversational contexts. There are relatively few instances of excessive repetition, and the use of pronouns and specific names (or nouns) seems proportional. Lexical complexity: The participant uses a wide range of vocabulary, with appropriate usage of uncommon and descriptive words. Overall score: 2/10. b. Tip-of-the-tongue phenomenon: The participant occasionally paused during the conversation, which could be instances of the tip-of-the-tongue phenomenon. However, it is difficult to confirm without more information. Based on available data, the severity would be rated 3/10.

Syntax and grammar features: a. Syntactic complexity and grammatical components: The participant constructs complex sentences using a variety of grammatical structures and demonstrates good subject-verb agreement, tense consistency, and proper use of pronouns, prepositions, and conjunctions. The usage of subordinate clauses is present, indicating good syntactic complexity. Overall score: 2/10. b. Difficulty with complex ideas: The participant seems to handle complex ideas fairly well and was able to construct narratives with a beginning, middle, and end. The score for difficulty with complex ideas is 3/10.

Semantics features: a. Semantic density and coherence: The participant produces text with a high amount of meaningful information (high semantic density), and demonstrates logical consistency and meaningfulness throughout the conversation (high semantic coherence). The balance between content words and function words seems appropriate. Overall score: 2/10. b. Memory issues: The participant retells the Cinderella story with minor deviations, but overall remembers major plot points and characters. They provided detailed responses to the picture descriptions and didn't repeat questions. There are minimal signs of memory issues in this context, so the score is 3/10.

Overall, the participant demonstrates mild issues in the conversation. However, these are well within the range of normal conversational variance. Based on this conversation alone, they would be classified as part of the Probably Normal Cognition Group
