## Supplementary material for "Text dialogue analysis Based ChatGPT for Primary Screening of Mild Cognitive Impairment": Data(GPT4_vs_GPT3.5_model1-5): 105_GPT4.docx

ChatGPT

Assessment of PARTICIPANT's language features:

Vocabulary features:

1. Word frequency: 233 unique words
2. Word ratio: 0.37 (233 unique words / 632 total words)
3. Phrase frequency: 107 unique phrases
4. Phrase ratio: 0.38 (107 unique phrases / 280 total phrases)
5. Difficulty finding the right words: Occasional use of filler words such as "um" and "uh"
6. Lexical complexity: Moderate variety and sophistication of vocabulary
7. Limited vocabulary: Slightly repetitive use of simple words

Semantics features:

1. Semantic density: Moderate amount of meaningful information in the text
2. Inability to convey meaning: Minimal difficulty in expressing ideas
3. Semantic coherence: Logical consistency and meaningfulness of the text are generally maintained
4. Lack of coherence: Occasional disjointed or unrelated thoughts

Syntax and Grammar features:

1. Syntactic complexity: Moderate complexity in sentence structure
2. Simplified sentence structure: Occasional use of short, simple sentences
3. Grammatical components: Generally correct usage of grammar
4. Grammatical errors: Minimal errors in grammar

Based on the language features assessed, I would assign PARTICIPANT to the Probably Normal group. There are some instances of filler words, slightly repetitive vocabulary, and occasional disjointed thoughts, but overall, the participant's language abilities appear to be within the normal range.
