## Supplementary material for "Text dialogue analysis Based ChatGPT for Primary Screening of Mild Cognitive Impairment": Data(GPT4_vs_GPT3.5_model1-5): 106_GPT3.5 (2).docx

ChatGPT

3 / 3

Let's analyze the dialogue from the perspective of the evaluation rules provided.

Vocabulary features:

- Word frequency: We count the occurrences of each unique word in the text. Here are the unique words used by the participant: "alright," "action," "oh," "well," "the," "little," "girl," "is," "reaching," "waiting," "for," "her," "brother," "to," "give," "a," "cookie," "boy," "has," "from," "jar," "but," "he's," "knocked," "lid," "off," "and," "he," "tumbling," "stool," "uh," "m:other," "drying," "dishes," "left," "hand," "just," "touching," "another," "right," "you," "only," "want," "say," "that's," "about," "it," "can," "tell," "me," "what," "story," "was," "anything," "remember," "well," "uncle," "Bill," "Johnny," "admired," "because," "had," "taken," "him," "on," "a," "camping," "trip." So, there are 70 unique words.
- Word ratio: The total number of words in the text is 103. The word ratio is calculated by dividing the number of unique words (70) by the total number of words (103), which results in approximately 0.68.

The participant's use of vocabulary seems to be fairly diverse, with a moderate word ratio.

Word-finding difficulties:

- The participant does not exhibit any noticeable word-finding difficulties. They speak fluently without frequent pauses or use of filler words.

Repetition:

- The participant does not repeat phrases or ideas excessively. Some repetition of words occurs, but it is not significant.

Lexical complexity:

- Based on the provided dialogue, it is difficult to assess the lexical complexity accurately. However, the participant uses a mix of simple and descriptive words, which suggests a moderate level of lexical complexity.

Limited vocabulary:

- The participant does not exhibit a significant limitation in vocabulary. They use a variety of words to convey their ideas.

Moving on to the semantics features:

Semantic density:

- To determine semantic density, we need to analyze the ratio of content words (nouns, verbs, adjectives, adverbs) to function words (prepositions, pronouns, conjunctions, etc.). In this short dialogue, the content words are: "little," "girl," "reaching," "waiting," "brother," "give," "cookie," "boy," "knocked," "lid," "tumbling," "stool," "uh," "drying," "dishes," "left," "hand," "touching," "another," "right," "want," "say," "tell," "story," "remember," "uncle," "Bill," "Johnny," "admired," "taken," "camping," "trip." The function words are: "alright," "oh," "well," "the," "is," "for," "her," "to," "a," "from," "but," "he's," "off," "and," "he," "out_o(f)," "and," "has," "in," "with," "you," "only," "that's," "about," "it," "can," "me," "what," "was," "anything," "well," "because," "had," "him," "on," "a." The ratio of content words to function words is approximately 31:34.

Inability to convey meaning:

- The participant is able to describe the scene clearly and convey their ideas without difficulty.

Semantic coherence:

- The participant's description of the scene is logically consistent and meaningful. The relationships between words, phrases, and sentences are coherent and cohesive.

Lack of coherence:

- The participant maintains a clear focus in the conversation and organizes their thoughts coherently.

Moving on to the syntax and grammar features:

Syntactic complexity:

- In this short dialogue, the sentence structures used by the participant are relatively simple. There are no complex sentence structures or subordinate clauses.

Simplified sentence structure:

- The participant primarily uses simple sentence structures.

Grammatical components:

- The participant demonstrates correct usage of grammar, with no apparent errors in subject-verb agreement, tense consistency, or the use of pronouns, prepositions, and conjunctions.

Grammatical errors:

- The participant does not make any significant grammatical errors. Their sentences are grammatically correct.

Based on the analysis of the provided dialogue, the participant does not exhibit significant difficulties with vocabulary, sentence structure, repetition, word-finding, coherence, or grammar. Therefore, based on this limited information, the participant is likely in the Probably Normal group rather than the Probably Mild Cognitive Impairment group. However, it's important to note that this initial screening is not definitive, and a comprehensive judgment would require further evaluation and assessment.
