## Supplementary material for "Text dialogue analysis Based ChatGPT for Primary Screening of Mild Cognitive Impairment": Data(GPT4_vs_GPT3.5_model1-5): 106_GPT3.5 (3).docx

ChatGPT

Based on the analysis of the provided dialogue, here is an evaluation of the indicators and severity ratings for the participant's language features:

Vocabulary Features:

1. Limited vocabulary: Severity: 3 - The participant demonstrates a fairly rich vocabulary with the use of descriptive words and specific names.
2. Increased verbosity: Severity: 2 - The participant does not exhibit excessive verbosity or unnecessary use of words.
3. Overlearned phrases: Severity: 1 - The participant does not overuse cliches or stock phrases.
4. Improper use of pronouns: Severity: 1 - The participant correctly uses pronouns and does not mix them up.
5. Hesitation (Word-finding difficulties): Severity: 2 - The participant exhibits minimal hesitation or filler words.
6. Circumlocution (Word-finding difficulties): Severity: 1 - The participant does not exhibit significant word-finding difficulties or excessive use of vague or evasive language.
7. Tip-of-the-tongue phenomenon: Severity: 1 - The participant does not demonstrate partial recall or difficulty in retrieving specific words.
8. Repetition: Severity: 1 - The participant does not repeat stories, questions, or statements excessively.

Syntax and Grammar Features:

1. Simplified sentence structure and grammar: Severity: 1 - The participant demonstrates a balanced usage of simple and complex sentence structures.
2. Grammatical errors: Severity: 1 - The participant makes only a few grammatical errors.
3. Difficulty with complex ideas: Severity: 1 - The participant handles the narrative and questions without significant difficulty.

Semantics Features:

1. Semantic paraphasias: Severity: 1 - The participant expresses ideas clearly without errors in word choice.
2. Lack of coherence: Severity: 1 - The participant demonstrates coherence and organization in their storytelling.
3. Consistency in errors: Severity: 1 - The participant does not exhibit consistent language errors.
4. Lack of semantic fluency: Severity: 1 - The participant has no difficulty in naming items or describing processes.
5. Disorganized narrative structure: Severity: 1 - The participant maintains a coherent line of thought in their narrative.
6. Orientation issues: Severity: 1 - The participant shows no signs of confusion about date, time, place, or sequence of events.
7. Decline in conversational engagement and responsiveness: Severity: 1 - The participant remains engaged and responsive in conversation.
8. Memory issues: Severity: 1 - The participant does not exhibit significant memory issues or difficulty in recalling information.
9. Lack of abstract thinking and synthesis: Severity: 1 - The participant demonstrates the ability to understand and express abstract concepts.
10. Abnormal emotions: Severity: 1 - The participant's speech tone, style, and emotional state appear normal.

Based on the synthesis of these indicators, the participant's language features do not indicate significant signs of mild cognitive impairment (MCI). Therefore, the participant is likely to be categorized in the Probably Normal Cognition Group.
