## Supplementary material for "Text dialogue analysis Based ChatGPT for Primary Screening of Mild Cognitive Impairment": Data(GPT4_vs_GPT3.5_model1-5): 106_GPT3.5 (4).docx

Please assess a participant with possible mild cognitive impairment (MCI) based on their language features. Note that I do not need to diagnose the participant but rather analyze and rate the severity of each evaluation indicator on a scale of 1 to 10 (1 being the least severe, and 10 being the most severe). Vocabulary features: a. Analyze word frequency, word ratio, phrase frequency, and phrase ratio. b. Calculate the lexical complexity. c. Assess and rate the severity of the following aspects: limited vocabulary, increased verbosity, overlearned phrases, improper use of pronouns, hesitation, circumlocution, tip-of-the-tongue phenomenon, and repetition. Syntax and grammar features: a. Analyze syntactic complexity and grammatical components. b. Assess and rate the severity of the following aspects: simplified sentence structure and grammar, grammatical errors, and difficulty with complex ideas. Semantics features: a. Analyze semantic density and coherence. b. Assess and rate the severity of the following aspects: semantic paraphasias, lack of coherence, consistency in errors, lack of semantic fluency, disorganized narrative structure, orientation issues, decline in conversational engagement and responsiveness, memory issues, lack of abstract thinking and synthesis, and abnormal emotions. Based on the comprehensive synthesis of all the indicators, classify the participant into either the Probably Normal Cognition Group or the Probably Mild Cognitive Impairment Group. Keep in mind that these issues might be present in individuals with normal cognition and that not all individuals with MCI will display all of these features. The final categorization should be based on a comprehensive synthesis of all the indicators. Please note: This evaluation isn't meant to provide a formal diagnosis but to serve as an initial screening tool for potential MCI. INVESTIGATOR: name me any action . INVESTIGATOR: anything you see in the picture . PARTICIPANT: alright . INVESTIGATOR: okay . PARTICIPANT: action . PARTICIPANT: oh well the little girl is reaching waiting for her brother to give her a cookie . PARTICIPANT: the boy has a cookie from a cookie jar but he's knocked the lid off and he is tumbling off a stool . PARTICIPANT: and …uh m:other is …uh drying dishes but she has left the spigot on and …uh water's pouring out_o(f) the sink . PARTICIPANT: and the little girl has her finger up to her mouth . PARTICIPANT: and a boy has one cookie in his hand for the girl in his left hand . PARTICIPANT: and he is just touching another cookie with his right hand . PARTICIPANT: you only want action you say ? INVESTIGATOR: mhm . PARTICIPANT: and that's about it . INVESTIGATOR: okay . INVESTIGATOR: okay can you tell me what the story was about ? INVESTIGATOR: anything you remember about that story . PARTICIPANT: well uncle Bill and Johnny . PARTICIPANT: and uncle … …uh Johnny admired uncle Bill because uncle Bill had taken him on a camping trip .

ChatGPT

Let's analyze the participant's language features based on the provided conversation and evaluate the severity of each indicator:

Vocabulary Features: a. Word frequency, word ratio, phrase frequency, and phrase ratio: We don't have access to the complete conversation, so we cannot calculate these measures.

b. Lexical complexity: Unable to assess without a broader context.

c. Severity Assessment:

- Limited vocabulary: 3
- Increased verbosity: 2
- Overlearned phrases: 1
- Improper use of pronouns: 1
- Hesitation: 1
- Circumlocution: 2
- Tip-of-the-tongue phenomenon: 1
- Repetition: 2

Syntax and Grammar Features: a. Syntactic complexity and grammatical components: The provided conversation consists mostly of simple and coherent sentences with no apparent syntactic complexity issues.

b. Severity Assessment:

- Simplified sentence structure and grammar: 2
- Grammatical errors: 1
- Difficulty with complex ideas: N/A (not observed in the conversation)

Semantics Features: a. Semantic density and coherence: The participant's responses show relatively coherent descriptions, though the overall semantic density cannot be fully assessed without more context.

b. Severity Assessment:

- Semantic paraphasias: N/A (not observed in the conversation)
- Lack of coherence: 1
- Consistency in errors: N/A (not observed in the conversation)
- Lack of semantic fluency: N/A (not observed in the conversation)
- Disorganized narrative structure: 1
- Orientation issues: N/A (not observed in the conversation)
- Decline in conversational engagement and responsiveness: N/A (not observed in the conversation)
- Memory issues: N/A (not observed in the conversation)
- Lack of abstract thinking and synthesis: N/A (not observed in the conversation)
- Abnormal emotions: N/A (not observed in the conversation)

Based on the available indicators, let's categorize the participant:

Probably Normal Cognition Group: The participant exhibits some minor issues in limited vocabulary, verbosity, circumlocution, and repetition. There are no apparent syntactic complexity or grammatical errors. The semantic features show a relatively coherent narrative structure, although there might be some minor lack of coherence.

Please note that this assessment is based on a limited portion of the conversation, and a comprehensive evaluation should consider a more extensive and diverse language sample, as well as other cognitive assessments, for a more accurate classification of cognitive impairment
