## Supplementary material for "Text dialogue analysis Based ChatGPT for Primary Screening of Mild Cognitive Impairment": Data(GPT4_vs_GPT3.5_model1-5): 106_GPT3.5 (5).docx

ChatGPT

3 / 3

Based on the provided conversation between the investigator and the participant, we can assess the participant's language features to determine the severity of potential mild cognitive impairment (MCI). Let's evaluate each indicator and rate its severity on a scale of 1 to 10.

Vocabulary Features: a. Analyzing word frequency, word ratio, phrase frequency, and phrase ratio can provide insights into the participant's lexical complexity and use of vocabulary. Since the text provided is limited, we can only perform a basic analysis. We'll rate the severity based on the available information.

- Word frequency and Word ratio: Due to the limited text, it's challenging to assess word frequency accurately. However, we can note that the participant uses relatively simple and common words, with a lack of specific names or nouns. Severity: 3/10
- Phrase frequency and Phrase ratio: Again, due to the limited text, it's challenging to evaluate phrase frequency accurately. However, the participant does use a few descriptive phrases. Severity: 3/10
- Lexical complexity: The vocabulary used by the participant is relatively straightforward, without many rare or sophisticated words. Severity: 4/10

b. Tip-of-the-tongue phenomenon: The participant does not display explicit signs of the tip-of-the-tongue phenomenon in this conversation. Severity: 1/10

Syntax and Grammar Features: a. Syntactic complexity and grammatical components provide insights into the participant's sentence structure and grammatical usage. Since the text is short, we can only perform a basic analysis.

- Syntactic complexity: The sentences used by the participant are relatively simple, with straightforward structures and minimal use of subordinate clauses. Severity: 2/10
- Grammatical components: The participant demonstrates reasonable usage of grammar, with no glaring errors in subject-verb agreement, tense consistency, or pronoun usage. Severity: 2/10

b. Difficulty with complex ideas: The participant struggles with complex ideas and narratives, as indicated by their response to the investigator's question about the story. Severity: 6/10

Semantics Features: a. Semantic density and coherence assess the meaningfulness and logical consistency of the participant's language. Again, due to the limited text, we can only provide a basic analysis.

- Semantic density: The participant's responses contain a decent balance between content words and function words, indicating some level of semantic density. Severity: 5/10
- Semantic coherence: The participant's responses generally display coherence and cohesion, with reasonable relationships between words, phrases, and sentences. Severity: 3/10

b. Memory issues: The participant struggles with memory, as evidenced by their difficulty in recalling details of the story or remembering major plot points or characters. Severity: 8/10

Based on the comprehensive synthesis of all the indicators, we can classify the participant into the Probably Mild Cognitive Impairment Group. The severity ratings for each indicator contribute to this classification, suggesting potential difficulties in vocabulary, syntax and grammar, comprehension of complex ideas, and memory recall. However, it's important to note that this assessment is based on a limited conversation, and a formal diagnosis would require a more comprehensive evaluation by a medical professional.
