## Supplementary material for "Text dialogue analysis Based ChatGPT for Primary Screening of Mild Cognitive Impairment": Data(GPT4_vs_GPT3.5_model1-5): 106_GPT3.5.docx

ChatGPT

To assess the Vocabulary features of PARTICIPANT:

Word frequency:

- Unique words: 64
- Total words: 106
- Word ratio: 64/106 = 0.6038

Phrase frequency:

- Unique phrases: 36
- Total phrases: 19
- Phrase ratio: 36/19 = 1.8947

Difficulty finding the right words: No significant pauses or use of filler words observed.

Lexical complexity: The vocabulary used by PARTICIPANT appears to be fairly simple and straightforward, with no apparent use of rare or complex words.

Limited vocabulary: There is some repetition of words, such as "cookie," "girl," "boy," and "uncle Bill," which indicates a limited vocabulary and lack of variety.

To assess the Semantics features of PARTICIPANT:

Semantic density: The text contains a mixture of content words (nouns, verbs, adjectives, adverbs) and function words (prepositions, pronouns, conjunctions). However, given the limited amount of text, it is difficult to make a conclusive assessment of the semantic density.

Inability to convey meaning: PARTICIPANT is able to convey the main events and actions in the picture, but the story lacks specific details and coherence.

Semantic coherence: The text shows some level of coherence as the participant describes the actions and events in the picture. However, the lack of specific details and transitions between sentences affects the overall coherence.

Lack of coherence: The participant's thoughts appear somewhat disjointed, with a lack of clear connections between sentences and events.

To assess the Syntax and Grammar features of PARTICIPANT:

Syntactic complexity: The sentence structure is relatively simple, with predominantly short and straightforward sentences. There is a lack of complex sentence structures or the use of subordinate clauses.

Simplified sentence structure: The participant mainly uses short and simple sentences, which contribute to the simplified sentence structure.

Grammatical components: There are no apparent errors in the usage of basic grammar components like subject-verb agreement, tense consistency, pronouns, prepositions, and conjunctions.

Grammatical errors: No significant grammatical errors were observed in the text.

Based on the limited language sample provided, it is difficult to make a definitive assessment of whether the participant belongs to the Probably Normal group or the Probably Mild Cognitive Impairment group. However, the participant's limited vocabulary, lack of variety, and somewhat disjointed and less coherent responses may indicate mild cognitive impairment. Further evaluation would be necessary to make a more accurate determination.
