## Supplementary material for "Text dialogue analysis Based ChatGPT for Primary Screening of Mild Cognitive Impairment": Data(GPT4_vs_GPT3.5_model1-5): 106_GPT4 (2).docx

INVESTIGATOR: name me any action . INVESTIGATOR: anything you see in the picture . PARTICIPANT: alright . INVESTIGATOR: okay . PARTICIPANT: action . PARTICIPANT: oh well the little girl is reaching waiting for her brother to give her a cookie . PARTICIPANT: the boy has a cookie from a cookie jar but he's knocked the lid off and he is tumbling off a stool . PARTICIPANT: and …uh m:other is …uh drying dishes but she has left the spigot on and …uh water's pouring out_o(f) the sink . PARTICIPANT: and the little girl has her finger up to her mouth . PARTICIPANT: and a boy has one cookie in his hand for the girl in his left hand . PARTICIPANT: and he is just touching another cookie with his right hand . PARTICIPANT: you only want action you say ? INVESTIGATOR: mhm . PARTICIPANT: and that's about it . INVESTIGATOR: okay . INVESTIGATOR: okay can you tell me what the story was about ? INVESTIGATOR: anything you remember about that story . PARTICIPANT: well uncle Bill and Johnny . PARTICIPANT: and uncle … …uh Johnny admired uncle Bill because uncle Bill had taken him on a camping trip .

ChatGPT

Let's analyze this transcript following the established criteria.

**Vocabulary Features:**

- Word frequency and Word ratio: The participant appears to use a varied range of unique words throughout the conversation.
- Phrase frequency and Phrase ratio: Similarly, the participant uses a variety of unique phrases.
- Word-finding difficulties: There are a few instances of the participant using filler words like "uh," indicating potential word-finding difficulties.
- Repetition: There is no significant repetition in the participant's responses.
- Lexical complexity: The participant uses both simple and complex words, indicating a moderate level of lexical complexity.
- Limited vocabulary: The participant does not seem to excessively repeat the same words or rely heavily on simple words, indicating a balanced vocabulary.

**Semantic Features:**

- Semantic density: The participant uses a good balance of content and function words, indicating an average semantic density.
- Inability to convey meaning: The participant is generally able to express their ideas clearly.
- Semantic coherence: The participant maintains coherence in their conversation, with logically consistent and meaningful phrases and sentences.
- Lack of coherence: There is no significant lack of coherence observed in the participant's speech.

**Syntax and Grammar Features:**

- Syntactic complexity: The participant uses a variety of sentence structures, indicating a moderate level of syntactic complexity.
- Simplified sentence structure: The participant does not overly simplify their sentences, indicating a balanced use of sentence structures.
- Grammatical components: The participant generally uses correct grammar, with appropriate use of subject-verb agreement, tense consistency, and pronouns, prepositions, and conjunctions.
- Grammatical errors: There are a few grammatical errors, but they are not significant or frequent.

Based on this analysis, the participant could be assigned to the "Probably Normal" group. The participant exhibits a few signs of word-finding struggles but otherwise maintains semantic coherence, uses correct grammar, and has no significant repetition. However, this determination is not definitive and would require a more comprehensive evaluation by a professional.
