## Supplementary material for "Text dialogue analysis Based ChatGPT for Primary Screening of Mild Cognitive Impairment": Data(GPT4_vs_GPT3.5_model1-5): 106_GPT4 (3).docx

ChatGPT

Thank you for the thorough framework. Below is the analysis and evaluation of the PARTICIPANT's responses:

**Vocabulary Features**:

- Word frequency and Word ratio: The word "cookie" is repeated 3 times, indicating some repetition. Also, usage of pronouns like "he", "her", and "his" is high but doesn't seem excessive compared to nouns.
- Phrase frequency and Phrase ratio: Unique phrases were used and no repeated phrase is seen in the conversation.
- Lexical complexity: The vocabulary used is relatively simple.

Assessment:

1. Limited vocabulary: The vocabulary appears relatively limited. Rating: 4/10.
2. Increased verbosity: There isn't significant verbosity in the response. Rating: 3/10.
3. Overlearned phrases: No overlearned phrases or cliches are used. Rating: 1/10.
4. Improper use of pronouns: There isn't a significant issue with pronoun use. Rating: 2/10.
5. Hesitation (Word-finding difficulties): There are several instances of filled pauses and hesitations. Rating: 6/10.
6. Circumlocution (Word-finding difficulties): There is some circumlocution, but it's not excessive. Rating: 4/10.
7. Tip-of-the-tongue phenomenon: This is not clearly observed. Rating: 2/10.
8. Repetition: Some repetition is noticed. Rating: 5/10.

**Syntax and Grammar Features**:

- Syntactic complexity: The sentences are not too complex, though multiple clauses are used.
- Grammatical components: There are no significant errors in grammar.

Assessment:

1. Simplified sentence structure and grammar: The sentence structures are relatively simple. Rating: 5/10.
2. Grammatical errors: There are no noticeable grammatical errors. Rating: 1/10.
3. Difficulty with complex ideas: The participant does seem to have some trouble with recalling complex ideas. Rating: 5/10.

**Semantics Features**:

- Semantic density: The amount of meaningful information is moderate.
- Semantic coherence: The participant's speech appears to be coherent, even though there are moments of hesitation and repetitiveness.

Assessment:

1. Semantic paraphasias: There aren't significant semantic paraphasias. Rating: 2/10.
2. Lack of coherence: Despite hesitations, the overall narrative is coherent. Rating: 3/10.
3. Consistency in errors: Errors do not seem consistent. Rating: 2/10.
4. Lack of semantic fluency: There are instances of semantic fluency problems. Rating: 5/10.
5. Disorganized narrative structure: The narrative structure is moderately organized. Rating: 4/10.
6. Orientation Issues: No clear orientation issues. Rating: 1/10.
7. Decline in conversational engagement and responsiveness: Some decline in conversational engagement is noticed. Rating: 4/10.
8. Memory issues: There are some signs of memory issues. Rating: 5/10.
9. Lack of abstract thinking and synthesis: This is not clearly observed. Rating: 2/10.
10. Abnormal emotions: This is not clearly observed. Rating: 1/10.

Based on this preliminary analysis, PARTICIPANT may fall into the Probably Mild Cognitive Impairment Group. There are noticeable difficulties in some areas, such as hesitations and some memory issues.
