## Supplementary material for "Text dialogue analysis Based ChatGPT for Primary Screening of Mild Cognitive Impairment": Data(GPT4_vs_GPT3.5_model1-5): 106_GPT4 (5).docx

ChatGPT

2 / 2

Based on the text provided, I'll provide an assessment of this participant's language features.

Vocabulary features: a. Word frequency, word ratio, phrase frequency, and phrase ratio seem relatively average for the length of the text provided. Pronoun to specific noun ratio appears slightly increased, indicating potential issues with specificity. The participant used a diverse range of words and phrases, but a formal calculation of lexical complexity isn't possible with the provided data. Estimated severity: 4. b. There is some evidence of the tip-of-the-tongue phenomenon, for instance with the "...uh" hesitations. Estimated severity: 4.

Syntax and grammar features: a. Syntactic complexity seems average. The participant uses a range of sentence structures including some use of subordinate clauses. Grammatical components are largely correct with occasional omissions ("he is tumbling off a stool"). Estimated severity: 3. b. The participant demonstrates some difficulty with complex ideas. Their recounting of the story is not as detailed or coherent as it could be. Estimated severity: 5.

Semantics features: a. Semantic density appears average. There's a good balance of content and function words, and the participant effectively conveys a variety of scenes. Semantic coherence is generally good, though there are some oddities in how the participant describes certain scenarios. Estimated severity: 4. b. The participant appears to have some difficulty recalling details of the story involving Uncle Bill and Johnny. They recount only a few aspects and fail to provide a comprehensive summary. This could indicate memory issues. Estimated severity: 6.

Comprehensive synthesis: Based on this analysis, the participant appears to fall into the Probably Mild Cognitive Impairment Group. Their language use indicates potential issues with vocabulary specificity, tip-of-the-tongue phenomenon, complexity of ideas, semantic coherence, and especially memory recall.

However, it's important to remember that this is a preliminary assessment based on a limited amount of text and might not accurately reflect the participant's overall cognitive function. These aspects should be further investigated and monitored over time. A formal diagnosis should only be made by a qualified healthcare professional based on a comprehensive examination.
