## Supplementary material for "Text dialogue analysis Based ChatGPT for Primary Screening of Mild Cognitive Impairment": Data(GPT4_vs_GPT3.5_model1-5): 106_GPT4.docx

ChatGPT

Assessment of PARTICIPANT's language features:

1. Vocabulary features:

- Word frequency: 80 unique words
- Word ratio: 80 unique words / 158 total words = 0.5063
- Phrase frequency: 37 unique phrases
- Phrase ratio: 37 unique phrases / 37 total phrases = 1
- Difficulty finding the right words: Several instances of "uh" suggesting some difficulty
- Lexical complexity: Moderate, with some repetition and a few less common words
- Limited vocabulary: Slightly limited, with some repetition and simple word choices

2. Semantics features:

- Semantic density: Moderate, with a balance between content and function words
- Inability to convey meaning: Minimal, as the participant is mostly able to express ideas clearly
- Semantic coherence: Generally coherent, with some minor issues in describing the scene
- Lack of coherence: Minimal, as the participant is able to maintain a logical flow of ideas

3. Syntax and Grammar features:

- Syntactic complexity: Moderate, with a mix of short and complex sentences
- Simplified sentence structure: Some instances of short and simple sentences
- Grammatical components: Mostly correct usage of grammar
- Grammatical errors: Minimal, with only a few mistakes

Given the assessment of the language features, it is difficult to confidently assign the participant to a specific group. However, considering the slight limitations in vocabulary and the instances of difficulty finding the right words, I would lean towards placing the PARTICIPANT in the Probably Mild Cognitive Impairment group. This decision is based on the subtle language gaps observed, but further assessment would be necessary for a more accurate diagnosis
