## Supplementary material for "Text dialogue analysis Based ChatGPT for Primary Screening of Mild Cognitive Impairment": Data(GPT4_vs_GPT3.5_model1-5): 107_GPT3.5 (4).docx

ChatGPT

Based on the provided language features and indicators, we can assess and rate the severity of each evaluation indicator on a scale of 1 to 10, with 1 being the least severe and 10 being the most severe. Please note that this analysis is based solely on the language features and does not provide a formal diagnosis. Additionally, it's important to consider that these issues may be present in individuals with normal cognition, and not all individuals with mild cognitive impairment (MCI) will display all of these features.

Vocabulary Features: a. Analyzing word frequency, word ratio, phrase frequency, and phrase ratio: N/A (Not provided) b. Calculating lexical complexity: N/A (Not provided) c. Assessing and rating the severity of the following aspects:

- Limited vocabulary: 3
- Increased verbosity: 3
- Overlearned phrases: 2
- Improper use of pronouns: 1
- Hesitation: 1
- Circumlocution: 2
- Tip-of-the-tongue phenomenon: 1
- Repetition: 1

Syntax and Grammar Features: a. Analyzing syntactic complexity and grammatical components: N/A (Not provided) b. Assessing and rating the severity of the following aspects:

- Simplified sentence structure and grammar: 2
- Grammatical errors: 1
- Difficulty with complex ideas: 2

Semantics Features: a. Analyzing semantic density and coherence: N/A (Not provided) b. Assessing and rating the severity of the following aspects:

- Semantic paraphasias: N/A (Not provided)
- Lack of coherence: 2
- Consistency in errors: 1
- Lack of semantic fluency: 2
- Disorganized narrative structure: 2
- Orientation issues: 1
- Decline in conversational engagement and responsiveness: 1
- Memory issues: 1
- Lack of abstract thinking and synthesis: 2
- Abnormal emotions: N/A (Not provided)

Based on the comprehensive synthesis of the indicators provided, we can classify the participant into the Probably Mild Cognitive Impairment Group. The participant displays several mild to moderate indicators of cognitive impairment across vocabulary, syntax and grammar, and semantics features. However, a formal diagnosis would require a comprehensive evaluation by a qualified healthcare professional.
