## Supplementary material for "Text dialogue analysis Based ChatGPT for Primary Screening of Mild Cognitive Impairment": Data(GPT4_vs_GPT3.5_model1-5): 107_GPT3.5 (5).docx

ChatGPT

2 / 2

Based on the provided language features, here is an assessment of the participant's possible mild cognitive impairment (MCI) based on each evaluation indicator:

Vocabulary features: a. Analyzing word frequency, word ratio, phrase frequency, and phrase ratio requires a larger sample of text. The provided text is relatively short and limited, making it difficult to assess these indicators accurately.

b. The tip-of-the-tongue phenomenon is not explicitly evident in the participant's responses, as they are able to recall and describe the scene with some level of detail. Therefore, it is not possible to rate the severity of this aspect based on the given text.

Syntax and grammar features: a. Analyzing syntactic complexity and grammatical components requires a larger sample of text. The provided text consists of short phrases and sentences, limiting the analysis of these indicators.

b. The participant does not exhibit any noticeable difficulty with complex ideas in the given text, as they are able to comprehend and respond appropriately to the questions asked.

Semantics features: a. Analyzing semantic density and coherence requires a larger sample of text. The provided text is relatively short and limited, making it difficult to assess these indicators accurately.

b. The participant does not display any significant memory issues in the given text. They are able to recall details of the scene, describe the actions, and provide relevant information when prompted.

Based on the limited information available, it is challenging to make a conclusive assessment of the participant's cognitive status. However, based on the provided text, there are no clear indications of severe impairments in vocabulary, syntax, grammar, semantics, or memory. Therefore, based on the available information, the participant is more likely to be classified into the Probably Normal Cognition Group rather than the Probably Mild Cognitive Impairment Group. It is important to note that this assessment is based solely on the provided text and should not be considered a formal diagnosis. A comprehensive evaluation by a healthcare professional is necessary for an accurate assessment of cognitive impairment.
