## Supplementary material for "Text dialogue analysis Based ChatGPT for Primary Screening of Mild Cognitive Impairment": Data(GPT4_vs_GPT3.5_model1-5): 107_GPT3.5.docx

PARTICIPANT's word frequency assessment:

- "the": 18
- "is": 12
- "and": 11
- "in": 9
- "a": 8
- "to": 7
- "of": 6
- "cookies": 5
- "boy": 4
- "mother": 4

Word ratio: The word ratio is the proportion of unique words to the total number of words in the text.

PARTICIPANT's word ratio assessment: Total number of words: 119 Number of unique words: 67 Word ratio: 67/119 ≈ 0.563

Phrase frequency: To assess phrase frequency, we count the occurrences of each unique phrase in the text. Unique phrases are distinct combinations of words that appear in the text, not taking into account their repetitions.

PARTICIPANT's phrase frequency assessment:

- "the mother is": 4
- "the window is": 2
- "the curtains are": 2
- "the little girl is": 1
- "it seems to be": 1
- "there's grass outside": 1
- "mother's finished certain": 1
- "the mother seems to have": 1
- "the children look to be": 1
- "they're dressed for summer": 1

Phrase ratio: The phrase ratio is the proportion of unique phrases to the total number of phrases in the text.

PARTICIPANT's phrase ratio assessment: Total number of phrases: 25 Number of unique phrases: 10 Phrase ratio: 10/25 = 0.4

Difficulty finding the right words: There are no significant pauses or use of filler words in the conversation, indicating that the participant does not exhibit difficulty finding the right words.

Lexical complexity: The participant's vocabulary shows some variety and includes descriptive words such as "gentle," "tidy," and "obviously." However, the use of vocabulary is generally simple and lacks sophistication.

Limited vocabulary: The participant tends to repeat certain words such as "the," "is," and "and." Additionally, there is a lack of diversity and sophistication in the vocabulary used.

Assessment of Semantics features:

Semantic density: The participant's responses contain a reasonable amount of meaningful information, with the inclusion of content words such as nouns ("sink," "stool," "cookies"), verbs ("washing," "drying"), and adjectives ("open," "gentle").

Inability to convey meaning: The participant is generally able to express ideas clearly, with no significant difficulty in conveying meaning.

Semantic coherence: The participant's responses demonstrate logical consistency and meaningfulness. The relationships between words, phrases, and sentences are coherent, and there is a clear understanding of the scene depicted in the picture.

Lack of coherence: There is no evident lack of coherence in the participant's responses. The thoughts flow logically and appear related to each other.

Assessment of Syntax and Grammar features:

Syntactic complexity: The participant's sentences are mostly simple, with few instances of complex sentence structures. There is a frequent use of coordinating conjunctions ("and") to connect ideas. The average length of sentences is relatively short.

Simplified sentence structure: The participant predominantly uses short and simple sentences, with a lack of complex sentence structures.

Grammatical components: The participant demonstrates overall correct usage of grammar, including subject-verb agreement, tense consistency, and proper use of pronouns, prepositions, and conjunctions.

Grammatical errors: There are no significant grammatical errors observed in the participant's responses.

Assignment:

Based on the provided conversation, the participant does not exhibit any significant language gaps or subtle cognitive impairments. Therefore, the participant can be assigned to the Probably Normal group.
