## Supplementary material for "Text dialogue analysis Based ChatGPT for Primary Screening of Mild Cognitive Impairment": Data(GPT4_vs_GPT3.5_model1-5): 107_GPT4 (2).docx

ChatGPT

Let's analyse the PARTICIPANT's language performance based on the given parameters.

1. Vocabulary:
   - Word frequency and ratio: While a complete count of unique words and their frequencies would be impossible here, the participant does seem to use a varied vocabulary with several unique words and phrases. The repeated words are mainly common words which are to be expected in a conversational setting.
   - Phrase frequency and ratio: Again, while it's not feasible to count the unique phrases and their frequencies, the participant seems to use a variety of unique phrases, indicating no severe cognitive impairment.
   - Word-finding difficulties: The participant does use fillers like "uh" and "um" fairly often, indicating some difficulty in finding words.
   - Repetition: There is some repetition in the participant's statements, but it doesn't seem overly frequent or disruptive.
   - Lexical complexity: The participant uses a mix of simple and complex words, indicating a reasonable lexical complexity.
   - Limited vocabulary: The participant doesn't seem to overly rely on a limited set of words or phrases.
2. Semantics:
   - Semantic density: The participant's statements seem to be fairly dense with content words.
   - Inability to convey meaning: The participant is generally successful in conveying meaning, although there are some instances where the message might not be completely clear.
   - Semantic coherence: The participant maintains reasonable coherence throughout the discussion, with ideas logically connected and consistent.
   - Lack of coherence: The participant does not show a significant lack of coherence.
3. Syntax and Grammar:
   - Syntactic complexity: The participant uses a mix of simple and complex sentence structures, indicating a reasonable level of syntactic complexity.
   - Simplified sentence structure: Although some sentences are simple, the participant does not exclusively rely on overly simplified structures.
   - Grammatical components: The participant mostly uses grammar correctly, with subject-verb agreement, tense consistency, and proper use of pronouns, prepositions, and conjunctions.
   - Grammatical errors: There are a few instances of grammatical errors or inconsistencies, such as "…uh the children look to be almost about the same size."

Given the above analysis, the participant does show some signs of mild cognitive impairment, such as word-finding difficulties, usage of fillers, and minor grammatical errors. However, these signs are not significantly disruptive to their overall communication ability. They maintain a reasonable level of vocabulary, semantic density, and syntactic complexity. Therefore, based on this language sample, the participant would probably be classified under the Probably Normal group. It's worth noting that this is a preliminary assessment and further comprehensive evaluation might be needed to confirm the cognitive status of the participant
