## Supplementary material for "Text dialogue analysis Based ChatGPT for Primary Screening of Mild Cognitive Impairment": Data(GPT4_vs_GPT3.5_model1-5): 107_GPT4 (3).docx

ChatGPT

Based on the text provided, below is a high-level evaluation of the possible cognitive impairment for the PARTICIPANT.

Vocabulary Features:

1. Limited Vocabulary: Score 2/10. There's a variety in words used, however, there is repetition of certain terms such as "mother", "dishes", "cookies", "kitchen", etc.
2. Increased Verbosity: Score 2/10. The verbosity is acceptable, with adequate and necessary details provided to describe the picture.
3. Overlearned Phrases: Score 1/10. No notable overuse of cliches or stock phrases observed.
4. Improper Use of Pronouns: Score 1/10. The use of pronouns seems correct and appropriate.
5. Hesitation (Word-finding difficulties): Score 3/10. Frequent use of "uh" and "um" was observed.
6. Circumlocution (Word-finding difficulties): Score 2/10. There are no significant instances of roundabout conversation.
7. Tip-of-the-tongue Phenomenon: Score 1/10. No notable instances of partial recall.
8. Repetition: Score 2/10. Some repetitions (like "mother is washing dishes") but not excessive.

Syntax and Grammar Features:

1. Simplified Sentence Structure and Grammar: Score 3/10. The sentences used were generally simple.
2. Grammatical Errors: Score 1/10. There is a lack of significant grammatical errors.
3. Difficulty with Complex Ideas: Score 1/10. No difficulties observed in explaining the complex narrative.

Semantics Features:

1. Semantic Paraphasias: Score 1/10. No noticeable difficulty in expressing ideas.
2. Lack of Coherence: Score 1/10. Conversations seem coherent with logical flow.
3. Consistency in Errors: Score 1/10. No consistent errors noticed.
4. Lack of Semantic Fluency: Score 1/10. No trouble noted in describing the scene or events.
5. Disorganized Narrative Structure: Score 2/10. There are some minor issues in organizing the narrative.
6. Orientation Issues: Score 1/10. No confusion observed regarding the context of the picture.
7. Decline in Conversational Engagement and Responsiveness: Score 1/10. PARTICIPANT appears engaged and responsive.
8. Memory Issues: Score 1/10. No apparent memory issues, as PARTICIPANT was able to maintain consistent narrative across conversations.
9. Lack of Abstract Thinking and Synthesis: Score 1/10. PARTICIPANT seems able to synthesize the overall situation.
10. Abnormal Emotions: Score 1/10. No abnormal emotions are observed in the text.

In conclusion, the PARTICIPANT is showing signs of mild cognitive impairment, specifically in areas like hesitation (word-finding difficulties) and a slight disorganized narrative structure. These, however, could also be normal speech characteristics and not necessarily indicative of MCI.

Considering the overall scores and understanding that this analysis is based only on a single sample, the PARTICIPANT is dichotomised into the Probably Normal Cognition Group. However, a more comprehensive analysis involving additional samples and possibly a professional clinical diagnosis would be necessary for a definitive conclusion
