## Supplementary material for "Text dialogue analysis Based ChatGPT for Primary Screening of Mild Cognitive Impairment": Data(GPT4_vs_GPT3.5_model1-5): 107_GPT4 (5).docx

ChatGPT

Based on the provided text, here's the analysis of the participant's language features:

Vocabulary features: a. The participant repeats certain phrases and words, such as "the mother", "the little boy", and "cookie". This results in a lower unique word ratio. The ratio of pronouns to specific names or nouns appears relatively balanced, with both represented in approximately equal amounts. Overall, the lexical complexity seems moderately diverse, with a mix of common and less frequently used words, such as "shrubbery", "cupboards", and "sandals". Lexical complexity score: 6/10 b. There were several instances of the tip-of-the-tongue phenomenon, where the participant struggled to find the correct words or hesitated in the middle of sentences, e.g., "…uh the mother is washing dishes." and "…um mother was daydreaming and forgot and left the water...". Tip-of-the-tongue score: 7/10

Syntax and grammar features: a. The participant's sentences are generally grammatically correct with proper use of verbs, pronouns, prepositions, and conjunctions. However, the syntactic complexity is moderate, with sentences typically composed of one or two clauses. Syntactic complexity score: 5/10 b. There doesn't appear to be any noticeable difficulty with complex ideas. The participant narrates complex scenes with relative clarity, but at times shows some hesitations or redundancies, e.g., "because he did not place his weight correctly on the stool he's about to fall and probably hurt himself". Difficulty with complex ideas score: 3/10

Semantic features: a. Semantic density appears moderately high, as the participant provides meaningful information in most sentences. There's a balance between content words and function words, indicating a fair amount of detailed information. Semantic coherence is relatively good, with a logical sequence of events and the relationships between words, phrases, and sentences seem coherent. Semantic density score: 7/10, Semantic coherence score: 7/10 b. The participant does not seem to exhibit significant memory issues in this text. They don't repeat questions, and they recall a detailed sequence of events and characters. Memory issues score: 2/10

Based on a comprehensive synthesis of all the indicators, the participant would probably be classified in the Probably Normal Cognition Group. They display some features associated with MCI, such as tip-of-the-tongue phenomenon, but overall they maintain a coherent narrative, exhibit complex thinking, and do not show significant memory issues. However, this should be used only as an initial screening tool and not a formal diagnosis.
