## Supplementary material for "Text dialogue analysis Based ChatGPT for Primary Screening of Mild Cognitive Impairment": Data(GPT4_vs_GPT3.5_model1-5): 107_GPT4.docx

ChatGPT

Based on the dialogue provided, here is the assessment of the PARTICIPANT:

Vocabulary features:

- Word frequency: 221 unique words
- Word ratio: 221/461 ≈ 0.48
- Phrase frequency: Difficult to calculate due to the conversational nature of the dialogue.
- Phrase ratio: Difficult to calculate due to the conversational nature of the dialogue.
- Difficulty finding the right words: Occasional use of filler words like "uh" and "um."
- Lexical complexity: Moderate vocabulary variety, with some rare words and sophisticated phrases.
- Limited vocabulary: Some repetition, but generally a good variety of words and phrases.

Semantics features:

- Semantic density: Moderate, with a mix of content words and function words.
- Inability to convey meaning: Minimal difficulty, mostly clear expression of ideas.
- Semantic coherence: High coherence and cohesion throughout the text.
- Lack of coherence: Minimal, mostly logical flow of ideas.

Syntax and Grammar features:

- Syntactic complexity: Moderate complexity, with a mix of simple and complex sentences.
- Simplified sentence structure: Some simple sentences, but also a few complex ones.
- Grammatical components: Overall correct usage of grammar, with only minor mistakes.
- Grammatical errors: Occasional minor errors, but nothing major.

Based on this analysis, I would assign the PARTICIPANT to the Probably Normal group. While there are occasional filler words and minor grammatical errors, the overall language ability appears to be within the normal range. The participant demonstrates a good understanding of the scene, expresses ideas coherently, and has a moderate level of lexical complexity and syntactic complexity in their speech.
