## Supplementary material for "Text dialogue analysis Based ChatGPT for Primary Screening of Mild Cognitive Impairment": Data(GPT4_vs_GPT3.5_model1-5): 108_GPT3.5 (3).docx

ChatGPT

Based on the provided conversation, here is an analysis of the indicators and their severity ratings for the PARTICIPANT:

Vocabulary Features:

- Limited vocabulary: Severity rating: 3
- Increased verbosity: Severity rating: 4
- Overlearned phrases: Severity rating: 2
- Improper use of pronouns: Severity rating: 3
- Hesitation (Word-finding difficulties): Severity rating: 3
- Circumlocution (Word-finding difficulties): Severity rating: 2
- Tip-of-the-tongue phenomenon: Severity rating: 4
- Repetition: Severity rating: 2

Syntax and Grammar Features:

- Simplified sentence structure and grammar: Severity rating: 2
- Grammatical errors: Severity rating: 2
- Difficulty with complex ideas: Severity rating: 2

Semantics Features:

- Semantic paraphasias: Severity rating: 2
- Lack of coherence: Severity rating: 2
- Consistency in errors: Severity rating: 1
- Lack of semantic fluency: Severity rating: 3
- Disorganized narrative structure: Severity rating: 2
- Orientation issues: Severity rating: 2
- Decline in conversational engagement and responsiveness: Severity rating: 3
- Memory issues: Severity rating: 2
- Lack of abstract thinking and synthesis: Severity rating: 2
- Abnormal emotions: Severity rating: 1

Based on the comprehensive synthesis of the indicators, it appears that the PARTICIPANT shows mild cognitive impairment (MCI) symptoms. While some indicators suggest normal cognition, such as grammatical errors and simplified sentence structure, other indicators point towards MCI, such as limited vocabulary, increased verbosity, word-finding difficulties, repetition, lack of coherence, and decline in conversational engagement and responsiveness. The severity ratings suggest a moderate level of impairment overall. However, it's important to note that further evaluation by a medical professional specializing in cognitive assessments is necessary for an accurate diagnosis.
