## Supplementary material for "Text dialogue analysis Based ChatGPT for Primary Screening of Mild Cognitive Impairment": Data(GPT4_vs_GPT3.5_model1-5): 108_GPT3.5 (4).docx

ChatGPT

Based on the language features exhibited by the participant, we can assess and rate the severity of each evaluation indicator as follows:

Vocabulary features: a. Word frequency, word ratio, phrase frequency, and phrase ratio: Not provided in the conversation. b. Lexical complexity: Not provided in the conversation. c. Severity assessment:

- Limited vocabulary: 3/10 - The participant uses a limited range of vocabulary and doesn't demonstrate a wide lexical repertoire.
- Increased verbosity: 4/10 - The participant tends to provide excessive details and elaborations, resulting in verbose speech.
- Overlearned phrases: 3/10 - The participant does not exhibit significant overuse of learned phrases.
- Improper use of pronouns: Not observed in the conversation.
- Hesitation: 2/10 - The participant displays some hesitations and pauses while speaking.
- Circumlocution: Not observed in the conversation.
- Tip-of-the-tongue phenomenon: Not observed in the conversation.
- Repetition: 3/10 - The participant repeats certain information but not to a significant extent.

Syntax and grammar features: a. Syntactic complexity and grammatical components: Not provided in the conversation. b. Severity assessment:

- Simplified sentence structure and grammar: 3/10 - The participant generally uses simple sentence structures and demonstrates basic grammatical competence.
- Grammatical errors: Not observed in the conversation.
- Difficulty with complex ideas: Not observed in the conversation.

Semantics features: a. Semantic density and coherence: Not provided in the conversation. b. Severity assessment:

- Semantic paraphasias: Not observed in the conversation.
- Lack of coherence: 3/10 - The participant's speech lacks smooth coherence and organization at times.
- Consistency in errors: Not observed in the conversation.
- Lack of semantic fluency: 2/10 - The participant experiences occasional difficulties in generating fluent and coherent responses.
- Disorganized narrative structure: 2/10 - The participant's narrative lacks a clear and structured organization.
- Orientation issues: Not observed in the conversation.
- Decline in conversational engagement and responsiveness: Not observed in the conversation.
- Memory issues: Not observed in the conversation.
- Lack of abstract thinking and synthesis: Not observed in the conversation.
- Abnormal emotions: Not observed in the conversation.

Based on the comprehensive synthesis of all the indicators, the participant can be classified into the Probably Normal Cognition Group. While the participant exhibits some language features that may indicate mild cognitive impairment (e.g., limited vocabulary, increased verbosity, lack of coherence), the severity of these features is relatively mild and not definitive of MCI. It's important to note that this assessment is based on a single conversation, and a comprehensive evaluation involving multiple measures would be necessary for a more accurate diagnosis.
