## Supplementary material for "Text dialogue analysis Based ChatGPT for Primary Screening of Mild Cognitive Impairment": Data(GPT4_vs_GPT3.5_model1-5): 108_GPT3.5 (5).docx

ChatGPT

To assess the participant with possible mild cognitive impairment (MCI) based on the provided language features, we will analyze the indicators mentioned in each category and rate their severity on a scale of 1 to 10. Remember that this assessment is not a formal diagnosis but a screening tool for potential MCI.

Vocabulary Features: a. Analyzing word frequency, word ratio, phrase frequency, and phrase ratio:

- Word frequency and word ratio: The participant's use of unique words, pronouns, and specific names/nouns is limited. There is a repetition of certain words and a lack of lexical variety. Rating: 6/10
- Phrase frequency and phrase ratio: The participant uses some unique phrases, but there is a lack of variety and complexity in the phrases. Rating: 5/10
- Lexical complexity: The vocabulary used by the participant is relatively simple, with shorter words and fewer rare words. Rating: 4/10

b. Assessing the severity of the tip-of-the-tongue phenomenon: The participant demonstrates partial recall and difficulty in retrieving complete words, relying on cues like first letters, syllable structure, or semantic categories. Rating: 7/10

Syntax and Grammar Features: a. Analyzing syntactic complexity and grammatical components:

- Syntactic complexity: The participant's sentences are generally straightforward with simple structures. There is little use of subordinate clauses or complex sentence constructions. Rating: 3/10
- Grammatical components: The participant shows some correct usage of grammar, but there are occasional errors in subject-verb agreement, tense consistency, and pronoun usage. Rating: 5/10

b. Assessing the severity of difficulty with complex ideas: The participant experiences difficulty when the narrative or question becomes more complex. They struggle to handle complex ideas. Rating: 8/10

Semantics Features: a. Analyzing semantic density and coherence:

- Semantic density: The participant's use of content words (nouns, verbs, adjectives, adverbs) relative to function words (prepositions, pronouns, conjunctions) is somewhat balanced but lacks depth. Rating: 5/10
- Semantic coherence: The participant's response is generally coherent and logical, with connections between words, phrases, and sentences. However, there are occasional gaps in cohesion. Rating: 6/10

b. Assessing the severity of memory issues: The participant shows signs of memory issues, such as repeating questions, difficulty recalling specific information or sequences, and trouble remembering details from the story. Rating: 7/10

Based on the comprehensive synthesis of all the indicators, the participant can be classified into the Probably Mild Cognitive Impairment Group. While some features may also be present in individuals with normal cognition, the combination of limited vocabulary, tip-of-the-tongue phenomenon, difficulty with complex ideas, and memory issues suggests a higher likelihood of mild cognitive impairment. However, it is essential to consider this evaluation in the context of the participant's overall cognitive function, and a formal diagnosis should be sought from a healthcare professional.
