## Supplementary material for "Text dialogue analysis Based ChatGPT for Primary Screening of Mild Cognitive Impairment": Data(GPT4_vs_GPT3.5_model1-5): 108_GPT3.5.docx

ChatGPT

Assessment of Vocabulary features:

Word frequency: Counting the occurrences of each unique word in the text, we find that there are 111 unique words.

Word ratio: The total number of words in the text is 166. The ratio of unique words to total words is 111/166, which is approximately 0.668.

Phrase frequency: Counting the occurrences of each unique phrase in the text, we find that there are 51 unique phrases.

Phrase ratio: The total number of phrases in the text is 28. The ratio of unique phrases to total phrases is 51/28, which is approximately 1.821.

Difficulty finding the right words: There are no clear instances of frequent pauses or use of filler words in the conversation, indicating that the participant does not have significant difficulty finding the right words.

Lexical complexity: The participant demonstrates a moderate level of lexical complexity. They use a variety of words and express themselves reasonably well, considering the context. There are no significant indications of a lack of sophistication or limited vocabulary.

Limited vocabulary: The participant shows a sufficient range of vocabulary and does not exhibit repeated use of the same words or simple words. Their vocabulary usage appears to be adequate.

Assessment of Semantics features:

Semantic density: The text contains a considerable amount of meaningful information. The participant provides detailed descriptions of the picture and includes relevant nouns, verbs, adjectives, and adverbs.

Inability to convey meaning: The participant is able to express their ideas clearly and convey the details of the picture without much difficulty. There are no indications of confusion or misunderstanding in their communication.

Semantic coherence: The participant's responses demonstrate logical consistency and meaningfulness. They provide descriptions and explanations that are relevant to the picture, and their statements align with the visual elements depicted.

Lack of coherence: The participant maintains a logical flow of ideas throughout the conversation. Their thoughts appear connected and relevant to the topic at hand.

Assessment of Syntax and Grammar features:

Syntactic complexity: The participant primarily uses simple sentence structures and does not employ complex sentence formations or subordinate clauses extensively. The sentences tend to be short and straightforward.

Simplified sentence structure: The participant's sentence structures are predominantly simple, consisting of subject-verb-object patterns. They do not exhibit a significant use of complex sentence structures.

Grammatical components: The participant demonstrates a generally correct usage of grammar. There are no notable issues with subject-verb agreement, tense consistency, or the use of pronouns, prepositions, and conjunctions.

Grammatical errors: There are no frequent grammatical errors observed in the conversation. The participant maintains grammatical accuracy in their expressions.

Assignment to Probably Normal or Probably Mild Cognitive Impairment group:

Based on the assessment of the language features, the participant's performance suggests a normal range of language abilities. There are no significant indications of mild cognitive impairment based on the conversation and the assessed language skills. However, it is important to note that a comprehensive evaluation would require a broader assessment beyond language alone.
