## Supplementary material for "Text dialogue analysis Based ChatGPT for Primary Screening of Mild Cognitive Impairment": Data(GPT4_vs_GPT3.5_model1-5): 108_GPT4 (2).docx

ChatGPT

Let's analyze the PARTICIPANT's language and communication features step by step according to the guidelines provided.

**Vocabulary features:**

- Word frequency and ratio: Due to the limitations of this format, it's not possible for me to provide an exact calculation. However, it seems that the participant uses a variety of words and phrases, which is a good sign.
- Phrase frequency and ratio: Similarly, I can't calculate exact numbers, but the variety of phrases used is quite high, indicating a rich vocabulary.
- Word-finding difficulties: The participant did use filler words like "uh" and "um" frequently and there were pauses, indicating some difficulty in word-finding.
- Repetition: The participant repeated certain information, like the mother doing dishes and the water running out of the sink.
- Lexical complexity: The participant used a variety of words and phrases of moderate complexity, indicating a good vocabulary.
- Limited vocabulary: There wasn't significant limitation in vocabulary observed.

**Semantics features:**

- Semantic density: The participant made use of content words more than function words, indicating a good semantic density.
- Inability to convey meaning: The participant was able to describe the scene quite clearly, indicating no significant inability to convey meaning.
- Semantic coherence: The participant maintained a logical flow of ideas, showing good semantic coherence.
- Lack of coherence: The participant didn't struggle significantly with organizing thoughts coherently.

**Syntax and Grammar features:**

- Syntactic complexity: The participant used sentences of moderate complexity. They didn't use overly simplified structures.
- Simplified sentence structure: The participant didn't exhibit a significant lack of complex sentence structures.
- Grammatical components: The participant made correct usage of grammar, including subject-verb agreement, tense consistency, and proper use of pronouns, prepositions, and conjunctions.
- Grammatical errors: The participant didn't make frequent grammatical errors.

Based on this analysis, the PARTICIPANT would likely be assigned to the Probably Normal group. There are some signs of difficulty with word-finding and repetition, but overall, the participant maintains good semantic coherence, uses a rich vocabulary, and makes few grammatical errors. Please note that this is an initial analysis and not a diagnostic tool. A comprehensive evaluation by healthcare professionals is needed to accurately assess cognitive function
