## Supplementary material for "Text dialogue analysis Based ChatGPT for Primary Screening of Mild Cognitive Impairment": Data(GPT4_vs_GPT3.5_model1-5): 108_GPT4 (3).docx

ChatGPT

Thank you for providing the dialogues. Based on the given conversations and criteria for assessment, the following analysis can be made:

Vocabulary features:

1. Limited vocabulary: Severity - PARTICIPANT: 5/10, more frequent use of simple words and common terms.
2. Increased verbosity: Severity - PARTICIPANT: 6/10, occasionally uses more words than necessary.
3. Overlearned phrases: Severity - PARTICIPANT: 2/10, there are no significant overuses of cliches or stock phrases.
4. Improper use of pronouns: Severity - PARTICIPANT: 1/10, pronoun use is normal.
5. Hesitation (Word-finding difficulties): Severity - PARTICIPANT: 7/10, noticeable pauses and filler words like "uh".
6. Circumlocution (Word-finding difficulties): Severity - PARTICIPANT: 5/10, some instances of using many words where fewer would do.
7. Tip-of-the-tongue phenomenon: Severity - PARTICIPANT: 4/10, occasional partial recall.
8. Repetition: Severity - PARTICIPANT: 6/10, repeats descriptions of scenes.

Syntax and Grammar features:

1. Simplified sentence structure and grammar: Severity - PARTICIPANT: 5/10, frequently opts for simpler structures.
2. Grammatical errors: Severity - PARTICIPANT: 2/10, a few errors, but not significant.
3. Difficulty with complex ideas: Severity - PARTICIPANT: 4/10, handles simpler sentences and ideas, struggles slightly when the narrative becomes more complex.

Semantics features:

1. Semantic paraphasias: Severity - PARTICIPANT: 3/10, occasional odd combinations of words.
2. Lack of coherence: Severity - PARTICIPANT: 4/10, generally coherent but some abrupt topic changes.
3. Consistency in errors: Severity - PARTICIPANT: 3/10, some consistent language errors.
4. Lack of semantic fluency: Severity - PARTICIPANT: 4/10, slight trouble describing processes or sequence of events.
5. Disorganized narrative structure: Severity - PARTICIPANT: 4/10, occasional lack of coherent line of thought.
6. Orientation Issues: Severity - PARTICIPANT: 2/10, no significant issues.
7. Decline in conversational engagement and responsiveness: Severity - PARTICIPANT: 4/10, some reduced spontaneity and less initiation of conversation.
8. Memory issues: Severity - PARTICIPANT: 6/10, repeats descriptions and questions.
9. Lack of abstract thinking and synthesis: Severity - PARTICIPANT: 3/10, slight difficulty in synthesizing the overall situation.
10. Abnormal emotions: Severity - PARTICIPANT: 2/10, no significant issues.

Based on the evaluation of all the indicators, the PARTICIPANT can be categorized into the "Probably Mild Cognitive Impairment Group". The higher levels of repetition, verbosity, word-finding difficulties, and memory issues indicate possible signs of mild cognitive impairment. However, this should not be used as a diagnosis and it is recommended that a professional neuropsychological assessment is done for a precise diagnosis
