## Supplementary material for "Text dialogue analysis Based ChatGPT for Primary Screening of Mild Cognitive Impairment": Data(GPT4_vs_GPT3.5_model1-5): 108_GPT4 (4).docx

ChatGPT

Based on the text provided, here is an assessment of the participant's language features:

Vocabulary features: a. Word and phrase frequency seems to be generally normal. There are repeated phrases like "the water's running" and "the mother's doing the dishes," but this might be due to the context of the conversation. b. The lexical complexity appears to be relatively normal, but some simplification is noted. c.

- Limited vocabulary: 3/10 (most words were simple, yet appropriate to the context)
- Increased verbosity: 1/10 (participant did not appear to be excessively verbose)
- Overlearned phrases: 2/10 (common phrases were used, but they weren't excessive or inappropriate)
- Improper use of pronouns: 1/10 (pronouns were appropriately used)
- Hesitation: 5/10 (several instances of hesitation or pausing were observed)
- Circumlocution: 1/10 (circumlocution was not prominent)
- Tip-of-the-tongue phenomenon: 1/10 (participant didn't appear to struggle with word retrieval)
- Repetition: 4/10 (some repetition was present, but it might be due to the nature of the task)

Syntax and grammar features: a. The participant's syntactic complexity and grammatical components appeared to be fairly normal for the task. b.

- Simplified sentence structure and grammar: 3/10 (sentences were fairly simple, but they were complex enough for the task)
- Grammatical errors: 2/10 (few grammatical errors were present)
- Difficulty with complex ideas: 1/10 (the task did not require expressing complex ideas)

Semantics features: a. Semantic density and coherence seemed to be within the normal range. b.

- Semantic paraphasias: 1/10 (participant made no clear substitutions of words with different meanings)
- Lack of coherence: 2/10 (the participant was mostly coherent)
- Consistency in errors: 2/10 (not enough errors to assess consistency)
- Lack of semantic fluency: 2/10 (participant was mostly fluent)
- Disorganized narrative structure: 2/10 (the narrative was relatively organized)
- Orientation issues: 1/10 (no orientation issues were apparent)
- Decline in conversational engagement and responsiveness: 2/10 (participant seemed engaged)
- Memory issues: 3/10 (there was some repetition which may indicate memory issues)
- Lack of abstract thinking and synthesis: 1/10 (the task did not require significant abstract thinking)
- Abnormal emotions: 1/10 (no emotional content to assess)

Based on these indicators, it would be most appropriate to classify this participant in the "Probably Normal Cognition Group". However, some mild hesitation and repetition were observed. Further observation or more complex tasks might be needed for a more accurate classification. As mentioned earlier, this evaluation isn't meant to provide a formal diagnosis but to serve as an initial screening tool for potential MCI
