## Supplementary material for "Text dialogue analysis Based ChatGPT for Primary Screening of Mild Cognitive Impairment": Data(GPT4_vs_GPT3.5_model1-5): 108_GPT4 (5).docx

ChatGPT

Vocabulary features: a. This participant's lexical complexity seems moderate. There is a good number of unique words and phrases, such as "cookie jar," "stool," "cabinets," etc. However, there's some repetition, which might be a sign of memory issues. Most nouns and verbs are simple and commonly used. The word ratio leans heavily towards common words, with limited usage of pronouns and specific names. Overall, I would rate lexical complexity at 5/10. b. In terms of the tip-of-the-tongue phenomenon, the participant shows some signs with a few instances of hesitation ("…uh", "…w well like …", "…ab what else", "…uh …uh sink board", "…wa doing"). I would rate this at 5/10.

Syntax and grammar features: a. The syntactic complexity is relatively low. The sentences are generally short and direct, without many subordinate clauses. The grammar is mostly correct, but there's a notable lack of verb tenses variation (mainly present tense). Therefore, I would rate syntactic complexity at 4/10. b. The participant seems to have little difficulty expressing complex ideas, maintaining the coherence of the narrative despite some difficulty recalling specific details. I would rate the difficulty with complex ideas at 3/10.

Semantics features: a. The semantic density is relatively high, with a large number of content words and meaningful information. The participant describes the scene in the picture with reasonable detail. However, semantic coherence is compromised by occasional repetitions and hesitations. Overall, I would rate semantic density at 7/10 and semantic coherence at 6/10. b. Memory issues are present, shown by repeating phrases ("the water's running out of the sink", "the mother's doing the dishes", "the boy is…stealing the cookies out of the cookie jar", "she has a dish towel in her hand") and asking whether a piece of information was previously stated ("did I say that before?"). I would rate memory issues at 6/10.

In conclusion, based on this language evaluation, the participant could possibly fall into the "Probably Mild Cognitive Impairment Group". Although there are several indicators that are on the less severe end of the scale, some signs (such as memory issues and the tip-of-the-tongue phenomenon) are more prominent and may suggest some degree of cognitive impairment. However, this should not be considered a definitive assessment, as other factors and more thorough evaluations would need to be considered for a formal diagnosis.
