## Supplementary material for "Text dialogue analysis Based ChatGPT for Primary Screening of Mild Cognitive Impairment": Data(GPT4_vs_GPT3.5_model1-5): 108_GPT4.docx

ChatGPT

Assessment:

Vocabulary features:

1. Word frequency: 92 unique words
2. Word ratio: 0.33 (92 unique words / 280 total words)
3. Phrase frequency: 51 unique phrases
4. Phrase ratio: 0.55 (51 unique phrases / 93 total phrases)
5. Difficulty finding the right words: Mild (frequent pauses, use of filler words like "uh" and "um")
6. Lexical complexity: Moderate (some sophisticated vocabulary, but also simple words)
7. Limited vocabulary: Mild (some repetition and simple words)

Semantics features:

1. Semantic density: Moderate (a balance of content and function words)
2. Inability to convey meaning: Mild (some difficulty, but overall able to convey meaning)
3. Semantic coherence: Moderate (generally coherent, but some disjointed thoughts)
4. Lack of coherence: Mild (some difficulty maintaining logical flow, but generally coherent)

Syntax and Grammar features:

1. Syntactic complexity: Low (simple sentence structures, few subordinate clauses)
2. Simplified sentence structure: Predominant (short and simple sentences)
3. Grammatical components: Moderate (some mistakes, but mostly correct usage)
4. Grammatical errors: Mild (occasional errors in tense, subject-verb agreement, etc.)

Based on the assessment, I would assign PARTICIPANT to the Probably Mild Cognitive Impairment group. The participant exhibited mild difficulty finding the right words, some lack of coherence, and simplified sentence structures. While they were able to convey meaning and maintain some level of coherence, the language gaps were noticeable enough to suggest mild cognitive impairment.
