## Supplementary material for "Text dialogue analysis Based ChatGPT for Primary Screening of Mild Cognitive Impairment": Data(GPT4_vs_GPT3.5_model1-5): 109_GPT3.5 (2).docx

Attention! You do not need to diagnose, just follow the steps below to initially screen for PARTICIPANT with possibly mild cognitive impairment (MCI). Some differences between MCI and normal cognition are also given, please analyse them in detail, step by step. Firstly, please assess the Vocabulary features of PARTICIPANT at the Word frequency and Word ratio, Phrase frequency and Phrase ratio, Word-finding difficulties, Repetition, Lexical complexity, Limited vocabulary. Word frequency: Count the occurrences of each unique word in the text. Unique words are distinct words that appear in the text, not taking into account their repetitions. Each word is counted only once, regardless of how many times it occurs. Word ratio: Calculate the proportion of unique words to the total number of words in the text. Phrase frequency: Count the occurrences of each unique phrases in the text. Unique phrases are distinct combinations of words that appear in the text, not taking into account their repetitions. Phrase ratio: Calculate the proportion of unique phrases to the total number of phrases in the text. Word-finding difficulties: Frequent pauses or use of filler words, like "uh" or "um,". MCI: more frequent instances, longer time to find the right word, or using more general terms. Normal: fewer instances, and better lexical retrieval. Repetition: The participant repeats certain phrases or ideas a few times throughout the conversation. MCI: more frequent repetition. Normal: less frequent repetition (some repetition of words but not excessive) and better memory of what has been mentioned. Lexical complexity: Measure the variety and sophistication of the vocabulary used, considering factors, including word length and frequency of rare words. Limited vocabulary: Repeated use of the same words or simple words. MCI: slightly reduced vocabulary, and more frequent use of simpler words or common terms; Normal: richer vocabulary (low limitation). Secondly, please assess the Semantics features of PARTICIPANT, from Semantic density and Inability to convey meaning, Semantic coherence and Lack of coherence. Semantic density: Measure the amount of meaningful information in the text, and considering factor is the number of content words (nouns, verbs, adjectives, adverbs) relative to function words (prepositions, pronouns, conjunctions, etc.). Inability to convey meaning: Difficulty in expressing ideas clearly, leading to confusion or misunderstanding. Normal: the participant was able to describe the scene clearly. Semantic coherence: Assess the logical consistency and meaningfulness of the text, which can be done by analyzing the relationships between words, phrases, and sentences, and checking for coherence and cohesion. Lack of coherence: Difficulty in maintaining a logical flow of ideas, with thoughts appearing disjointed or unrelated. MCI: struggle with organizing thoughts coherently or maintaining a clear focus in conversation; Normal: greater coherence and organization in their storytelling and conversations. Thirdly, please assess the Syntax and Grammar features of PARTICIPANT, from Syntactic complexity and Simplified sentence structure, Grammatical components and Grammatical errors. Syntactic complexity: Measure the complexity of the sentence structure in the text, considering factors such as the number of clauses per sentence, the average length of sentences, and the use of subordinate clauses. Simplified sentence structure: A lack of complex sentence structures. MCI: simpler sentence structures; Normal: in longer dialogues there is a greater variety of sentence structures (moderately complex), but in short dialogues the sentences may turn out to be simple. Grammatical components: Analyze the text for correct usage of grammar, such as subject-verb agreement, tense consistency, and proper use of pronouns, prepositions, and conjunctions. MCI: avoidance of complex grammatical constructions; Normal: better use of complex grammatical constructions. Grammatical errors: Frequent mistakes in verb tense, subject-verb agreement, or other aspects of grammar. MCI: more frequent grammatical errors, such as mistakes in verb tense and subject-verb agreement; Normal: fewer grammatical errors (a few errors, but not significant). Finally, Please dichotomise PARTICIPANT into the Probably Normal group or the Probably Mild Cognitive Impairment group. PARTICIPANT with MCI might exhibit more difficulty with vocabulary, sentence structure, repetition, word-finding, coherence, and grammar. However, it is not definitive and requires a comprehensive judgement. Now, you do not have to respond to this dialogue. When I have entered THE DIALOGUE BETWEEN PARTICIPANT AND INVESTIGATOR, evaluate PARTICIPANT according to the evaluation rules above. INVESTIGATOR: alright here is this picture . PARTICIPANT: uhhuh . INVESTIGATOR: all of the action you can see going on . PARTICIPANT: all of the action ? INVESTIGATOR: mhm . PARTICIPANT: well the little boy is gonna fall off the stool . PARTICIPANT: he's getting a cookie . PARTICIPANT: do you want me to go slower ? INVESTIGATOR: no you go ahead . PARTICIPANT: okay the little girl is asking for a cookie . PARTICIPANT: she's … the mother is drying dishes . PARTICIPANT: and the sink is overflowing with water . PARTICIPANT: she's gettin(g) water all over the floor . PARTICIPANT: and let me see if anything is going on outside here . PARTICIPANT: …hm well I can't see anything else as far as action goes . PARTICIPANT: is there more ? INVESTIGATOR: okay . INVESTIGATOR: no that's fine thank_you . INVESTIGATOR: okay there it is . INVESTIGATOR: what do you see going on in that picture ? PARTICIPANT: well the boy is getting cookies and he's gonna fall off the stool . PARTICIPANT: and the mother is doing dishes . PARTICIPANT: and the water is running over out_of the sink . PARTICIPANT: she has the faucet on . PARTICIPANT: she's spilling water all over the floor . INVESTIGATOR: anything else ? PARTICIPANT: well the boy is handing the little girl a cookie . INVESTIGATOR: anything else ? PARTICIPANT: …uh nothing that I can see of action . INVESTIGATOR: okay fine . INVESTIGATOR: going on in the picture . INVESTIGATOR: all of the action . PARTICIPANT: okay . PARTICIPANT: …um you mean drying dishes ? INVESTIGATOR: okay . PARTICIPANT: …uh overflowing … the water's overflowing out_of the sink . PARTICIPANT: climbing a stool . PARTICIPANT: getting a cookie . PARTICIPANT: giving the cookie to the girl . PARTICIPANT: raising her arm . PARTICIPANT: …uh stepping in the water . PARTICIPANT: looking out the window . PARTICIPANT: hm that's about all I can see . PARTICIPANT: is there more ? INVESTIGATOR: okay that's fine . INVESTIGATOR: you can start now . PARTICIPANT: there hasta be action . PARTICIPANT: oh the stool … the boy is falling off the stool . PARTICIPANT: …uh the lady is drying dishes . PARTICIPANT: the water is running out_of the sink . PARTICIPANT: …uh …hm … I don't know what else they would want . INVESTIGATOR: just tell me when you think that you've told me everything that you see going on in the picture . PARTICIPANT: that's about all I can see . PARTICIPANT: those three things . INVESTIGATOR: xxx . PARTICIPANT: everything that you see ? INVESTIGATOR: mhm . PARTICIPANT: that I see them ? INVESTIGATOR: see going on in the picture . PARTICIPANT: alright . PARTICIPANT: the mother is drying dishes . PARTICIPANT: the water is running out_of the sink . PARTICIPANT: the boy is going to fall off the stool . PARTICIPANT: he's getting cookies . PARTICIPANT: and the girl is waiting for a cookie from his ( her ) brother . PARTICIPANT: is there more ? INVESTIGATOR: you just tell me when you think you've got it all . PARTICIPANT: …hm I don't … I don't see any other action there . INVESTIGATOR: okay that's fine .

ChatGPT

Let's analyze the dialogue between the participant and the investigator based on the evaluation criteria provided.

Firstly, let's assess the Vocabulary features of the participant.

- Word frequency: Count the occurrences of each unique word in the text. Unique words are distinct words that appear in the text, not taking into account their repetitions. Each word is counted only once, regardless of how many times it occurs.

In the dialogue, the participant uses a limited range of words, with repeated occurrences of certain words such as "action," "cookie," "dishes," and "water." The participant does not demonstrate a diverse vocabulary.

- Word ratio: Calculate the proportion of unique words to the total number of words in the text.

The participant's word ratio is low since they repeat certain words and use a limited vocabulary. The participant tends to use the same words repeatedly throughout the conversation.

- Phrase frequency: Count the occurrences of each unique phrase in the text. Unique phrases are distinct combinations of words that appear in the text, not taking into account their repetitions.

The participant repeats certain phrases like "the boy is getting cookies" and "the mother is drying dishes." The occurrence of these phrases indicates some repetition in their speech.

- Phrase ratio: Calculate the proportion of unique phrases to the total number of phrases in the text.

Since the participant repeats certain phrases, the phrase ratio is low. They rely on a limited set of phrases to describe the scene.

- Word-finding difficulties: Frequent pauses or use of filler words, like "uh" or "um."

The participant exhibits some word-finding difficulties, as indicated by their use of filler words like "uh" and "um." They pause at times, suggesting a struggle to find the right words or recall specific terms.

- Repetition: The participant repeats certain phrases or ideas a few times throughout the conversation.

The participant repeats phrases such as "the boy is getting cookies" and "the mother is drying dishes" multiple times. This repetition is more frequent than what would be expected in a conversation with normal cognition.

- Lexical complexity: Measure the variety and sophistication of the vocabulary used, considering factors such as word length and frequency of rare words.

The participant's vocabulary is relatively simple and lacks complexity. They use common words and do not demonstrate the use of rare or sophisticated vocabulary.

- Limited vocabulary: Repeated use of the same words or simple words.

The participant's speech shows a limited vocabulary, as they repeat certain words and rely on simpler terms. They use the same words repeatedly, which indicates a reduced range of vocabulary.

Based on the assessment of Vocabulary features, the participant demonstrates limited vocabulary, word-finding difficulties, repetition, and reduced lexical complexity. These characteristics align with the features associated with mild cognitive impairment (MCI).

Next, let's assess the Semantics features of the participant.

- Semantic density: Measure the amount of meaningful information in the text, considering factors such as the number of content words (nouns, verbs, adjectives, adverbs) relative to function words (prepositions, pronouns, conjunctions, etc.).

The participant's dialogue contains a moderate amount of meaningful information, with a focus on describing the actions and events in the picture. They use content words like "boy," "fall," "cookie," "mother," "dishes," and "water" to convey meaning.

- Inability to convey meaning: Difficulty in expressing ideas clearly, leading to confusion or misunderstanding.

The participant does not demonstrate a significant inability to convey meaning. They generally succeed in describing the actions and events in the picture, although with some repetition and word-finding difficulties.

- Semantic coherence: Assess the logical consistency and meaningfulness of the text, analyzing the relationships between words, phrases, and sentences.

The participant's dialogue maintains some level of semantic coherence. They describe the actions and events in a somewhat logical manner, linking different elements together.

- Lack of coherence: Difficulty in maintaining a logical flow of ideas, with thoughts appearing disjointed or unrelated.

The participant's speech does not exhibit a lack of coherence. Although there are occasional pauses and repetitions, the participant is able to maintain a relatively coherent flow of ideas and provide a reasonable description of the picture.

Based on the assessment of Semantics features, the participant demonstrates moderate semantic density, some ability to convey meaning, coherence in describing the picture, and no significant lack of coherence.

Moving on, let's assess the Syntax and Grammar features of the participant.

- Syntactic complexity: Measure the complexity of the sentence structure in the text, considering factors such as the number of clauses per sentence, the average length of sentences, and the use of subordinate clauses.

The participant's sentence structures are relatively simple. They tend to use short, straightforward sentences without much complexity or the inclusion of subordinate clauses.

- Simplified sentence structure: A lack of complex sentence structures.

The participant's sentences predominantly follow a simple structure. They do not employ complex sentence structures or incorporate a variety of sentence types.

- Grammatical components: Analyze the text for correct usage of grammar, such as subject-verb agreement, tense consistency, and proper use of pronouns, prepositions, and conjunctions.

The participant demonstrates generally correct usage of grammatical components. There are no significant errors in subject-verb agreement, tense consistency, or the use of pronouns, prepositions, and conjunctions.

- Grammatical errors: Frequent mistakes in verb tense, subject-verb agreement, or other aspects of grammar.

The participant's dialogue does not contain frequent grammatical errors. While there are some pauses and occasional word omissions, there are no significant mistakes in verb tense, subject-verb agreement, or other grammatical aspects.

Based on the assessment of Syntax and Grammar features, the participant exhibits simplified sentence structures, moderately complex sentence structures in longer dialogues, correct usage of grammatical components, and no significant grammatical errors.

In conclusion, based on the evaluation of the participant's dialogue according to the provided criteria, there are indications of mild cognitive impairment (MCI). The participant demonstrates features such as limited vocabulary, word-finding difficulties, repetition, reduced lexical complexity, moderately coherent semantic expression, simplified sentence structures, and generally correct grammar usage. However, a comprehensive judgment is necessary to definitively determine if the participant falls into the "Probably Mild Cognitive Impairment" group.
