## Supplementary material for "Text dialogue analysis Based ChatGPT for Primary Screening of Mild Cognitive Impairment": Data(GPT4_vs_GPT3.5_model1-5): 109_GPT3.5 (3).docx

ChatGPT

Based on the provided conversation, we can analyze the indicators and evaluate the severity for each aspect as follows:

Vocabulary features:

- Limited vocabulary: Severity: 3/10. The participant demonstrates a slightly reduced vocabulary with more frequent use of simple words or common terms.
- Increased verbosity: Severity: 2/10. The participant uses a few more words than necessary but not excessively.
- Overlearned phrases: Severity: 1/10. There is no evidence of overuse of cliches or stock phrases.
- Improper use of pronouns: Severity: 1/10. There is no difficulty observed in using pronouns correctly.
- Hesitation (Word-finding difficulties): Severity: 2/10. The participant exhibits a few instances of hesitation and filler words.
- Circumlocution (Word-finding difficulties): Severity: 1/10. There is no clear evidence of difficulty finding specific words or deliberate vagueness.
- Tip-of-the-tongue phenomenon: Severity: 1/10. There is no indication of partial recall or difficulty remembering specific words.
- Repetition: Severity: 1/10. There is no repetition of stories, questions, or statements within a short span of time.

Syntax and Grammar features:

- Simplified sentence structure and grammar: Severity: 2/10. The participant demonstrates a preference for simpler sentence structures in both long and short sentences but without significant avoidance of complex grammatical constructions.
- Grammatical errors: Severity: 1/10. There are no significant grammatical errors observed.
- Difficulty with complex ideas: Severity: 1/10. The participant handles the given narrative adequately without difficulty.

Semantics features:

- Semantic paraphasias: Severity: 1/10. There is no difficulty observed in expressing ideas clearly or making errors in word choice.
- Lack of coherence: Severity: 1/10. The participant maintains coherence and focus in conversation without frequent changes in topic.
- Consistency in errors: Severity: 1/10. There are no consistent language errors observed.
- Lack of semantic fluency: Severity: 1/10. The participant demonstrates no notable difficulty in naming items or describing processes.
- Disorganized narrative structure: Severity: 1/10. The participant maintains a coherent line of thought without jumping around or lacking key elements.
- Orientation issues: Severity: 1/10. There is no confusion or difficulty observed regarding date, time, place, or sequence of events.
- Decline in conversational engagement and responsiveness: Severity: 1/10. The participant actively engages in conversation and responds appropriately.
- Memory issues: Severity: 1/10. There is no significant difficulty observed in memory recall.
- Lack of abstract thinking and synthesis: Severity: 1/10. The participant demonstrates the ability to synthesize information and understand abstract concepts.
- Abnormal emotions: Severity: 1/10. There are no apparent changes in tone or style of speech, lack of initiative, or abnormal emotions.

Based on the synthesis of the indicators, the participant's performance does not indicate significant signs of mild cognitive impairment (MCI). The participant's language use falls within the range of normal cognition, with minor deviations observed in limited vocabulary and increased verbosity. However, it is important to note that this analysis is based on a single conversation and a more comprehensive assessment is necessary to make a conclusive determination about cognitive impairment.
