## Supplementary material for "Text dialogue analysis Based ChatGPT for Primary Screening of Mild Cognitive Impairment": Data(GPT4_vs_GPT3.5_model1-5): 109_GPT3.5 (4).docx

ChatGPT

Let's assess the participant's language features based on the provided conversation and evaluate the severity of each indicator.

Vocabulary features: a. Analyzing word frequency, word ratio, phrase frequency, and phrase ratio is not feasible based on the provided conversation. b. Calculating lexical complexity is not possible based on the provided conversation. c. Assessing and rating the severity of the following aspects:

- Limited vocabulary: 3 - The participant demonstrates a basic vocabulary but does not exhibit an extensive range of words.
- Increased verbosity: 2 - The participant tends to provide additional information but not excessively so.
- Overlearned phrases: 1 - No evidence of overlearned phrases.
- Improper use of pronouns: 1 - No evidence of improper pronoun usage.
- Hesitation: 2 - The participant exhibits some hesitation but not significantly.
- Circumlocution: 1 - No evidence of excessive circumlocution.
- Tip-of-the-tongue phenomenon: 1 - No evidence of experiencing the tip-of-the-tongue phenomenon.
- Repetition: 1 - No evidence of excessive repetition.

Syntax and grammar features: a. Analyzing syntactic complexity and grammatical components is not feasible based on the provided conversation. b. Assessing and rating the severity of the following aspects:

- Simplified sentence structure and grammar: 2 - The participant generally constructs coherent sentences with some minor errors.
- Grammatical errors: 2 - The participant makes occasional grammatical errors but not consistently.
- Difficulty with complex ideas: 1 - No evidence of difficulty with complex ideas.

Semantics features: a. Analyzing semantic density and coherence is not feasible based on the provided conversation. b. Assessing and rating the severity of the following aspects:

- Semantic paraphasias: 1 - No evidence of semantic paraphasias.
- Lack of coherence: 2 - The participant's responses are generally coherent, but there are occasional gaps in information.
- Consistency in errors: 1 - No evidence of consistent errors.
- Lack of semantic fluency: 1 - The participant exhibits adequate semantic fluency.
- Disorganized narrative structure: 1 - The participant's narrative structure is relatively organized.
- Orientation issues: 1 - No evidence of orientation issues.
- Decline in conversational engagement and responsiveness: 1 - The participant is engaged and responsive throughout the conversation.
- Memory issues: 1 - No evidence of significant memory issues.
- Lack of abstract thinking and synthesis: 1 - The participant demonstrates adequate abstract thinking and synthesis.
- Abnormal emotions: 1 - No evidence of abnormal emotions.

Based on the comprehensive synthesis of all the indicators, it appears that the participant falls into the Probably Normal Cognition Group. The language features observed do not indicate a significant impairment or suggestive of mild cognitive impairment (MCI). However, it's important to note that this assessment is based solely on the provided conversation and further evaluation may be necessary to confirm the cognitive status of the participant.
