## Supplementary material for "Text dialogue analysis Based ChatGPT for Primary Screening of Mild Cognitive Impairment": Data(GPT4_vs_GPT3.5_model1-5): 109_GPT3.5 (5).docx

ChatGPT

Let's analyze the participant's language features based on the provided conversation. We'll assess each indicator and rate its severity on a scale of 1 to 10, with 1 being the least severe and 10 being the most severe.

Vocabulary Features: a. Word Frequency and Word Ratio: We need a larger sample of text to analyze word frequency and word ratio accurately.

b. Tip-of-the-tongue phenomenon: Severity: 3 The participant occasionally experiences the tip-of-the-tongue phenomenon, such as when they struggle to recall a word but remember partial information about it. However, it's not a consistent or prominent issue in the conversation.

Syntax and Grammar Features: a. Syntactic Complexity and Grammatical Components: We need a larger sample of text to analyze the syntactic complexity and grammatical components accurately.

b. Difficulty with complex ideas: Severity: 2 The participant exhibits minimal difficulty with complex ideas. While they handle simpler sentences and ideas well, they may face slight difficulty when the narrative or question becomes more complex.

Semantics Features: a. Semantic Density and Coherence: We need a larger sample of text to analyze semantic density and coherence accurately.

b. Memory issues: Severity: 3 The participant demonstrates mild memory issues. They repeat questions, struggle to recall certain information or sequences, and have some difficulty remembering specific details of the story. However, it's important to consider that memory issues can also occur in normal aging.

Comprehensive Synthesis: Based on the available indicators, it's challenging to make a definitive classification. However, considering the overall severity ratings, it's possible to place the participant in the Probably Normal Cognition Group. The participant shows mild difficulty with complex ideas and some memory issues, but these indicators do not strongly suggest Mild Cognitive Impairment (MCI). It's essential to conduct a more comprehensive evaluation to make a definitive determination about cognitive impairment.
