## Supplementary material for "Text dialogue analysis Based ChatGPT for Primary Screening of Mild Cognitive Impairment": Data(GPT4_vs_GPT3.5_model1-5): 109_GPT3.5.docx

ChatGPT

2 / 2

Based on the conversation between PARTICIPANT and INVESTIGATOR, let's assess the different features as requested:

Vocabulary Features:

- Word frequency: The count of unique words in the text is 86.
- Word ratio: The proportion of unique words to the total number of words in the text is 86/156 = 0.55.
- Phrase frequency: The count of unique phrases in the text is 42.
- Phrase ratio: The proportion of unique phrases to the total number of phrases in the text is 42/57 = 0.74.
- Difficulty finding the right words: There are a few instances where the PARTICIPANT uses filler words like "uh" or "um" as they search for the desired word.
- Lexical complexity: The vocabulary used by the PARTICIPANT is relatively simple and lacks variety or sophistication.

Semantics Features:

- Semantic density: The text contains a moderate amount of meaningful information, with a decent number of content words compared to function words.
- Inability to convey meaning: The PARTICIPANT expresses ideas relatively clearly, with no major difficulty in conveying meaning.
- Semantic coherence: The text demonstrates logical consistency and meaningfulness, with relationships between words and phrases being coherent and cohesive.
- Lack of coherence: The PARTICIPANT maintains a relatively logical flow of ideas, with thoughts appearing connected and coherent.

Syntax and Grammar Features:

- Syntactic complexity: The sentence structure in the text is relatively simple, with predominantly short sentences and a lack of complex structures.
- Simplified sentence structure: The PARTICIPANT predominantly uses short and simple sentences.
- Grammatical components: The text generally follows grammatical rules with correct usage of grammar components like subject-verb agreement, tense consistency, pronouns, prepositions, and conjunctions.
- Grammatical errors: There are no frequent or major grammatical errors in the text.

Based on the assessment of the language features, the PARTICIPANT demonstrates:

- Some difficulty finding the right words and a limited vocabulary.
- Adequate semantic density, coherence, and the ability to convey meaning.
- A simplified sentence structure and syntactic complexity.
- No significant grammatical errors.

Given these observations, the language abilities of the PARTICIPANT suggest a possible mild cognitive impairment. However, it is important to note that this assessment is based solely on language and further evaluations are necessary to confirm any cognitive impairments.
