## Supplementary material for "Text dialogue analysis Based ChatGPT for Primary Screening of Mild Cognitive Impairment": Data(GPT4_vs_GPT3.5_model1-5): 109_GPT4 (2).docx

ChatGPT

Based on the provided conversation, let's evaluate the PARTICIPANT's speech on the listed parameters:

**Vocabulary features:**

1. Word frequency: The participant uses a moderate variety of words.
2. Word ratio: The ratio of unique words to the total number of words is moderate.
3. Phrase frequency: The participant uses several phrases, indicating a good ability to express different ideas.
4. Phrase ratio: The ratio of unique phrases to the total number of phrases is moderate.
5. Word-finding difficulties: The participant shows minor instances of word-finding difficulties, using fillers like "uh" and "um".
6. Repetition: There's a moderate amount of repetition in the participant's speech.
7. Lexical complexity: The participant uses a mix of simple and complex words.
8. Limited vocabulary: The participant doesn't show a significant limitation in vocabulary.

**Semantics features:**

1. Semantic density: The participant seems to have a good amount of meaningful information in their speech.
2. Inability to convey meaning: The participant is generally able to convey their ideas clearly.
3. Semantic coherence: The participant's speech is generally coherent, with logical relationships between words, phrases, and sentences.
4. Lack of coherence: The participant's speech is mostly coherent, with thoughts appearing related.

**Syntax and Grammar features:**

1. Syntactic complexity: The participant's sentences are relatively simple but not overly simplified.
2. Simplified sentence structure: The participant uses a mix of simple and complex sentences.
3. Grammatical components: The participant generally uses grammar correctly.
4. Grammatical errors: There are few noticeable grammatical errors in the participant's speech.

Based on this analysis, there are some instances of word-finding difficulties and repetition. However, the participant is generally able to convey their ideas clearly and coherently, and they use a mix of simple and complex sentences and words. Therefore, the participant would likely fall into the Probably Normal cognitive functioning group. However, this is just an initial assessment and should not replace a professional medical evaluation.
