## Supplementary material for "Text dialogue analysis Based ChatGPT for Primary Screening of Mild Cognitive Impairment": Data(GPT4_vs_GPT3.5_model1-5): 109_GPT4 (3).docx

ChatGPT

Based on the presented analysis, here are the scoring of the evaluation indicators:

**Vocabulary features:**

1. Limited vocabulary: 3 (Some repetitive use of phrases but not overly limited vocabulary)
2. Increased verbosity: 2 (Not significant verbosity, mostly succinct in responses)
3. Overlearned phrases: 2 (Not many cliches or overused phrases were used)
4. Improper use of pronouns: 2 (Slight misuse of pronouns ("his (her) brother") but relatively rare)
5. Hesitation (Word-finding difficulties): 4 (Some hesitation evident but not severe)
6. Circumlocution (Word-finding difficulties): 3 (Occasional circumlocution, but not pervasive)
7. Tip-of-the-tongue phenomenon: 2 (Rarely seen)
8. Repetition: 4 (Repeated the scene description several times, could be due to the nature of the task)

**Syntax and Grammar features:**

1. Simplified sentence structure and grammar: 3 (Uses simple structures more frequently)
2. Grammatical errors: 2 (A few minor errors but not significant)
3. Difficulty with complex ideas: 2 (Did not seem to have difficulty describing the scene)

**Semantics features:**

1. Semantic paraphasias: 2 (Overall clarity in expressing ideas was maintained)
2. Lack of coherence: 3 (Some occasional lack of coherence)
3. Consistency in errors: 2 (Some repetition, but not unusual for educational or cultural background)
4. Lack of semantic fluency: 3 (Slight struggle in narrating the sequence of events)
5. Disorganized narrative structure: 3 (Narrative structure is slightly disorganized due to repetitions)
6. Orientation Issues: 1 (No evident confusion about the sequence of events)
7. Decline in conversational engagement and responsiveness: 2 (Maintains decent engagement, responds to prompts appropriately)
8. Memory issues: 2 (Some repetition might indicate minor memory issues, but it could also be due to the nature of the task)
9. Lack of abstract thinking and synthesis: 2 (Was able to synthesize the overall situation of the scene)
10. Abnormal emotions: 1 (No abnormal emotions observed in the conversation)

With a holistic view of the participant's language and communication skills, we have more indicators pointing towards normal cognition rather than mild cognitive impairment. Therefore, I would place the participant in the **Probably Normal Cognition Group**. It should be stressed, however, that a comprehensive clinical evaluation would be necessary for an accurate diagnosis
