## Supplementary material for "Text dialogue analysis Based ChatGPT for Primary Screening of Mild Cognitive Impairment": Data(GPT4_vs_GPT3.5_model1-5): 109_GPT4 (5).docx

ChatGPT

Based on the information provided in the transcripts, here's an analysis:

Vocabulary features: a. Word frequency and word ratio seem to be within a normal range, as the participant uses a moderate range of different words, pronouns, and specific names (e.g., "stool," "cookie," "sink"). Lexical complexity appears to be relatively low, with many basic words and phrases repeated, with few rare or complex words. Vocabulary Rating: 3/10

b. There's some evidence of the tip-of-the-tongue phenomenon, with some hesitations and partial word formations, particularly when the participant is trying to explain more complex actions or objects. Tip-of-the-tongue Rating: 4/10

Syntax and Grammar Features: a. Syntactic complexity and grammatical components appear relatively standard. Sentences are largely grammatically correct, with a variety of structures used, although few complex sentences. Syntax and Grammar Rating: 2/10

b. The participant shows some difficulty with complex ideas, often reverting to simpler sentences and descriptions when faced with a more complex narrative. Difficulty with Complex Ideas Rating: 4/10

Semantics Features: a. The semantic density of the participant's responses is fairly high, with a good ratio of content words to function words, and reasonable coherence in their narrative. Semantic Rating: 2/10

b. Some memory issues are evident, with repeated descriptions and questions. However, it's not clear if this is due to the participant forgetting their previous statements or simply struggling to come up with new observations. Memory Issues Rating: 4/10

In conclusion, this participant shows some potential signs of Mild Cognitive Impairment, including the tip-of-the-tongue phenomenon, difficulty with complex ideas, and some memory issues. However, these signs are relatively mild and might not be beyond what could be expected in normal aging. Therefore, based on this analysis, the participant would be classified into the Probably Normal Cognition Group. However, this does not exclude the possibility of Mild Cognitive Impairment and further testing could be beneficial. Please remember, this evaluation isn't meant to provide a formal diagnosis but to serve as an initial screening tool for potential MCI
