## Supplementary material for "Text dialogue analysis Based ChatGPT for Primary Screening of Mild Cognitive Impairment": Data(GPT4_vs_GPT3.5_model1-5): 109_GPT4.docx

ChatGPT

Assessment:

Vocabulary features:

1. Word frequency: 103 unique words
2. Word ratio: 0.34 (103 unique words / 299 total words)
3. Phrase frequency: 59 unique phrases
4. Phrase ratio: 0.57 (59 unique phrases / 103 total phrases)
5. Difficulty finding the right words: Slight (occasional pauses and filler words)
6. Lexical complexity: Moderate (some sophisticated vocabulary, but also simple words)
7. Limited vocabulary: Slight (some repetition and simple words, but within normal range)

Semantics features:

1. Semantic density: Moderate (a balance of content and function words)
2. Inability to convey meaning: Slight (minor difficulty, but overall able to convey meaning)
3. Semantic coherence: Good (generally coherent with occasional disjointed thoughts)
4. Lack of coherence: Slight (some difficulty maintaining logical flow, but mostly coherent)

Syntax and Grammar features:

1. Syntactic complexity: Low to Moderate (simple sentence structures, few subordinate clauses)
2. Simplified sentence structure: Predominant (short and simple sentences, but within normal range)
3. Grammatical components: Moderate (some mistakes, but mostly correct usage)
4. Grammatical errors: Slight (occasional errors in tense, subject-verb agreement, etc.)

Based on this assessment, I would assign PARTICIPANT to the Probably Normal group. While there are some minor language gaps and occasional difficulty finding the right words, these issues fall within the normal range, considering that it can be difficult to distinguish between mild cognitive impairment and normalcy by language alone.
