## Supplementary material for "Text dialogue analysis Based ChatGPT for Primary Screening of Mild Cognitive Impairment": Data(GPT4_vs_GPT3.5_model1-5): 110_GPT3.5 (2).docx

Attention! You do not need to diagnose, just follow the steps below to initially screen for PARTICIPANT with possibly mild cognitive impairment (MCI). Some differences between MCI and normal cognition are also given, please analyse them in detail, step by step. Firstly, please assess the Vocabulary features of PARTICIPANT at the Word frequency and Word ratio, Phrase frequency and Phrase ratio, Word-finding difficulties, Repetition, Lexical complexity, Limited vocabulary. Word frequency: Count the occurrences of each unique word in the text. Unique words are distinct words that appear in the text, not taking into account their repetitions. Each word is counted only once, regardless of how many times it occurs. Word ratio: Calculate the proportion of unique words to the total number of words in the text. Phrase frequency: Count the occurrences of each unique phrases in the text. Unique phrases are distinct combinations of words that appear in the text, not taking into account their repetitions. Phrase ratio: Calculate the proportion of unique phrases to the total number of phrases in the text. Word-finding difficulties: Frequent pauses or use of filler words, like "uh" or "um,". MCI: more frequent instances, longer time to find the right word, or using more general terms. Normal: fewer instances, and better lexical retrieval. Repetition: The participant repeats certain phrases or ideas a few times throughout the conversation. MCI: more frequent repetition. Normal: less frequent repetition (some repetition of words but not excessive) and better memory of what has been mentioned. Lexical complexity: Measure the variety and sophistication of the vocabulary used, considering factors, including word length and frequency of rare words. Limited vocabulary: Repeated use of the same words or simple words. MCI: slightly reduced vocabulary, and more frequent use of simpler words or common terms; Normal: richer vocabulary (low limitation). Secondly, please assess the Semantics features of PARTICIPANT, from Semantic density and Inability to convey meaning, Semantic coherence and Lack of coherence. Semantic density: Measure the amount of meaningful information in the text, and considering factor is the number of content words (nouns, verbs, adjectives, adverbs) relative to function words (prepositions, pronouns, conjunctions, etc.). Inability to convey meaning: Difficulty in expressing ideas clearly, leading to confusion or misunderstanding. Normal: the participant was able to describe the scene clearly. Semantic coherence: Assess the logical consistency and meaningfulness of the text, which can be done by analyzing the relationships between words, phrases, and sentences, and checking for coherence and cohesion. Lack of coherence: Difficulty in maintaining a logical flow of ideas, with thoughts appearing disjointed or unrelated. MCI: struggle with organizing thoughts coherently or maintaining a clear focus in conversation; Normal: greater coherence and organization in their storytelling and conversations. Thirdly, please assess the Syntax and Grammar features of PARTICIPANT, from Syntactic complexity and Simplified sentence structure, Grammatical components and Grammatical errors. Syntactic complexity: Measure the complexity of the sentence structure in the text, considering factors such as the number of clauses per sentence, the average length of sentences, and the use of subordinate clauses. Simplified sentence structure: A lack of complex sentence structures. MCI: simpler sentence structures; Normal: in longer dialogues there is a greater variety of sentence structures (moderately complex), but in short dialogues the sentences may turn out to be simple. Grammatical components: Analyze the text for correct usage of grammar, such as subject-verb agreement, tense consistency, and proper use of pronouns, prepositions, and conjunctions. MCI: avoidance of complex grammatical constructions; Normal: better use of complex grammatical constructions. Grammatical errors: Frequent mistakes in verb tense, subject-verb agreement, or other aspects of grammar. MCI: more frequent grammatical errors, such as mistakes in verb tense and subject-verb agreement; Normal: fewer grammatical errors (a few errors, but not significant). Finally, Please dichotomise PARTICIPANT into the Probably Normal group or the Probably Mild Cognitive Impairment group. PARTICIPANT with MCI might exhibit more difficulty with vocabulary, sentence structure, repetition, word-finding, coherence, and grammar. However, it is not definitive and requires a comprehensive judgement. Now, you do not have to respond to this dialogue. When I have entered THE DIALOGUE BETWEEN PARTICIPANT AND INVESTIGATOR, evaluate PARTICIPANT according to the evaluation rules above. INVESTIGATOR: tell me all the action you see going on in the picture . PARTICIPANT: mother's dryin(g) the dishes . PARTICIPANT: Junior's in the cookie jar handin(g) a cookie to his little sister . PARTICIPANT: the water's spilling out o(f) the sink . PARTICIPANT: the window is open . PARTICIPANT: the door to the cupboard where the cookie jar is open and the … the top of the cookie jar is off . PARTICIPANT: cookie jar is quite full . PARTICIPANT: mother has two cups and a dish on the sink . PARTICIPANT: looks like a nice day . PARTICIPANT: …uh action we're lookin(g) for action . PARTICIPANT: the kitchen stool that Junior's on is about ready to fall over . PARTICIPANT: what else do we have ? PARTICIPANT: …uh that's all the action I see . PARTICIPANT: oh …uh Sissy is holdin(g) her finger up to her mouth like . PARTICIPANT: don't make any noise or mother (wi)ll turn around . PARTICIPANT: I … I've already said he's reachin(g) a cookie to her and goin(g) for another one just before the stool falls down . PARTICIPANT: his socks are droopin(g) . PARTICIPANT: does that count ? INVESTIGATOR: … PARTICIPANT: …uh I don't see anything outside . PARTICIPANT: …uh …uh the curtains don't look like they're blowin(g) or anything . PARTICIPANT: mother's just standin(g) lookin(g) . PARTICIPANT: she's gonna get her feet wet in a minute . INVESTIGATOR: okay that's fine . PARTICIPANT: okay . INVESTIGATOR: tell me everything that you see going on in this picture . INVESTIGATOR: tell me everything that's happening there . PARTICIPANT: young boy is up at the cookie jar . PARTICIPANT: he has the cookie jar opened . PARTICIPANT: he's reachin(g) for one for himself . PARTICIPANT: he's handing one down to his sister . PARTICIPANT: the three legged stool is coming down with him . PARTICIPANT: …uh they're nicely dressed kids but …uh the boy needs his socks pulled up . PARTICIPANT: …mm that's all I see there . PARTICIPANT: mother … I presume it's mother is at the sink . PARTICIPANT: the water is …r running over out_of sink . PARTICIPANT: she's drying a dish . PARTICIPANT: she's looking out the window . PARTICIPANT: there are two cups and a dish, I presume that's a dish, that are already dry I guess and sitting on the side of the sink . PARTICIPANT: …uh nicely groomed . PARTICIPANT: nice curtains pulled back . PARTICIPANT: it's apparently summer outside because the window's open and it looks like shrubs are growing . PARTICIPANT: you can see a window in the other part of the house . PARTICIPANT: I presume that's still their house . PARTICIPANT: and there's a driveway bordered with grass . PARTICIPANT: and there's a tree out there somewhere that you can see part of the trunk and part of the foliage . PARTICIPANT: and …uh I see cabinets around the kitchen, other cabinets . PARTICIPANT: and that's about it . INVESTIGATOR: okay .

ChatGPT

Based on the provided dialogue between the investigator and participant, let's evaluate the participant according to the evaluation rules provided.

Assessment of Vocabulary Features:

1. Word frequency and Word ratio: To assess the word frequency, we count the occurrences of each unique word in the text. However, since the given dialogue is in spoken form and not a complete text, we cannot accurately count the word frequency. The word ratio, which calculates the proportion of unique words to the total number of words, cannot be determined without an accurate word count.
2. Phrase frequency and Phrase ratio: Similar to word frequency, we cannot accurately count the phrase frequency and calculate the phrase ratio without a complete text or accurate phrase count.
3. Word-finding difficulties: The participant shows some word-finding difficulties. They use filler words like "uh" and "um" and have instances where they pause or take longer to find the right word. For example, in the first statement, the participant says, "mother's dryin(g) the dishes," with a hesitation in finding the word "drying." This suggests mild word-finding difficulties.
4. Repetition: The participant repeats certain phrases, like "Junior's in the cookie jar handing a cookie to his little sister," and "the water's spilling out of the sink." This repetition is within normal limits, as some repetition is common in everyday conversation.
5. Lexical complexity: Assessing lexical complexity based on the given dialogue is challenging since the analysis requires a complete text and consideration of factors such as word length and frequency of rare words. Without a broader context, it is difficult to make a conclusive judgment about the participant's lexical complexity.
6. Limited vocabulary: The participant does not exhibit a limited vocabulary based on the given dialogue. They use words and terms appropriate to describe the scene and convey their observations. There is no evidence of a significantly reduced vocabulary or frequent use of simpler words.

Assessment of Semantics Features:

1. Semantic density: Assessing semantic density requires considering the number of content words relative to function words in the text. However, in the provided dialogue, we don't have access to the complete text, and thus, an accurate assessment of semantic density is not possible.
2. Inability to convey meaning: The participant demonstrates the ability to express ideas clearly and describes the scene with reasonable clarity. There is no significant difficulty in conveying meaning or causing confusion or misunderstanding.
3. Semantic coherence: The participant's statements show a reasonable level of semantic coherence. They provide descriptions of various elements in the picture and maintain meaningful relationships between words and phrases. The descriptions follow a logical flow and maintain cohesion within the context of the conversation.
4. Lack of coherence: There is no evidence of a lack of coherence in the participant's statements. The thoughts expressed are not disjointed or unrelated. The participant maintains a clear focus on describing the scene and organizes their thoughts coherently.

Assessment of Syntax and Grammar Features:

1. Syntactic complexity and Simplified sentence structure: The participant's sentences generally exhibit a simple sentence structure. They use short, straightforward sentences to describe the scene. In longer dialogues, there might be a greater variety of sentence structures, but in the provided dialogue, the sentences are relatively simple.
2. Grammatical components: The participant demonstrates the correct usage of grammar. There is subject-verb agreement, tense consistency, and appropriate use of pronouns, prepositions, and conjunctions. The participant doesn't avoid complex grammatical constructions, and their statements maintain grammatical accuracy.
3. Grammatical errors: There are no frequent grammatical errors in the participant's statements. Although the participant says, "his socks are drooping," it is not a significant grammatical error. Overall, the participant's grammar is mostly correct, with only minor deviations.

Based on the evaluation, the participant shows some mild word-finding difficulties but generally exhibits a reasonable vocabulary and semantic coherence. They use a simplified sentence structure but maintain grammatical accuracy with minimal errors. Considering these factors, it is more likely that the participant falls into the Probably Normal group rather than the Probably Mild Cognitive Impairment group. However, it's important to note that a comprehensive judgment would require a more thorough evaluation and consideration of additional factors.
