## Supplementary material for "Text dialogue analysis Based ChatGPT for Primary Screening of Mild Cognitive Impairment": Data(GPT4_vs_GPT3.5_model1-5): 110_GPT3.5 (3).docx

ChatGPT

2 / 2

Based on the provided dialogue, let's analyze the indicators and evaluate the severity for each indicator mentioned:

Vocabulary Features:

1. Limited vocabulary: Severity: 3 - The participant demonstrates a moderately reduced vocabulary and occasionally repeats words.
2. Increased verbosity: Severity: 2 - The participant uses more words than necessary but not excessively.
3. Overlearned phrases: Severity: 1 - The participant does not exhibit overuse of cliches or stock phrases.
4. Improper use of pronouns: Severity: 1 - The participant uses pronouns appropriately without difficulty.
5. Hesitation (Word-finding difficulties): Severity: 2 - The participant occasionally hesitates and uses filler words.
6. Circumlocution (Word-finding difficulties): Severity: 2 - The participant sometimes uses more words than necessary but not to the extent of deliberate vagueness or evasiveness.
7. Tip-of-the-tongue phenomenon: Severity: 1 - The participant does not demonstrate difficulty recalling words or partial recall.
8. Repetition: Severity: 1 - The participant does not excessively repeat stories, questions, or statements.

Syntax and Grammar Features:

1. Simplified sentence structure and grammar: Severity: 1 - The participant exhibits a balanced usage of simple and complex sentences.
2. Grammatical errors: Severity: 1 - The participant makes few grammatical errors.
3. Difficulty with complex ideas: Severity: 1 - The participant can handle complex ideas without difficulty.

Semantics Features:

1. Semantic paraphasias: Severity: 1 - The participant expresses ideas clearly without errors in word choice.
2. Lack of coherence: Severity: 2 - The participant occasionally struggles with organizing thoughts coherently and changing topics abruptly.
3. Consistency in errors: Severity: 1 - The participant does not exhibit consistent language errors.
4. Lack of semantic fluency: Severity: 1 - The participant can name items and describe processes without difficulty.
5. Disorganized narrative structure: Severity: 1 - The participant maintains a coherent line of thought without jumping chronologically or lacking key elements.
6. Orientation issues: Severity: 1 - The participant shows no signs of confusion about date, time, place, or sequence of events.
7. Decline in conversational engagement and responsiveness: Severity: 1 - The participant remains actively engaged and responds appropriately in the conversation.
8. Memory issues: Severity: 1 - The participant does not demonstrate difficulty in recalling information or major details.
9. Lack of abstract thinking and synthesis: Severity: 1 - The participant can understand and express abstract concepts.
10. Abnormal emotions: Severity: 1 - The participant does not exhibit abnormal emotions or changes in speech style.

Based on the analysis of the indicators, the severity ratings, and the synthesis of all the indicators, the participant is likely in the Probably Normal Cognition Group. The participant shows some mild language-related issues, such as limited vocabulary and occasional difficulties with coherence and verbosity, but overall does not exhibit significant signs of mild cognitive impairment (MCI). However, a comprehensive evaluation should be conducted by a healthcare professional to provide a definitive diagnosis.
