## Supplementary material for "Text dialogue analysis Based ChatGPT for Primary Screening of Mild Cognitive Impairment": Data(GPT4_vs_GPT3.5_model1-5): 110_GPT3.5 (4).docx

ChatGPT

2 / 2

Let's assess the participant's language features for possible mild cognitive impairment (MCI) based on the indicators provided. We'll rate the severity of each evaluation indicator on a scale of 1 to 10, with 1 being the least severe and 10 being the most severe.

Vocabulary Features: a. Analyzing word frequency, word ratio, phrase frequency, and phrase ratio: Not provided in the conversation.

b. Calculating lexical complexity: Not applicable.

c. Assessing and rating the severity of various aspects:

- Limited vocabulary: 3
- Increased verbosity: 2
- Overlearned phrases: 2
- Improper use of pronouns: 1
- Hesitation: 1
- Circumlocution: 2
- Tip-of-the-tongue phenomenon: 1
- Repetition: 1

Syntax and Grammar Features: a. Analyzing syntactic complexity and grammatical components: Not applicable.

b. Assessing and rating the severity of various aspects:

- Simplified sentence structure and grammar: 1
- Grammatical errors: 1
- Difficulty with complex ideas: 2

Semantics Features: a. Analyzing semantic density and coherence: Not applicable.

b. Assessing and rating the severity of various aspects:

- Semantic paraphasias: 1
- Lack of coherence: 1
- Consistency in errors: 1
- Lack of semantic fluency: 2
- Disorganized narrative structure: 2
- Orientation issues: 1
- Decline in conversational engagement and responsiveness: 1
- Memory issues: 1
- Lack of abstract thinking and synthesis: 2
- Abnormal emotions: 1

Based on the comprehensive synthesis of the indicators, we can categorize the participant into the Probably Mild Cognitive Impairment Group. While some indicators suggest normal cognition or only mild impairment, others point to potential cognitive difficulties, such as limited vocabulary, increased verbosity, overlearned phrases, difficulty with complex ideas, lack of semantic fluency, disorganized narrative structure, and decline in abstract thinking and synthesis.

Please note that this assessment is based solely on the provided conversation and cannot replace a formal diagnosis. It is always recommended to consult with a healthcare professional for a proper evaluation.
