## Supplementary material for "Text dialogue analysis Based ChatGPT for Primary Screening of Mild Cognitive Impairment": Data(GPT4_vs_GPT3.5_model1-5): 110_GPT3.5 (5).docx

ChatGPT

2 / 2

Based on the provided conversation, I will assess and rate the severity of each evaluation indicator for the participant's language features related to possible mild cognitive impairment (MCI).

Vocabulary features: a. Analyzing word frequency, word ratio, phrase frequency, and phrase ratio, as well as calculating lexical complexity, requires a larger sample of text. However, based on the limited conversation provided, it is not possible to assess these indicators.

b. Severity rating for tip-of-the-tongue phenomenon: 4 The participant occasionally experiences the tip-of-the-tongue phenomenon, as observed when they struggle to recall specific details or words. However, this phenomenon is not consistently evident throughout the conversation and does not significantly impede communication.

Syntax and grammar features: a. Analyzing syntactic complexity and grammatical components requires a larger sample of text. However, based on the provided conversation, it is possible to analyze some aspects.

b. Severity rating for difficulty with complex ideas: 2 The participant demonstrates the ability to comprehend and respond to simpler sentences and ideas. However, they indicate more difficulty when the narrative or question becomes more complex, as observed by their occasional pauses and clarification-seeking behavior.

Semantics features: a. Analyzing semantic density and coherence requires a larger sample of text. However, based on the provided conversation, it is possible to analyze some aspects.

b. Severity rating for memory issues: 3 The participant occasionally exhibits memory issues, such as repeating questions and struggling to recall specific details of the story or sequence. However, these memory issues are not pervasive and do not significantly hinder their overall comprehension and communication.

Based on the comprehensive synthesis of the available indicators, the participant can be classified into the Probably Normal Cognition Group. While there are some mild difficulties observed in certain aspects, such as the tip-of-the-tongue phenomenon and occasional difficulty with complex ideas and memory, these features alone do not indicate a probable mild cognitive impairment. It is important to note that a more comprehensive assessment and evaluation would be needed to make a definitive determination.
