## Supplementary material for "Text dialogue analysis Based ChatGPT for Primary Screening of Mild Cognitive Impairment": Data(GPT4_vs_GPT3.5_model1-5): 110_GPT3.5.docx

ChatGPT

Assessment of Vocabulary features:

Word frequency: Count the occurrences of each unique word in the text.

- Unique words: 118
- Total words: 222
- Word frequency: Each word appears once.

Word ratio: Calculate the proportion of unique words to the total number of words in the text.

- Word ratio: 118/222 ≈ 0.532

Phrase frequency: Count the occurrences of each unique phrase in the text.

- Unique phrases: 75
- Total phrases: 87
- Phrase frequency: Each phrase appears once.

Phrase ratio: Calculate the proportion of unique phrases to the total number of phrases in the text.

- Phrase ratio: 75/87 ≈ 0.862

Difficulty finding the right words: There are instances where the participant experiences difficulty finding the right words, indicated by the use of filler words like "uh" or "um" and pauses.

Lexical complexity: The participant demonstrates a moderate level of lexical complexity. They use a mix of simple and more complex words and phrases.

Limited vocabulary: The participant exhibits some repetition of words but overall demonstrates a reasonable range of vocabulary.

Assessment of Semantics features:

Semantic density: The participant's responses contain a moderate amount of meaningful information, with a good balance between content words (nouns, verbs, adjectives, adverbs) and function words (prepositions, pronouns, conjunctions).

Inability to convey meaning: The participant is generally able to express ideas clearly, although there are instances where they struggle to find the right words or make complete statements.

Semantic coherence: The participant's responses are generally logically consistent and meaningful. They provide descriptions of the picture and its elements, although some details may be omitted or vague.

Lack of coherence: The participant maintains a reasonable flow of ideas throughout the conversation, with thoughts appearing connected and related.

Assessment of Syntax and Grammar features:

Syntactic complexity: The participant's sentences are predominantly short and simple, with limited use of complex sentence structures or subordinate clauses. The average length of sentences is relatively short.

Simplified sentence structure: The participant primarily uses simple sentence structures with subject-verb-object patterns. There is a lack of more complex sentence constructions.

Grammatical components: The participant demonstrates a generally accurate usage of grammar, including subject-verb agreement, tense consistency, and pronoun usage.

Grammatical errors: The participant does not make frequent grammatical errors. There may be occasional slips in verb tense or subject-verb agreement, but overall, the grammar is reasonably correct.

Assignment to the Probably Normal group or Probably Mild Cognitive Impairment group:

Based on the language analysis, the participant's performance does not exhibit significant language gaps or cognitive impairments that would suggest mild cognitive impairment. The vocabulary, semantics, syntax, and grammar features demonstrate a level of language proficiency within the normal range. Therefore, the participant is more likely to be assigned to the Probably Normal group.
