## Supplementary material for "Text dialogue analysis Based ChatGPT for Primary Screening of Mild Cognitive Impairment": Data(GPT4_vs_GPT3.5_model1-5): 110_GPT4 (3).docx

ChatGPT

Analyzing the language sample provided by the PARTICIPANT using the indicators you have outlined, we can note the following:

Vocabulary Features:

1. Limited vocabulary: The participant uses simple words, and repeats certain phrases, which might indicate a slightly reduced vocabulary. Rating: 4/10
2. Increased verbosity: The participant uses a fair amount of detail in their descriptions, but doesn't seem excessively verbose. Rating: 2/10
3. Overlearned phrases: The participant uses some common phrases, but doesn't overuse cliches or stock phrases. Rating: 3/10
4. Improper use of pronouns: The participant doesn't seem to struggle with pronoun usage in this sample. Rating: 1/10
5. Hesitation (Word-finding difficulties): The participant uses "uh" and "…", indicating some hesitation. Rating: 5/10
6. Circumlocution (Word-finding difficulties): There is some evidence of circumlocution with phrases like "I presume that's a dish". Rating: 5/10
7. Tip-of-the-tongue phenomenon: There doesn't seem to be evidence of this issue in the sample. Rating: 1/10
8. Repetition: The participant repeats a few details, such as the boy reaching for cookies, but it's not excessive. Rating: 3/10

Syntax and Grammar Features:

1. Simplified sentence structure and grammar: Participant uses mostly simple sentences, but a few sentences contain subordinate clauses. Rating: 3/10
2. Grammatical errors: The participant makes very few grammatical errors in this sample. Rating: 2/10
3. Difficulty with complex ideas: The participant seems to handle the complex scene well, breaking it down into understandable descriptions. Rating: 2/10

Semantics Features:

1. Semantic paraphasias: The participant seems to express their ideas clearly. Rating: 2/10
2. Lack of coherence: The participant stays on topic and maintains focus. Rating: 1/10
3. Consistency in errors: There are few errors, and they don't appear to be consistent. Rating: 1/10
4. Lack of semantic fluency: The participant has no apparent trouble naming items in the scene or describing the sequence of events. Rating: 2/10
5. Disorganized narrative structure: The participant's narrative is mostly organized and chronological. Rating: 2/10
6. Orientation Issues: The participant seems clear on the sequence of events and the details of the scene. Rating: 1/10
7. Decline in conversational engagement and responsiveness: The participant actively engages with the task and provides detailed descriptions. Rating: 2/10
8. Memory issues: The participant repeats a few details, but it's not excessive, and may not indicate a memory issue. Rating: 3/10
9. Lack of abstract thinking and synthesis: The participant synthesizes the overall situation. Rating: 1/10
10. Abnormal emotions: There's no clear indication of abnormal emotions from the text provided. Rating: 1/10

Given the analysis above, the PARTICIPANT shows some signs of mild cognitive impairment, particularly in vocabulary (hesitation and circumlocution), but there are many areas where they perform well, particularly in maintaining coherence, semantic fluency, and engagement in conversation. Therefore, this evidence does not seem to clearly place them in either the Probably Normal Cognition Group or the Probably Mild Cognitive Impairment Group. It's a mixed presentation, with some signs of potential cognitive issues but also signs of normal cognitive function. It would be beneficial to have more data or conduct more comprehensive tests to draw a solid conclusion. However, if I were forced to dichotomise the PARTICIPANT, I would lean towards Probably Normal Cognition Group based on the current data. Please remember, this is a preliminary judgment and may not represent the actual cognitive condition of the PARTICIPANT. The professional clinical diagnosis is needed for accurate evaluation
