## Supplementary material for "Text dialogue analysis Based ChatGPT for Primary Screening of Mild Cognitive Impairment": Data(GPT4_vs_GPT3.5_model1-5): 110_GPT4 (4).docx

- Limited vocabulary: 2/10 (the participant's vocabulary seems appropriate for the task)
- Increased verbosity: 3/10 (participant was not excessively verbose but gave a detailed description)
- Overlearned phrases: 1/10 (no evidence of overused phrases)
- Improper use of pronouns: 1/10 (pronouns were used correctly)
- Hesitation: 4/10 (there are instances of hesitation and pauses in speech)
- Circumlocution: 1/10 (participant was straightforward)
- Tip-of-the-tongue phenomenon: 1/10 (there is no clear instance of the struggle in word finding)
- Repetition: 2/10 (the participant repeated certain phrases but not excessively)

Syntax and grammar features: a. Syntactic complexity appears normal; grammatical components are mostly well used. b.

- Simplified sentence structure and grammar: 2/10 (the task does not require complex sentence structure, but the participant demonstrated the ability to use complete sentences)
- Grammatical errors: 1/10 (few grammatical errors were noticed)
- Difficulty with complex ideas: 1/10 (the participant did not show difficulty with complex ideas)

Semantics features: a. Semantic density and coherence seem to be within the normal range. b.

- Semantic paraphasias: 1/10 (no instances of semantic paraphasias were noticed)
- Lack of coherence: 1/10 (the participant's responses were coherent)
- Consistency in errors: 1/10 (participant's errors did not seem to follow a specific pattern)
- Lack of semantic fluency: 2/10 (participant hesitated a few times)
- Disorganized narrative structure: 1/10 (there was no noticeable disorganized narrative structure)
- Orientation issues: 1/10 (no orientation issues were apparent)
- Decline in conversational engagement and responsiveness: 1/10 (participant was engaged and responsive)
- Memory issues: 1/10 (no clear memory issues are apparent)
- Lack of abstract thinking and synthesis: 1/10 (the task did not require significant abstract thinking)
- Abnormal emotions: 1/10 (no emotional content to assess)

Based on the evaluation of the indicators, it would be most appropriate to classify this participant in the "Probably Normal Cognition Group". The participant showed some hesitation but managed to give a detailed and coherent description of the picture. As always, these observations are not intended to provide a formal diagnosis but to serve as an initial screening tool for potential MCI. Further tests are recommended to obtain a more accurate evaluation.
