## Supplementary material for "Text dialogue analysis Based ChatGPT for Primary Screening of Mild Cognitive Impairment": Data(GPT4_vs_GPT3.5_model1-5): 110_GPT4 (5).docx

ChatGPT

Vocabulary features:

a. Lexical complexity is average, with some variation in the words used to describe the scene (e.g., "stool," "sink," "cookie jar") and limited use of uncommon or complex words. Word frequency and ratio show a good balance of unique words, pronouns, and specific names. The phrase frequency and ratio are also balanced, with some repetition of phrases related to the main actions in the picture (e.g., "cookie jar," "sink"). Based on these aspects, I would rate this at a 4 out of 10, considering that the person is capable of using varied vocabulary but doesn't reach high levels of complexity or uniqueness.

b. Tip-of-the-tongue phenomenon: There were a few pauses (marked by "uh" and "mm"), but these don't necessarily indicate a tip-of-the-tongue phenomenon. It's common for individuals to pause during speech. There's no evidence of the participant struggling to recall specific words. Rating: 3 out of 10.

Syntax and grammar features:

a. The syntactic complexity of the sentences is moderate, with a mixture of simple and compound sentences. There are a few instances of coordination and subordination (e.g., "mother's just standin(g) lookin(g)."). Grammatical components such as subject-verb agreement, tense consistency, and proper use of pronouns, prepositions, and conjunctions are mostly correct. I would rate this at 4 out of 10, as there is some complexity in the sentence structures and correct use of grammar.

b. Difficulty with complex ideas: The participant does not seem to struggle significantly with complex ideas or narratives. While the descriptions are mainly simple and straightforward, the participant manages to explain the potential future event (e.g., "stool that Junior's on is about ready to fall over"). Rating: 3 out of 10.

Semantics features:

a. Semantic density and coherence: The text is quite dense with meaningful information, with a high proportion of content words. The participant effectively connects ideas and maintains coherence throughout the description. Rating: 3 out of 10, as the participant's language is coherent and dense with content words.

b. Memory issues: The participant repeats some descriptions (e.g., the boy reaching for the cookies) but it's not clear if this is due to a memory issue or due to the repetition of the investigator's question. The participant seems to remember details well and provides a consistent narrative. Rating: 4 out of 10, considering that some repetition is present but it doesn't appear to be due to significant memory difficulties.

Based on the comprehensive synthesis of all the indicators, the participant is classified into the Probably Normal Cognition Group. However, slight issues with vocabulary complexity and minor repetition should be monitored in future evaluations
