## Supplementary material for "Text dialogue analysis Based ChatGPT for Primary Screening of Mild Cognitive Impairment": Data(GPT4_vs_GPT3.5_model1-5): 110_GPT4.docx

ChatGPT

Assessment:

Vocabulary features:

- Word frequency: 109 unique words
- Word ratio: 109 unique words / 268 total words = 0.407
- Phrase frequency: 75 unique phrases
- Phrase ratio: 75 unique phrases / 94 total phrases = 0.798
- Difficulty finding the right words: Mild difficulty observed with a few instances of "uh" and "um."
- Lexical complexity: Moderate complexity, with a mix of common and less frequent words.
- Limited vocabulary: Limited repetition, some variety in words used.

Semantics features:

- Semantic density: Moderate density, with a balance of content and function words.
- Inability to convey meaning: Minimal difficulty in expressing ideas clearly.
- Semantic coherence: Good coherence, with logical consistency and meaningfulness in the text.
- Lack of coherence: Minimal difficulty in maintaining a logical flow of ideas.

Syntax and Grammar features:

- Syntactic complexity: Moderate complexity, with some longer sentences and a few subordinate clauses.
- Simplified sentence structure: Some short sentences, but also more complex structures.
- Grammatical components: Mostly correct usage of grammar.
- Grammatical errors: Occasional dropping of "g" in gerunds (e.g., "standin(g)").

Group Assignment: Based on the language analysis above, I would assign PARTICIPANT to the Probably Normal group. While there are occasional difficulties with finding the right words and some grammatical errors, these issues do not seem significant enough to indicate mild cognitive impairment
