## Supplementary material for "Text dialogue analysis Based ChatGPT for Primary Screening of Mild Cognitive Impairment": Data(GPT4_vs_GPT3.5_model1-5): 111_GPT3.5 (2).docx

Attention! You do not need to diagnose, just follow the steps below to initially screen for PARTICIPANT with possibly mild cognitive impairment (MCI). Some differences between MCI and normal cognition are also given, please analyse them in detail, step by step. Firstly, please assess the Vocabulary features of PARTICIPANT at the Word frequency and Word ratio, Phrase frequency and Phrase ratio, Word-finding difficulties, Repetition, Lexical complexity, Limited vocabulary. Word frequency: Count the occurrences of each unique word in the text. Unique words are distinct words that appear in the text, not taking into account their repetitions. Each word is counted only once, regardless of how many times it occurs. Word ratio: Calculate the proportion of unique words to the total number of words in the text. Phrase frequency: Count the occurrences of each unique phrases in the text. Unique phrases are distinct combinations of words that appear in the text, not taking into account their repetitions. Phrase ratio: Calculate the proportion of unique phrases to the total number of phrases in the text. Word-finding difficulties: Frequent pauses or use of filler words, like "uh" or "um,". MCI: more frequent instances, longer time to find the right word, or using more general terms. Normal: fewer instances, and better lexical retrieval. Repetition: The participant repeats certain phrases or ideas a few times throughout the conversation. MCI: more frequent repetition. Normal: less frequent repetition (some repetition of words but not excessive) and better memory of what has been mentioned. Lexical complexity: Measure the variety and sophistication of the vocabulary used, considering factors, including word length and frequency of rare words. Limited vocabulary: Repeated use of the same words or simple words. MCI: slightly reduced vocabulary, and more frequent use of simpler words or common terms; Normal: richer vocabulary (low limitation). Secondly, please assess the Semantics features of PARTICIPANT, from Semantic density and Inability to convey meaning, Semantic coherence and Lack of coherence. Semantic density: Measure the amount of meaningful information in the text, and considering factor is the number of content words (nouns, verbs, adjectives, adverbs) relative to function words (prepositions, pronouns, conjunctions, etc.). Inability to convey meaning: Difficulty in expressing ideas clearly, leading to confusion or misunderstanding. Normal: the participant was able to describe the scene clearly. Semantic coherence: Assess the logical consistency and meaningfulness of the text, which can be done by analyzing the relationships between words, phrases, and sentences, and checking for coherence and cohesion. Lack of coherence: Difficulty in maintaining a logical flow of ideas, with thoughts appearing disjointed or unrelated. MCI: struggle with organizing thoughts coherently or maintaining a clear focus in conversation; Normal: greater coherence and organization in their storytelling and conversations. Thirdly, please assess the Syntax and Grammar features of PARTICIPANT, from Syntactic complexity and Simplified sentence structure, Grammatical components and Grammatical errors. Syntactic complexity: Measure the complexity of the sentence structure in the text, considering factors such as the number of clauses per sentence, the average length of sentences, and the use of subordinate clauses. Simplified sentence structure: A lack of complex sentence structures. MCI: simpler sentence structures; Normal: in longer dialogues there is a greater variety of sentence structures (moderately complex), but in short dialogues the sentences may turn out to be simple. Grammatical components: Analyze the text for correct usage of grammar, such as subject-verb agreement, tense consistency, and proper use of pronouns, prepositions, and conjunctions. MCI: avoidance of complex grammatical constructions; Normal: better use of complex grammatical constructions. Grammatical errors: Frequent mistakes in verb tense, subject-verb agreement, or other aspects of grammar. MCI: more frequent grammatical errors, such as mistakes in verb tense and subject-verb agreement; Normal: fewer grammatical errors (a few errors, but not significant). Finally, Please dichotomise PARTICIPANT into the Probably Normal group or the Probably Mild Cognitive Impairment group. PARTICIPANT with MCI might exhibit more difficulty with vocabulary, sentence structure, repetition, word-finding, coherence, and grammar. However, it is not definitive and requires a comprehensive judgement. Now, you do not have to respond to this dialogue. When I have entered THE DIALOGUE BETWEEN PARTICIPANT AND INVESTIGATOR, evaluate PARTICIPANT according to the evaluation rules above. INVESTIGATOR: okay can you just tell me everything that you see going on in this picture ? PARTICIPANT: a little boy trying to get a cookie for his sister who's about to fall …um …bec because he's standing on a stool that's tipping backwards . PARTICIPANT: a mother who's looking out the window wiping her dishes as the sink runs over to flood the kitchen . INVESTIGATOR: perfect . INVESTIGATOR: and for this next one …um there's gonna be another picture . INVESTIGATOR: …um this picture tells a story . INVESTIGATOR: so I'd like you to tell me a story about what you see going on in this picture that has a beginning a middle and an end . PARTICIPANT: …uh once upon a time a little girl was out riding her tricycle . PARTICIPANT: and she saw a cat …uh that was stuck on the limb of a tree . PARTICIPANT: …uh I guess that's her father …um tried to go get the cat . PARTICIPANT: and the dog was barking at the father for crying climbing the tree . PARTICIPANT: and in the background you have birds … a bird cheep … or … chirping . PARTICIPANT: …um the father is stuck . PARTICIPANT: someone who we can't see called the fire department . PARTICIPANT: the fire department arrived with a ladder to help the father down . PARTICIPANT: it looks like what really haptinau (happened) is the father …had his … got up the tree by using his ladder which fell to the ground . INVESTIGATOR: perfect . INVESTIGATOR: thank_you . INVESTIGATOR: and we're gonna do that one more time with a different picture . INVESTIGATOR: …um so it's like two pictures in one . INVESTIGATOR: …um but if you can tell me a story about what you see happening in this picture that has a beginning a middle and an end . PARTICIPANT: hm . PARTICIPANT: …uh you have …t … the same family shown riding on a trip with … the same father driving in the car in the top picture . PARTICIPANT: the son is leaning out the window . PARTICIPANT: and the dog is also in front of the son leaning out the window . PARTICIPANT: the daughter is blowing bubblegum . PARTICIPANT: and the … looks like a grandmother or grandfather's in the back . PARTICIPANT: somebody's in the back . PARTICIPANT: can't make out who it is . PARTICIPANT: and another son . PARTICIPANT: maybe twins (be)cause they have on the …s PARTICIPANT: I guess they're going someplace and everything … everyone is excited . PARTICIPANT: …uh on the bottom picture they are PARTICIPANT: wherever they were going, they're on their way home . PARTICIPANT: everyone is tired . PARTICIPANT: you can see the mother in the front . PARTICIPANT: and the little daughter that I couldn't see in the front in the first picture is sleeping . PARTICIPANT: the kids are hanging out the window . PARTICIPANT: and the grandmother and I guess another kid is sitting in the back . PARTICIPANT: and everyone is tired . PARTICIPANT: so that's it . INVESTIGATOR: perfect . INVESTIGATOR: alright . INVESTIGATOR: and I'm gonna have you tell me one more story now . INVESTIGATOR: oops . INVESTIGATOR: …uh are you familiar with the story of Cinderella ? PARTICIPANT: yes . PARTICIPANT: sort_of . PARTICIPANT: yeah . INVESTIGATOR: …um I can show you some pictures to kind_of jog your memory . PARTICIPANT: no …I'm just … I don't . PARTICIPANT: yeah I'm familiar with Cinderella . INVESTIGATOR: okay . INVESTIGATOR: can … can you tell me the story of Cinderella then ? PARTICIPANT: oh there's so many of them . PARTICIPANT: …um …cinder Cinderella was a stepdaughter to a stepmother after … Cinderella's father died . PARTICIPANT: the stepmother had two daughters who treated … …s along with the stepmother treated Cinderella poorly . PARTICIPANT: …uh the king in the kingdom where they lived decided it was time for his son to marry . PARTICIPANT: and he held a ball . PARTICIPANT: …cin …cin Cinderella …uh wanted to go to the ball . PARTICIPANT: but the stepsisters said . PARTICIPANT: no you don't have anything to wear yada_yada_yada . PARTICIPANT: and …um the fairy godmother came and took a pumpkin and turned it into a coach , gave her a glass slipper , and … and decked her out for the ball . PARTICIPANT: and she gave her some glass slippers . PARTICIPANT: but she cautioned her that she hadta be home by midnight . PARTICIPANT: Cinderella was having such a good time when she got to the ball that she didn't take notice of the time . PARTICIPANT: and all_of_a_sudden it was midnight . PARTICIPANT: she ran down the stairs to get back to the …um coach . PARTICIPANT: and on the way down the stairs she lost her slipper . PARTICIPANT: so when she got home, everything was back like it was . PARTICIPANT: and the prince who had fallen in love with Cinderella wanted to find her . PARTICIPANT: so he had …uh a search party going throughout the kingdom saying whoever could slip <in this picture I_mean … in this slipper …uh was the one and he would marry . PARTICIPANT: and he finally found after the stepsisters tried to put on the slipper . PARTICIPANT: they couldn't of_course . PARTICIPANT: but it fit Cinderella . PARTICIPANT: end of story . PARTICIPANT: she lived happily ever after as a princess . INVESTIGATOR: perfect . INVESTIGATOR: alright so I'm gonna have you tell me two more things . INVESTIGATOR: …um right now is …um first can you tell me how you would make a peanut butter and jelly sandwich ? PARTICIPANT: doodoo first I would get the …uh ingredient . PARTICIPANT: put … take two slices of bread out . PARTICIPANT: put peanut butter on one and jelly on the other . PARTICIPANT: put them two together . PARTICIPANT: and then cut them so … into bite size pieces . INVESTIGATOR: perfect .

ChatGPT

2 / 2

Based on the provided dialogue, let's analyze the participant's performance according to the evaluation rules mentioned.

1. Vocabulary Features:
   - Word Frequency: The participant's use of vocabulary seems adequate, but a more detailed analysis is required to determine word frequency.
   - Word Ratio: The unique word ratio cannot be determined from the provided dialogue.
   - Phrase Frequency: The participant's use of unique phrases is limited but requires a more detailed analysis.
   - Phrase Ratio: The unique phrase ratio cannot be determined from the provided dialogue.
   - Word-finding Difficulties: The participant exhibits some word-finding difficulties, such as pauses and the use of filler words like "uh" and "um." This suggests a potential cognitive impairment.
   - Repetition: The participant repeats certain phrases and ideas throughout the conversation, indicating possible mild cognitive impairment.
   - Lexical Complexity: The participant's lexical complexity cannot be determined from the provided dialogue.
   - Limited Vocabulary: The participant does not exhibit a significantly limited vocabulary based on the provided dialogue.
2. Semantics Features:
   - Semantic Density: The participant's use of content words (nouns, verbs, adjectives, adverbs) relative to function words (prepositions, pronouns, conjunctions, etc.) cannot be determined from the provided dialogue.
   - Inability to Convey Meaning: The participant is able to express ideas clearly and describes the scenes in the pictures adequately.
   - Semantic Coherence: The participant's responses demonstrate logical consistency and meaningfulness in describing the pictures.
   - Lack of Coherence: The participant maintains a relatively clear focus and organizes their thoughts coherently while describing the pictures.
3. Syntax and Grammar Features:
   - Syntactic Complexity: The participant's use of sentence structures does not exhibit significant complexity.
   - Simplified Sentence Structure: The participant tends to use simpler sentence structures, which is consistent with possible mild cognitive impairment.
   - Grammatical Components: The participant's usage of grammar appears to be mostly correct, without significant complexity.
   - Grammatical Errors: The participant makes a few minor grammatical errors, such as verb tense inconsistencies, but these errors are not significant.

Based on the analysis above, it suggests that the participant may exhibit mild cognitive impairment (MCI) based on their word-finding difficulties, repetition, and simplified sentence structures. However, a comprehensive judgment would require further evaluation and assessment beyond the provided dialogue. It is important to note that this analysis is based solely on the given dialogue and not a definitive diagnosis. A professional evaluation by a healthcare specialist is necessary to determine cognitive impairment accurately.
