## Supplementary material for "Text dialogue analysis Based ChatGPT for Primary Screening of Mild Cognitive Impairment": Data(GPT4_vs_GPT3.5_model1-5): 111_GPT3.5 (3).docx

ChatGPT

Based on the analysis of the indicators you provided, here is the evaluation of each indicator for the participant:

Vocabulary features:

1. Limited vocabulary: Severity: 3
2. Increased verbosity: Severity: 2
3. Overlearned phrases: Severity: 1
4. Improper use of pronouns: Severity: 2
5. Hesitation (Word-finding difficulties): Severity: 2
6. Circumlocution (Word-finding difficulties): Severity: 1
7. Tip-of-the-tongue phenomenon: Severity: 1
8. Repetition: Severity: 1

Syntax and Grammar features:

1. Simplified sentence structure and grammar: Severity: 2
2. Grammatical errors: Severity: 1
3. Difficulty with complex ideas: Severity: 1

Semantics features:

1. Semantic paraphasias: Severity: 1
2. Lack of coherence: Severity: 2
3. Consistency in errors: Severity: 1
4. Lack of semantic fluency: Severity: 1
5. Disorganized narrative structure: Severity: 2
6. Orientation issues: Severity: 1
7. Decline in conversational engagement and responsiveness: Severity: 1
8. Memory issues: Severity: 1
9. Lack of abstract thinking and synthesis: Severity: 1
10. Abnormal emotions: Severity: 1

Based on the overall evaluation of the indicators, it appears that the participant shows mild cognitive impairment (MCI). However, it is important to note that not all evaluation indicators are present, and some of the issues described can also be observed in individuals with normal cognitive function. Therefore, further comprehensive assessment and evaluation would be required to confirm the participant's cognitive status.
