## Supplementary material for "Text dialogue analysis Based ChatGPT for Primary Screening of Mild Cognitive Impairment": Data(GPT4_vs_GPT3.5_model1-5): 111_GPT3.5 (5).docx

ChatGPT

Based on the provided conversation with the participant, let's assess the indicators of possible mild cognitive impairment (MCI) based on the language features you mentioned.

Vocabulary features: a. Word frequency, word ratio, phrase frequency, and phrase ratio: It's difficult to analyze these features without the full text or a larger sample. However, from the given conversation, we can see that the participant uses a range of words and phrases, including some specific names (e.g., Cinderella, father, stepmother). We cannot determine the actual frequency or ratio without a more extensive analysis. b. Tip-of-the-tongue phenomenon: There is no clear evidence of the tip-of-the-tongue phenomenon in the participant's responses.

Syntax and grammar features: a. Syntactic complexity and grammatical components: The participant generally constructs grammatically correct sentences and uses a mixture of sentence structures. However, there are instances of incomplete sentences and sentence fragments, indicating some syntactic simplification. b. Difficulty with complex ideas: The participant appears to have more difficulty when the narrative or question becomes more complex, as seen in the third story prompt where they mentioned there are multiple versions of the Cinderella story. They also struggled to recall details and sequence of events.

Semantics features: a. Semantic density and coherence: The participant uses a reasonable amount of meaningful content words, but it's challenging to assess the semantic density without a larger sample. In terms of coherence, the participant's responses generally make sense and maintain logical consistency within the given context. b. Memory issues: The participant displays some memory issues, such as difficulty recalling specific details of the stories or sequence of events. However, it's important to note that memory issues can also occur in individuals with normal cognition, so it should be considered in the context of overall cognitive function.

Based on the provided conversation and the analysis of language features, we can make a preliminary assessment:

Severity ratings (1-10, with 1 being the least severe and 10 being the most severe):

1. Vocabulary features: Insufficient data to provide a rating.
2. Syntax and grammar features:
   - Syntactic complexity and grammatical components: 4
   - Difficulty with complex ideas: 6
3. Semantics features:
   - Semantic density and coherence: Insufficient data to provide a rating.
   - Memory issues: 4

Considering the ratings and the participant's performance, it's challenging to make a conclusive categorization into either the Probably Normal Cognition Group or the Probably Mild Cognitive Impairment Group. However, the participant's difficulty with complex ideas and memory issues may suggest a potential mild cognitive impairment. Further evaluation, including a comprehensive assessment, would be necessary to make a more accurate determination.
