## Supplementary material for "Text dialogue analysis Based ChatGPT for Primary Screening of Mild Cognitive Impairment": Data(GPT4_vs_GPT3.5_model1-5): 111_GPT3.5.docx

ChatGPT

2 / 2

Assessment of Vocabulary Features:

Word frequency: The participant uses a variety of words, including both common and more specific vocabulary. Word ratio: The proportion of unique words to total words is relatively high, indicating a good variety of vocabulary. Phrase frequency: The participant uses distinct phrases, but some repetitions are present. Phrase ratio: The proportion of unique phrases to total phrases is relatively high, indicating a decent variety of expressions. Difficulty finding the right words: The participant experiences occasional pauses and filler words ("uh," "um") while searching for specific words. Lexical complexity: The participant demonstrates a moderate level of lexical complexity, utilizing words of varying lengths and some less common terms. Limited vocabulary: There is no significant evidence of limited vocabulary, as the participant uses diverse words and phrases.

Assessment of Semantics Features:

Semantic density: The participant provides meaningful information and uses content words (nouns, verbs, adjectives, adverbs) to convey their ideas. Inability to convey meaning: The participant is generally able to express their ideas clearly, with only minor instances of ambiguity. Semantic coherence: The participant's responses show logical consistency and meaningful connections between words, phrases, and sentences. Lack of coherence: There is no significant lack of coherence, as the participant maintains a logical flow of ideas throughout the conversation.

Assessment of Syntax and Grammar Features:

Syntactic complexity: The participant primarily uses simple sentence structures with a few instances of more complex sentences containing multiple clauses. Simplified sentence structure: The participant tends to favor shorter, simpler sentences but demonstrates occasional use of complex structures. Grammatical components: The participant generally follows correct grammar rules, including subject-verb agreement and proper use of pronouns, prepositions, and conjunctions. Grammatical errors: There are minimal grammatical errors, and the participant demonstrates overall grammatical accuracy.

Assignment to Probably Normal group or Probably Mild Cognitive Impairment group:

Based on the conversation, the participant does not exhibit significant language gaps or difficulties that would suggest mild cognitive impairment. They demonstrate adequate vocabulary, semantic coherence, and syntactic skills, with only minor instances of word-finding difficulties. Therefore, the participant can be assigned to the Probably Normal group. However, it's important to note that this assessment is based solely on the linguistic aspects of the conversation and cannot provide a comprehensive diagnosis. A thorough evaluation by a healthcare professional would be necessary for an accurate assessment of cognitive functioning.
