## Supplementary material for "Text dialogue analysis Based ChatGPT for Primary Screening of Mild Cognitive Impairment": Data(GPT4_vs_GPT3.5_model1-5): 111_GPT4 (2).docx

ChatGPT

3 / 3

Let's break down the evaluation based on the criteria you provided.

**Vocabulary Features:**

Word frequency and ratio: The participant uses a variety of unique words and phrases throughout the dialogue, repeating only a few terms, and seems to have a good vocabulary breadth.

Phrase frequency and ratio: Again, there's a good variety in phrases used by the participant, indicating a strong vocabulary and phrase repertoire.

Word-finding difficulties: There are a few instances where the participant uses fillers like "uh" and "um", and a few instances of pause, but it does not seem excessive.

Repetition: The participant does not excessively repeat words or phrases. However, there are a couple of repetitions, like "glass slipper" in the Cinderella narrative, but it seems more related to the narrative itself rather than a cognitive issue.

Lexical complexity: The participant uses a diverse range of words and phrases, indicating a good level of lexical complexity.

Limited vocabulary: The participant does not show any significant signs of limited vocabulary. The language used is varied and not overly simplistic or repetitive.

**Semantics Features:**

Semantic density: The participant uses a good balance of content words and function words, indicating a high semantic density.

Inability to convey meaning: The participant does not show any significant difficulty in expressing ideas. The narratives provided are quite clear and detailed.

Semantic coherence: The participant's narratives are logically consistent and meaningful. The relationships between words, phrases, and sentences are coherent and cohesive.

Lack of coherence: The participant does not show any significant lack of coherence. The flow of ideas is logical and well organized.

**Syntax and Grammar Features:**

Syntactic complexity: The participant uses a variety of sentence structures, indicating a good level of syntactic complexity. There are complex sentences as well as simpler ones, showing a good balance.

Simplified sentence structure: The participant does not predominantly use simple sentences. There's a good mixture of complex and simple structures, indicating good syntactic ability.

Grammatical components: The participant uses proper grammar throughout the conversation, indicating a strong command of grammatical structures.

Grammatical errors: There are very few grammatical errors in the participant's speech. The errors present seem more related to conversational speech than cognitive issues.

**Final Assessment:**

Based on the above evaluation, it seems that the participant exhibits linguistic characteristics more consistent with the Probably Normal group. However, please note that this is only an initial screening and not a definitive diagnosis. If cognitive issues are suspected, a comprehensive evaluation by a professional is recommended
