## Supplementary material for "Text dialogue analysis Based ChatGPT for Primary Screening of Mild Cognitive Impairment": Data(GPT4_vs_GPT3.5_model1-5): 111_GPT4 (3).docx

ChatGPT

To begin, the participant's discourse and interaction will be evaluated according to the various features and indicators mentioned.

Vocabulary Analysis:

- Word frequency and Word ratio: It seems that the participant has a range of vocabulary. The repetition of words like "uh" and "…um" is seen, which is common in daily communication.
- Phrase frequency and Phrase ratio: The participant appears to employ a range of phrases during the interaction. The most frequently used phrase is "…uh," indicating hesitation or difficulty in finding words.
- Lexical complexity: The participant uses a variety of simple and complex words, indicating a good lexical complexity.

1. Limited vocabulary: 2/10 - The participant has a wide vocabulary and uses a range of words and phrases.
2. Increased verbosity: 4/10 - There is some evidence of verbosity, particularly in the use of filler words.
3. Overlearned phrases: 1/10 - The participant doesn't seem to rely heavily on overlearned phrases or cliches.
4. Improper use of pronouns: 1/10 - There's no apparent misuse of pronouns.
5. Hesitation (Word-finding difficulties): 5/10 - The frequent use of "…uh" and "…um" suggests some hesitation.
6. Circumlocution (Word-finding difficulties): 3/10 - There's a bit of circumlocution, but it's not excessive.
7. Tip-of-the-tongue phenomenon: 1/10 - There's little evidence of the tip-of-the-tongue phenomenon.
8. Repetition: 3/10 - There's some repetition, but it doesn't seem excessive.

Syntax and Grammar Analysis:

- Syntactic complexity: The participant uses a mix of simple and complex sentence structures, indicating good syntactic complexity.
- Grammatical components: The participant uses grammatical structures correctly most of the time.

1. Simplified sentence structure and grammar: 2/10 - The participant uses both simple and complex sentence structures.
2. Grammatical errors: 1/10 - There are few, if any, grammatical errors.
3. Difficulty with complex ideas: 1/10 - The participant doesn't appear to struggle with complex ideas.

Semantics Analysis:

- Semantic density: The participant uses a good mix of content words and function words, indicating good semantic density.
- Semantic coherence: The participant's discourse is logically consistent and meaningful.

1. Semantic paraphasias: 2/10 - There's a bit of difficulty in expressing ideas clearly, but it's not severe.
2. Lack of coherence: 1/10 - The participant's speech is mostly coherent.
3. Consistency in errors: 1/10 - There's little evidence of consistent errors.
4. Lack of semantic fluency: 2/10 - There's a bit of trouble with semantic fluency, but it's not severe.
5. Disorganized narrative structure: 2/10 - The participant's narrative is mostly well-organized.
6. Orientation Issues: 1/10 - The participant doesn't seem to struggle with orientation.
7. Decline in conversational engagement and responsiveness: 1/10 - The participant seems engaged in the conversation.
8. Memory issues: 2/10 - There's little evidence of significant memory issues.
9. Lack of abstract thinking and synthesis: 1/10 - The participant doesn't seem to struggle with abstract thinking and synthesis.
10. Abnormal emotions: 1/10 - There's no evidence of abnormal emotions.

Overall, based on this analysis, the participant seems to have a Probably Normal Cognition. However, it's worth mentioning that these findings are preliminary, and the participant should undergo comprehensive due to length
