## Supplementary material for "Text dialogue analysis Based ChatGPT for Primary Screening of Mild Cognitive Impairment": Data(GPT4_vs_GPT3.5_model1-5): 111_GPT4 (4).docx

ChatGPT

First, let's analyze the participant's language features:

**Vocabulary Features:**

a. There appears to be a moderate word and phrase frequency, although some pronouns such as 'he', 'it', 'her', 'they' are overused.

b. Lexical complexity appears to be about average. The participant uses a variety of vocabulary, though the language tends to be on the simpler side.

c.

- Limited vocabulary: 3/10
- Increased verbosity: 2/10
- Overlearned phrases: 3/10
- Improper use of pronouns: 3/10
- Hesitation: 5/10
- Circumlocution: 3/10
- Tip-of-the-tongue phenomenon: 4/10
- Repetition: 3/10

**Syntax and Grammar Features:**

a. The syntactic complexity is fairly typical, and the participant's grammar is mostly correct.

b.

- Simplified sentence structure and grammar: 4/10
- Grammatical errors: 3/10
- Difficulty with complex ideas: 3/10

**Semantics Features:**

a. Semantic density seems generally good, with a few instances of less coherence in the story of Cinderella and other narratives.

b.

- Semantic paraphasias: 3/10
- Lack of coherence: 4/10
- Consistency in errors: 3/10
- Lack of semantic fluency: 3/10
- Disorganized narrative structure: 3/10
- Orientation issues: 2/10
- Decline in conversational engagement and responsiveness: 3/10
- Memory issues: 3/10
- Lack of abstract thinking and synthesis: 3/10
- Abnormal emotions: 1/10

Based on the comprehensive synthesis of all the indicators, the participant would probably be categorized in the **Probably Normal Cognition Group**. While there are a few markers for potential MCI, they are generally mild and are not consistent or severe enough to suggest a probable MCI. Nonetheless, it may be beneficial for the participant to undergo further formal evaluation and monitoring, particularly if there are other symptoms or risk factors for MCI.

Remember, this analysis does not provide a diagnosis, but rather serves as an initial screening tool for potential MCI. Always consult with a healthcare provider for a comprehensive assessment.
