## Supplementary material for "Text dialogue analysis Based ChatGPT for Primary Screening of Mild Cognitive Impairment": Data(GPT4_vs_GPT3.5_model1-5): 111_GPT4 (5).docx

ChatGPT

Analyzing the language features of the participant, let's look into the evaluation indicators as per the provided criteria.

**Vocabulary features:** a. Overall, the participant uses a fairly diverse set of words, with clear nouns, verbs, and pronouns. Some sentences are highly detailed, which shows a strong ability to name objects and actions. Despite this, there are several instances of repetitive use of words and phrases, which could indicate a decrease in lexical complexity.

- Word Frequency: 6/10
- Word Ratio: 7/10
- Phrase Frequency: 6/10
- Phrase Ratio: 6/10
- Lexical Complexity: 6/10

b. The participant does display some difficulty in recalling specific words, which is evident from the use of phrases like "um", "uh", and hesitations before certain words. The tip-of-the-tongue phenomenon can be observed.

- Severity of Tip-of-the-tongue Phenomenon: 5/10

**Syntax and grammar features:** a. While the participant is generally able to form complete sentences, there are multiple instances of grammatical mistakes and pauses, which could indicate issues with syntax.

- Syntactic Complexity: 7/10
- Correct Use of Grammatical Components: 6/10

b. The participant does appear to have some difficulty with complex ideas. While they are able to recount the story of Cinderella, there are several hesitations and repeated phrases.

- Severity of Difficulty with Complex Ideas: 5/10

**Semantics features:** a. The participant is able to convey meaningful information, with logical progressions in their narrative. The coherence of the text is generally maintained, though it does falter in certain areas due to the pauses and hesitations.

- Semantic Density: 7/10
- Semantic Coherence: 7/10

b. The participant seems to remember the plot of Cinderella quite well, though there are some slight inconsistencies and hesitations. There aren't any noticeable repeated questions or significant difficulties in recalling information, aside from the occasional hesitations.

- Severity of Memory Issues: 4/10

**Overall Assessment:**

Based on this analysis, it seems the participant may have slight difficulties with word recall, syntax, and semantics, which can be indicative of Mild Cognitive Impairment. However, their memory seems to be functioning fairly well, as they are able to recall and recount the story of Cinderella with relatively few issues.

Taking into account all these factors, the participant appears to fall in the "Probably Mild Cognitive Impairment Group", but the signs are relatively mild and could also be explained by normal cognitive aging. Further and more comprehensive assessments would be necessary to determine a more precise classification. Remember that this analysis isn't a formal diagnosis, but merely a tool for initial screening and identification of potential MCI
