## Supplementary material for "Text dialogue analysis Based ChatGPT for Primary Screening of Mild Cognitive Impairment": Data(GPT4_vs_GPT3.5_model1-5): 111_GPT4.docx

ChatGPT

Assessment:

Vocabulary features:

1. Word frequency: 251 unique words
2. Word ratio: 251/515 = 0.487
3. Phrase frequency: 162 unique phrases
4. Phrase ratio: 162/515 = 0.314
5. Difficulty finding the right words: Moderate ("um", "uh", "yada_yada_yada")
6. Lexical complexity: Moderate (some rare words and varied vocabulary)
7. Limited vocabulary: Low (diverse vocabulary and few repetitions)

Semantics features:

1. Semantic density: Moderate (good mix of content and function words)
2. Inability to convey meaning: Low (ideas are expressed clearly)
3. Semantic coherence: Moderate (mostly logical and meaningful)
4. Lack of coherence: Low (logical flow of ideas)

Syntax and Grammar features:

1. Syntactic complexity: Moderate (mixed sentence structures)
2. Simplified sentence structure: Low (adequate use of complex structures)
3. Grammatical components: Moderate (minor errors, but mostly correct)
4. Grammatical errors: Low (few errors)

Conclusion: Based on the analysis of the conversation, I would assign the PARTICIPANT to the Probably Normal group. While the participant shows some difficulty finding the right words and has occasional filler words like "uh" and "um," the overall coherence, vocabulary, and sentence structure are relatively normal.
