## Supplementary material for "Text dialogue analysis Based ChatGPT for Primary Screening of Mild Cognitive Impairment": Data(GPT4_vs_GPT3.5_model1-5): 112_GPT3.5 (2).docx

Attention! You do not need to diagnose, just follow the steps below to initially screen for PARTICIPANT with possibly mild cognitive impairment (MCI). Some differences between MCI and normal cognition are also given, please analyse them in detail, step by step. Firstly, please assess the Vocabulary features of PARTICIPANT at the Word frequency and Word ratio, Phrase frequency and Phrase ratio, Word-finding difficulties, Repetition, Lexical complexity, Limited vocabulary. Word frequency: Count the occurrences of each unique word in the text. Unique words are distinct words that appear in the text, not taking into account their repetitions. Each word is counted only once, regardless of how many times it occurs. Word ratio: Calculate the proportion of unique words to the total number of words in the text. Phrase frequency: Count the occurrences of each unique phrases in the text. Unique phrases are distinct combinations of words that appear in the text, not taking into account their repetitions. Phrase ratio: Calculate the proportion of unique phrases to the total number of phrases in the text. Word-finding difficulties: Frequent pauses or use of filler words, like "uh" or "um,". MCI: more frequent instances, longer time to find the right word, or using more general terms. Normal: fewer instances, and better lexical retrieval. Repetition: The participant repeats certain phrases or ideas a few times throughout the conversation. MCI: more frequent repetition. Normal: less frequent repetition (some repetition of words but not excessive) and better memory of what has been mentioned. Lexical complexity: Measure the variety and sophistication of the vocabulary used, considering factors, including word length and frequency of rare words. Limited vocabulary: Repeated use of the same words or simple words. MCI: slightly reduced vocabulary, and more frequent use of simpler words or common terms; Normal: richer vocabulary (low limitation). Secondly, please assess the Semantics features of PARTICIPANT, from Semantic density and Inability to convey meaning, Semantic coherence and Lack of coherence. Semantic density: Measure the amount of meaningful information in the text, and considering factor is the number of content words (nouns, verbs, adjectives, adverbs) relative to function words (prepositions, pronouns, conjunctions, etc.). Inability to convey meaning: Difficulty in expressing ideas clearly, leading to confusion or misunderstanding. Normal: the participant was able to describe the scene clearly. Semantic coherence: Assess the logical consistency and meaningfulness of the text, which can be done by analyzing the relationships between words, phrases, and sentences, and checking for coherence and cohesion. Lack of coherence: Difficulty in maintaining a logical flow of ideas, with thoughts appearing disjointed or unrelated. MCI: struggle with organizing thoughts coherently or maintaining a clear focus in conversation; Normal: greater coherence and organization in their storytelling and conversations. Thirdly, please assess the Syntax and Grammar features of PARTICIPANT, from Syntactic complexity and Simplified sentence structure, Grammatical components and Grammatical errors. Syntactic complexity: Measure the complexity of the sentence structure in the text, considering factors such as the number of clauses per sentence, the average length of sentences, and the use of subordinate clauses. Simplified sentence structure: A lack of complex sentence structures. MCI: simpler sentence structures; Normal: in longer dialogues there is a greater variety of sentence structures (moderately complex), but in short dialogues the sentences may turn out to be simple. Grammatical components: Analyze the text for correct usage of grammar, such as subject-verb agreement, tense consistency, and proper use of pronouns, prepositions, and conjunctions. MCI: avoidance of complex grammatical constructions; Normal: better use of complex grammatical constructions. Grammatical errors: Frequent mistakes in verb tense, subject-verb agreement, or other aspects of grammar. MCI: more frequent grammatical errors, such as mistakes in verb tense and subject-verb agreement; Normal: fewer grammatical errors (a few errors, but not significant). Finally, Please dichotomise PARTICIPANT into the Probably Normal group or the Probably Mild Cognitive Impairment group. PARTICIPANT with MCI might exhibit more difficulty with vocabulary, sentence structure, repetition, word-finding, coherence, and grammar. However, it is not definitive and requires a comprehensive judgement. Now, you do not have to respond to this dialogue. When I have entered THE DIALOGUE BETWEEN PARTICIPANT AND INVESTIGATOR, evaluate PARTICIPANT according to the evaluation rules above. INVESTIGATOR: there is the picture . PARTICIPANT: doesn't matter where you start then right ? INVESTIGATOR: nope . PARTICIPANT: alright . PARTICIPANT: there's …uh a little boy on a …uh stepstool which is tilted . PARTICIPANT: and he's getting some cookies . PARTICIPANT: now do you want descriptive as far as the cookie lids or … or how they are or just the actions ? INVESTIGATOR: just the action . PARTICIPANT: just the action . PARTICIPANT: okay . PARTICIPANT: and looks like he's trying to hand a cookie to his sister . PARTICIPANT: she kind_of has her finger up towards her mouth or something like she might be giggling . PARTICIPANT: …uh the mother's drying dishes but the water is overflowing onto the floor . PARTICIPANT: …uh I don't know, the silk ( sink ) … the sink looks like it's almost tilted . PARTICIPANT: now we should call the … …uh the … …sau the cup and the saucers . PARTICIPANT: that's not action right is it ? INVESTIGATOR: no you don't needta mention those . PARTICIPANT: …uh the breeze might be blowing . PARTICIPANT: the curtains look like they might be ... PARTICIPANT: any other action . PARTICIPANT: I don't see anything else . PARTICIPANT: I just see into the outdoors but I don't see anything moving . INVESTIGATOR: …hm okay . PARTICIPANT: that's about it I guess . INVESTIGATOR: a little bit xxx . PARTICIPANT: … INVESTIGATOR: this picture . INVESTIGATOR: and I want you to tell me everything that you see happening there everything that's going on . PARTICIPANT: okay . PARTICIPANT: anywhere ? INVESTIGATOR: everything that's happening . PARTICIPANT: …um the little girl's laughing . PARTICIPANT: the little boy's getting cookies and he's falling . PARTICIPANT: …uh I guess the mother would be doing dishes and the water's overflowing out_of the sink . PARTICIPANT: …uh the window's open and I would say the wind is blowing . INVESTIGATOR: okay ? PARTICIPANT: I guess that's it . INVESTIGATOR: alright .

ChatGPT

Based on the provided dialogue between the investigator and participant, we can assess the participant's language abilities using the criteria you mentioned. Please note that this analysis is based solely on the provided dialogue and should be considered as an initial screening, not a definitive diagnosis. Here's the evaluation based on the specified features:

Vocabulary features:

- Word frequency: We would need the full text to count the occurrences of each unique word, so we cannot assess this feature based on the provided dialogue.
- Word ratio: We cannot calculate the word ratio without the full text.
- Phrase frequency: We would need the full text to count the occurrences of each unique phrase, so we cannot assess this feature based on the provided dialogue.
- Phrase ratio: We cannot calculate the phrase ratio without the full text.
- Word-finding difficulties: The participant did not display significant word-finding difficulties in this dialogue. They were able to express their ideas relatively smoothly without frequent pauses or the use of filler words.
- Repetition: The participant did not excessively repeat phrases or ideas in this dialogue. Some repetition occurred, but it did not seem excessive.
- Lexical complexity: Without the full text, it's difficult to assess the variety and sophistication of the participant's vocabulary.

Semantics features:

- Semantic density: We cannot measure semantic density without the full text.
- Inability to convey meaning: The participant was able to convey their ideas relatively clearly in this dialogue.
- Semantic coherence: The participant's responses showed logical consistency and meaningfulness. Their descriptions of the scene were coherent and cohesive.
- Lack of coherence: The participant did not display a lack of coherence in this dialogue. Their thoughts appeared connected and organized.

Syntax and Grammar features:

- Syntactic complexity: The participant used mostly simple sentence structures in this dialogue. There were no complex sentence structures observed.
- Simplified sentence structure: The participant's sentences were relatively simple, with straightforward structures.
- Grammatical components: The participant showed basic grammatical competence in this dialogue, using correct subject-verb agreement and proper use of pronouns, prepositions, and conjunctions.
- Grammatical errors: There were no significant grammatical errors observed in the participant's dialogue. The participant demonstrated overall grammatical accuracy.

Based on this initial assessment, the participant did not exhibit significant difficulties with vocabulary, sentence structure, repetition, word-finding, coherence, or grammar in the provided dialogue. However, please note that a comprehensive evaluation should involve more extensive language samples and assessments to make a definitive judgment about cognitive impairment.
