## Supplementary material for "Text dialogue analysis Based ChatGPT for Primary Screening of Mild Cognitive Impairment": Data(GPT4_vs_GPT3.5_model1-5): 112_GPT3.5 (3).docx

ChatGPT

2 / 2

Based on the provided conversation, here is an analysis of the indicators and evaluation for the participant regarding mild cognitive impairment (MCI):

Vocabulary features:

- Word frequency and Word ratio: The participant uses a limited range of words and tends to repeat certain words. (Severity: 6)
- Phrase frequency and Phrase ratio: The participant doesn't use a wide variety of phrases and tends to repeat certain phrases. (Severity: 4)
- Lexical complexity: The participant's vocabulary lacks sophistication and contains mostly simple words. (Severity: 5)

Evaluation indicators based on Vocabulary features:

1. Limited vocabulary: The participant exhibits a slightly reduced vocabulary and more frequent use of the same words or simple/common terms. (Severity: 6)
2. Increased verbosity: The participant tends to be more verbose and uses more words than necessary. (Severity: 3)
3. Overlearned phrases: There are no indications of overuse of cliches or stock phrases. (Severity: 1)
4. Improper use of pronouns: There are no indications of difficulty with pronouns or improper use. (Severity: 1)
5. Hesitation (Word-finding difficulties): The participant shows some hesitations, using filler words such as "uh" and "um." (Severity: 4)
6. Circumlocution (Word-finding difficulties): The participant doesn't exhibit significant circumlocution. (Severity: 1)
7. Tip-of-the-tongue phenomenon: There are no indications of the tip-of-the-tongue phenomenon. (Severity: 1)
8. Repetition: The participant repeats certain statements within a short span of time. (Severity: 6)

Syntax and Grammar features:

- Syntactic complexity: The participant demonstrates a relatively simplified sentence structure and grammar. (Severity: 4)
- Grammatical components: There are no indications of significant grammatical errors. (Severity: 1)

Evaluation indicators based on Syntax and Grammar features:

1. Simplified sentence structure and grammar: The participant tends to prefer simpler structures in both long and short sentences and avoids complex grammatical constructions. (Severity: 5)
2. Grammatical errors: The participant doesn't make significant grammatical errors. (Severity: 1)
3. Difficulty with complex ideas: The participant doesn't exhibit difficulty with complex ideas. (Severity: 1)

Semantics features:

- Semantic density: The participant's speech contains a moderate amount of meaningful information. (Severity: 5)
- Semantic coherence: The participant's speech lacks coherence at times, with changes in topic and indirect speech. (Severity: 6)

Evaluation indicators based on Semantics features:

1. Semantic paraphasias: There are no indications of difficulty in word choice or odd word combinations. (Severity: 1)
2. Lack of coherence: The participant struggles with organizing thoughts coherently and may change topics frequently. (Severity: 7)
3. Consistency in errors: There are no indications of consistent language errors. (Severity: 1)
4. Lack of semantic fluency: There are no indications of difficulty in naming items or describing processes. (Severity: 1)
5. Disorganized narrative structure: The participant struggles to maintain a coherent line of thought and the narrative lacks key elements. (Severity: 7)
6. Orientation issues: There are no indications of orientation issues. (Severity: 1)
7. Decline in conversational engagement and responsiveness: The participant shows decreased initiation, difficulties in following a conversation, and a tendency to steer conversations back to familiar topics. (Severity: 6)
8. Memory issues: There are no indications of memory issues in the provided conversation. (Severity: 1)
9. Lack of abstract thinking and synthesis: There are no indications of difficulty in abstract thinking or synthesis. (Severity: 1)
10. Abnormal emotions: There are no indications of abnormal emotions. (Severity: 1)

Based on the comprehensive analysis of the indicators, the participant's conversation suggests a probable mild cognitive impairment (MCI). However, it is important to note that some indicators may also be present in individuals with normal cognitive function, and the assessment should be evaluated in the context of the individual's overall cognitive abilities.
