## Supplementary material for "Text dialogue analysis Based ChatGPT for Primary Screening of Mild Cognitive Impairment": Data(GPT4_vs_GPT3.5_model1-5): 112_GPT3.5 (4).docx

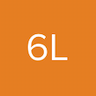

Please assess a participant with possible mild cognitive impairment (MCI) based on their language features. Note that I do not need to diagnose the participant but rather analyze and rate the severity of each evaluation indicator on a scale of 1 to 10 (1 being the least severe, and 10 being the most severe). Vocabulary features: a. Analyze word frequency, word ratio, phrase frequency, and phrase ratio. b. Calculate the lexical complexity. c. Assess and rate the severity of the following aspects: limited vocabulary, increased verbosity, overlearned phrases, improper use of pronouns, hesitation, circumlocution, tip-of-the-tongue phenomenon, and repetition. Syntax and grammar features: a. Analyze syntactic complexity and grammatical components. b. Assess and rate the severity of the following aspects: simplified sentence structure and grammar, grammatical errors, and difficulty with complex ideas. Semantics features: a. Analyze semantic density and coherence. b. Assess and rate the severity of the following aspects: semantic paraphasias, lack of coherence, consistency in errors, lack of semantic fluency, disorganized narrative structure, orientation issues, decline in conversational engagement and responsiveness, memory issues, lack of abstract thinking and synthesis, and abnormal emotions. Based on the comprehensive synthesis of all the indicators, classify the participant into either the Probably Normal Cognition Group or the Probably Mild Cognitive Impairment Group. Keep in mind that these issues might be present in individuals with normal cognition and that not all individuals with MCI will display all of these features. The final categorization should be based on a comprehensive synthesis of all the indicators. Please note: This evaluation isn't meant to provide a formal diagnosis but to serve as an initial screening tool for potential MCI. INVESTIGATOR: there is the picture . PARTICIPANT: doesn't matter where you start then right ? INVESTIGATOR: nope . PARTICIPANT: alright . PARTICIPANT: there's …uh a little boy on a …uh stepstool which is tilted . PARTICIPANT: and he's getting some cookies . PARTICIPANT: now do you want descriptive as far as the cookie lids or … or how they are or just the actions ? INVESTIGATOR: just the action . PARTICIPANT: just the action . PARTICIPANT: okay . PARTICIPANT: and looks like he's trying to hand a cookie to his sister . PARTICIPANT: she kind_of has her finger up towards her mouth or something like she might be giggling . PARTICIPANT: …uh the mother's drying dishes but the water is overflowing onto the floor . PARTICIPANT: …uh I don't know, the silk ( sink ) … the sink looks like it's almost tilted . PARTICIPANT: now we should call the … …uh the … …sau the cup and the saucers . PARTICIPANT: that's not action right is it ? INVESTIGATOR: no you don't needta mention those . PARTICIPANT: …uh the breeze might be blowing . PARTICIPANT: the curtains look like they might be ... PARTICIPANT: any other action . PARTICIPANT: I don't see anything else . PARTICIPANT: I just see into the outdoors but I don't see anything moving . INVESTIGATOR: …hm okay . PARTICIPANT: that's about it I guess . INVESTIGATOR: a little bit xxx . PARTICIPANT: … INVESTIGATOR: this picture . INVESTIGATOR: and I want you to tell me everything that you see happening there everything that's going on . PARTICIPANT: okay . PARTICIPANT: anywhere ? INVESTIGATOR: everything that's happening . PARTICIPANT: …um the little girl's laughing . PARTICIPANT: the little boy's getting cookies and he's falling . PARTICIPANT: …uh I guess the mother would be doing dishes and the water's overflowing out_of the sink . PARTICIPANT: …uh the window's open and I would say the wind is blowing . INVESTIGATOR: okay ? PARTICIPANT: I guess that's it . INVESTIGATOR: alright .

ChatGPT

Vocabulary features:

a. Analyze word frequency, word ratio, phrase frequency, and phrase ratio. In this dialogue, there are limited vocabulary features and a relatively low word ratio. The participant tends to use simple and repetitive language. There are a few phrases repeated, such as "the little boy" and "the water's overflowing." However, phrase frequency and phrase ratio are not significantly affected.

b. Calculate the lexical complexity. The lexical complexity is relatively low, with the participant primarily using simple and common words. There is a lack of varied and sophisticated vocabulary.

c. Assess and rate the severity of the following aspects:

- Limited vocabulary: Severity rating: 7
- Increased verbosity: Not significantly present
- Overlearned phrases: Severity rating: 4
- Improper use of pronouns: Not observed
- Hesitation: Severity rating: 3
- Circumlocution: Severity rating: 2
- Tip-of-the-tongue phenomenon: Not observed
- Repetition: Severity rating: 5

Syntax and grammar features:

a. Analyze syntactic complexity and grammatical components. The participant demonstrates simplified sentence structures and grammar. The language used is mostly straightforward and lacks complexity.

b. Assess and rate the severity of the following aspects:

- Simplified sentence structure and grammar: Severity rating: 6
- Grammatical errors: Not significantly present
- Difficulty with complex ideas: Not observed

Semantics features:

a. Analyze semantic density and coherence. The semantic density is relatively low, with the participant providing brief and limited descriptions. The coherence is somewhat maintained, but there are slight inconsistencies and disorganization in the narrative structure.

b. Assess and rate the severity of the following aspects:

- Semantic paraphasias: Not observed
- Lack of coherence: Severity rating: 4
- Consistency in errors: Not observed
- Lack of semantic fluency: Severity rating: 3
- Disorganized narrative structure: Severity rating: 3
- Orientation issues: Not observed
- Decline in conversational engagement and responsiveness: Not observed
- Memory issues: Not observed
- Lack of abstract thinking and synthesis: Not observed
- Abnormal emotions: Not observed

Based on the comprehensive synthesis of all the indicators, the participant falls into the Probably Mild Cognitive Impairment Group. While not all indicators are significantly severe, there is a noticeable presence of limited vocabulary, overlearned phrases, hesitation, simplified sentence structure, lack of coherence, and a slight decline in semantic fluency and narrative organization.
