## Supplementary material for "Text dialogue analysis Based ChatGPT for Primary Screening of Mild Cognitive Impairment": Data(GPT4_vs_GPT3.5_model1-5): 112_GPT3.5 (5).docx

ChatGPT

2 / 2

Let's analyze the language features of the participant and rate the severity of each evaluation indicator:

Vocabulary features: a. Word frequency and word ratio: The participant's use of words seems to be relatively straightforward and without any noticeable issues. They are able to articulate their observations using appropriate vocabulary. Severity rating: 1/10 b. Phrase frequency and phrase ratio: The participant utilizes simple phrases to describe the scene. There is no evidence of any notable difficulty with phrase production. Severity rating: 1/10 c. Lexical complexity: The participant's vocabulary appears to be adequate for describing the scene, without significant use of rare words or overly complex language. Severity rating: 1/10

Syntax and grammar features: a. Syntactic complexity: The participant constructs simple sentences and does not exhibit any evident difficulty with sentence structure. Severity rating: 1/10 b. Grammatical components: The participant demonstrates proper usage of grammar, including subject-verb agreement and appropriate use of pronouns, prepositions, and conjunctions. Severity rating: 1/10

Semantics features: a. Semantic density: The participant's use of content words (nouns, verbs, adjectives, adverbs) and function words (prepositions, pronouns, conjunctions) seems balanced and appropriate for the task. Severity rating: 1/10 b. Semantic coherence: The participant's responses exhibit logical consistency and meaningfulness. There is no evident lack of coherence or cohesion in their descriptions. Severity rating: 1/10

Overall assessment: Based on the analysis of the language features, the participant does not exhibit significant signs of language impairment or cognitive decline. The observed language features are consistent with normal cognition, and there is no strong evidence to suggest mild cognitive impairment (MCI). Therefore, the participant can be classified into the Probably Normal Cognition Group.

It's important to note that this assessment is based solely on the provided language sample, and a comprehensive evaluation of cognitive function requires a broader assessment encompassing various domains beyond language. A professional evaluation by a healthcare provider is necessary for an accurate diagnosis or further assessment of cognitive impairment.
