## Supplementary material for "Text dialogue analysis Based ChatGPT for Primary Screening of Mild Cognitive Impairment": Data(GPT4_vs_GPT3.5_model1-5): 112_GPT3.5.docx

ChatGPT

Based on the assessment of language features, here is the analysis for each category:

Vocabulary features:

- Word frequency: The participant uses a variety of unique words, indicating a decent vocabulary range.
- Word ratio: The proportion of unique words to total words is relatively high, suggesting a good vocabulary diversity.
- Phrase frequency: The participant uses a reasonable number of unique phrases, resulting in fewer repetitions.
- Phrase ratio: The proportion of unique phrases to total phrases is relatively high, indicating a good phrase diversity.
- Difficulty finding the right words: The participant does not exhibit frequent pauses or use filler words while searching for the desired word.
- Lexical complexity: The vocabulary used by the participant shows some variety and sophistication, with a mix of basic and more complex words.
- Limited vocabulary: The participant uses a decent range of words, avoiding excessive repetition and demonstrating some variety and sophistication.

Semantics features:

- Semantic density: The participant provides a moderate amount of meaningful information in their responses, with a balanced ratio of content words to function words.
- Inability to convey meaning: The participant expresses ideas relatively clearly, with less difficulty in conveying meaning compared to previous examples.
- Semantic coherence: The participant's responses show logical consistency and meaningfulness, with better coherence and cohesion between words, phrases, and sentences.
- Lack of coherence: The participant maintains a logical flow of ideas, with thoughts appearing more connected and related.

Syntax and Grammar features:

- Syntactic complexity: The participant uses a mix of sentence structures, including some complex sentences and variations in clause usage.
- Simplified sentence structure: The participant demonstrates a good balance between simple and complex sentence structures, showing more variety.
- Grammatical components: The participant generally displays correct usage of grammar, including subject-verb agreement, tense consistency, and proper use of pronouns, prepositions, and conjunctions.
- Grammatical errors: The participant makes fewer grammatical errors, showing improvement in verb tense, subject-verb agreement, and other aspects of grammar.

Based on the language assessment, the participant demonstrates better language skills compared to previous examples. The vocabulary usage, semantic coherence, and syntactic complexity are more developed, indicating a higher level of language proficiency. Therefore, based on the provided conversation, the participant is more likely to belong to the Probably Normal group. However, it's important to note that this is an initial screening and further evaluation would be required to make a definitive determination regarding cognitive impairment
