## Supplementary material for "Text dialogue analysis Based ChatGPT for Primary Screening of Mild Cognitive Impairment": Data(GPT4_vs_GPT3.5_model1-5): 112_GPT4 (2).docx

ChatGPT

Given the information, the participant's language abilities will be analyzed according to the provided guidelines.

**Vocabulary Features:**

*Word Frequency and Word Ratio:* This would require a full transcript and computational analysis for accurate results. Given the scope of this text, the participant seems to repeat words such as "uh", "the", and "and" often.

*Phrase Frequency and Phrase Ratio:* Similar to the word frequency and word ratio, a complete transcript and computational analysis would be required for this.

*Word-finding Difficulties:* There are several instances of filler words like "uh" and pauses which may indicate some difficulty in finding words.

*Repetition:* The participant repeats certain ideas such as the boy getting cookies, the mother drying dishes, and the water overflowing.

*Lexical Complexity:* The participant uses a relatively simple vocabulary, with occasional use of more complex phrases such as "finger up towards her mouth or something like she might be giggling."

*Limited Vocabulary:* The participant does use a number of repeated words and simple phrases, but also incorporates some unique words and descriptions.

**Semantics Features:**

*Semantic Density:* The participant uses a significant number of content words relative to function words, indicating a reasonable level of semantic density.

*Inability to Convey Meaning:* The participant is able to convey the scene clearly, with only a few instances of uncertainty or confusion.

*Semantic Coherence:* The participant is generally coherent in their descriptions, with logical connections between phrases and sentences.

*Lack of Coherence:* The participant does not struggle significantly with maintaining a logical flow of ideas, although there are a few instances of disjointed thoughts, particularly when considering the action in the picture.

**Syntax and Grammar Features:**

*Syntactic Complexity:* The participant uses a mix of simple and complex sentences, with some usage of subordinate clauses.

*Simplified Sentence Structure:* There are instances of simplified sentence structures, but also instances of more complex sentences.

*Grammatical Components:* The participant uses grammar correctly for the most part, with appropriate usage of subject-verb agreement, tense consistency, and pronouns, prepositions, and conjunctions.

*Grammatical Errors:* There are minimal grammatical errors noted in the text.

Based on these observations, the participant may fall into the "Probably Normal" group, as they display a reasonable level of vocabulary, semantics, syntax, and grammar abilities. However, the use of filler words and some repetition may suggest potential areas to monitor. As always, it is important to note that this is a preliminary assessment and a more comprehensive evaluation would be necessary for a definitive conclusion
