## Supplementary material for "Text dialogue analysis Based ChatGPT for Primary Screening of Mild Cognitive Impairment": Data(GPT4_vs_GPT3.5_model1-5): 112_GPT4 (3).docx

ChatGPT

Given the provided transcript of the conversation, let's analyze the different linguistic aspects for indicators of possible mild cognitive impairment (MCI). Please note that as an AI language model, I don't have the ability to listen to the tone, pitch or any non-verbal communication in the interaction, which are significant factors in such an evaluation.

Firstly, let's look at the Vocabulary features:

1. Limited vocabulary: The participant uses simple words and doesn't exhibit a rich vocabulary, for instance "falling", "laughing", "blowing". This indicates some reduction in vocabulary. Score: 5/10
2. Increased verbosity: The participant doesn't appear to be particularly verbose. Score: 3/10
3. Overlearned phrases: There are a few potentially overlearned phrases, such as "doesn't matter", "alright", "okay". Score: 4/10
4. Improper use of pronouns: The participant uses pronouns correctly. Score: 2/10
5. Hesitation (Word-finding difficulties): The participant frequently pauses and uses fillers like "uh", suggesting word-finding difficulties. Score: 7/10
6. Circumlocution (Word-finding difficulties): The participant does not seem to talk in a roundabout way but seems to struggle with finding specific words. Score: 6/10
7. Tip-of-the-tongue phenomenon: The participant does not exhibit the tip-of-the-tongue phenomenon. Score: 2/10
8. Repetition: There is some repetition, like "just the action" and "okay", but not excessive. Score: 4/10

Secondly, let's evaluate the Syntax and Grammar features:

1. Simplified sentence structure and grammar: The participant uses simple sentences and structures, but this could be due to the simplicity of the task. Score: 4/10
2. Grammatical errors: There is one error ("the silk (sink)"), indicating some problems with word-finding or articulation. Score: 6/10
3. Difficulty with complex ideas: The participant does not seem to have difficulty with complex ideas. Score: 3/10

Lastly, we'll analyze the Semantics features:

1. Semantic paraphasias: The participant has some difficulty expressing ideas clearly and makes some errors in word choice. Score: 6/10
2. Lack of coherence: The participant is mostly coherent, but has some minor issues with clarity. Score: 4/10
3. Consistency in errors: The participant doesn't make consistent language errors. Score: 2/10
4. Lack of semantic fluency: The participant has some issues with naming items or describing a sequence of events. Score: 5/10
5. Disorganized narrative structure: The participant is generally organized in their narrative. Score: 3/10
6. Orientation Issues: There are no obvious issues with orientation. Score: 2/10
7. Decline in conversational engagement and responsiveness: The participant seems to engage in conversation, but there are occasional difficulties in following. Score: 4/10
8. Memory issues: No obvious memory issues are observed. Score: 2/10
9. Lack of abstract thinking and synthesis: The participant does not seem to have major issues with abstract thinking. Score: 3/10
10. Abnormal emotions: No apparent abnormal emotions. Score: 2/10

In summary, based on the transcript, there are some potential indicators of MCI, such as hesitation (word-finding difficulties), simplification in sentence structure and grammar, semantic paraphasias, and lack of semantic fluency. However, these indicators are relatively mild. Therefore, the participant would likely be placed in the Probably Normal Cognition Group, with a recommendation for further testing due to the presence of a few potential MCI indicators. Please note that a qualified healthcare professional should make the final diagnosis
