## Supplementary material for "Text dialogue analysis Based ChatGPT for Primary Screening of Mild Cognitive Impairment": Data(GPT4_vs_GPT3.5_model1-5): 112_GPT4 (4).docx

ChatGPT

To analyze the language features of the participant, we'll examine vocabulary, syntax and grammar, and semantic features. Keep in mind this evaluation is not exhaustive due to the limited data provided and without real-time interaction or further context. Here is an assessment based on the information given:

Vocabulary Features: a. Word frequency seems to be relatively consistent with some repeated phrases such as "the little boy", "the cookies", and "the mother". Phrase frequency is also consistent and normal for the context. b. Lexical complexity seems to be in a normal range. The participant was able to use a variety of words to describe the scene. c. As for the various aspects:

- Limited vocabulary: 3/10. The participant was able to use a variety of words and phrases to describe the scene. However, there is still some repetition.
- Increased verbosity: 2/10. There's no notable verbosity.
- Overlearned phrases: 3/10. The participant did use some common phrases, but they were contextually appropriate.
- Improper use of pronouns: 1/10. The participant seemed to use pronouns appropriately.
- Hesitation: 4/10. The participant demonstrated some hesitation, but it was not excessive.
- Circumlocution: 2/10. Minimal circumlocution was noted.
- Tip-of-the-tongue phenomenon: 3/10. There were instances of this, for example, the moment with "…sau the cup and the saucers."
- Repetition: 3/10. There was some repetition, but it was not excessive.

Syntax and Grammar Features: a. The participant's syntactic complexity and grammatical components seem to be within normal limits. The sentences were well-structured, and there were both simple and complex sentences. b. In terms of the different aspects:

- Simplified sentence structure and grammar: 2/10. The participant used a mix of sentence structures, demonstrating a good grasp of grammar.
- Grammatical errors: 2/10. There were a few instances, but they were minor.
- Difficulty with complex ideas: 3/10. The participant was able to tell stories using complex ideas.

Semantic Features: a. Semantic density and coherence seemed normal. The participant was able to maintain a coherent narrative and used appropriate semantic content for the context. b. As for the various aspects:

- Semantic paraphasias: 1/10. There was no clear evidence of this.
- Lack of coherence: 2/10. The participant was able to maintain a coherent narrative.
- Consistency in errors: 2/10. There were some consistency in the participant's minor errors, but not severe.
- Lack of semantic fluency: 3/10. There were minor issues, but the participant was generally fluent.
- Disorganized narrative structure: 2/10. The narrative was mostly well-organized.
- Orientation issues: 1/10. No clear evidence of this.
- Decline in conversational engagement and responsiveness: 1/10. The participant was actively engaged and responsive.
- Memory issues: 1/10. No clear evidence of memory problems based on this extract.
- Lack of abstract thinking and synthesis: 2/10. The participant was able to think abstractly, synthesizing a story from the picture.
- Abnormal emotions: 1/10. There were no apparent signs of abnormal emotions.

Based on this brief analysis, the participant seems to belong to the Probably Normal Cognition Group, but keep in mind that this is a very tentative classification based on a short sample of their speech and is not a definitive assessment. An in-person, comprehensive evaluation by a healthcare provider is always necessary for accurate diagnosis
