## Supplementary material for "Text dialogue analysis Based ChatGPT for Primary Screening of Mild Cognitive Impairment": Data(GPT4_vs_GPT3.5_model1-5): 112_GPT4 (5).docx

ChatGPT

Vocabulary features:

a. There is a relatively broad variety of unique words and phrases used in the text, demonstrating a fairly decent lexical complexity. The proportion of unique words to the total number of words is moderate. There is a significant repetition of certain phrases related to the context of the picture, which is expected in this situation. However, the participant does show some use of synonyms (e.g., "the little boy", "his sister", "the mother") which indicates a moderate level of lexical complexity.

Word Frequency: 6 Word Ratio: 6 Phrase Frequency: 5 Phrase Ratio: 5 Lexical Complexity: 6

b. There are instances where the participant seems to struggle to find the right words (e.g., "…uh the silk ( sink )", "… …uh the … …sau the cup and the saucers"), but it's not prevalent and does not seem to significantly impact communication.

Tip-of-the-tongue phenomenon: 5

Syntax and grammar features:

a. The syntactic complexity and grammatical components used by the participant are generally correct. Sentences are not overly complex but still maintain a certain level of complexity, with the usage of subordinate clauses and proper subject-verb agreement.

Syntactic Complexity: 5 Grammatical Components: 6

b. There is no clear evidence of difficulty with complex ideas. The participant appears to describe the picture effectively with relatively simple and straightforward language.

Difficulty with complex ideas: 2

Semantics features:

a. The semantic density of the text is fairly high, with a good number of content words used in relation to function words. The semantic coherence of the text is also quite high, as the participant describes the picture in a coherent, logical manner.

Semantic Density: 7 Semantic Coherence: 7

b. There are no clear signs of memory issues present in the text. The participant does not repeat questions or show difficulty in recalling information.

Memory Issues: 2

Based on these indicators, it would appear that the participant falls into the Probably Normal Cognition Group. There are a few minor areas of concern (such as occasional word-finding difficulty), but they do not appear to significantly impact the participant's communication or comprehension. However, these concerns may warrant further monitoring and evaluation. As always, this evaluation should be supplemented by additional diagnostic measures and considered in the broader context of the participant's overall cognitive function.
