## Supplementary material for "Text dialogue analysis Based ChatGPT for Primary Screening of Mild Cognitive Impairment": Data(GPT4_vs_GPT3.5_model1-5): 112_GPT4.docx

ChatGPT

Assessment:

Vocabulary features:

1. Word frequency: 129 unique words
2. Word ratio: 129/202 = 0.638
3. Phrase frequency: 96 unique phrases
4. Phrase ratio: 96/202 = 0.475
5. Difficulty finding the right words: Moderate ("uh", "um")
6. Lexical complexity: Moderate (some rare words and varied vocabulary)
7. Limited vocabulary: Low (diverse vocabulary and few repetitions)

Semantics features:

1. Semantic density: Moderate (good mix of content and function words)
2. Inability to convey meaning: Low (ideas are expressed clearly)
3. Semantic coherence: High (logical and meaningful)
4. Lack of coherence: Low (logical flow of ideas)

Syntax and Grammar features:

1. Syntactic complexity: Moderate (mixed sentence structures)
2. Simplified sentence structure: Low (adequate use of complex structures)
3. Grammatical components: Moderate (minor errors, but mostly correct)
4. Grammatical errors: Low (few errors)

Based on the assessment of the conversation between PARTICIPANT and INVESTIGATOR, I would assign the PARTICIPANT to the Probably Normal group. The participant demonstrates moderate difficulty finding the right words, but overall, their language features are relatively strong. Semantic coherence is high, and there are few instances of limited vocabulary or simplified sentence structures.
