## Supplementary material for "Text dialogue analysis Based ChatGPT for Primary Screening of Mild Cognitive Impairment": Data(GPT4_vs_GPT3.5_model1-5): 113_GPT3.5 (2).docx

Attention! You do not need to diagnose, just follow the steps below to initially screen for PARTICIPANT with possibly mild cognitive impairment (MCI). Some differences between MCI and normal cognition are also given, please analyse them in detail, step by step. Firstly, please assess the Vocabulary features of PARTICIPANT at the Word frequency and Word ratio, Phrase frequency and Phrase ratio, Word-finding difficulties, Repetition, Lexical complexity, Limited vocabulary. Word frequency: Count the occurrences of each unique word in the text. Unique words are distinct words that appear in the text, not taking into account their repetitions. Each word is counted only once, regardless of how many times it occurs. Word ratio: Calculate the proportion of unique words to the total number of words in the text. Phrase frequency: Count the occurrences of each unique phrases in the text. Unique phrases are distinct combinations of words that appear in the text, not taking into account their repetitions. Phrase ratio: Calculate the proportion of unique phrases to the total number of phrases in the text. Word-finding difficulties: Frequent pauses or use of filler words, like "uh" or "um,". MCI: more frequent instances, longer time to find the right word, or using more general terms. Normal: fewer instances, and better lexical retrieval. Repetition: The participant repeats certain phrases or ideas a few times throughout the conversation. MCI: more frequent repetition. Normal: less frequent repetition (some repetition of words but not excessive) and better memory of what has been mentioned. Lexical complexity: Measure the variety and sophistication of the vocabulary used, considering factors, including word length and frequency of rare words. Limited vocabulary: Repeated use of the same words or simple words. MCI: slightly reduced vocabulary, and more frequent use of simpler words or common terms; Normal: richer vocabulary (low limitation). Secondly, please assess the Semantics features of PARTICIPANT, from Semantic density and Inability to convey meaning, Semantic coherence and Lack of coherence. Semantic density: Measure the amount of meaningful information in the text, and considering factor is the number of content words (nouns, verbs, adjectives, adverbs) relative to function words (prepositions, pronouns, conjunctions, etc.). Inability to convey meaning: Difficulty in expressing ideas clearly, leading to confusion or misunderstanding. Normal: the participant was able to describe the scene clearly. Semantic coherence: Assess the logical consistency and meaningfulness of the text, which can be done by analyzing the relationships between words, phrases, and sentences, and checking for coherence and cohesion. Lack of coherence: Difficulty in maintaining a logical flow of ideas, with thoughts appearing disjointed or unrelated. MCI: struggle with organizing thoughts coherently or maintaining a clear focus in conversation; Normal: greater coherence and organization in their storytelling and conversations. Thirdly, please assess the Syntax and Grammar features of PARTICIPANT, from Syntactic complexity and Simplified sentence structure, Grammatical components and Grammatical errors. Syntactic complexity: Measure the complexity of the sentence structure in the text, considering factors such as the number of clauses per sentence, the average length of sentences, and the use of subordinate clauses. Simplified sentence structure: A lack of complex sentence structures. MCI: simpler sentence structures; Normal: in longer dialogues there is a greater variety of sentence structures (moderately complex), but in short dialogues the sentences may turn out to be simple. Grammatical components: Analyze the text for correct usage of grammar, such as subject-verb agreement, tense consistency, and proper use of pronouns, prepositions, and conjunctions. MCI: avoidance of complex grammatical constructions; Normal: better use of complex grammatical constructions. Grammatical errors: Frequent mistakes in verb tense, subject-verb agreement, or other aspects of grammar. MCI: more frequent grammatical errors, such as mistakes in verb tense and subject-verb agreement; Normal: fewer grammatical errors (a few errors, but not significant). Finally, Please dichotomise PARTICIPANT into the Probably Normal group or the Probably Mild Cognitive Impairment group. PARTICIPANT with MCI might exhibit more difficulty with vocabulary, sentence structure, repetition, word-finding, coherence, and grammar. However, it is not definitive and requires a comprehensive judgement. Now, you do not have to respond to this dialogue. When I have entered THE DIALOGUE BETWEEN PARTICIPANT AND INVESTIGATOR, evaluate PARTICIPANT according to the evaluation rules above. INVESTIGATOR: what I'd like you to do is just to tell me everything that you see going on in the picture . INVESTIGATOR: okay ? INVESTIGATOR: everything you see going on in the picture . PARTICIPANT: well Johnny's in the cookie jar handin(g) …um cookies to his sister who has her fingers up to her lips . PARTICIPANT: they're at the … …uh on the … at the … he's at the …uh cupboard above the countertop . PARTICIPANT: the mother has her back to them washing … …uh drying dishes . PARTICIPANT: the water's overflowing from the sink . PARTICIPANT: the window's up . PARTICIPANT: and she has on a dress with an apron . PARTICIPANT: and …um cups are on the countertop . PARTICIPANT: and the water is on the floor . PARTICIPANT: oh and … and he's about to fall because the …uh stepstool is tilting because he's up there getting the cookie jar … getting into the cookie jar . PARTICIPANT: that's all I can … that's there that I can see . " 7:25 " PARTICIPANT: well the family was very happy that uncle Bill was coming in on the fourth of July . PARTICIPANT: and Johnny was especially happy because he could remember when he and his uncle went trout fishing on the river . PARTICIPANT: and they had a very good time . PARTICIPANT: and that's all I remember . INVESTIGATOR: okay that's fine . " 21:33 " INVESTIGATOR: can you think of anything from that story I read you ? PARTICIPANT: the … the uncle was coming in …uh to visit this family …uh on the fourth of July . PARTICIPANT: and the … the little boy . PARTICIPANT: Johnny I think his name was . PARTICIPANT: was very happy to see him because he …uh remembered what good times they had had going trout fishing . INVESTIGATOR: (o)kay anything else ? PARTICIPANT: hmhunh . INVESTIGATOR: okay that's fine . " 23:20 " INVESTIGATOR: anything at all that you can think of from that story . PARTICIPANT: Melody's grandfather visited them for Thanksgiving . PARTICIPANT: and she was very happy to have him visit because she enjoyed the …s storytelling . PARTICIPANT: and one day he … she wanted to visit his childhood home . PARTICIPANT: but she was very frightened of the noise in the city . PARTICIPANT: and was very glad ... PARTICIPANT: although she enjoyed … (en)joyed going there . PARTICIPANT: but she was very glad when she came home . INVESTIGATOR: okay . INVESTIGATOR: okay good . INVESTIGATOR: here's the first word . INVESTIGATOR: pencil . PARTICIPANT: the pencil fell on the floor . INVESTIGATOR: tree . PARTICIPANT: the tree is growing beautifully . INVESTIGATOR: now I'm gonna give you two words . INVESTIGATOR: don't worry about the order that I give you the two words in . INVESTIGATOR: just use both of them in one short simple sentence . INVESTIGATOR: child , hospital . PARTICIPANT: what is it ? INVESTIGATOR: child , hospital . PARTICIPANT: oh ! PARTICIPANT: the child is in the hospital . INVESTIGATOR: cold , winter . PARTICIPANT: yesterday was a cold winter day . INVESTIGATOR: now I'm gonna give you three words . INVESTIGATOR: I don't want you to worry about the order that I give them to you . INVESTIGATOR: just give me a short simple sentence using all three . INVESTIGATOR: chair , doctor , sit . PARTICIPANT: sit in the doctor's chair . INVESTIGATOR: bureau , open , drawer . PARTICIPANT: open the bureau drawer .

ChatGPT

Based on the provided dialogue between the investigator and the participant, let's assess the participant's performance according to the evaluation rules outlined.

First, let's analyze the Vocabulary features:

- Word frequency: It's not possible to count the occurrences of each unique word in the given dialogue without the complete transcript.
- Word ratio: Similarly, we cannot calculate the proportion of unique words to the total number of words without the complete transcript.
- Phrase frequency: Again, we cannot determine the occurrences of unique phrases without the complete transcript.
- Phrase ratio: Similarly, we cannot calculate the proportion of unique phrases to the total number of phrases without the complete transcript.
- Word-finding difficulties: The participant shows some instances of pauses and filler words ("uh" and "um") but demonstrates relatively good lexical retrieval. They are able to find the right words, although there are a few instances of longer pauses or general terms used.
- Repetition: The participant does not exhibit excessive repetition in their dialogue. Some repetition of phrases or ideas occurs, but it is not significantly frequent.
- Lexical complexity: Without the complete transcript, it is difficult to measure the variety and sophistication of the participant's vocabulary in detail.
- Limited vocabulary: The participant does not display a significant limitation in their vocabulary. They use descriptive words and provide some details in their responses.

Next, let's assess the Semantics features:

- Semantic density: It's not possible to measure the semantic density without the complete transcript.
- Inability to convey meaning: The participant is able to express their ideas clearly and convey the scene in the picture adequately.
- Semantic coherence: The participant's responses demonstrate logical consistency and meaningfulness. They provide information about the picture and follow the conversation coherently.
- Lack of coherence: The participant does not exhibit a lack of coherence in their storytelling. Their thoughts are organized, and there is a clear focus in the conversation.

Moving on to the Syntax and Grammar features:

- Syntactic complexity: It's challenging to assess the complexity of the participant's sentence structure without the complete transcript.
- Simplified sentence structure: Based on the provided dialogue, the participant's sentences are mostly simple in structure, with straightforward subject-verb-object patterns.
- Grammatical components: The participant generally uses correct grammar in their responses, including subject-verb agreement and proper use of pronouns, prepositions, and conjunctions.
- Grammatical errors: There are no significant grammatical errors in the participant's dialogue. They make occasional minor mistakes, but these errors are not frequent or substantial.

Based on the limited information provided in the dialogue, it's difficult to make a definitive judgment about whether the participant falls into the Probably Normal group or the Probably Mild Cognitive Impairment group. However, from the given dialogue, the participant demonstrates relatively good performance in terms of vocabulary, semantics, syntax, and grammar. They exhibit some mild word-finding difficulties and simplified sentence structures, but overall their performance does not strongly suggest mild cognitive impairment. A comprehensive evaluation would require additional information and assessment.
