## Supplementary material for "Text dialogue analysis Based ChatGPT for Primary Screening of Mild Cognitive Impairment": Data(GPT4_vs_GPT3.5_model1-5): 113_GPT3.5 (3).docx

ChatGPT

Based on the provided conversation, here is the analysis and evaluation of the indicators for the participant:

Vocabulary Features:

- Word frequency and Word ratio: Not analyzed in the given conversation.
- Phrase frequency and Phrase ratio: Not analyzed in the given conversation.
- Lexical complexity: Not analyzed in the given conversation.

Evaluation Indicators:

1. Limited vocabulary: Not observed.
2. Increased verbosity: Not observed.
3. Overlearned phrases: Not observed.
4. Improper use of pronouns: Not observed.
5. Hesitation (Word-finding difficulties): Some instances of hesitation observed, such as "uh" and "um." Severity: 3
6. Circumlocution (Word-finding difficulties): Not observed.
7. Tip-of-the-tongue phenomenon: Not observed.
8. Repetition: Some repetition observed, such as mentioning the cookie jar multiple times. Severity: 2

Syntax and Grammar Features:

- Syntactic complexity: Not analyzed in the given conversation.
- Grammatical components: Not analyzed in the given conversation.

Evaluation Indicators:

1. Simplified sentence structure and grammar: Not analyzed.
2. Grammatical errors: Not analyzed.
3. Difficulty with complex ideas: Not observed.

Semantics Features:

- Semantic density: Not analyzed in the given conversation.
- Semantic coherence: Not analyzed in the given conversation.

Evaluation Indicators:

1. Semantic paraphasias: Not observed.
2. Lack of coherence: Some instances of changing topics and going off on tangents. Severity: 4
3. Consistency in errors: Not observed.
4. Lack of semantic fluency: Not observed.
5. Disorganized narrative structure: Some difficulty in maintaining a coherent line of thought. Severity: 3
6. Orientation issues: Not observed.
7. Decline in conversational engagement and responsiveness: Not observed.
8. Memory issues: Not observed.
9. Lack of abstract thinking and synthesis: Not observed.
10. Abnormal emotions: Not observed.

Dichotomization: Based on the observed indicators, it is likely that the participant falls into the Probably Normal Cognition Group. The indicators of mild cognitive impairment (MCI) are either not observed or present at a low severity level. However, it's important to note that the analysis is based on a limited conversation and should be evaluated in the context of the participant's overall cognitive function.
