## Supplementary material for "Text dialogue analysis Based ChatGPT for Primary Screening of Mild Cognitive Impairment": Data(GPT4_vs_GPT3.5_model1-5): 113_GPT3.5 (4).docx

ChatGPT

Vocabulary Features: a. Word frequency, word ratio, phrase frequency, and phrase ratio: Without access to the full transcript or a larger sample of language, it is not possible to analyze these features accurately. More extensive language samples would be needed to assess the participant's word and phrase usage patterns.

b. Lexical complexity: The provided dialogue does not provide sufficient information to evaluate the participant's lexical complexity.

c. Assessment and rating of severity:

- Limited vocabulary: The participant demonstrates a relatively limited vocabulary, using simple and common words. Severity: 5
- Increased verbosity: The participant tends to provide more information than necessary, using additional words and phrases. Severity: 4
- Overlearned phrases: There is no evidence of overlearned phrases in the given conversation.
- Improper use of pronouns: There is no evidence of improper use of pronouns in the given conversation.
- Hesitation: The participant displays hesitation, indicated by the use of filler words such as "um" and pauses. Severity: 4
- Circumlocution: The participant employs circumlocution in some instances, struggling to find precise words or phrases. Severity: 3
- Tip-of-the-tongue phenomenon: There is no evidence of tip-of-the-tongue phenomenon in the given conversation.
- Repetition: The participant repeats certain words and phrases. Severity: 3

Syntax and Grammar Features: a. Syntactic complexity and grammatical components: The provided dialogue does not offer enough information to analyze the participant's syntactic complexity and grammatical components.

b. Assessment and rating of severity:

- Simplified sentence structure and grammar: The participant demonstrates relatively simple sentence structures and grammar, using straightforward sentence constructions. Severity: 4
- Grammatical errors: There is no clear evidence of grammatical errors in the given conversation.
- Difficulty with complex ideas: The participant does not exhibit difficulty with complex ideas in the provided dialogue.

Semantics Features: a. Semantic density and coherence: Without access to a larger language sample, it is difficult to assess semantic density and coherence accurately.

b. Assessment and rating of severity:

- Semantic paraphasias: There are no evident semantic paraphasias in the given conversation.
- Lack of coherence: The participant's responses generally maintain coherence and logical connections. Severity: 2
- Consistency in errors: There is no clear evidence of consistent errors in the given conversation.
- Lack of semantic fluency: The participant displays some difficulty in generating ideas and providing detailed descriptions. Severity: 3
- Disorganized narrative structure: The participant's narrative structure appears relatively organized with identifiable story elements.
- Orientation issues: There is no evidence of orientation issues in the given conversation.
- Decline in conversational engagement and responsiveness: The participant actively engages in the conversation and responds to the investigator's prompts. Severity: 2
- Memory issues: There is no clear evidence of memory issues in the given conversation.
- Lack of abstract thinking and synthesis: The participant demonstrates a limited ability to engage in abstract thinking and synthesis. Severity: 4
- Abnormal emotions: There is no evidence of abnormal emotions in the given conversation.

Based on the comprehensive synthesis of all the indicators, it is challenging to definitively categorize the participant into either the Probably Normal Cognition Group or the Probably Mild Cognitive Impairment Group due to the limited language sample provided. However, some features, such as limited vocabulary, increased verbosity, hesitation, circumlocution, repetition, simplified sentence structures, and a limited ability for abstract thinking and synthesis, suggest the possibility of mild cognitive impairment. It is important to note that a more extensive evaluation would be necessary to make a more accurate determination.
