## Supplementary material for "Text dialogue analysis Based ChatGPT for Primary Screening of Mild Cognitive Impairment": Data(GPT4_vs_GPT3.5_model1-5): 113_GPT3.5 (5).docx

Severity rating: 4 The participant demonstrates relatively average vocabulary features. There are no major issues with word frequency, word ratio, phrase frequency, or phrase ratio. The lexical complexity is difficult to assess without a larger sample of text, but there are no clear indications of low variety or excessive reliance on simple vocabulary.

b. Assessing the severity of the tip-of-the-tongue phenomenon:

Severity rating: 3 There is no explicit evidence of the tip-of-the-tongue phenomenon in the provided conversation. The participant does not display notable difficulties in recalling words or experiencing partial recall. However, a thorough assessment requires additional context and information.

Syntax and Grammar Features: a. Analyzing syntactic complexity and grammatical components:

- Syntactic complexity: Measure the complexity of sentence structure, considering factors such as the number of clauses per sentence, average sentence length, and use of subordinate clauses.
- Grammatical components: Analyze the text for correct grammar usage, including subject-verb agreement, tense consistency, and proper use of pronouns, prepositions, and conjunctions.

Severity rating: 5 The participant's syntax and grammar features are relatively average. The sentences are generally coherent, and there are no major grammatical errors or obvious issues with sentence complexity. However, a more comprehensive analysis would require a larger sample of text to assess the participant's overall syntactic ability.

b. Assessing the severity of difficulty with complex ideas:

Severity rating: 6 While the participant can handle simpler sentences and ideas, they display some difficulty when the narrative or question becomes more complex. This is evidenced by their incomplete responses and occasional hesitations when asked to recall details or provide additional information. However, further assessment is necessary to confirm the extent of the difficulty.

Semantics Features: a. Analyzing semantic density and coherence:

- Semantic density: Measure the amount of meaningful information in the text, considering the ratio of content words (nouns, verbs, adjectives, adverbs) to function words (prepositions, pronouns, conjunctions, etc.).
- Semantic coherence: Assess the logical consistency and meaningfulness of the text, checking for coherence and cohesion.

Severity rating: 4 The participant's semantic features are relatively average. The provided responses contain meaningful information, and there are no obvious incoherencies or inconsistencies. However, a more extensive analysis would require a larger sample of text to evaluate semantic density and coherence accurately.

b. Assessing the severity of memory issues:

Severity rating: 4 While the participant doesn't explicitly exhibit memory issues in the provided conversation, it is mentioned that they have difficulty recalling certain information or sequences. However, memory issues can also be a part of normal aging, so a comprehensive evaluation is needed to determine the significance of the memory difficulties.

Based on the comprehensive synthesis of all indicators, the participant can be classified as being in the Probably Normal Cognition Group. While there are some indications of potential difficulties with complex ideas and memory, the severity ratings suggest that the participant's language features are within the range of what could be expected for someone with normal cognition. However, it is important to conduct a thorough evaluation to make a definitive determination.
