## Supplementary material for "Text dialogue analysis Based ChatGPT for Primary Screening of Mild Cognitive Impairment": Data(GPT4_vs_GPT3.5_model1-5): 113_GPT4 (2).docx

ChatGPT

Let's analyze the text based on the given instructions.

1. Vocabulary features:
   - Word frequency & Word ratio: The participant uses a variety of unique words, demonstrating a fairly wide range of vocabulary. There aren't many repeated words, suggesting a good word ratio.
   - Phrase frequency & Phrase ratio: The participant uses a diverse range of phrases. There isn't an excessive repetition of phrases, which suggests a good phrase ratio.
   - Word-finding difficulties: The participant exhibits some word-finding difficulties as indicated by frequent use of filler words like "uh" and pauses, suggesting potential mild cognitive impairment.
   - Repetition: The participant repeats certain information, but it's not excessive, which could be seen as normal.
   - Lexical complexity: The participant uses a mixture of simple and moderately complex words, indicating a moderate level of lexical complexity.
   - Limited vocabulary: The participant does not excessively use the same words or overly simple words, suggesting a normal level of vocabulary.
2. Semantics features:
   - Semantic density: The participant uses a fair amount of content words relative to function words, indicating a good semantic density.
   - Inability to convey meaning: The participant is able to express ideas clearly, suggesting normal cognitive functioning.
   - Semantic coherence & Lack of coherence: The participant maintains logical consistency and meaningfulness in the text. The thoughts are organized and related, suggesting normal cognitive functioning.
3. Syntax and Grammar features:
   - Syntactic complexity & Simplified sentence structure: The participant uses sentences of moderate complexity, with a mix of simple and complex sentence structures, suggesting normal cognitive functioning.
   - Grammatical components & Grammatical errors: The participant uses correct grammar most of the time, although there are a few instances of grammatical errors. However, these mistakes are not frequent, indicating normal cognitive functioning.

Based on this analysis, I would assign the PARTICIPANT to the "Probably Normal" group. While there are some signs of word-finding difficulties, these are not excessive and the participant demonstrates good vocabulary, semantic, and syntactic abilities overall. It's important to note that this is a preliminary analysis and more comprehensive assessments would be required for a definitive conclusion.
