## Supplementary material for "Text dialogue analysis Based ChatGPT for Primary Screening of Mild Cognitive Impairment": Data(GPT4_vs_GPT3.5_model1-5): 113_GPT4 (3).docx

ChatGPT

Analyzing the provided text, we'll evaluate the linguistic and cognitive indicators to screen for potential Mild Cognitive Impairment (MCI) in the PARTICIPANT.

1. Vocabulary features:
   - Word and Phrase frequency and ratio: The participant uses a variety of words and phrases, indicating normal vocabulary. However, there are instances of repetition and hesitation, suggesting possible MCI (e.g., "the water's overflowing from the sink... the water is on the floor").
   - Lexical complexity: The participant uses a mix of simple and complex words and phrases, indicating normal cognitive function.
   - Limited vocabulary: Severity 3/10. The participant uses a mix of simple and complex vocabulary, but there are instances of repetition and filler words.
   - Increased verbosity: Severity 1/10. The participant is concise and to the point in most of their statements.
   - Overlearned phrases: Severity 2/10. The participant does not seem to overly rely on cliches or stock phrases.
   - Improper use of pronouns: Severity 2/10. The participant appears to use pronouns correctly.
   - Hesitation (Word-finding difficulties): Severity 4/10. There are several instances of pauses and filler words, suggesting some word-finding difficulty.
   - Circumlocution (Word-finding difficulties): Severity 3/10. Some instances suggest circumlocution, but it's not a prevalent issue.
   - Tip-of-the-tongue phenomenon: Severity 3/10. Some pauses may indicate this phenomenon, but it's not clearly manifested.
   - Repetition: Severity 3/10. There are some instances of repetition but not excessively.
2. Syntax and Grammar features:
   - Syntactic complexity: The participant uses a mix of simple and complex sentences, suggesting normal cognitive function.
   - Grammatical components: Few grammar errors are evident, suggesting normal cognitive function.
   - Simplified sentence structure and grammar: Severity 2/10. The participant primarily uses correct and varied sentence structures.
   - Grammatical errors: Severity 2/10. There are few grammatical errors in the participant's speech.
   - Difficulty with complex ideas: Severity 2/10. The participant appears to be able to handle complex ideas well.
3. Semantics features:
   - Semantic density and coherence: The participant's narrative is generally coherent and dense with information, suggesting normal cognitive function.
   - Semantic paraphasias: Severity 2/10. The participant generally expresses ideas clearly with few errors in word choice.
   - Lack of coherence: Severity 3/10. The participant's narratives are generally coherent, but some instances show a slight lack of focus.
   - Consistency in errors: Severity 2/10. No consistent language errors are evident in the participant's speech.
   - Lack of semantic fluency: Severity 2/10. The participant seems to have little trouble naming items or describing sequences of events.
   - Disorganized narrative structure: Severity 3/10. The participant's narratives are generally well-structured, but some instances show a slight lack of coherence.
   - Orientation Issues: Severity 2/10. The participant seems to have an accurate sense of time and place.
   - Decline in conversational engagement and responsiveness: Severity 2/10. The participant appears engaged and responsive in the conversation.
   - Memory issues: Severity 3/10. There are a few instances where the participant seems to struggle with recalling information, but not consistently.
   - Lack of abstract thinking and synthesis: Severity 2/10. The participant seems capable of abstract thinking and synthesis.
   - Abnormal emotions: Severity 1/10. There are to: ChatGPT.AIAssistant<|im_sep|>ANALYSIS:
4. Vocabulary features

- Word frequency and Word ratio: The participant used a variety of unique words and pronouns. There was a balance between the use of pronouns and specific names, indicating a good grasp of pronouns. Score: 2
- Phrase frequency and Phrase ratio: There was a varied usage of phrases, indicating good command over language. Score: 2
- Lexical complexity: The participant used a good mix of simple and complex words, with no indication of a limited vocabulary. Score: 2
- Limited vocabulary: The participant demonstrated a varied vocabulary without significant repetition. Score: 2
- Increased verbosity: There were instances of verbosity and hesitations, but this was not excessive. Score: 3
- Overlearned phrases: There were no cliches or stock phrases used. Score: 1
- Improper use of pronouns: The participant used pronouns appropriately. Score: 1
- Hesitation (Word-finding difficulties): Some hesitation and filler words were present, which may suggest minor word-finding difficulties. Score: 3
- Circumlocution (Word-finding difficulties): No evidence of circumlocution. Score: 1
- Tip-of-the-tongue phenomenon: No instances were detected. Score: 1
- Repetition: Some repetition of phrases, but not significantly severe. Score: 2

2. Syntax and Grammar features

- Syntactic complexity: The participant used both simple and complex sentence structures. Score: 2
- Grammatical components: Grammatical components were used correctly. Score: 1
- Simplified sentence structure and grammar: No significant simplification in sentence structure or grammar was detected. Score: 2
- Grammatical errors: Few grammatical errors were made. Score: 2
- Difficulty with complex ideas: The participant seemed to handle complex ideas well. Score: 2

3. Semantics features

- Semantic density: The participant provided a good amount of meaningful information in their responses. Score: 2
- Semantic coherence: The participant's responses were mostly coherent and logical. Score: 2
- Semantic paraphasias: No semantic paraphasias were detected. Score: 1
- Lack of coherence: The participant was generally coherent. Score: 2
- Consistency in errors: The errors made were not consistent. Score: 2
- Lack of semantic fluency: The participant was able to name items and describe processes. Score: 1
- Disorganized narrative structure: No evidence of disorganized narrative structure. Score: 1
- Orientation Issues: No confusion about date, time, place, or sequence of events. Score: 1
- Decline in conversational engagement and responsiveness: The participant was engaged and responsive in conversation. Score: 1
- Memory issues: Some memory issues were evident, but these were not severe. Score: 3
- Lack of abstract thinking and synthesis: The participant was able to express abstract thoughts. Score: 1
- Abnormal emotions: No abnormal emotions were detected. Score: 1

CONCLUSION: Overall, the participant's cognitive function seems to be largely normal, with minor instances of hesitations, some verbosity, and a few memory issues. However, these are not severe and could potentially be attributed to factors such as stress or fatigue rather than MCI. Therefore, based on the responses in this context, the participant is grouped under the Probably Normal Cognition Group. Remember, this is an AI-based assessment and any concerns about cognitive function should be followed up with a healthcare professional
