## Supplementary material for "Text dialogue analysis Based ChatGPT for Primary Screening of Mild Cognitive Impairment": Data(GPT4_vs_GPT3.5_model1-5): 113_GPT4 (4).docx

ChatGPT

In assessing the language features of this participant, it's important to take into account the context and format of the interaction as well as the individual performance. Let's examine vocabulary, syntax and grammar, and semantic features.

Vocabulary Features: a. The participant uses a variety of words and phrases to describe scenes and recall stories. There is no unusual word or phrase repetition. b. Lexical complexity appears normal, as the participant uses a mix of simple and more complex words. c. Assessment of specific aspects:

- Limited vocabulary: 2/10. The participant uses a good variety of words and phrases.
- Increased verbosity: 1/10. The participant is not overly verbose.
- Overlearned phrases: 2/10. The participant uses some repeated phrases, but they are contextually appropriate.
- Improper use of pronouns: 1/10. Pronoun use seems correct.
- Hesitation: 4/10. There are moments of hesitation but they do not seem abnormal.
- Circumlocution: 2/10. Minimal circumlocution is noted.
- Tip-of-the-tongue phenomenon: 3/10. Some evidence during the narrative.
- Repetition: 2/10. Only occasional repetition observed.

Syntax and Grammar Features: a. Syntactic complexity and grammatical components are normal. The participant produces grammatically correct sentences with both simple and complex structures. b. As for the specific aspects:

- Simplified sentence structure and grammar: 2/10. There is a mix of sentence structures.
- Grammatical errors: 2/10. Only occasional minor errors observed.
- Difficulty with complex ideas: 2/10. The participant seems capable of understanding and expressing complex ideas.

Semantic Features: a. Semantic density and coherence are normal, with the participant maintaining a coherent narrative throughout. b. Assessment of specific aspects:

- Semantic paraphasias: 2/10. No clear evidence of this.
- Lack of coherence: 2/10. The participant's responses are generally coherent.
- Consistency in errors: 2/10. There are no consistent errors in the participant's language.
- Lack of semantic fluency: 2/10. The participant is able to respond fluently.
- Disorganized narrative structure: 2/10. The participant's narratives are organized.
- Orientation issues: 2/10. The participant seems oriented to the task.
- Decline in conversational engagement and responsiveness: 2/10. The participant remains engaged and responsive.
- Memory issues: 3/10. Slight issues in recalling stories.
- Lack of abstract thinking and synthesis: 2/10. The participant seems capable of abstract thinking and synthesis.
- Abnormal emotions: 1/10. No clear evidence of this.

Based on this analysis, the participant seems to belong to the Probably Normal Cognition Group. It's important to remember that this is a preliminary assessment based on limited data and is not meant to provide a formal diagnosis. Further comprehensive evaluation would be required to confirm any indications of MCI
