## Supplementary material for "Text dialogue analysis Based ChatGPT for Primary Screening of Mild Cognitive Impairment": Data(GPT4_vs_GPT3.5_model1-5): 113_GPT4 (5).docx

ChatGPT

Vocabulary features:

a. The participant demonstrates a good variety of unique words and phrases, with moderate word repetition and use of pronouns, suggesting a balanced word and phrase ratio. However, there is some difficulty with specific names, such as 'the sink' and 'the saucers,' indicating a potential mild issue with noun recall. The lexical complexity is average - most of the vocabulary is relatively simple and commonly used, but the participant also uses a few less frequent words like 'countertop' and 'trout fishing.' Overall, I'd rate the severity of this indicator as a 3.

b. The tip-of-the-tongue phenomenon is evident in this case, particularly when the participant is trying to recall specific names or terms, for example, when the participant was struggling to say 'sink' or 'saucers.' The severity of this issue is not extreme, but it's enough to be noticeable. Therefore, I'd rate the severity of this indicator as a 4.

Syntax and grammar features:

a. The syntactic complexity and grammatical components of the participant's language are mostly adequate. The sentences are largely grammatically correct and vary in length and complexity, indicating a reasonably intact ability to structure and generate sentences. However, there is some minor difficulty with more complex sentence structures, which is evidenced by pauses and filler words ('um,' 'uh'). I'd rate the severity of this indicator as a 4.

b. The participant does not seem to have substantial difficulty with complex ideas. They are able to understand and respond appropriately to the investigator's instructions, and they can generate somewhat complex narratives about the pictures and the stories they're asked to recall. However, there are instances of struggle in constructing more complex narratives, which might suggest some mild difficulty with complex ideas. I'd rate the severity of this indicator as a 4.

Semantics features:

a. The semantic density and coherence of the participant's language are generally good. The participant uses a balance of content and function words and maintains a coherent narrative, despite some pauses and repetition. Therefore, I'd rate the severity of this indicator as a 3.

b. The participant does show some signs of memory issues. They struggle to recall certain details and often repeat questions or phrases. However, they do demonstrate the ability to recall major plot points of stories and some specific details, indicating that these memory issues might be mild. I'd rate the severity of this indicator as a 5.

Based on the comprehensive synthesis of all the indicators, the participant's language and cognitive abilities seem to fall somewhere between normal cognition and mild cognitive impairment. However, the slight issues with recall, memory, and handling complex ideas might suggest a potential mild cognitive impairment. It's important to note that a more comprehensive assessment would be needed for a definitive categorization. Therefore, I would place the participant in the Probably Mild Cognitive Impairment Group, but close to the boundary with the Probably Normal Cognition Group.
