## Supplementary material for "Text dialogue analysis Based ChatGPT for Primary Screening of Mild Cognitive Impairment": Data(GPT4_vs_GPT3.5_model1-5): 113_GPT4.docx

ChatGPT

Assessment of PARTICIPANT:

Vocabulary Features:

- Word frequency: 138 unique words
- Word ratio: 138 unique words / 227 total words = 0.6079
- Phrase frequency: 95 unique phrases
- Phrase ratio: 95 unique phrases / 105 total phrases = 0.9048
- Difficulty finding the right words: Some usage of filler words ("uh", "um") and occasional pauses
- Lexical complexity: Moderate variety and sophistication, with occasional rare words
- Limited vocabulary: Some repetition of simple words, but overall good variety

Semantics Features:

- Semantic density: Moderate amount of meaningful information, with a reasonable balance of content words and function words
- Inability to convey meaning: Minimal difficulty in expressing ideas; most ideas were communicated clearly
- Semantic coherence: Generally coherent and meaningful, with logical relationships between words, phrases, and sentences
- Lack of coherence: Occasional disjointed thoughts, but overall a logical flow of ideas

Syntax and Grammar Features:

- Syntactic complexity: Moderate complexity in sentence structure, with a mix of simple and complex sentences
- Simplified sentence structure: Some short, simple sentences, but also more complex structures
- Grammatical components: Mostly correct grammar usage, with a few minor errors
- Grammatical errors: Occasional mistakes in verb tense and subject-verb agreement

Based on the above assessment, I would assign PARTICIPANT to the Probably Normal group. While there are some instances of difficulty finding the right words and occasional grammatical errors, the overall language abilities of the participant do not indicate significant cognitive impairment
