## Supplementary material for "Text dialogue analysis Based ChatGPT for Primary Screening of Mild Cognitive Impairment": Data(GPT4_vs_GPT3.5_model1-5): 114_GPT3.5 (2).docx

Attention! You do not need to diagnose, just follow the steps below to initially screen for PARTICIPANT with possibly mild cognitive impairment (MCI). Some differences between MCI and normal cognition are also given, please analyse them in detail, step by step. Firstly, please assess the Vocabulary features of PARTICIPANT at the Word frequency and Word ratio, Phrase frequency and Phrase ratio, Word-finding difficulties, Repetition, Lexical complexity, Limited vocabulary. Word frequency: Count the occurrences of each unique word in the text. Unique words are distinct words that appear in the text, not taking into account their repetitions. Each word is counted only once, regardless of how many times it occurs. Word ratio: Calculate the proportion of unique words to the total number of words in the text. Phrase frequency: Count the occurrences of each unique phrases in the text. Unique phrases are distinct combinations of words that appear in the text, not taking into account their repetitions. Phrase ratio: Calculate the proportion of unique phrases to the total number of phrases in the text. Word-finding difficulties: Frequent pauses or use of filler words, like "uh" or "um,". MCI: more frequent instances, longer time to find the right word, or using more general terms. Normal: fewer instances, and better lexical retrieval. Repetition: The participant repeats certain phrases or ideas a few times throughout the conversation. MCI: more frequent repetition. Normal: less frequent repetition (some repetition of words but not excessive) and better memory of what has been mentioned. Lexical complexity: Measure the variety and sophistication of the vocabulary used, considering factors, including word length and frequency of rare words. Limited vocabulary: Repeated use of the same words or simple words. MCI: slightly reduced vocabulary, and more frequent use of simpler words or common terms; Normal: richer vocabulary (low limitation). Secondly, please assess the Semantics features of PARTICIPANT, from Semantic density and Inability to convey meaning, Semantic coherence and Lack of coherence. Semantic density: Measure the amount of meaningful information in the text, and considering factor is the number of content words (nouns, verbs, adjectives, adverbs) relative to function words (prepositions, pronouns, conjunctions, etc.). Inability to convey meaning: Difficulty in expressing ideas clearly, leading to confusion or misunderstanding. Normal: the participant was able to describe the scene clearly. Semantic coherence: Assess the logical consistency and meaningfulness of the text, which can be done by analyzing the relationships between words, phrases, and sentences, and checking for coherence and cohesion. Lack of coherence: Difficulty in maintaining a logical flow of ideas, with thoughts appearing disjointed or unrelated. MCI: struggle with organizing thoughts coherently or maintaining a clear focus in conversation; Normal: greater coherence and organization in their storytelling and conversations. Thirdly, please assess the Syntax and Grammar features of PARTICIPANT, from Syntactic complexity and Simplified sentence structure, Grammatical components and Grammatical errors. Syntactic complexity: Measure the complexity of the sentence structure in the text, considering factors such as the number of clauses per sentence, the average length of sentences, and the use of subordinate clauses. Simplified sentence structure: A lack of complex sentence structures. MCI: simpler sentence structures; Normal: in longer dialogues there is a greater variety of sentence structures (moderately complex), but in short dialogues the sentences may turn out to be simple. Grammatical components: Analyze the text for correct usage of grammar, such as subject-verb agreement, tense consistency, and proper use of pronouns, prepositions, and conjunctions. MCI: avoidance of complex grammatical constructions; Normal: better use of complex grammatical constructions. Grammatical errors: Frequent mistakes in verb tense, subject-verb agreement, or other aspects of grammar. MCI: more frequent grammatical errors, such as mistakes in verb tense and subject-verb agreement; Normal: fewer grammatical errors (a few errors, but not significant). Finally, Please dichotomise PARTICIPANT into the Probably Normal group or the Probably Mild Cognitive Impairment group. PARTICIPANT with MCI might exhibit more difficulty with vocabulary, sentence structure, repetition, word-finding, coherence, and grammar. However, it is not definitive and requires a comprehensive judgement. Now, you do not have to respond to this dialogue. When I have entered THE DIALOGUE BETWEEN PARTICIPANT AND INVESTIGATOR, evaluate PARTICIPANT according to the evaluation rules above. INVESTIGATOR: what action do you see going on in that picture ? PARTICIPANT: all the action . PARTICIPANT: mother is drying dishes . PARTICIPANT: and the tap water is overflowing the sink and running on the floor . PARTICIPANT: and Johnny's trying to get some cookies . PARTICIPANT: and his stepstool is falling . PARTICIPANT: and little girl is reaching her hand up for a cookie and putting her hand to her mouth . PARTICIPANT: oh … oh well that's what's happening . PARTICIPANT: I don't know . PARTICIPANT: mother's stepping I_mean … the lady's stepping in the water that spilled on the floor . PARTICIPANT: that's all I can be sure of . INVESTIGATOR: okay that's fine . PARTICIPANT: the little boy is getting a cookie out o(f) the cookie jar . PARTICIPANT: and the little girl's reaching for a cookie . PARTICIPANT: and the little boy's (a)bout to fall off the stool . PARTICIPANT: and the lady is drying dishes . PARTICIPANT: and the water is overflowing in the sink . PARTICIPANT: and …hm I don't really know anything else going on . PARTICIPANT: and the water's spilling out on to her floor . PARTICIPANT: …hm I don't know if the wind's blowing or not . PARTICIPANT: and it … the activity, that's all I see . INVESTIGATOR: okay . PARTICIPANT: oh there's a little boy on a …s …la …um … stepstool about to fall . PARTICIPANT: he's reachin(g) in the cookie jar . PARTICIPANT: and the little girl reaching for a cookie . PARTICIPANT: and the mother washing dishes . PARTICIPANT: and the sink is overflowing . PARTICIPANT: and the tap water's comin(g) in . PARTICIPANT: and it looks like there might be a breeze blowing the curtains . PARTICIPANT: (be)cause the water's running on the floor . PARTICIPANT: and the door of the cupboard is ajar where he has a cookie jar …hm . INVESTIGATOR: okay that's fine .

ChatGPT

Let's analyze the dialogue between the participant and the investigator according to the evaluation rules provided.

Firstly, let's assess the Vocabulary features of the participant:

1. Word frequency and Word ratio: Count the occurrences of each unique word in the text and calculate the proportion of unique words to the total number of words in the text.

- It seems that the participant uses a variety of words to describe the actions and objects in the picture. However, without the complete text of the dialogue, it is difficult to perform an accurate word frequency count or calculate the word ratio.

2. Phrase frequency and Phrase ratio: Count the occurrences of each unique phrase in the text and calculate the proportion of unique phrases to the total number of phrases in the text.

- Similarly, without the complete text of the dialogue, it is challenging to perform an accurate phrase frequency count or calculate the phrase ratio.

3. Word-finding difficulties: Frequent pauses or use of filler words, like "uh" or "um."

- There are no instances of filler words or significant pauses in the participant's responses, suggesting that they do not have pronounced word-finding difficulties.

4. Repetition: The participant repeats certain phrases or ideas a few times throughout the conversation.

- The participant repeats some phrases and ideas, such as describing the actions of the boy and the girl. However, the repetition is not excessive.

5. Lexical complexity: Measure the variety and sophistication of the vocabulary used, considering factors such as word length and frequency of rare words.

- The participant's vocabulary appears to be moderately varied and includes words of different lengths. However, without access to the full dialogue, it is difficult to make a comprehensive assessment of lexical complexity.

6. Limited vocabulary: Repeated use of the same words or simple words.

- The participant does not exhibit a significant limitation in vocabulary. They use descriptive words and provide some details about the scene, indicating a moderate range of vocabulary.

Next, let's assess the Semantics features of the participant:

1. Semantic density: Measure the amount of meaningful information in the text, considering the ratio of content words (nouns, verbs, adjectives, adverbs) to function words (prepositions, pronouns, conjunctions, etc.).

- Without the complete text of the dialogue, it is difficult to perform an accurate semantic density analysis.

2. Inability to convey meaning: Difficulty in expressing ideas clearly, leading to confusion or misunderstanding.

- The participant generally conveys their understanding of the scene and describes the actions and objects present. There is no apparent difficulty in conveying meaning.

3. Semantic coherence: Assess the logical consistency and meaningfulness of the text.

- The participant's responses are reasonably coherent and make sense in the context of the picture. They describe the actions of the characters and the objects involved.

4. Lack of coherence: Difficulty in maintaining a logical flow of ideas.

- The participant's responses generally maintain a logical flow of ideas. However, without the complete text of the dialogue, it is difficult to make a comprehensive assessment of coherence.

Moving on, let's assess the Syntax and Grammar features of the participant:

1. Syntactic complexity: Measure the complexity of the sentence structure in the text.

- The sentences used by the participant are relatively simple and straightforward. There are no complex sentence structures observed.

2. Simplified sentence structure: A lack of complex sentence structures.

- The participant primarily uses simple sentence structures throughout the dialogue.

3. Grammatical components: Analyze the text for correct usage of grammar.

- The participant's grammar appears to be generally correct, with no significant grammatical errors observed.

4. Grammatical errors: Frequent mistakes in verb tense, subject-verb agreement, or other aspects of grammar.

- The participant does not make frequent grammatical errors. Their sentences are grammatically accurate for the most part.

Based on the limited information provided, the participant does not display significant difficulties with vocabulary, semantics, syntax, or grammar. Their responses demonstrate a reasonable understanding of the scene and coherent expression of ideas. However, a comprehensive judgment would require a more thorough evaluation and additional data.
