## Supplementary material for "Text dialogue analysis Based ChatGPT for Primary Screening of Mild Cognitive Impairment": Data(GPT4_vs_GPT3.5_model1-5): 114_GPT3.5 (3).docx

ChatGPT

Based on the provided conversation, here is an analysis of the indicators and evaluation indicators for the participant's possible mild cognitive impairment (MCI):

1. Limited vocabulary: Severity: 3 The participant demonstrates slightly reduced vocabulary with more frequent use of simple words and common terms.
2. Increased verbosity: Severity: 4 The participant tends to be more verbose and uses more words than necessary to convey their ideas.
3. Overlearned phrases: Severity: 2 There is no significant evidence of overuse of cliches or stock phrases.
4. Improper use of pronouns: Severity: 3 The participant tends to use more pronouns than specific names (or nouns) and occasionally has difficulty with pronouns.
5. Hesitation (Word-finding difficulties): Severity: 4 The participant exhibits hesitation with frequent pauses and filler words, indicating word-finding difficulties.
6. Circumlocution (Word-finding difficulties): Severity: 2 There is no clear evidence of circumlocution or deliberate attempts to be vague or evasive.
7. Tip-of-the-tongue phenomenon: Severity: 3 The participant experiences partial recall or difficulty retrieving specific words, indicating the tip-of-the-tongue phenomenon.
8. Repetition: Severity: 2 The participant does not exhibit excessive repetition of stories, questions, or statements.
9. Simplified sentence structure and grammar: Severity: 3 The participant shows a preference for simpler sentence structures, especially in longer sentences, but also demonstrates some complexity.
10. Grammatical errors: Severity: 2 There are a few grammatical errors, but not significant enough to indicate a major issue.
11. Difficulty with complex ideas: Severity: 3 The participant can handle simpler sentences and ideas but may struggle with more complex narratives or questions.
12. Semantic paraphasias: Severity: 2 There is no clear evidence of semantic paraphasias or errors in word choice.
13. Lack of coherence: Severity: 3 The participant sometimes struggles with organizing thoughts coherently and may change topics frequently or go off on tangents.
14. Consistency in errors: Severity: 2 There is no clear evidence of consistent language errors unusual for the participant's background.
15. Lack of semantic fluency: Severity: 2 The participant does not exhibit significant difficulty in naming items or describing processes.
16. Disorganized narrative structure: Severity: 2 There is no significant evidence of a disorganized narrative structure.
17. Orientation issues: Severity: 2 There is no clear evidence of confusion about date, time, place, or sequence of events.
18. Decline in conversational engagement and responsiveness: Severity: 3 The participant shows some decreased initiation and spontaneity in conversation and may have difficulties following and responding to questions.
19. Memory issues: Severity: 2 The participant does not exhibit clear signs of memory issues, such as repetitive questioning or difficulty recalling information.
20. Lack of abstract thinking and synthesis: Severity: 2 There is no clear evidence of difficulty in understanding or expressing abstract concepts.
21. Abnormal emotions: Severity: 2 There is no clear evidence of abnormal emotions, tone changes, or increased irritability, depression, or apathy.

Based on the comprehensive synthesis of all the indicators, the participant's cognitive function falls within the Probably Normal Cognition Group. However, it's important to note that some indicators are present but not significant enough to conclusively determine mild cognitive impairment. A more comprehensive evaluation by a healthcare professional specializing in cognitive assessments is recommended for a definitive diagnosis.
