## Supplementary material for "Text dialogue analysis Based ChatGPT for Primary Screening of Mild Cognitive Impairment": Data(GPT4_vs_GPT3.5_model1-5): 114_GPT3.5 (4).docx

ChatGPT

Let's assess the participant based on the provided language features:

Vocabulary features: a. Word frequency, word ratio, phrase frequency, and phrase ratio: Without access to the actual word and phrase frequency data, we cannot make an assessment based on this information.

b. Lexical complexity: We don't have specific measures to assess the lexical complexity of the participant's language in this dialogue.

c. Severity assessment:

- Limited vocabulary: The participant shows some diversity in their vocabulary, mentioning different objects and actions in the scene. Severity: 3.
- Increased verbosity: The participant tends to provide additional information, elaborating on the actions and objects in the scene. Severity: 4.
- Overlearned phrases: No evidence of overlearned phrases in the dialogue.
- Improper use of pronouns: No significant improper use of pronouns observed. Severity: 1.
- Hesitation: The participant occasionally hesitates in their responses. Severity: 2.
- Circumlocution: The participant uses some circumlocution when describing certain objects and actions. Severity: 4.
- Tip-of-the-tongue phenomenon: No evidence of tip-of-the-tongue phenomenon in the dialogue.
- Repetition: The participant repeats some phrases, but not excessively. Severity: 3.

Syntax and grammar features: a. Syntactic complexity and grammatical components: The participant's sentences are generally syntactically simple and lack complex structures. There are no detailed grammatical components to assess.

b. Severity assessment:

- Simplified sentence structure and grammar: The participant's sentences are straightforward, lacking complexity. Severity: 6.
- Grammatical errors: No significant grammatical errors observed. Severity: 1.
- Difficulty with complex ideas: The participant does not struggle with complex ideas in this dialogue. Severity: 1.

Semantics features: a. Semantic density and coherence: Without access to the actual semantic density data, we cannot make an assessment based on this information.

b. Severity assessment:

- Semantic paraphasias: No semantic paraphasias observed. Severity: 1.
- Lack of coherence: The participant's responses generally maintain coherence. Severity: 2.
- Consistency in errors: No consistent errors observed. Severity: 1.
- Lack of semantic fluency: The participant shows reasonable fluency in describing the scene. Severity: 2.
- Disorganized narrative structure: The participant's narrative structure is relatively organized. Severity: 2.
- Orientation issues: No orientation issues observed. Severity: 1.
- Decline in conversational engagement and responsiveness: The participant remains engaged and responsive. Severity: 1.
- Memory issues: No significant memory issues observed in this dialogue. Severity: 1.
- Lack of abstract thinking and synthesis: No significant lack of abstract thinking or synthesis observed. Severity: 1.
- Abnormal emotions: No evidence of abnormal emotions in the dialogue.

Based on the comprehensive synthesis of the indicators, we can classify the participant into the Probably Normal Cognition Group. While there are some language features that indicate mild difficulties, overall, the participant's language does not show severe impairments typically associated with mild cognitive impairment (MCI). It's important to note that this assessment is based on the provided dialogue only and a comprehensive evaluation by a trained professional is required for an accurate diagnosis or classification of cognitive impairment.
