## Supplementary material for "Text dialogue analysis Based ChatGPT for Primary Screening of Mild Cognitive Impairment": Data(GPT4_vs_GPT3.5_model1-5): 114_GPT3.5 (5).docx

ChatGPT

Let's analyze the participant's language features based on the provided text:

Vocabulary features: a. Word frequency and word ratio: We can count the occurrences of each unique word, pronoun, and specific name (noun) in the text and calculate the proportion of unique words to the total number of words in the text. We can also calculate the ratio of pronouns to specific names (or nouns). b. Phrase frequency and phrase ratio: We can count the occurrences of each unique phrase in the text and calculate the proportion of unique phrases to the total number of phrases in the text. c. Lexical complexity: We can measure the variety and sophistication of the vocabulary used, considering factors such as word length and frequency of rare words.

Syntax and grammar features: a. Syntactic complexity: We can measure the complexity of the sentence structure in the text, considering factors such as the number of clauses per sentence, the average length of sentences, and the use of subordinate clauses. b. Grammatical components: We can analyze the text for correct usage of grammar, such as subject-verb agreement, tense consistency, and proper use of pronouns, prepositions, and conjunctions.

Semantics features: a. Semantic density: We can measure the amount of meaningful information in the text by considering the number of content words (nouns, verbs, adjectives, adverbs) relative to function words (prepositions, pronouns, conjunctions, etc.). b. Semantic coherence: We can assess the logical consistency and meaningfulness of the text by analyzing the relationships between words, phrases, and sentences and checking for coherence and cohesion.

Now let's evaluate the severity of each indicator on a scale of 1 to 10, with 1 being the least severe and 10 being the most severe.

Vocabulary features: a. Word frequency and word ratio: 5 - The participant uses a mix of common and specific words, but there is no clear indication of a severe deficit in word frequency or word ratio. b. Phrase frequency and phrase ratio: 4 - The participant uses a decent variety of phrases, but there is some repetition and limited use of unique phrases. c. Lexical complexity: 3 - The participant demonstrates an average lexical complexity with no strong evidence of either a sophisticated or limited vocabulary.

Syntax and grammar features: a. Syntactic complexity: 4 - The participant's sentences are generally well-structured, but there is a lack of complexity in terms of the number of clauses and sentence length. b. Grammatical components: 5 - The participant shows mostly correct grammar usage, although there are a few minor errors observed (e.g., "the lady's stepping").

Semantics features: a. Semantic density: 6 - The participant includes a decent amount of meaningful information in the text, but there is room for improvement in terms of using more content words relative to function words. b. Semantic coherence: 5 - The participant's responses generally make sense and show logical consistency, although there are occasional inconsistencies or gaps in the narrative.

Based on the comprehensive synthesis of all the indicators, it appears that the participant exhibits some mild language impairments, but the overall severity is not significant. Therefore, based on the provided information, the participant is likely to be categorized into the Probably Normal Cognition Group. However, it's important to note that this analysis is based on a limited text sample, and a comprehensive evaluation by a qualified healthcare professional is necessary to confirm any cognitive impairment.
