## Supplementary material for "Text dialogue analysis Based ChatGPT for Primary Screening of Mild Cognitive Impairment": Data(GPT4_vs_GPT3.5_model1-5): 114_GPT4 (2).docx

ChatGPT

To assess the cognitive status of the participant, I'll follow the steps you've laid out. It's important to remember that this analysis is based on a single written sample and may not provide a complete or entirely accurate picture of the participant's cognitive health.

1. Vocabulary features:
   - Word frequency & Word ratio: Since the task does not provide a total number of unique words and total words, these cannot be calculated.
   - Phrase frequency & Phrase ratio: Similar to word frequency and ratio, without a specific count of total unique phrases and total phrases, these cannot be calculated.
   - Word-finding difficulties: The participant occasionally uses filler words like "um" and "oh" and seems to pause to find words, suggesting some difficulty with word-finding.
   - Repetition: The participant repeats several ideas, such as the boy reaching for the cookie jar, the girl reaching for a cookie, and the water overflowing from the sink.
   - Lexical complexity: The participant uses a mix of simple and moderately complex words. There is some variation in word length and no obvious use of rare words.
   - Limited vocabulary: The participant uses some repeated and simple words, but also uses a variety of different words to describe the scene.
2. Semantic features:
   - Semantic density: The participant uses a reasonable amount of content words relative to function words.
   - Inability to convey meaning: The participant seems capable of expressing their ideas clearly for the most part, but there are instances of uncertainty.
   - Semantic coherence: The participant's description of the scene appears logically consistent and meaningful. The relationships between words, phrases, and sentences generally exhibit coherence and cohesion.
   - Lack of coherence: The participant generally maintains a logical flow of ideas. There are a few instances where the participant appears to lose focus or struggle with organizing thoughts, but these are not frequent.
3. Syntax and Grammar features:
   - Syntactic complexity: The participant primarily uses simple sentences. There are a few instances of more complex structures, but these are not the norm.
   - Simplified sentence structure: The sentence structures used by the participant are mostly simple, suggesting a tendency towards simpler structures.
   - Grammatical components: The participant uses mostly correct grammar. However, there are instances where the participant avoids more complex grammatical constructions.
   - Grammatical errors: There are a few instances of incorrect verb tenses and other grammatical errors, but these are not frequent.

Given the analysis above, the participant exhibits some characteristics of both normal cognition and Mild Cognitive Impairment (MCI), including occasional word-finding difficulties, repetition, simplified sentence structure, and slightly reduced vocabulary. However, the participant also exhibits a logical flow of ideas and mostly accurate grammar, which are more characteristic of normal cognition.

Based on this single interaction, it is challenging to definitively assign the participant to one group or another. However, due to the presence of certain features associated with MCI, such as word-finding difficulties, repetition, and simplified sentence structure, it might be appropriate to consider the participant as "Probably Mild Cognitive Impairment". However, this is not a definitive conclusion, and further comprehensive assessment would be necessary for a more accurate determination.
