## Supplementary material for "Text dialogue analysis Based ChatGPT for Primary Screening of Mild Cognitive Impairment": Data(GPT4_vs_GPT3.5_model1-5): 114_GPT4 (3).docx

ChatGPT

Based on the responses of the participant and the guidelines provided, here are my initial assessments of the various categories:

1. Vocabulary features:
   - Word frequency and Word ratio: The participant has used a variety of words but the word "cookie" was used quite frequently. Pronouns to specific names ratio seems balanced. (Severity: 3)
   - Phrase frequency and Phrase ratio: There is repetition of phrases such as "the little boy" and "the cookie jar". However, there is also a good variety of phrases. (Severity: 3)
   - Lexical complexity: The vocabulary used is relatively simple, but the conversation context doesn't necessitate complex language. (Severity: 3)
   - Limited vocabulary: The participant's vocabulary appears slightly reduced, and there is some repetition of common words. (Severity: 3)
   - Increased verbosity: Not particularly verbose. (Severity: 1)
   - Overlearned phrases: No cliches or stock phrases are evident. (Severity: 1)
   - Improper use of pronouns: No notable issues. (Severity: 1)
   - Hesitation (Word-finding difficulties): Some hesitations noted, but not very severe. (Severity: 2)
   - Circumlocution (Word-finding difficulties): Not evident. (Severity: 1)
   - Tip-of-the-tongue phenomenon: Not evident. (Severity: 1)
   - Repetition: Some repetition of phrases was observed. (Severity: 2)
2. Syntax and Grammar features:
   - Syntactic complexity: Sentence structure is relatively simple, but appropriate for the conversation. (Severity: 3)
   - Grammatical components: Few minor errors, but generally correct usage. (Severity: 2)
   - Simplified sentence structure and grammar: Participant used simpler structures, but this is contextually appropriate. (Severity: 3)
   - Grammatical errors: Few errors, such as incomplete sentences or phrases. (Severity: 2)
   - Difficulty with complex ideas: No complex ideas were presented, so this can't be accurately assessed.
3. Semantics features:
   - Semantic density: Adequate use of content words vs function words. (Severity: 1)
   - Semantic coherence: The conversation is coherent. (Severity: 1)
   - Semantic paraphasias: Not evident. (Severity: 1)
   - Lack of coherence: The participant maintained coherence throughout. (Severity: 1)
   - Consistency in errors: Not evident. (Severity: 1)
   - Lack of semantic fluency: Not evident. (Severity: 1)
   - Disorganized narrative structure: Not evident. (Severity: 1)
   - Orientation Issues: Not evident. (Severity: 1)
   - Decline in conversational engagement and responsiveness: Not evident. (Severity: 1)
   - Memory issues: Not evident. (Severity: 1)
   - Lack of abstract thinking and synthesis: Not evident. (Severity: 1)
   - Abnormal emotions: Not evident. (Severity: 1)

Given the overall low scores, the participant would be dichotomized into the Probably Normal Cognition Group. However, it's important to note that this assessment can only serve as an initial screening. Further, comprehensive clinical assessments would be necessary for an accurate diagnosis.
