## Supplementary material for "Text dialogue analysis Based ChatGPT for Primary Screening of Mild Cognitive Impairment": Data(GPT4_vs_GPT3.5_model1-5): 114_GPT4 (4).docx

ChatGPT

Based on the provided text, let's assess the participant's language features.

Vocabulary Features: a. The participant repeats phrases such as "the little boy," "cookie jar," "reaching for a cookie," "the water is overflowing." The word and phrase ratio seems balanced, not skewed towards unusual repetition. b. Lexical complexity seems average. The participant uses a variety of everyday words to describe scenes. c. Specific aspects:

- Limited vocabulary: 2/10. Vocabulary seems varied and appropriate.
- Increased verbosity: 1/10. The participant's speech does not appear excessively verbose.
- Overlearned phrases: 2/10. The participant does not significantly overuse any particular phrases.
- Improper use of pronouns: 1/10. No issues noted.
- Hesitation: 3/10. There are instances of hesitation but within normal limits.
- Circumlocution: 2/10. Not significantly present.
- Tip-of-the-tongue phenomenon: 2/10. Some evidence, but within a normal range.
- Repetition: 3/10. Some repetition in narrative descriptions, but not concerning.

Syntax and Grammar Features: a. The participant uses a variety of sentence structures, indicating normal syntactic complexity and understanding of grammatical components. b. Specific aspects:

- Simplified sentence structure and grammar: 2/10. Some simplification but within normal limits.
- Grammatical errors: 1/10. The participant makes very few, if any, grammatical errors.
- Difficulty with complex ideas: 2/10. The participant does not seem to have trouble with complex ideas.

Semantic Features: a. The participant maintains semantic density and coherence throughout their speech. b. Specific aspects:

- Semantic paraphasias: 1/10. No clear instances identified.
- Lack of coherence: 2/10. The participant's narrative is mostly coherent.
- Consistency in errors: 2/10. No consistent errors are noted.
- Lack of semantic fluency: 2/10. The participant seems fluent.
- Disorganized narrative structure: 2/10. Some repetition, but the overall structure is organized.
- Orientation issues: 2/10. The participant seems appropriately oriented to the task.
- Decline in conversational engagement and responsiveness: 2/10. The participant remains engaged and responsive.
- Memory issues: 2/10. No significant memory issues are noted.
- Lack of abstract thinking and synthesis: 2/10. The participant seems capable of abstract thinking and synthesis.
- Abnormal emotions: 1/10. No clear evidence of this.

Based on these scores, the participant seems to fit in the Probably Normal Cognition Group. Please remember that this assessment is not a formal diagnosis and is intended only as an initial screening tool for potential MCI. A more comprehensive evaluation would be needed for a formal diagnosis.
