## Supplementary material for "Text dialogue analysis Based ChatGPT for Primary Screening of Mild Cognitive Impairment": Data(GPT4_vs_GPT3.5_model1-5): 114_GPT4 (5).docx

ChatGPT

Based on the provided text, the following evaluations can be made:

Vocabulary Features: a. The participant does repeat certain phrases (like "cookie jar," "water is overflowing") and words ("cookie," "mother," "water"), but there is a decent variety of words used as well. The participant does not seem to struggle with finding the right words to describe the scene and has a relatively good ratio of unique words to the total number of words. Lexical complexity appears to be average, with some longer words like "overflowing," and "stepstool." It doesn't appear that there is a frequent use of rare words. Score: 3/10 b. The participant does show some signs of the tip-of-the-tongue phenomenon, with multiple instances of hesitation and use of filler words like "um" and "hm." However, the participant always eventually finds the word they are looking for, indicating this issue is not severely impacting their ability to communicate. Score: 4/10

Syntax and Grammar Features: a. The participant uses mostly simple sentence structures, with a few more complex ones. There are few subordinate clauses, and the average sentence length is relatively short. However, the participant's sentences are generally grammatically correct, demonstrating a sound understanding of basic syntax. Score: 3/10 b. The participant seems to handle the complexity of describing the scene reasonably well. However, there are a few instances where the participant seems unsure ("I don't know if the wind's blowing or not"), which might suggest some difficulties with complex ideas. Score: 3/10

Semantic Features: a. The participant's speech contains a good amount of meaningful information, with a decent ratio of content words to function words. The participant effectively communicates the main events in the scene. Semantic coherence is mostly good, though there are a few points where the participant jumps between ideas without clear transitions. Score: 2/10 b. The participant does repeat certain phrases and concepts, which might suggest some memory issues. However, the participant does not seem to forget major elements of the scene, and there are no instances of the participant asking the investigator to repeat questions. Score: 3/10

In conclusion, this participant shows some signs that could be consistent with Mild Cognitive Impairment, such as some difficulties with the tip-of-the-tongue phenomenon and potential memory issues. However, these issues are relatively mild and do not severely impact the participant's ability to communicate or understand the scene they're describing. As such, based on this limited sample, the participant could be classified in the Probably Normal Cognition Group. But, a more detailed cognitive assessment would be needed to confirm this.
