## Supplementary material for "Text dialogue analysis Based ChatGPT for Primary Screening of Mild Cognitive Impairment": Data(GPT4_vs_GPT3.5_model1-5): 114_GPT4.docx

ChatGPT

Based on the conversation between PARTICIPANT and INVESTIGATOR, here is the assessment of the language features:

Vocabulary features:

- Word frequency: 80 unique words
- Word ratio: 80/128 = 0.625
- Phrase frequency: 44 unique phrases
- Phrase ratio: 44/48 = 0.917
- Difficulty finding the right words: Occasional use of filler words and pauses
- Lexical complexity: Moderate variety in vocabulary with some rare words
- Limited vocabulary: Some repetition, but overall a decent variety

Semantics features:

- Semantic density: Moderate content words in relation to function words
- Inability to convey meaning: Participant is mostly able to express ideas clearly
- Semantic coherence: Fairly consistent and meaningful text with logical relationships between words and phrases
- Lack of coherence: Occasional disjointed thoughts, but largely coherent

Syntax and Grammar features:

- Syntactic complexity: Moderate complexity with some use of subordinate clauses
- Simplified sentence structure: Mix of simple and slightly complex sentences
- Grammatical components: Mostly correct usage of grammar
- Grammatical errors: Occasional errors, but not frequent

Given the above assessment, the PARTICIPANT can be assigned to the Probably Normal group. However, it's important to note that this is only an initial screening and not a definitive diagnosis
