## Supplementary material for "Text dialogue analysis Based ChatGPT for Primary Screening of Mild Cognitive Impairment": Data(GPT4_vs_GPT3.5_model1-5): 115_GPT3.5 (2).docx

Attention! You do not need to diagnose, just follow the steps below to initially screen for PARTICIPANT with possibly mild cognitive impairment (MCI). Some differences between MCI and normal cognition are also given, please analyse them in detail, step by step. Firstly, please assess the Vocabulary features of PARTICIPANT at the Word frequency and Word ratio, Phrase frequency and Phrase ratio, Word-finding difficulties, Repetition, Lexical complexity, Limited vocabulary. Word frequency: Count the occurrences of each unique word in the text. Unique words are distinct words that appear in the text, not taking into account their repetitions. Each word is counted only once, regardless of how many times it occurs. Word ratio: Calculate the proportion of unique words to the total number of words in the text. Phrase frequency: Count the occurrences of each unique phrases in the text. Unique phrases are distinct combinations of words that appear in the text, not taking into account their repetitions. Phrase ratio: Calculate the proportion of unique phrases to the total number of phrases in the text. Word-finding difficulties: Frequent pauses or use of filler words, like "uh" or "um,". MCI: more frequent instances, longer time to find the right word, or using more general terms. Normal: fewer instances, and better lexical retrieval. Repetition: The participant repeats certain phrases or ideas a few times throughout the conversation. MCI: more frequent repetition. Normal: less frequent repetition (some repetition of words but not excessive) and better memory of what has been mentioned. Lexical complexity: Measure the variety and sophistication of the vocabulary used, considering factors, including word length and frequency of rare words. Limited vocabulary: Repeated use of the same words or simple words. MCI: slightly reduced vocabulary, and more frequent use of simpler words or common terms; Normal: richer vocabulary (low limitation). Secondly, please assess the Semantics features of PARTICIPANT, from Semantic density and Inability to convey meaning, Semantic coherence and Lack of coherence. Semantic density: Measure the amount of meaningful information in the text, and considering factor is the number of content words (nouns, verbs, adjectives, adverbs) relative to function words (prepositions, pronouns, conjunctions, etc.). Inability to convey meaning: Difficulty in expressing ideas clearly, leading to confusion or misunderstanding. Normal: the participant was able to describe the scene clearly. Semantic coherence: Assess the logical consistency and meaningfulness of the text, which can be done by analyzing the relationships between words, phrases, and sentences, and checking for coherence and cohesion. Lack of coherence: Difficulty in maintaining a logical flow of ideas, with thoughts appearing disjointed or unrelated. MCI: struggle with organizing thoughts coherently or maintaining a clear focus in conversation; Normal: greater coherence and organization in their storytelling and conversations. Thirdly, please assess the Syntax and Grammar features of PARTICIPANT, from Syntactic complexity and Simplified sentence structure, Grammatical components and Grammatical errors. Syntactic complexity: Measure the complexity of the sentence structure in the text, considering factors such as the number of clauses per sentence, the average length of sentences, and the use of subordinate clauses. Simplified sentence structure: A lack of complex sentence structures. MCI: simpler sentence structures; Normal: in longer dialogues there is a greater variety of sentence structures (moderately complex), but in short dialogues the sentences may turn out to be simple. Grammatical components: Analyze the text for correct usage of grammar, such as subject-verb agreement, tense consistency, and proper use of pronouns, prepositions, and conjunctions. MCI: avoidance of complex grammatical constructions; Normal: better use of complex grammatical constructions. Grammatical errors: Frequent mistakes in verb tense, subject-verb agreement, or other aspects of grammar. MCI: more frequent grammatical errors, such as mistakes in verb tense and subject-verb agreement; Normal: fewer grammatical errors (a few errors, but not significant). Finally, Please dichotomise PARTICIPANT into the Probably Normal group or the Probably Mild Cognitive Impairment group. PARTICIPANT with MCI might exhibit more difficulty with vocabulary, sentence structure, repetition, word-finding, coherence, and grammar. However, it is not definitive and requires a comprehensive judgement. Now, you do not have to respond to this dialogue. When I have entered THE DIALOGUE BETWEEN PARTICIPANT AND INVESTIGATOR, evaluate PARTICIPANT according to the evaluation rules above. PARTICIPANT: well the mother's washing the dishes . PARTICIPANT: and the boy is reaching for cookies and he's falling off the chair ( stool ) . PARTICIPANT: and the little girl's laughing at him of … I think . PARTICIPANT: and the water's spilling over onto the floor . PARTICIPANT: … PARTICIPANT: I can't see any other action . PARTICIPANT: I guess she's drying her dishes not washing them . PARTICIPANT: she's doing something with them . INVESTIGATOR: okay . INVESTIGATOR: okay . INVESTIGATOR: just tell me everything that you see happening in this picture . INVESTIGATOR: just everything that you see going on . PARTICIPANT: okay . PARTICIPANT: the little boy is climbing up <in the> … getting cookies out o(f) the cookie jar and is … he's falling . PARTICIPANT: the stool's falling over . PARTICIPANT: and the little girl has her hand up that she wants a cookie and he's trying to hand her one . PARTICIPANT: and the door to the cabinet is open . PARTICIPANT: and the mother is washing dishes . PARTICIPANT: and the … …dishwash the dish water is overflowing in the sink and it's running on to the floor and she's standing in the water . PARTICIPANT: and there's … she's drying a plate or washing it . PARTICIPANT: whatever . PARTICIPANT: and there's two cups and a plate on the counter top . PARTICIPANT: and it looks like the window is open . PARTICIPANT: and it has curtains pulled back at the window . PARTICIPANT: and there's a little lane going around the house towards the back I imagine with shrubs . PARTICIPANT: and you can see another window .

ChatGPT

2 / 2

Based on the provided dialogue, let's evaluate the participant according to the evaluation rules mentioned earlier.

1. Vocabulary features:
   - Word frequency: Count the occurrences of each unique word in the text.
     - The participant mentions various words related to the scene, such as "mother," "washing," "dishes," "boy," "reaching," "cookies," "falling," "chair," "girl," "laughing," "water," "spilling," "floor," "action," "drying," "plate," "washing," "cups," "counter," "window," "open," "curtains," "shrub," etc.
   - Word ratio: Calculate the proportion of unique words to the total number of words in the text.
     - To determine this, we need the total number of words in the text, including repetitions, which is not provided.
   - Phrase frequency: Count the occurrences of each unique phrase in the text.
     - The participant mentions several unique phrases, such as "mother's washing the dishes," "the boy is reaching for cookies and he's falling off the chair (stool)," "the little girl's laughing at him," "the water's spilling over onto the floor," "she's drying her dishes not washing them," etc.
   - Phrase ratio: Calculate the proportion of unique phrases to the total number of phrases in the text.
     - To determine this, we need the total number of phrases in the text, which is not provided.
   - Word-finding difficulties:
     - The participant doesn't exhibit significant word-finding difficulties, as they are able to describe various objects, actions, and events in the scene without struggling or using filler words excessively.
   - Repetition:
     - The participant repeats some phrases, like "washing the dishes" and "drying a plate," but the repetition is not excessive.
   - Lexical complexity:
     - The participant uses a mix of simple and descriptive words to convey their observations, which suggests a moderately varied vocabulary. However, without access to the full text, it's challenging to assess the complexity accurately.
2. Semantics features:
   - Semantic density:
     - Without the total number of content words and function words in the text, it's difficult to calculate the semantic density accurately.
   - Inability to convey meaning:
     - The participant is able to express their ideas clearly and describe the scene in detail, indicating no significant difficulty in conveying meaning.
   - Semantic coherence:
     - The participant's descriptions show logical consistency and meaningfulness. They provide coherent details about the actions, objects, and relationships in the scene.
   - Lack of coherence:
     - The participant maintains a logical flow of ideas and provides relevant information without appearing disjointed or unrelated.
3. Syntax and Grammar features:
   - Syntactic complexity:
     - The sentence structures used by the participant are mainly simple, with a few complex sentences. However, the dialogue is relatively short, making it difficult to assess the overall complexity accurately.
   - Simplified sentence structure:
     - The participant predominantly uses simple sentence structures to convey their thoughts.
   - Grammatical components:
     - The participant generally demonstrates correct usage of grammar, such as subject-verb agreement and appropriate use of pronouns and prepositions.
   - Grammatical errors:
     - The provided dialogue does not contain any significant grammatical errors.

Based on the evaluation of the participant's vocabulary, semantics, and syntax/grammar features in the given dialogue, there are no clear indications of mild cognitive impairment (MCI). The participant exhibits reasonably coherent and meaningful descriptions, with a mix of simple and descriptive vocabulary. However, it's important to note that this evaluation is based on a short dialogue, and a comprehensive assessment would require more extensive and varied conversations over time to make a more accurate judgment.
